# Estimating national-level measles case fatality ratios: an updated systematic review and modelling study

**DOI:** 10.1101/2022.10.05.22280730

**Authors:** Alyssa N. Sbarra, Jonathan F. Mosser, Mark Jit, Matthew Ferrari, Rebecca E. Ramshaw, Patrick O’Connor, L. Kendall Krause, Emma L. B. Rogowski, Allison Portnoy

## Abstract

Background: To understand current and mitigate future measles mortality burden, it is critical to have robust estimates of measles case fatality. Estimates of measles case fatality ratios (CFR) that are age-, location-, and time-specific are essential to capture variation in underlying population-level factors—such as vaccination coverage and measles incidence—that contribute systematically to increases or decreases in case fatality. In this study, we updated estimates of measles CFR by expanding upon previous systematic reviews and implementing a meta-regression model.

Methods: We conducted a literature review of all available data on measles case fatality from 1980 through 2019 from low- and middle-income countries and extracted the most granular information available on measles cases and deaths. Using this data and a suite of covariates related to measles CFR, we implemented a Bayesian meta-regression model to produce estimates of measles CFR by location and age from 1990 to 2019.

Findings: We identified 245 sources that contained information on both measles cases and deaths. In 2019, we estimated a mean all-age CFR among community-based settings of 1.32% (95% Uncertainty Interval (UI): 1.28 – 1.36%) and among hospital-based settings of 5.35% (95% UI: 5.08 – 5.64%). In community-based settings, we estimated 2019 CFR to be 3.03% (95% UI: 2.89 – 3.16%) among under-one year olds, 1.63% (95% UI: 1.58 – 1.68%) among 1 to 4 year olds, 0.84% (95% UI: 0.80 – 0.87%) among 5 to 9 year olds, and 0.67% (95% UI: 0.64 – 0.70%) among 10 to 14 year olds. Between 1990 and 2019, we estimated measles vaccination has averted approximately 71 million deaths due to decreased measles mortality.

Interpretation: While CFRs have declined, there are still large heterogeneities across locations and ages. Our updated methodologic framework and estimates can be used to evaluate the effect of measles control and vaccination programmes on reducing preventable measles mortality burden.

**Research in context:** *Evidence before this study:* Two previous systematic reviews have synthesized individual studies of measles CFR. The first review, by Wolfson et al., was published in 2009 and used 58 community-based studies in 29 countries to provide global estimates of measles CFR. Wolfson and colleagues published a descriptive analysis suggesting global estimates of CFR with a mean of 3.3%, a median of 3.9%, and range from 0 – 40.1%. For outbreak investigations, results suggested a median CFR of 5.2% (95% CI: 2.6 – 11.6%). These results were the first figures of measles CFR beyond single country-year studies, reports, and investigations; however, this study only included community-based studies, did not produce estimates for other locations or years, and did not stratify by other underlying determinants of mortality, such as development status of each country. The later review by Portnoy and others was published in 2019 and included data from 1980 to 2016 from low- and middle-income countries; studies included reports from both community-based (n=85) and hospital-based (n=39) settings. Following the review, authors used a log-linear prediction model with a select set of covariates, generally understood to be related to measles CFR (previous vaccination history [first dose MCV coverage used as a proxy], estimated measles attack rate) and indirectly associated with measles CFR (under-5 mortality [U5M], total fertility rate, proportion of population living in urban areas, population density). The authors reported predicted CFR stratified by year, country-development status, under-5 mortality rate, care-setting (community versus hospital), age (under- or over-5 years), and calendar year from 1990 to 2030. Results predicted a mean CFR of 2.2% (95% CI: 0.7 – 4.5%) for years 1990–2015, with stratification for community (CFR: 1.5, 95% CI: 0.5 – 3.1%) and hospital-based studies (CFR: 2.9, 95% CI: 0.9 – 6.0%).

*Added value of this study:* Our study produces age-, location- and year-specific estimates of measles CFR from 1990 to 2019 by building on previous estimates in three ways. First, it updates the existing body of evidence to those published through 2020 and non-English studies. Second, it incorporates an explicit conceptual framework based on literature review and expert consultation to identify a suite of covariates demonstrated to be related to measles CFR at the population level. Last, it uses a Bayesian meta-regression model, with a flexible spline component to better capture variation in CFR by age.

*Implications of all the available evidence:* This model, along with corresponding estimates, can contribute to a deeper understanding of measles CFR and allow for a more robust assessment of vaccination programmes and other interventions to reduce measles mortality burden.

## Introduction

In 2019, over 207,500 deaths were estimated to be attributable to measles.^1^ However, the exact figure cannot be directly measured because there is a lack of reliable data on measles mortality from most high-burden settings. Instead, measles mortality is usually estimated by combining information on estimates of measles incidence and measles case fatality ratios (CFRs).^2^ Hence an accurate understanding of measles CFRs across different times and geographies is vital for disease burden estimation. Additionally, a robust understanding of country-level CFRs can help to identify opportunities for health system strengthening and inform assessments of the impact of vaccination programmes. Cohort-based and cross-sectional studies and outbreak investigations provide reports of measles CFR in the literature, but are often limited to specific locations, years, and vaccine coverage levels.^3^

Previous work has reviewed the body of available published data on measles CFR^4^; more recently an additional study^5^ has also modelled estimates of measles CFR for low- and middle-income countries (LMICs) among persons aged under and over 5-years old and in both community- and hospital-based settings. Time-varying estimates of measles CFR are critical to understanding evolving patterns of measles mortality across time and location and have been instrumental in understanding acute measles deaths and the impact of various vaccination scenarios.^6^ While a major advancement, these previously published estimates do not include CFR data from the most recent years, nor an underlying conceptual model between CFR and associated covariates.^7^

Additionally, CFR estimates stratified by broad age categories may obscure important variation within these age groups, particularly for younger children. Both previous systematic review studies have shown that measles CFR is higher in persons younger than five years of age compared with children five years of age or older. However, there are likely to be critically important age-specific variations in infants and younger children related to maternal antibody presence, immune system maturation and vaccination status, among other factors, that go uncaptured in a composite estimate of CFR among all under-five year olds.^8^ Given that measles incidence tends to be highest among young children who are unvaccinated^9^, an accurate understanding of CFR among these ages is critical for understanding measles mortality burden and developing targeted interventions.

In this study, we conducted a full literature review of measles CFR data representing both community and hospital cases in LMICs. We expanded on previous reviews by including data from non-English studies, examining all studies for the most granular age data available, and extending the scope to include data published through 2020 (representing cases occurring through 2019). Additionally, we developed a Bayesian meta-regression model to produce location-, year-, and age-specific estimates of measles CFR from 1990 to 2019.

## Methods

### Literature review and data extraction

We extended previously published systematic literature reviews on measles CFRs in LMICs to include data through 2020, and non-English studies. To do this, we searched PubMed from January 1, 1980 through December 31, 2020 using the following search string:

> ((((measles[MeSH Terms] OR measles) AND (mortality[MeSH Terms] OR mortality OR “case fatality rate” OR “case fatality ratio” OR “case fatality”)))

In addition to the literature search, we added studies from previous systematic reviews^3,5^ and the Global Burden of Diseases, Injuries, and Risk Factors study^10^ prior to de-duplicating and screening. We included studies if they were included in the previous systematic reviews or, if upon screening, they contained primary data on measles cases and deaths from hospital-, community-, or surveillance-based reports, including outbreak investigations. We excluded studies if they were not in humans, contained non-original or non-primary data (i.e., reported on outcomes of another study), reported on data from global or regional surveillance, or did not contain relevant information on measles cases and deaths. Additionally, like the previous reviews, we excluded studies: (1) only reporting on measles cases and deaths among restricted populations, such as from communities of internally displaced persons or among persons living with HIV; (2) only reporting on long-term measles mortality, such as from subacute sclerosing panencephalitis; or (3) from a high-income country as defined by the World Bank country income classification in 2017.

We extracted the following data from each study: number of measles cases, number of measles deaths, study year, age, geographic location, whether the source was from a hospital- or community-based setting, and whether the source was reporting on an outbreak setting. As available, we also extracted laboratory confirmation of measles cases and length of time following onset of rash for a death to be considered attributable to measles. For each study, we computed the annual age-specific case-fatality ratios; we included all suspected measles cases and considered all deaths within 30 days of rash onset, unless cases or acute deaths were defined otherwise in each study.

### Covariate selection and analysis

An overview of our entire covariate selection and modelling process can be found in Supplementary Figure 1. Previous work identified measles incidence and age to be critical covariates when assessing measles CFR.^3,5,7^ We additionally selected covariates based on a systematic review and expert consultation^7^ that identified five possible underlying mechanisms contributing to systematic increases or decreases in measles CFR (health system access and care seeking behaviours, health system quality, nutritional status, measles control and epidemiology, and risk of secondary infection) and related population-level indicators with evidence of an association with measles CFR (average household size, educational attainment, first-dose coverage of measles-containing vaccine (MCV1), human immunodeficiency virus (HIV) prevalence, level of health care availability, second-dose coverage of measles-containing vaccine (MCV2), stunting prevalence, surrounding conflict, travel time to nearest health care facility, under-five mortality rate, underweight prevalence, vitamin A deficiency prevalence, vitamin A treatment prevalence, and wasting prevalence).

**Figure 1.**
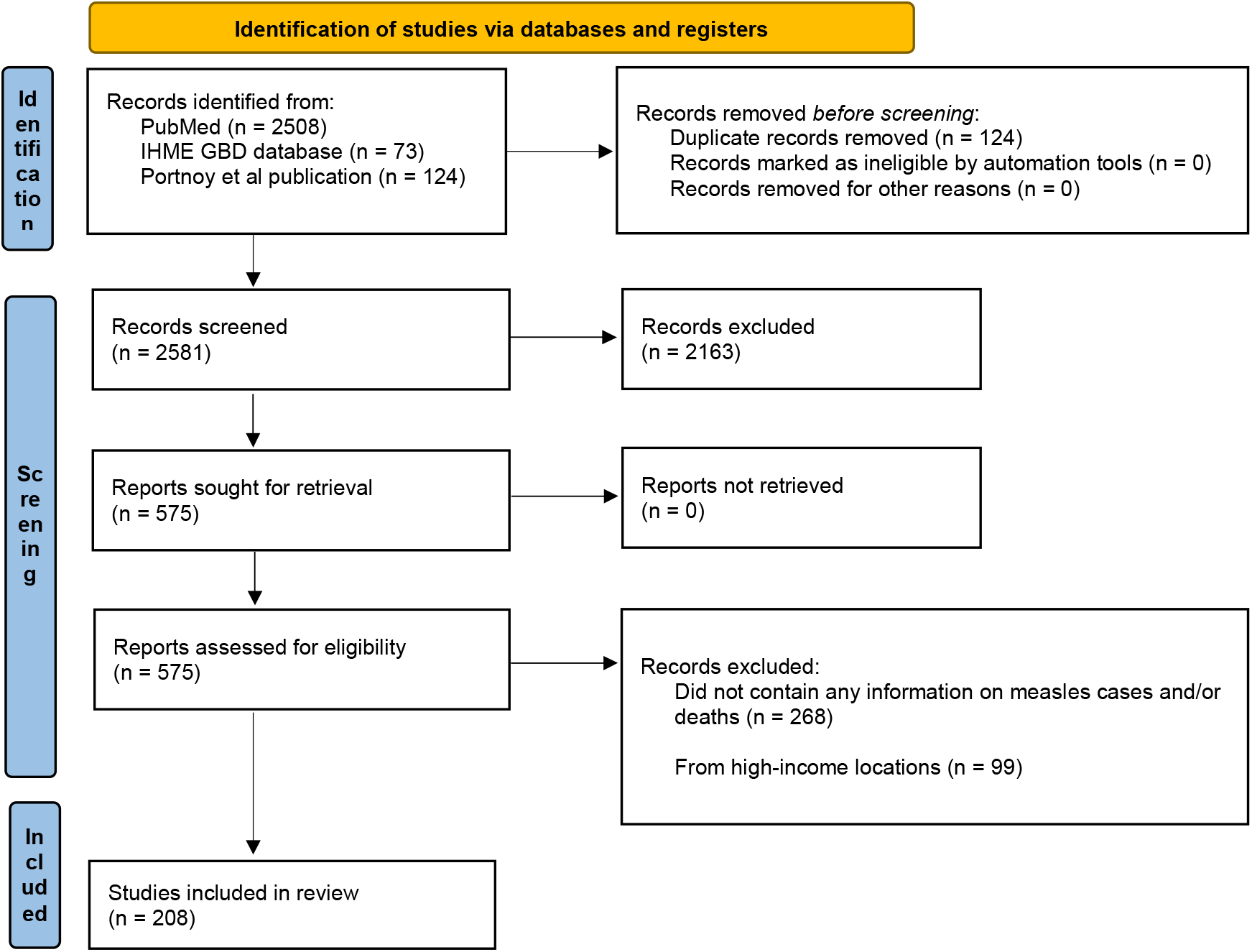
PRISMA diagram. The number of studies at each stage of the review is shown for the systematic review.

We used estimates of country-specific annual measles incidence that were generated using previously described methods.^11^ For the remaining possible covariates, we searched databases of health-related indicators (from World Health Organization, United Nations, World Bank, Global Burden of Disease (GBD) Study) to identify possible covariate sets that could be used to represent each indicator. For the following indicators, we were able to find an appropriate covariate set available for nearly all countries and years from 1980 to 2019: HIV prevalence^10^, MCV1 coverage^12^, under-five mortality rate^13^, vitamin A deficiency prevalence^10^, and wasting prevalence^10^. If a covariate set was not available, we identified a proxy covariate set if an appropriate alternative existed based on expert group review. Proxy covariate sets, identified by expert consultation, include the following: gross domestic product (GDP) per capita^13^ (for level of health care available), maternal education^10^ (for educational attainment), proportion living in an urban setting^13^ (for travel time to nearest health facility), total fertility rate^13^ (for average household size), and mortality rate due to war and terrorism^10^ (for surrounding conflict). For vitamin A treatment, we were unable to identify an appropriate proxy covariate set; therefore, vitamin A treatment was excluded as a covariate in our model.

If specific country-years were missing in covariate sets, we either computed an interpolated or projected value if there were fewer than 20% of years missing per country for the covariate or used the GBD regional average of covariate values for a country if there was 20% missingness or more. For covariates with less than 20% missingness per country, we linearly interpolated missing years using values from adjacent available years of covariate values. If missing covariate values were at the beginning or end of the covariate time series, we used an annualized rate of change with exponential weights to compute the projected covariate values either forwards or backwards in time to complete the full time series, such that weights are more representative of years in the time-series that are closer to either the most recent year for forwards projections and most historic year for backwards projections. For wasting prevalence specifically, which was only available from 1990 onwards, we held the 1990 value constant from 1980 to 1990. We assumed that all covariates took their values in 1980 for all years before 1980.

We conducted a two-step data analysis of the covariate sets to determine the strength of relationship and predictive capability of each covariate in describing underlying trends in CFR. Covariates were grouped into five mechanisms as previously described.^7^ To examine the correlation of covariates, we then calculated the pairwise correlation of each of the covariates in each mechanistic group. If there was a correlation > 0.8 for any pairwise comparison, covariates were removed sequentially based on the average highest collinearity between all other covariates in the mechanistic group. Next, as a second step, we performed a simple linear regression of the remaining covariates per mechanistic group with the measles CFR dataset.

Covariates were removed as uninformative if they had a p-value greater than two-times the average p-value across all covariates (i.e., greater than 0.33). The final list of covariates selected for inclusion were: age, a categorical indicator for community versus hospital studies, measles incidence, mortality rate due to war and terrorism, maternal education, GDP per capita, HIV prevalence, MCV1 coverage, total fertility rate, under-5 mortality rate, proportion living in urban settings, vitamin A deficiency, and wasting prevalence; specific details of this process can be found in Supplementary Information Section 1.

Finally, we selected a transformation (log, logit, or untransformed) for each covariate by fitting separate linear regressions with each version of the transformed covariate as a predictor and an outcome of logit CFR. Transformation was selected based on the corresponding model with the lowest Akaike information criterion (AIC) score. Then, to improve model stability, we standardized each transformed covariate by subtracting the mean of the transformed covariate and dividing by the standard deviation.

### Model fitting

Some studies presented measles deaths aggregated into very large age bands. This may bias results if mortality is higher in the lower end of the age band. To reduce this bias, we fit the model in data in two stages. First, we fit a model to only data for which there was age granularity representing groupings five years wide or smaller; data used in this model included 0- to 34-year-olds. This model used the age-granular data and transformed and standardized covariate values for each study midpoint year and fit a Bayesian fixed-effects meta-regression model^14^ with the outcome variable as the logit CFR; for details on model selection see Supplementary Information Section 2. We computed standard error in logit space per study using the delta method transformation^15^ and used these values as weights in the meta-regression. To represent the relationship between logit CFR and age, we used a quadratic spline with 5 knots, with 3 internal knots, placed uniformly based on data density (i.e., equal proportions of input data represented between each knot) resulting in internal knots placed at ages 0.68, 1.31, and 3.83 years. Next, we split cases and deaths from each input data source reporting age bins wider than 1 year differentially based on estimates of country- and age-specific incidence^11^ and the overall relative age pattern of CFR estimated in the first stage model. We then recalculated logit of CFR and standard error per the newly adjusted number of deaths and cases per new granular age group.

Then, in a second stage model, we used the same general model formula previously described, maintaining the spline knot locations identified in the first stage model, and fit our outcome of logit CFR to all data following age-splitting (Supplementary Information Section 3). To ensure the correct direction of association between each covariate and CFR, defined as the direction described previously^7^, we placed priors on each regression coefficient. We generated 1000 samples of the regression coefficients from their fitted joint posterior distribution and predicted country- and age-specific measles CFR in LMICs from 1990 to 2019. We assumed CFR varied up to age 34 (the maximum age for which we had age-specific data) and held CFR constant for older ages.

To understand the effect of modelling changes, adding new covariates, and updating our dataset on our estimates of CFR relative to those produced by Portnoy and colleagues^5^, we conducted a decomposition analysis comparing this new modelling framework to Portnoy and colleagues’ (Supplementary Information Section 4). Additionally, we computed in-sample and five-fold out-of-sample cross validation metrics to assess model performance. We produced mean estimates of CFR at the age-, region-, or year-level by using the case-weighted average of age-, country-, year-specific CFR estimates. We performed all analyses and produced all figures within the R computing environment (version 5.4).

## Results

From our literature search, we identified 2581 articles for review. Following our inclusion and exclusion criteria, we extracted information on measles cases and deaths from 245 studies (Figure 1). A full list of source-specific extractions can be found in Supplementary Information Table 2. One hundred seventy-five of the sources were from community-based settings, 66 were from hospital-based settings, and four contained observations from both community- and hospital-based settings. One hundred twenty-six studies contained granular information on age (i.e., at least one age group less than or equal to five years wide) and 67 studies only presented information without age granularity. Among all sources, 57 overall unique age groups were represented. Eighty-eight sources provided information on laboratory confirmation of cases and 84 sources provided information on a definition for a measles-related death.

**Table 2.** Case-weighted CFR by region, year, and setting (community- versus hospital-based), with 95% confidence interval, for no-vaccination scenario.

|  | 1990 |  | 2000 |  | 2010 |  | 2019 |  |
| --- | --- | --- | --- | --- | --- | --- | --- | --- |
|  | Community-based | Hospital-based | Community-based | Hospital-based | Community-based | Hospital-based | Community-based | Hospital-based |
| North Africa and Middle East | 5.04%<br>(4.66 – 5.48%) | 18.26%<br>(17.03 – 19.65%) | 4.51%<br>(4.19 – 4.90%) | 16.45%<br>(15.39 – 17.67%) | 3.52%<br>(3.25 – 3.82%) | 13.25%<br>(12.33 – 14.29%) | 1.83%<br>(1.68 – 1.99%) | 7.19%<br>(6.65 – 7.79%) |
| Sub-Saharan Africa | 5.32%<br>(5.04 – 5.62%) | 19.03%<br>(18.30 – 19.87%) | 4.15%<br>(3.93 – 4.41%) | 15.41%<br>(14.73 – 16.17%) | 3.67%<br>(3.46 – 3.92%) | 13.79%<br>(13.07 – 14.65%) | 2.77%<br>(2.59 – 2.96%) | 10.77%<br>(10.16 – 11.51%) |
| Central Europe, Eastern Europe, and Central Asia | 1.97%<br>(1.78 – 2.14%) | 7.97%<br>(7.31 – 8.61%) | 0.56%<br>(0.50 – 0.63%) | 2.39%<br>(2.14 – 2.67%) | 0.37%<br>(0.33 – 0.42%) | 1.57%<br>(1.39 – 1.77%) | 0.33%<br>(0.29 – 0.38%) | 1.42%<br>(1.25 – 1.62%) |
| South Asia | 4.50%<br>(4.25 – 4.76%) | 16.88%<br>(16.03 – 17.79%) | 3.42%<br>(3.20 – 3.65%) | 13.21%<br>(12.46 – 14.05%) | 2.49%<br>(2.31 – 2.67%) | 9.89%<br>(9.25 – 10.63%) | 1.61%<br>(1.49 – 1.74%) | 6.57%<br>(6.11 – 7.11%) |
| Latin America and Caribbean | 2.75%<br>(2.48 – 3.01%) | 10.80%<br>(9.84 – 11.84%) | 1.88%<br>(1.68 – 2.07%) | 7.58%<br>(6.81 – 8.41%) | 1.24%<br>(1.08 – 1.40%) | 5.12%<br>(4.50 – 5.75%) | 0.88%<br>(0.77 – 1.01%) | 3.66%<br>(3.18 – 4.16%) |
| Southeast Asia, East Asia, and Oceania | 1.64%<br>(1.52 – 1.78%) | 6.68%<br>(6.21 – 7.21%) | 1.16%<br>(1.06 – 1.26%) | 4.79%<br>(4.40 – 5.21%) | 0.83%<br>(0.75 – 0.91%) | 3.45%<br>(3.13 – 3.79%) | 0.75%<br>(0.68 – 0.83%) | 3.15%<br>(2.86 – 3.47%) |

There were 44 studies with information on measles cases and deaths before 1980, 119 from 1980–1989, 84 from 1990–1999, 67 from 2000–2009, and 71 from 2010–2019. Seventy-five countries were represented among sources. Among 3,477,901 measles cases represented across all observations, the crude mean CFR was 6.3% and median CFR was 2.2%. Among hospital-based studies, the crude mean CFR was 8.9%, and among community-based studies the crude mean CFR was 5.1%.

In 1990, our model estimated the mean community-based CFR across all locations to be 2.60% (95% Uncertainty Interval (UI): 2.52 − 2.69%), and in 2019, the mean modeled estimate was 1.32% (95% UI: 1.28 − 1.36%). Among hospital-based settings, the mean estimated CFR across all locations in 1990 was 10.13% (95% UI: 9.67 – 10.60%), and in 2019 was 5.35% (95% UI: 5.08 – 5.64%). In all regions, estimated measles CFRs decreased from 1990 to 2019 in both community- and hospital-based settings (Table 1). Across all regions, CFRs were estimated to be highest in the sub-Saharan Africa region in 2019 in both community- and hospital-based settings.

**Table 1.**
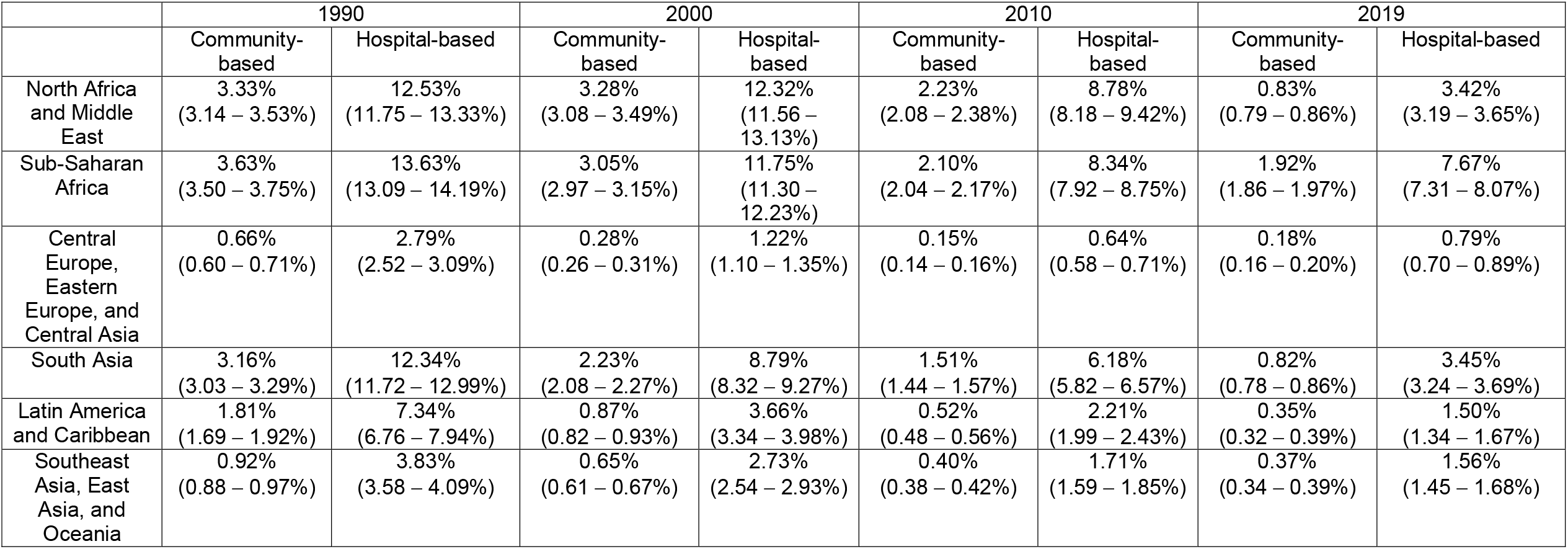
Case-weighted CFR by region, year, and setting (community- versus hospital-based), with 95% confidence interval.

The median country-specific case-weighted CFR estimates as well as the range in CFR estimates by country decreased across the study period (Figure 2). CFR decreased in all estimated LMICs from 1990 to 2019. As mean all-age estimates of CFR have been case-weighted, country- and year-specific means were influenced by the underlying distribution of the ages of cases within that specific country and year; a relative distribution of the ages of cases is shown in Supplementary Figure 6. Age-standardized CFR estimates, which show that the declining CFR trends persist after age-standardization, can be found in Supplementary Information Section 5. Country-specific CFR results can be found in Supplementary Information Appendix 2, and validation metrics can be found in Supplementary Tables 8–9.

**Figure 2.**
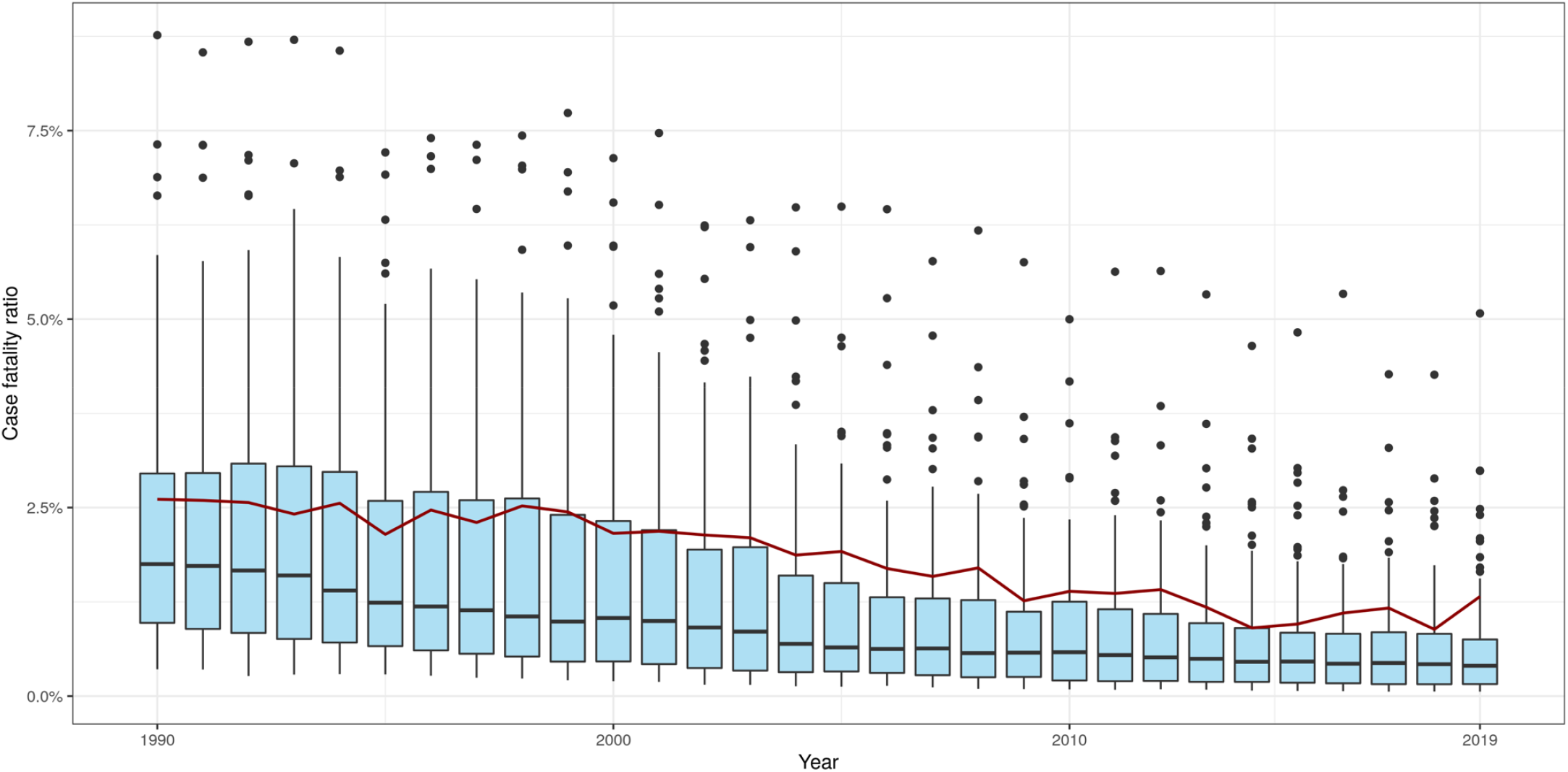
Box plots of country-specific, community-based case fatality ratios (CFRs) by year, with case-weighted LMIC mean CFR value by year (red line).

We estimated CFR to be highest among infants and to decline monotonically as age increased (Figure 3a). This general trend was consistent across regions (Figure 3b) and over time. In 2019 across LMICs in community-based settings, we estimated that CFR among under-one-year-olds was 3.03% (95% UI: 2.89 – 3.16%), 1.63% (95% UI: 1.58 – 1.68%) among 1- to 4-year-olds, 0.84% (95% UI: 0.80 – 0.87%) among 5- to 9-year-olds, and 0.67% (95% UI: 0.64 – 0.70%) among 10- to 14-year-olds. Among LMICs in 2019 in hospital-based settings, CFR among under-one-year-olds was 5.33% (95% UI: 5.06 – 5.59%), among 1- to 4-year-olds was 2.80% (95% UI: 2.70 – 2.90%), among 5- to 9-year-olds was 1.50% (95% UI: 1.44 – 1.57%), and among 10- to 14-year-olds was 0.87% (95% UI: 0.83 – 0.91%).

**Figure 3a.**
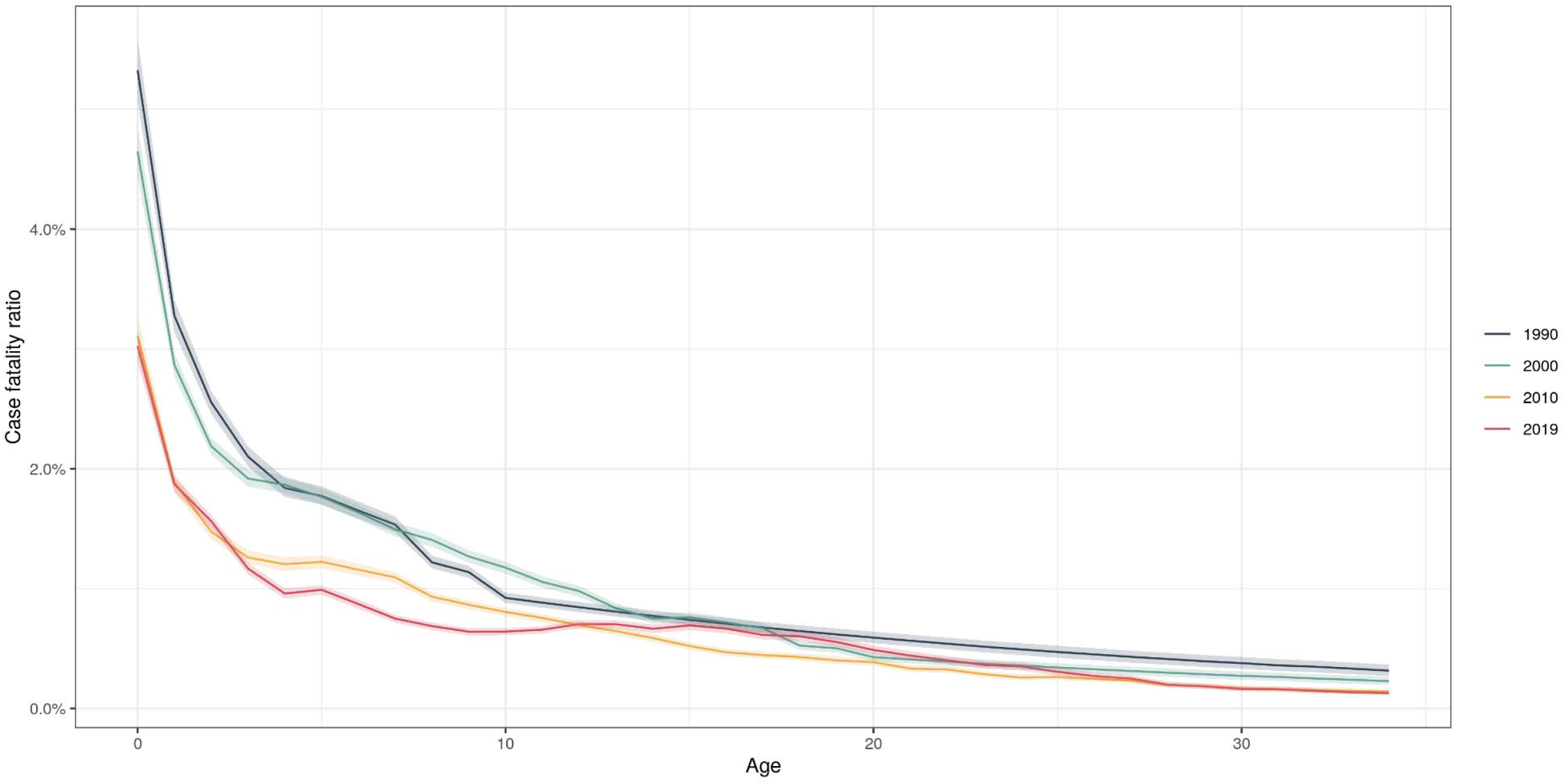
Age-specific, community-based, case-weighted case fatality ratio (CFR) estimates among 0–14-year-olds, by years 1990, 2000, 2010, and 2019 for low- and middle-income countries.

**Figure 3b.**
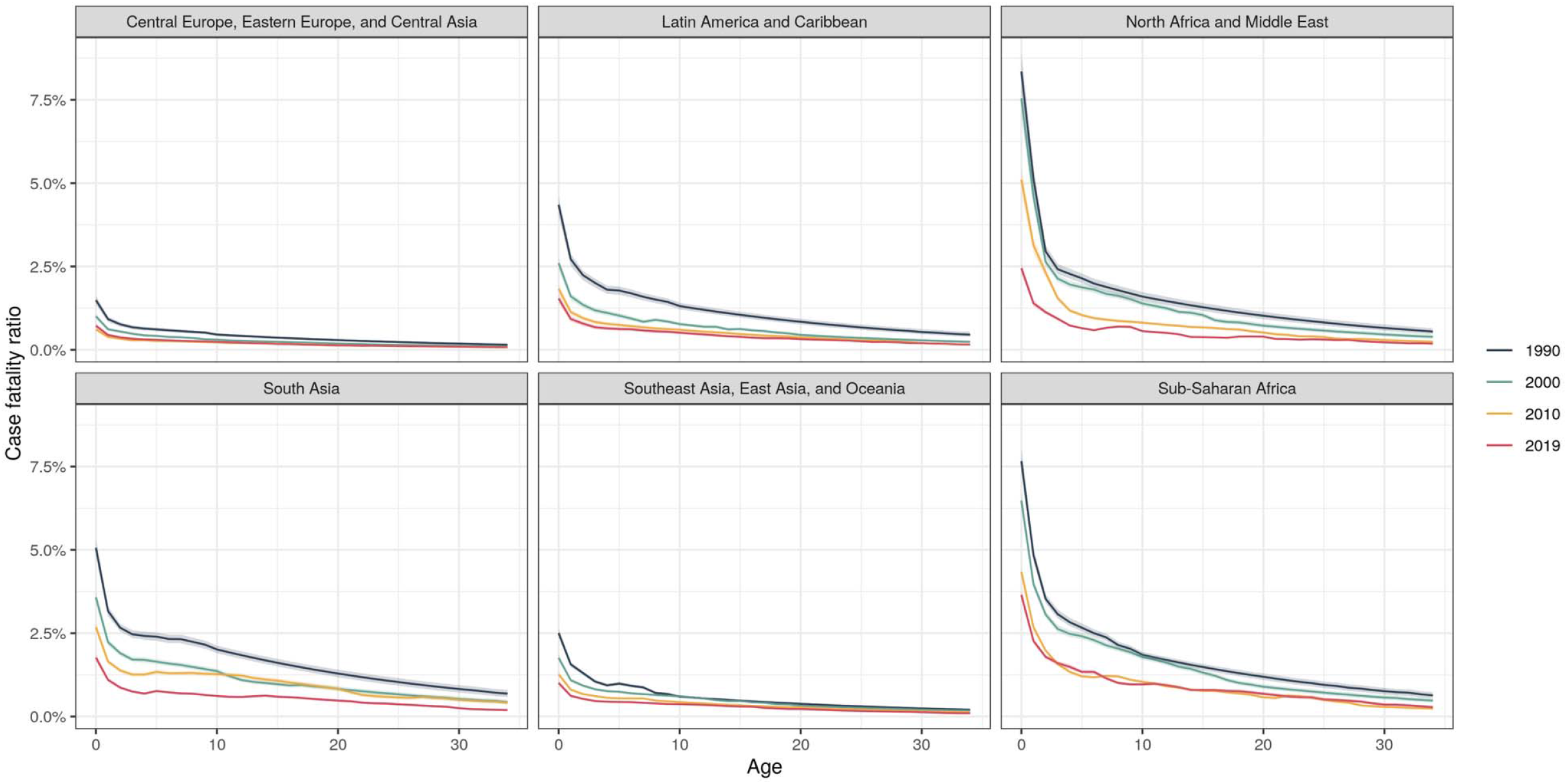
Age-specific, community-based, case-weighted case fatality ratio (CFR) (CFR) estimates among 0–14-year-olds, by years 1990, 2000, 2010, and 2019 by region.

In addition to our baseline scenario, we predicted results for a scenario in which there had been no vaccine introduction (i.e., MCV1 coverage was 0% in all countries and years, and incidence values also reflect a lack of vaccination). Methods for estimating incidence in a no-vaccination scenario have been described at length elsewhere.^16^ CFR estimates by region, year, and community- and hospital-based settings are larger relative to the baseline vaccination scenario (Table 2). As a result of these differences in CFR and measles incidence in both scenarios, we estimated that from years 1990 to 2019 there have been approximately 71 million deaths averted that are attributable to measles vaccination in these LMICs.

## Discussion

Until 2019, there was only a single systematic review of measles CFR, which was limited to community-based settings and did not examine changes in CFR over time.^3^ In 2019, an updated systematic review expanded literature to additionally include CFRs among hospital-based settings, and used a time-varying model to estimate CFR by location and year.^5^ While more comprehensive, this updated review did not include age-specificity beyond variation among under- and over 5-year-olds, and covariates included were not selected via a transparent, systematic process. To address these shortcomings, our new study: (1) updated the former literature searches; (2) based covariate selection on widespread exert consultation, a literature review, and selection through a statistical process; and (3) accounted for the distribution of age in the modelling process. Our study included 40 new sources on measles CFR from 21 new countries up to 2019. We statistically tested new covariates representing known relations to measles CFR for inclusion such as vitamin A deficiency prevalence, wasting prevalence, HIV prevalence and mortality rate due to war and terrorism.

We estimated CFR in community-based settings in 1990 to be 2.60% and to have declined to 1.32% by 2019. We estimated higher CFRs in hospital-based settings relative to community-based settings, consistent with previous findings^5^ likely observing higher severity cases requiring hospitalization. We estimated infants to have the highest CFRs. In this age group, the risk for infection with measles is influenced by the persistence of maternal antibodies, which in turn depends on gestational age and underlying maternal immunity rates.^8^ On an individual level, the presence of maternal antibodies may also mitigate the severity of measles infection, potentially leading to lower CFRs. In our analysis, after controlling for study-level covariates, our model suggests that population-level CFR decreases monotonically with age, consistent with previous studies.^17^ This may be because infants who acquire measles do not have sufficient maternal antibodies to prevent infection. More detailed and robust data collection in these youngest ages will be critical for assessing this relationship further.

This study has several limitations. First, we did not include studies representing special populations, including some that might be particularly vulnerable to both measles infection and also increased case fatality such as refugees and internally displaced persons. These populations should not be neglected in the global measles control agenda; however, at this time data on these subpopulations is too sparse to accurately assess their current situation. Next, we assumed that the case fatality estimates presented in each study were nationally representative, which may have biased the relationship between CFR and national-level covariates.

Additionally, we assumed that the age distribution of measles cases represented in studies without reported age-specificity followed the same relative age distribution of cases estimated nationally in that country and year during our age-splitting process.

Also, we were constrained by the limited number of studies reporting information on both laboratory confirmation of measles cases and a definition of what was considered a death attributable to measles. Therefore, we chose to include all available studies in the analysis regardless of their reporting on case confirmation or death definition to avoid introducing a compositional bias, stemming from differences in study level demographics, in our estimation framework.

Next, we were not able to incorporate uncertainty from our covariates, including measles incidence, and model specification in our modeling framework, and as such, our uncertainty estimates only reflect uncertainty in the CFR modeling process itself without any additional factors. Additionally, for all covariates that passed our statistical analysis checks, we included each in our modeling framework with priors to govern the direction of association estimated by the model. In this process, four covariates (mortality rate due to war and terrorism, wasting prevalence, HIV prevalence, and GDP per capita) no longer contributed significantly to our model, so they were removed. It is important to emphasize that these covariates may still be significantly related to measles CFR; their exclusion was a result of the underlying collinearity of our covariates that suggested little added predictive benefit to their inclusion in the final model. Lastly, we did not examine individual-level relationships between measles CFR and the covariates included in our modeling framework but rather assessed population-level trends for use in population-level modeling. Therefore, the presented associations between covariates and CFR should not be considered causal relationships.

Our study improves upon previous estimates of measles CFR by incorporating new data sources, systematically identifying covariates, and including improved age-specific variation.

These estimates and methodology will be essential in future assessment of measles mortality and vaccination programmes by global- and country-level decision makers.

## Data Availability

All data produced in the present study are available upon reasonable request to the authors and will be published publicly at time of future publication in journal.

## Acknowledgements

We thank Natasha Crowcroft, Felicity Cutts, Emily Dansereau, Deepa Gamage, Katy Gaythorpe, Katrina Kretsinger, Kevin McCarthy, Mark Papania, and Niket Thakkar for their participation in a working group of experts and for the Immunization and Vaccines related Implementation Research advisory committee (IVIR-AC) for their valuable comments on multiple versions of this work. A.N.S, J.F.M., and A.P. received funding for this work from the Bill & Melinda Gates Foundation (INV-019383). A.N.S. received additional support from the National Institutes of Health (F31AI167535). The content of this work is the sole responsibility of the authors and does not represent official views of the National Institutes of Health. M.J. was supported by Gavi, the Vaccine Alliance and the Bill & Melinda Gates Foundation through the Vaccine Impact Modelling Consortium.

## References

1. Patel MK, Goodson JL, Alexander Jr JP, et al. Progress Toward Regional Measles Elimination — Worldwide, 2000–2019. MMWR Morb Mortal Wkly Rep 2020; 69(45): 1700–5.

2. World Health Organization. Reducing global measles mortality. 2003.

3. Wolfson LJ, Grais RF, Luquero FJ, Birmingham ME, Strebel PM. Estimates of measles case fatality ratios: a comprehensive review of community-based studies. International Journal of Epidemiology 2009; 38(1): 192–205.

4. Wolfson LJ, Grais RF, Luquero FJ, Birmingham ME, Strebel PM. Estimates of measles case fatality ratios: a comprehensive review of community-based studies. Int J Epidemiol 2009; 38(1): 192–205.

5. Portnoy A, Jit M, Ferrari M, Hanson M, Brenzel L, Verguet S. Estimates of case-fatality ratios of measles in low-income and middle-income countries: a systematic review and modelling analysis. Lancet Glob Health 2019; 7(4): E472–E81.

6. Portnoy A, Hsieh YL, Abbas K, et al. Differential health impact of intervention programs for time-varying disease risk: a measles vaccination modeling study. BMC Medicine 2022; 20.

7. Sbarra AN, Jit M, Mosser JF, et al. Population-level risk factors related to measles case fatality: a conceptual framework and systematic review of evidence. medRxiv 2022.

8. Guerra FM, Crowcroft NS, Friedman L, et al. Waning of measles maternal antibody in infants in measles elimination settings - A systematic literature review. Vaccine 2018; 36(10): 1248–55.

9. Goodson JL, Masresha BG, Wannemuehler K, Uzicanin A, Cochi S. Changing Epidemiology of Measles in Africa. Journal of Infectious Diseases 2011; 204: S205–S14.

10. Vos T, Lim SS, Abbafati C, et al. Global burden of 369 diseases and injuries in 204 countries and territories, 1990–2019: a systematic analysis for the Global Burden of Disease Study 2019. The Lancet 2020; 396(10258): 1204–22.

11. Simons E, Ferrari M, Fricks J, et al. Assessment of the 2010 global measles mortality reduction goal: results from a model of surveillance data. The Lancet 2012; 379(9832): 2173–8.

12. WHO. WHO/UNICEF coverage estimates for 1980-2021. 2022. https://www.who.int/data/gho/data/themes/immunization (accessed July 15 2022).

13. World Bank. World Development Indicators. 2022. https://databank.worldbank.org/source/world-development-indicators (accessed July 15 2022).

14. Zheng P, Barber R, Sorensen RJ, Murray CJ. Trimmed constrained mixed effects models: formulations and algorithms. Journal of Computational and Graphical Statistics 2021; 30(3): 1–13.

15. Levy Y. The delta method and its implementation in R. 2019. https://www.r-bloggers.com/2019/03/the-delta-method-and-its-implementation-in-r/ (accessed August 2022.

16. Toor J, Echeverria-Londono S, Li X, et al. Lives saved with vaccinatioon for 10 pathogens across 112 countries in a pre-COVID-19 world. eLife 2021; 10: e67635.

17. Aaby P, Martins CL, Garly M, Rodrigues A, Benn CS, Whittle H. The optimal age of measles immunisation in low-income countries: a secondary analysis of the assumptions underlying the current policy. BMJ Open 2012; 2.

