## Supplementary Information 1 for "Estimating national-level measles case fatality ratios: an updated systematic review and modelling study"

### ***Section 1.*** *Covariate selection via statistical analysis*

***Section 1.1.*** *Additional details on covariate interpolation*

The following covariate sets did not require interpolation or use of regional values: education, maternal education, war rate due to mortality and terrorism, health access and quality index, universal health coverage, sociodemographic index, stunting prevalence, wasting prevalence, underweight prevalence, vitamin A deficiency prevalence, HIV prevalence, and MCV2 coverage. For 12 countries, we interpolated covariate values for GDP per capita; we also used regional values in 23 countries. For 7 countries, we interpolated covariate values for under-5 mortality rate; we also used regional values in 2 countries. We used regional covariate values in 6 countries for total fertility rate. We used regional covariate values in 14 countries for MCV1 coverage. For 1 country, we interpolated covariate values for proportion living in urban settings; we also used regional values in 2 countries.

***Section 1.2.*** *Test for collinearity per underlying mechanism*

For the underlying mechanism of “health care access and care seeking”, we tested covariate sets for education, maternal education, proportion living in an urban setting, and mortality rate due to war and terrorism. Education was correlated with maternal education; correlation coefficients shown below. As education was more correlated with the other covariates relative to maternal education, it was removed from further analysis. Covariates moving on to the second step of data analysis for the “health care access and care seeking” mechanism were: maternal education, proportion living in an urban setting, and mortality rate due to war and terrorism.

|  | Education | Maternal education | Prop. living in urban setting | War mortality rate |
| --- | --- | --- | --- | --- |
| Education | 1.0 | 0.9965 | 0.6539 | -0.0648 |
| Maternal education |  | 1.0 | 0.6523 | -0.0645 |
| Prop. living in urban setting |  |  | 1.0 | -0.537 |
| War mortality rate |  |  |  | 1.0 |

For the underlying mechanism of “health care quality”, we tested covariate sets for under-5 mortality rate, health access and quality index, universal health coverage, GDP per capita, and sociodemographic index. Under-5 mortality rate, health access and quality index, universal health coverage and sociodemographic index were all correlated with each other; correlation coefficients shown below. Health access and quality index, universal health coverage, and sociodemographic index were more correlated with the other covariates relative to under-5 mortality rate, and so they were removed from further analysis. Covariates moving on to the second step of data analysis for the “health care quality” mechanism were: under-5 mortality rate, and GDP per capita.

|  | Under-5 mortality rate | Health access and quality index | Universal health coverage | GDP per capita | Sociodemographic Index |
| --- | --- | --- | --- | --- | --- |
| Under-5 mortality rate | 1.0 | -0.8027 | -0.8346 | -0.3920 | -0.8525 |
| Heath access and quality index |  | 1.0 | 0.9916 | 0.6428 | 0.9365 |
| Universal health coverage |  |  | 1.0 | 0.6311 | 0.9417 |
| GDP per capita |  |  |  | 1.0 | 0.6159 |
| Sociodemographic Index |  |  |  |  | 1.0 |

For the underlying mechanism of “nutritional status”, we tested covariate sets for stunting prevalence, wasting prevalence, underweight prevalence, and vitamin A deficiency prevalence. Stunting prevalence, underweight prevalence, and wasting prevalence were correlated with each other; correlation coefficients shown below. Stunting prevalence, and underweight prevalence were more correlated with the other covariates relative to wasting prevalence, and so they were removed from further analysis. Covariates moving on to the second step of data analysis for the “nutritional status” mechanism were: wasting prevalence and vitamin A deficiency prevalence.

|  | Stunting | Wasting | Underweight | Vitamin A deficiency |
| --- | --- | --- | --- | --- |
| Stunting | 1.0 | 0.7597 | 0.8970 | 0.6965 |
| Wasting |  | 1.0 | 0.8927 | 0.5687 |
| Underweight |  |  | 1.0 | 0.6509 |
| Vitamin A deficiency |  |  |  | 1.0 |

For the underlying mechanism of “risk of secondary infection”, we tested covariate sets for HIV prevalence and total fertility rate. The covariates were not correlated with each other; correlation coefficient shown below. Both covariates moved on to the second step of data analysis for the “risk of secondary infection” mechanism.

|  | HIV prevalence | TFR |
| --- | --- | --- |
| HIV prevalence | 1.0 | 0.2279 |
| TFR |  | 1.0 |

For the underlying mechanism of “measles control and epidemiology”, we tested covariate sets for MCV1 and MCV2 coverage. The covariates were not correlated with each other; correlation coefficient shown below. Both covariates moved on to the second step of data analysis for the “measles control and epidemiology” mechanism.

|  | MCV1 coverage | MCV2 coverage |
| --- | --- | --- |
| MCV1 coverage | 1.0 | 0.4713 |
| MCV2 coverage |  | 1.0 |

***Section 1.3.*** *Test for predictive capacity per underlying mechanism*

For the mechanism of “health care access and care seeking”, no covariates tested had p-values greater than 0.3 (see below). Therefore, all remaining covariates (maternal education, proportion living in urban setting, and mortality rate due to war and terrorism) were kept as covariate sets for the remainder of the analysis.

|  | Estimate | P-value |
| --- | --- | --- |
| Intercept | 0.0876 | < 2 e-16 |
| Maternal education | -0.0039 | 0.0192 |
| Prop urban | -0.0435 | 0.1163 |
| War mortality rate | 13.15116 | 0.1161 |

For the mechanism of “health care quality”, no covariates tested had p-values greater than 0.3 (see below). Therefore, all remaining covariates (under-5 mortality rate and GDP per capita) were kept as covariate sets for the remainder of the analysis.

|  | Estimate | P-value |
| --- | --- | --- |
| Intercept | -1.441e-04 | 0.9853 |
| Under-5 mortality rate | 4.097e-04 | 1.67e-10 |
| GDP per capita | 7.766e-07 | 0.0386 |

For the mechanism of “nutritional status”, no covariates tested had p-values greater than 0.3 (see below). Therefore, all remaining covariates (wasting prevalence and vitamin A deficiency prevalence) were kept as covariate sets for the remainder of the analysis.

|  | Estimate | P-value |
| --- | --- | --- |
| Intercept | 0.0097 | 0.192 |
| Wasting | 0.0901 | 0.273 |
| Vitamin A deficiency | 0.0767 | 0.130 |

For the mechanism of “risk of secondary infection”, no covariates tested had p-values greater than 0.3 (see below). Therefore, all remaining covariates (HIV prevalence and total fertility rate) were kept as covariate sets for the remainder of the analysis.

|  | Estimate | P-value |
| --- | --- | --- |
| Intercept | -1.540e-02 | 0.1686 |
| HIV prevalence | 2.675e-01 | 0.2462 |
| TFR | 8.409e-03 | 0.0003 |

For the mechanism of “measles control and epidemiology”, MCV2 coverage had a p-value greater than 0.3 (see below). Therefore, MCV1 coverage was the only covariate sets kept for the remainder of the analysis.

|  | Estimate | P-value |
| --- | --- | --- |
| Intercept | 0.1160 | <2 e-16 |
| MCV1 coverage | -0.1078 | 1.52e-14 |
| MCV2 coverage | 0.0021 | 0.863 |

### ***Section 2.*** *Model selection*

***Section 2.1.*** *First stage model with age granular data*

We analyzed the relationship between age and CFR in reported studies with age-specific data both with and without controlling for other covariates. There was a consistent relationship between covariate values and CFR values, particularly for measles incidence and MCV1 coverage (Supplementary Figures 4-5). Taken together, these suggested that the relationship between age and CFR was confounded by these other covariates, and therefore we elected to adjust for other covariates in our first-stage model.

***Section 2.2.*** *Knot selection*

We ran both first and second stage models with both 4 knots (with 2 internal) and 5 knots (with 3 internal) placed uniformly on data density and selected the best performing model based on the lowest Akaike information criterion (AIC) score among results from the second stage model. This process selected the model with 5 knots (AIC: 174907) instead of 4 knots (AIC: 175086).

***Section 2.3.*** *Inclusion of random effects*

We additionally tested the inclusion of random effects in our second stage model, by testing a random effect placed on each study. This approach caused the coefficient for the community versus hospital-setting indicator to become 0, with a non-significant p-value (p-value=1). Because we know these sets of studies (i.e. those from community-based settings and those from hospital-based settings) were collected from different underlying populations with known difference in measles severity, we elected to use a model without the inclusion of random effects.

### ***Section 3.*** *Final model structure*

Our final first stage CFR model (that only uses age specific input data) follows the following structure. Using transformed and standardized covariate values for each study midpoint year, we fit a Bayesian fixed-effects meta-regression model^1^ with the outcome variable of the logit of CFR. We computed standard error in logit space per study using the delta transformation and used these values as weights in the meta-regression. Before transforming to logit space, CFR ratios equalling 0 were offset to 0.0002202378 and ratios equal to 1 were offset to 0.9999999999999.

Our regression equation is as follows:

$$y_{i}=X_{i}\left( \beta\right)+ \epsilon_{i}$$

$$\epsilon_{i}\sim N(0,\Lambda)$$

where $y_{i}$ is the vector of observations of logit of CFR from the $i$^th^ study, $X_{i}$is a vector of covariates paired with each data observation, $\beta$ are regression coefficients, and $\epsilon_{i}$ are measurement errors with a given covariance $\Lambda$. For age, our $\beta$ coefficient is represented as a quadratic spline with 5 knots (3 internal) placed uniformly based on data density. We included a prior to ensure a right linear tail on our quadratic spline function.

Our final second stage model (that uses all data) is as follows. Model specifications are identical to the first stage as previously defined, except with the following additional priors:

$$\beta_{community indicator} \sim Uniform(-\infty, 0)$$

$$\beta_{incidence} \sim Uniform(0, \infty)$$

$$\beta_{mortality rate due to war and terrorism} \sim Uniform(0, \infty)$$

$$\beta_{maternal education} \sim Uniform(-\infty, 0)$$

$$\beta_{GDP per capita} \sim Uniform(-\infty, 0)$$

$$\beta_{HIV prevalence} \sim Uniform(0, \infty)$$

$$\beta_{MCV1 coverage} \sim Uniform(-\infty, 0)$$

$$\beta_{Total fertility rate} \sim Uniform(0, \infty)$$

$$\beta_{Under 5 mortality rate} \sim Uniform(0, \infty)$$

$$\beta_{Proportion living in urban setting} \sim Uniform(0, \infty)$$

$$\beta_{Vitamin A deficiency prevalence} \sim Uniform(0, \infty)$$

$$\beta_{Wasting prevalence} \sim Uniform(0, \infty)$$

### ***Section 4.*** *Decomposition analysis for validating changes to model structure, covariates, and input data*

To increase the robustness and rigor of measles CFR modeling, we considered various updates to the model structure, covariates, and input data sources relative to the model previously published by Portnoy et al.^2^ With updates to each component (model structure, covariates, and input data), we tracked the overall change in model performance to ensure updates were statistically beneficial in the estimation of measles CFR. Specific steps and validation at each step are described in each subsequent section.

***Section 4.1.*** *First stage, updates to model structure*

We made the following sequential adaptations to the log-linear model published previously:

- Model 0: Generalized linear model, with log link and cases as weights
- Model 1.A: Generalized linear model, with log(CFR) as outcome and cases as weights
- Model 1.B: Generalized linear model, with logit(CFR) as outcome and cases as weights
- Model 1.C: Bayesian meta-regression, with logit(CFR) as outcome and standard error as weights

The structure of Model 0 is identical to the model previously published^2^, and serves as our baseline. In order to more accurately represent CFR as a ratio bounded between 0 and 1, we first removed the log link from the model and instead log (Model 1.A) then logit (Model 1.B) transformed CFR as our outcome. In order to best capture the underlying uncertainty from the data, we then implemented a Bayesian meta-regression framework using standard errors as weights (Model 1.C). We compared both in- and out-of-sample validation for each model iteration. Model 1.C performed best among both in- and out-of-sample validation exercises across metrics (Supplementary Tables 4-5) yielding generally lower root mean squared error (RMSE), mean error and man absolute error and higher correlation.

***Section 4.2.*** *Second stage, updates to covariates*

We made the following sequential adaptations to the best model (previously referred to as Model 1.C) from our first decomposition step:

- Model 1: Previously described Model 1.C with original covariates and original data inputs
- Model 2: Previously described Model 1.C with updated covariates and original data inputs

We compared the best model using original covariates and data inputs to a new model fit using the updated covariate set. We compared the performance of these two models to the original model version (Model 0) in Supplementary Tables 6-7. Model 2 performed best across most in- and out-of-sample validation metrics yielding generally lower root mean squared error (RMSE), mean error and mean absolute error and higher correlation.

***Section 4.3.*** *Third stage, updates to input data sources*

We made the following sequential adaptations to the best model from our second decomposition step updates:

- Model 2: Previously described Model 1.C with updated covariates and original data inputs
- Model 3: Previously described Model 1.C with updated covariates and updated data inputs

There were 40 additional new studies added across 21 additional countries. Because the input data sources were changing, we did not compare validation metrics to previous decomposition steps. Full model validation can be found in Supplementary Tables 8-9.

### ***Section 5.*** *Supplementary Results*

***Section 5.1.*** *Age-standardized results*

Because the age distribution of cases within a country impacts the ability to compare trends across locations, we also computed country-specific age-standardized CFRs using a reference population of the global age pattern of cases from 1990 as well as the general population age distribution from the UN in 1990 (Supplementary Figure 7). Age-standardized estimates of CFR allow users to more directly compare estimates across locations and years.

***Section 5.2.*** *Sensitivity analyses*

We ran sensitivity analyses to investigate the implications of using all studies regardless of if they provided information on laboratory confirmation of cases or a definition of a death attributable to measles. Generally, studies that reported information on laboratory confirmation of cases were from countries and years with lower measles incidence, higher MCV1 coverage, and lower CFRs relative to studies that did not report information on laboratory confirmation (Supplementary Figures 8-10). Additionally, studies reporting definitions of deaths attributable to measles were most often from hospital-based settings rather than community-based settings (Chi-squared p-value < 0.001). In a sensitivity analysis excluding first studies without information on laboratory confirmation of cases and then following an additional sensitivity analysis excluding studies without information on a death definition, we were estimating systematically lower CFRs than when including all studies in our model (Supplementary Figures 11-12).

### ***Section 5.*** *Supplementary Tables*

#### **Supplementary Table 1.** GATHER compliance checklist.

| **Item number** | **Checklist item** | **Reported on page number(s):** |
| --- | --- | --- |
| **Objectives and funding** | | |
| 1 | Define the indicator(s), populations (including age, sex, and geographic entities), and time period(s) for which estimates were made. | Methods |
| 2 | List the funding sources for the work. | Acknowledgements |
| **Data inputs** | | |
| *For all data inputs from multiple sources that are synthesised as part of the study:* | | |
| 3 | Describe how the data were identified and how the data were accessed. | Methods |
| 4 | Specify the inclusion and exclusion criteria. Identify all ad hoc exclusions. | Methods |
| 5 | Provide information on all included data sources and their main characteristics. For each data source used, report reference information or contact name/institution, population represented, data collection method, year(s) of data collection, sex and age range, diagnostic criteria or measurement method, and sample size, as relevant. | Supplementary Table 2 |
| 6 | Identify and describe any categories of input data that have potentially important biases (e.g., based on characteristics listed in item 5). | Methods; Discussion |
| *For data inputs that contribute to the analysis but were not synthesised as part of the study:* | | |
| 7 | Describe and give sources for any other data inputs. | Methods |
| *For all data inputs:* | | |
| 8 | Provide all data inputs in a file format from which data can be efficiently extracted (e.g., a spreadsheet rather than a PDF), including all relevant meta-data listed in item 5. For any data inputs that cannot be shared because of ethical or legal reasons, such as third-party ownership, provide a contact name or the name of the institution that retains the right to the data. | Supplementary Table 2 |
| **Data analysis** | | |
| 9 | Provide a conceptual overview of the data analysis method. A diagram might be helpful. | Supplementary Figure 1 |
| 10 | Provide a detailed description of all steps of the analysis, including mathematical formulae. This description should cover, as relevant, data cleaning, data pre-processing, data adjustments and weighting of data sources, and mathematical or statistical model(s). | Methods; Supplementary Information Sections 1-3 |
| 11 | Describe how candidate models were evaluated and how the final model(s) were selected. | Methods; Supplementary Information Sections 2 |
| 12 | Provide the results of an evaluation of model performance, if done, as well as the results of any relevant sensitivity analysis. | Methods; Results; Supplementary Tables 8-9 |
| 13 | Describe methods for calculating uncertainty of the estimates. State which sources of uncertainty were, and were not, accounted for in the uncertainty analysis. | Methods |
| 14 | State how analytic or statistical source used to generate estimates can be accessed. | Methods |
| **Results and discussion** | | |
| 15 | Provide published estimates in a file format from which data can be efficiently extracted. | Results; Supplementary Appendix 2 |
| 16 | Report a quantitative measure of uncertainty of the estimates (e.g., uncertainty intervals). | Results |
| 17 | Interpret results in light of existing evidence. If updating a previous set of estimates, describe the reasons for changes in estimates. | Discussion; Research in Context |
| 18 | Discuss limitations that affect interpretation of the estimates. | Discussion |

#### **Supplementary Table 2.** Input data sources for final model.

| Citation | ISO3 | Midpoint Year | Community indicator | Minimum age (years) | Maximum age (years) |
| --- | --- | --- | --- | --- | --- |
| Arya LS, Azamy S, Ghani AR, Singh M. Outcome of measles in Afghanistan. Indian pediatrics. 1981 Feb;18(2):112-6. | AFG | 1978 | 0 | 0.4167 | 12 |
| Arya LS, Taana I, Tahiri C, Saidali A, Singh M. Spectrum of complications of measles in Afghanistan: a study of 784 cases. The Journal of tropical medicine and hygiene. 1987 Jun 1;90(3):117-22. | AFG | 1981 | 0 | 0.3333 | 12 |
| Choudhry VP, Atmar M, Amin I, Aram GN, Ghani R. Effect of protein energy malnutrition on the immediate outcome of measles. The Indian Journal of Pediatrics. 1987 Sep;54(5):717-22. | AFG | 1984 | 0 | 0 | 17 |
| Wakeham PF. Severe measles in Afghanistan. Journal of Tropical Pediatrics and Environmental Child Health. 1978;24(2):87-8. | AFG | 1971 | 1 | 0 | 99 |
| Chen RT, Weierbach R, Bisoffi Z, Cutts F, Rhodes P, Ramaroson S, Ntembagara C, Bizimana F. A ‘post-honeymoon period’ measles outbreak in Muyinga sector, Burundi. International journal of epidemiology. 1994 Feb 1;23(1):185-93. | BDI | 1988 | 1 | 0 | 5 |
| Kambiré C, Konde MK, Yaméogo A, Tiendrébéogo SR, Ouédraogo RT, Otten Jr MW, Cairns KL, Zuber PL. Measles incidence before and after mass vaccination campaigns in Burkina Faso. Journal of Infectious Diseases. 2003 May 15;187(Supplement_1):S80-5. | BFA | 2000 | 1 | 0 | 99 |
| Kidd S, Ouedraogo B, Kambire C, Kambou JL, McLean H, Kutty PK, Ndiaye S, Fall A, Alleman M, Wannemuehler K, Masresha B. Measles outbreak in Burkina Faso, 2009: a case–control study to determine risk factors and estimate vaccine effectiveness. Vaccine. 2012 Jul 13;30(33):5000-8. | BFA | 2009 | 1 | 0 | 99 |
| Sahuguède P, Roisin A, Sanou I, Nacro B, Tall F. Epidémie de rougeole au Burkina Faso: 714 cas hospitalisés à l'hôpital de Bobo-Dioulasso: étude des facteurs de risque. InAnnales de pédiatrie (Paris) 1989 (Vol. 36, No. 4, pp. 244-251). | BFA | 1986 | 0 | 0 | 18 |
| World Health Organization. Measles mortality reduction in West Africa, 1996-2002. Weekly Epidemiological Record= Relevé épidémiologique hebdomadaire. 2003;78(45):390-2. | BFA | 2002 | 1 | 0 | 99 |
| Bhuiya A, Wojtyniak B, D'Souza S, Nahar L, Shaikh K. Measles case fatality among the under-fives: a multivariate analysis of risk factors in a rural area of Bangladesh. Social science & medicine. 1987 Jan 1;24(5):439-43. | BGD | 1980 | 1 | 0 | 4 |
| Francisco AD, Fauveau V, Sarder AM, Chowdhury HR, Chakraborty J, Yunus MD. Measles in rural Bangladesh: issues of validation and age distribution. International journal of epidemiology. 1994 Apr 1;23(2):393-9. | BGD | 1989 | 1 | 0 | 5 |
| Fauveau V, Chakraborty J, Sarder AM, Khan MA, Koenig MA. Measles among under-9-month-olds in rural Bangladesh: its significance for age at immunization. Bulletin of the World Health organization. 1991;69(1):67. | BGD | 1980 | 1 | 0.5 | 99 |
| Koster FT, Curlin GC, Aziz KM, Haque A. Synergistic impact of measles and diarrhoea on nutrition and mortality in Bangladesh. Bulletin of the World Health Organization. 1981;59(6):901. | BGD | 1975 | 1 | 0 | 10 |
| Shahid NS, Clauquin P, Shaikh K, Zimicki S. Long-term complication of measles in rural Bangladesh. The Journal of Tropical Medicine and Hygiene. 1983 Apr 1;86(2):77-80. | BGD | 1980 | 1 | 0 | 1 |
| World Health Organization. Expanded Programme on Immunization: Public health importance of measles. Weekly Epidemiological Record. 1986;61(12):89-90. | BGD | 1984 | 1 | 0 | 99 |
| Tricou V, Pagonendji M, Manengu C, Mutombo J, Mabo RO, Gouandjika-Vasilache I. Measles outbreak in Northern Central African Republic 3 years after the last national immunization campaign. BMC Infectious Diseases. 2013 Dec;13(1):1-6. | CAF | 2011 | 1 | 0 | 99 |
| Aiqiang X, Zijian F, Wenbo X, Lixia W, Wanshen G, Qing X, Haijun S, Lee LA, Xiaofeng L. Active Case‐Based Surveillance for Measles in China: Lessons Learned from Shandong and Henan Provinces. Journal of Infectious Diseases. 2003 May 15;187(Supplement_1):S258-63. | CHN | 2001 | 1 | 0 | 53 |
| Ji Y, Zhang Y, Xu S, Zhu Z, Zuo S, Jiang X, Lu P, Wang C, Liang Y, Zheng H, Liu Y. Measles resurgence associated with continued circulation of genotype H1 viruses in China, 2005. Virology Journal. 2009 Dec;6(1):1-8. | CHN | 2005 | 1 | 0 | 99 |
| Ma C, Rodewald L, Hao L, Su Q, Zhang Y, Wen N, Fan C, Yang H, Luo H, Wang H, Goodson JL. Progress Toward Measles Elimination—China, January 2013-June 2019. China CDC Weekly. 2019 Dec;1(2):21. | CHN | 2013 | 1 | 0 | 99 |
|  | CHN | 2014 | 1 | 0 | 99 |
|  | CHN | 2015 | 1 | 0 | 99 |
|  | CHN | 2016 | 1 | 0 | 99 |
|  | CHN | 2017 | 1 | 0 | 99 |
|  | CHN | 2018 | 1 | 0 | 99 |
|  | CHN | 2019 | 1 | 0 | 99 |
| World Health Organization. Expanded Programme on Immunization: Measles outbreak= PROGRAMME ÉLARGI DE VACCINATION: Flambée de rougeole. 1990; 65(49): 379-81. | CHN | 1988 | 1 | 0 | 99 |
| Ye Y, Wang W, Wang X, Yu H. The clinical epidemiology of pediatric patients with measles from 2000 to 2009 in Shanghai, China. Clinical Pediatrics. 2011 Oct;50(10):916-22. | CHN | 2002 | 0 | 0 | 18 |
|  | CHN | 2007 | 0 | 0 | 18 |
| Yu X, Wang S, Guan J, Gou A, Liu Q, Jin X, Ghildyal R. Analysis of the cause of increased measles incidence in Xinjiang, China in 2004. The Pediatric infectious disease journal. 2007 Jun 1;26(6):513-8. | CHN | 2004 | 1 | 0 | 35 |
| Zhang RQ, Li HB, Li FY, Han LX, Xiong YM. Epidemiological characteristics of measles from 2000 to 2014: Results of a measles catch-up vaccination campaign in xianyang, china. Journal of Infection and Public Health. 2017 Sep 1;10(5):624-9. | CHN | 2009 | 1 | 0 | 99 |
| Njim T, Agyingi K, Aminde LN, Atunji EF. Trend in mortality from a recent measles outbreak in Cameroon: a retrospective analysis of 223 measles cases in the Benakuma Health District. The Pan African Medical Journal. 2016;23. | CMR | 2015 | 1 | 0 | 99 |
| Fischer PR. Measles in Zaire: 1987. Clinical pediatrics. 1988 May;27(5):234-5. | COD | 1986 | 0 | 0 | 15 |
| Gignoux E, Polonsky J, Ciglenecki I, Bichet M, Coldiron M, Thuambe Lwiyo E, Akonda I, Serafini M, Porten K. Risk factors for measles mortality and the importance of decentralized case management during an unusually large measles epidemic in eastern Democratic Republic of Congo in 2013. PloS one. 2018 Mar 14;13(3):e0194276. | COD | 2012 | 1 | 0 | 99 |
| Grout L, Minetti A, Hurtado N, François G, Fermon F, Chatelain A, Harczi G, de Dieu Ilunga Ngoie J, N’Goran A, Luquero FJ, Grais RF. Measles in Democratic Republic of Congo: an outbreak description from Katanga, 2010–2011. BMC infectious diseases. 2013 Dec;13(1):1-8. | COD | 2010 | 1 | 0 | 30 |
| Kasongo Project Team. Influence of measles vaccination on survival pattern of 7-35-month-old children in Kasongo, Zaire. The Lancet. 1981 Apr 4;317(8223):764-7. | COD | 1976 | 1 | 0 | 5 |
| Mancini S, Coldiron ME, Ronsse A, Ilunga BK, Porten K, Grais RF. Description of a large measles epidemic in Democratic Republic of Congo, 2010–2013. Conflict and health. 2014 Dec;8(1):1-8. | COD | 2010 | 1 | 0 | 99 |
|  | COD | 2011 | 1 | 0 | 99 |
|  | COD | 2012 | 1 | 0 | 99 |
|  | COD | 2013 | 1 | 0 | 99 |
| N’Goran AA, Ilunga N, Coldiron ME, Grais RF, Porten K. Community-based measles mortality surveillance in two districts of Katanga Province, Democratic Republic of Congo. BMC Research Notes. 2013 Dec;6(1):1-3. | COD | 2011 | 1 | 0 | 99 |
|  | COD | 2011 | 1 | 0 | 14 |
| Pan American Health Organization / World Health Organization. Epidemiological Update: Measles. 18 January 2019, Washington, D.C.: PAHO/WHO; 2019 | COL | 2018 | 1 | 0 | 99 |
| El Shazly MK, Atta HY, Kishk NA. Poliomyelitis, measles and neonatal tetanus: a hospital based epidemiological study. The Journal of the Egyptian Public Health Association. 1997 Jan 1;72(5-6):527-48. | EGY | 1994 | 0 | 0 | 60 |
| Belda K, Tegegne AA, Mersha AM, Bayenessagne MG, Hussein I, Bezabeh B. Measles outbreak investigation in Guji zone of Oromia Region, Ethiopia. The Pan African Medical Journal. 2017;27(Suppl 2). | ETH | 2015 | 1 | 0.25 | 30 |
| Gutu MA, Bekele A, Seid Y, Woyessa AB. Epidemiology of measles in Oromia region, Ethiopia, 2007-2016. The Pan African Medical Journal. 2020;37. | ETH | 2011 | 1 | 0 | 99 |
| Kalil FS, Gemeda DH, Bedaso MH, Wario SK. Measles outbreak investigation in Ginnir district of Bale zone, Oromia region, Southeast Ethiopia, May 2019. Pan African Medical Journal. 2020 May 14;36(1). | ETH | 2018 | 1 | 0 | 99 |
| Lindtjørn B. Severe measles in the Gardulla area of southwest Ethiopia. Journal of tropical pediatrics. 1986;32(5):234-9. | ETH | 1981 | 0 | 0 | 25 |
|  | ETH | 1981 | 1 | 0 | 4 |
| Mitiku K, Bedada T, Masresha BG, Kegne W, Nafo-Traoré F, Tesfaye N, Yigzaw A. Progress in measles mortality reduction in Ethiopia, 2002–2009. The Journal of infectious diseases. 2011 Jul 1;204(suppl_1):S232-8. | ETH | 2006 | 1 | 0.0833 | 65 |
| Navarro-Colorado C, Mahamud A, Burton A, Haskew C, Maina GK, Wagacha JB, Ahmed JA, Shetty S, Cookson S, Goodson JL, Schilperoord M. Measles outbreak response among adolescent and adult Somali refugees displaced by famine in Kenya and Ethiopia, 2011. The Journal of infectious diseases. 2014 Dec 15;210(12):1863-70. | ETH | 2011 | 0 | 0 | 99 |
| Poletti P, Parlamento S, Fayyisaa T, Feyyiss R, Lusiani M, Tsegaye A, Segafredo G, Putoto G, Manenti F, Merler S. The hidden burden of measles in Ethiopia: how distance to hospital shapes the disease mortality rate. BMC medicine. 2018 Dec;16(1):1-2. | ETH | 2015 | 0 | 0 | 65 |
| Tariku MK, Misikir SW. Measles outbreak investigation in Artuma Fursi Woreda, Oromia zone, Amhara region, Ethiopia, 2018: a case control study. BMC Research Notes. 2019 Dec;12(1):1-6. | ETH | 2018 | 1 | 0 | 99 |
| Güris D, Auerbach SB, Vitek C, Maes E, McCready J, Durand M, Cruz K, Iohp K, Haddock R, Rota J, Rota P. Measles outbreaks in Micronesia, 1991 to 1994. The Pediatric infectious disease journal. 1998 Jan 1;17(1):33-9. | FSM | 1992 | 1 | 0 | 19 |
| Bosu WK, Odoom S, Deiter P, Essel-Ahun M. Epidemiology of measles in the Central Region of Ghana: a five-year case review in three district hospitals. East African medical journal. 2003;80(6):312-7. | GHA | 1998 | 0 | 0 | 60 |
| Commey JO, Dekyem P. Measles in southern Ghana: 1985-1993. West African Journal of Medicine. 1994 Oct 1;13(4):223-6. | GHA | 1989 | 0 | 0.25 | 12 |
| Commey JO, Richardson JE. Measles in Ghana—1973–1982. Annals of tropical paediatrics. 1984 Sep 1;4(3):189-94. | GHA | 1977 | 0 | 0.3333 | 14 |
| Dollimore N, Cutts F, Binka FN, Ross DA, Morris SS, George Smith P. Measles incidence, case fatality, and delayed mortality in children with or without vitamin A supplementation in rural Ghana. American journal of epidemiology. 1997 Oct 15;146(8):646-54. | GHA | 1990 | 1 | 0 | 7 |
| Hull H, Williams PJ, Oldfield F. Measles mortality and vaccine efficacy in rural West Africa. The Lancet. 1983 Apr 30;321(8331):972-5. | GMB | 1981 | 1 | 0.25 | 10 |
| Hull HF. Increased measles mortality in households with multiple cases in the Gambia, 1981. Clinical Infectious Diseases. 1988 Mar 1;10(2):463-7. | GMB | 1981 | 1 | 0 | 10 |
| Lamb WH. Epidemic measles in a highly immunized rural West African (Gambian) village. Clinical Infectious Diseases. 1988 Mar 1;10(2):457-62. | GMB | 1984 | 1 | 0 | 99 |
| Williams PJ. Effect of measles immunization on child mortality in rural Gambia. Journal of Biosocial Science. 1989;21(S10):95-104. | GMB | 1984 | 1 | 0 | 15 |
| Williams PJ, Hull HF. Status of measles in the Gambia, 1981. Reviews of infectious diseases. 1983 May 1;5(3):391-4. | GMB | 1981 | 1 | 0.25 | 10 |
| Aaby P, Bukh J, Lisse IM, da Silva MC. Further community studies on the role of overcrowding and intensive exposure on measles mortality. Clinical Infectious Diseases. 1988 Mar 1;10(2):474-7. | GNB | 1981 | 1 | 0.42 | 18 |
| Aaby P, Bukh J, Lisse IM, Smits AJ. Introduction of measles into a highly immunised West African community: the role of health care institutions. Journal of Epidemiology & Community Health. 1985 Jun 1;39(2):113-6. | GNB | 1981 | 1 | 0 | 99 |
| Aaby P, Bukh J, Lisse IM, Smits AJ. Measles mortality, state of nutrition, and family structure: a community study from Guinea-Bissau. Journal of infectious diseases. 1983 Apr 1;147(4):693-701. | GNB | 1979 | 1 | 0 | 18 |
| Aaby P, Knudsen K, Jensen TG, Thirup J, Poulsen A, Sodemann M, da Silva MC, Whittle H. Measles incidence, vaccine efficacy, and mortality in two urban African areas with high vaccination coverage. Journal of infectious diseases. 1990 Nov 1;162(5):1043-8. | GNB | 1983 | 1 | 0 | 1 |
|  | GNB | 1986 | 1 | 0 | 1 |
| Aaby P, Martins C, Bale C, Garly ML, Rodrigues A, Biai S, Lisse IM, Whittle H, Benn CS. Sex differences in the effect of vaccines on the risk of hospitalization due to measles in Guinea-bissau. The Pediatric infectious disease journal. 2010 Apr 1;29(4):324-8. | GNB | 2003 | 0 | 0.5 | 5 |
| Aaby P, Bukh J, Lisse IM, da Silva MC. Decline in measles mortality: nutrition, age at infection, or exposure?. Br Med J (Clin Res Ed). 1988 Apr 30;296(6631):1225-8. | GNB | 1979 | 1 | 0 | 17 |
|  | GNB | 1982 | 1 | 0 | 17 |
| Aaby P, Bukh J, Lisse IM, Smits AJ, Gomes J, Fernandes MA, Indi F, Soares M. Determinants of measles mortality in a rural area of Guinea-Bissau: crowding, age, and malnutrition. Journal of tropical pediatrics. 1984 Jun 1;30(3):164-8. | GNB | 1980 | 1 | 0 | 20 |
| Martins CL, Garly ML, Balé C, Rodrigues A, Ravn H, Whittle HC, Lisse IM, Aaby P. Protective efficacy of standard Edmonston-Zagreb measles vaccination in infants aged 4.5 months: interim analysis of a randomised clinical trial. Bmj. 2008 Jul 24;337. | GNB | 2003 | 1 | 0.375 | 0.375 |
| Aaby P, Bukh J, Lisse IM, Smits AJ. Overcrowding and intensive exposure as determinants of measles mortality. American journal of epidemiology. 1984 Jul 1;120(1):49-63. | GNB | 1979 | 1 | 0 | 5 |
| Tollefson JE, Hospedales CJ, White FM. Epidemiological indicators and the epidemiology of measles in the English-speaking Caribbean and Suriname. The West Indian Medical Journal. 1992 Mar 1;41(1):2-7. | GUY | 1988 | 1 | 0 | 99 |
| World Health Organization. Measles surveillance: Measles in the Caribbean prior to the elimination campaign. Weekly Epidemiological Record= Relevé épidémiologique hebdomadaire. 1991;66(40):291-4. | GUY | 1988 | 1 | 0 | 99 |
| Lubis CP, Pasaribu S, Lubis MM. Morbidity and mortality of tetanus, diphtheria and morbilli (measles) cases (a 1982-1985 study at the Child Health Department, Dr. Pirngadi Hospital, Medan). The Journal of the Singapore Paediatric Society. 1987 Jan 1;29:66-72. | IDN | 1982 | 0 | 0 | 15 |
|  | IDN | 1983 | 0 | 0 | 15 |
|  | IDN | 1984 | 0 | 0 | 15 |
|  | IDN | 1985 | 0 | 0 | 15 |
|  | IDN | 1986 | 0 | 0 | 15 |
| Munir M, Mustadjab I, Wulur FH. Measles and its problems. A clinical analysis of hospitalized patients under 5 years of age. Paediatrica Indonesiana. 1982 Apr 30;22(3-4):49-64. | IDN | 1980 | 0 | 0 | 5 |
| Rangkuti SM, Nazir N, Sutanto AH, Lubis A, Siregar H. Measles morbidity and mortality in the Department of Child Health, Dr Pirngadi General Hospital, Medan, in 1973-1977. Paediatrica Indonesiana. 1980;20(7/8):139-44. | IDN | 1975 | 0 | 0 | 12 |
| Samsi TK, Ruspandji T, Susanto I, Gunawan K. Risk factors for severe measles. The Southeast Asian Journal of Tropical Medicine and Public Health. 1992 Sep 1;23(3):497-503. | IDN | 1984 | 0 | 0.5 | 18 |
| Agarwal DK, Dutta A, Arora RR, Nair MR. Natural history of measles in rural and urban community of Varanasi. Journal of Communicable Diseases. 1976;8(4):289-98. | IND | 1974 | 1 | 0 | 18 |
| Ananthakrishnan S, Srinivasan S, Mahadevan S. Vitamin A and post measles complications. Indian pediatrics. 1993;30(4):520-2. | IND | 1989 | 0 | 0.5 | 18 |
| Basa S, Das RR, Khan JA. Root-Cause Analytical Survey for Measles Outbreak: Vaccination or Vaccine?-A Study From Madhepura District, Bihar, India. Journal of Clinical and Diagnostic Research: JCDR. 2015 Jun;9(6):SC04. | IND | 2008 | 1 | 0 | 12 |
| Basu RN. Measles vaccine—feasiblity, efficacy and complication rates in a multicentric study. The Indian Journal of Pediatrics. 1984 Mar;51(2):139-43. | IND | 1982 | 1 | 0.75 | 2 |
| Bhatia R. Measles outbreak in village Tophema in Nagaland. Journal of communicable diseases. 1985;17(2):185-9. | IND | 1983 | 1 | 1 | 12 |
| Bose AS, Jafari H, Sosler S, Narula AP, Kulkarni VM, Ramamurty N, Oommen J, Jadi RS, Banpel RV, Henao-Restrepo AM. Case based measles surveillance in Pune: evidence to guide current and future measles control and elimination efforts in India. PLoS One. 2014 Oct 7;9(10):e108786. | IND | 2010 | 1 | 0 | 99 |
| Chand P, Rai RN, Chawla U, Tripathi KC, Datta KK. Epidemiology of measles--a thirteen years prospective study in a village. The Journal of Communicable Diseases. 1989 Sep 1;21(3):190-9. | IND | 1980 | 1 | 0 | 14 |
| Cherian T, Joseph A, John TJ. Low antibody response in infants with measles and children with subclinical measles virus infection. The Journal of Tropical Medicine and Hygiene. 1984 Feb 1;87(1):27-31. | IND | 1979 | 1 | 0 | 5 |
| Dhanoa JA, Cowan BE. Measles in the community-a study in non-hospitalised young children in Punjab. Journal of Tropical Pediatrics. 1982;28(2):59-61. | IND | 1980 | 1 | 0 | 2 |
| Gupta BP, Sharma S. Measles outbreak in a rural area near Shimla. Indian Journal of Community Medicine. 2006 Apr 1;31(2):106. | IND | 2004 | 1 | 0 | 14 |
| Gupta BP, Swami HM, Bhardwaj AK, Vaidya NK, Sharma CD, Kaushal RK. An outbreak of measles in a remote tribal area of Himachal Pradesh. Indian J Comm Health. 1989;5:25-8. | IND | 1986 | 1 | 0 | 14 |
| Jajoo UN, Chhabra S, Gupta OP, Jain AP. Measles epidemic in a rural community near Sevagram (Vidarbha). Indian journal of public health. 1984;28(4):204-7. | IND | 1982 | 1 | 0 | 10 |
| John S, Sanghi S, Prasad S, Bose A, George K. Two doses of measles vaccine: are some states in India ready for it?. Journal of tropical pediatrics. 2009 Aug 1;55(4):253-6. | IND | 1999 | 1 | 0 | 9 |
|  | IND | 2006 | 1 | 0 | 18 |
| John TJ, Joseph A, George TI, Radhakrishnan J, Singh RP, George K. Epidemiology and prevention of measles in rural south India. Indian Journal of Medical Research. 1980;72(August):153-8. | IND | 1977 | 1 | 0 | 9 |
| Kalita J, Mani VE, Bhoi SK, Misra UK. Spectrum and outcome of acute infectious encephalitis/encephalopathy in an intensive care unit from India. QJM: An International Journal of Medicine. 2017 Mar 1;110(3):141-8. | IND | 2013 | 0 | 2 | 85 |
| Lakhanpal U, Rathore MS. Epidemiology of measles in rural area of Punjab. Journal of communicable diseases. 1986;18(3):185-8. | IND | 1983 | 1 | 0 | 14 |
| Lobo J, Reddaiah VP, Kapoor SK, Nath LM. Epidemiology of measles in a rural community. The Indian Journal of Pediatrics. 1987 Mar;54(2):261-5. | IND | 1984 | 1 | 0 | 9 |
| Mangal N, Shah K, Sitaraman S. Epidemiological study of measles in urban (slum) area of Jaipur. Indian pediatrics. 1990;27(11):1216-7. | IND | 1985 | 1 | 0 | 9 |
| Mishra A, Mishra S, Jain P, Bhadoriya RS, Mishra R, Lahariya C. Measles related complications and the role of vitamin A supplementation. The Indian Journal of Pediatrics. 2008 Sep;75(9):887-90. | IND | 2004 | 1 | 0 | 18 |
| Murhekar MV, Ahmad M, Shukla H, Abhishek K, Perry RT, Bose AS, Shimpi R, Kumar A, Kaliaperumal K, Sethi R, Selvaraj V. Measles case fatality rate in Bihar, India, 2011–12. Plos one. 2014 May 13;9(5):e96668. | IND | 2011 | 1 | 0 | 99 |
| Murhekar MV, Hutin YJ, Ramakrishnan R, Ramachandran V, Biswas AK, Das PK, Gupta SN, Maji D, Martolia HC, Mohan A, Gupte MD. The heterogeneity of measles epidemiology in India: implications for improving control measures. The Journal of infectious diseases. 2011 Jul 1;204(suppl_1):S421-6. | IND | 2004 | 1 | 0 | 14 |
|  | IND | 2005 | 1 | 0 | 14 |
|  | IND | 2006 | 1 | 0 | 14 |
| NARAIN JP, KHARE S, Rana SR, Banerjee KB. Epidemic measles in an isolated unvaccinated population, India. International journal of epidemiology. 1989 Dec 1;18(4):952-8. | IND | 1986 | 0 | 0 | 99 |
|  | IND | 1986 | 1 | 0 | 99 |
| Phaneendra Rao RS, Kumari J, RAO K, Narasimham VL. Measles in a rural Community. Journal of communicable diseases. 1988;20(2):131-5. | IND | 1983 | 1 | 0 | 5 |
| Raoot A, Dewan DK, Dubey AP, Batra RK, Seth S. Measles outbreak in high risk areas of Delhi: epidemiological investigation and laboratory confirmation. The Indian Journal of Pediatrics. 2016 Mar;83(3):200-8. | IND | 2014 | 1 | 0 | 99 |
| Ratho RK, Mishra B, Singh T, Rao P, Kumar R. Measles outbreak in a migrant population. Indian J Pediatr. 2005;72(10):893-4. | IND | 2003 | 1 | 1 | 25 |
| Ray SK, Mallik S, Munsi AK, Mitra SP, Baur B, Kumar S. Epidemiological study of measles in slum areas of Kolkata. The Indian Journal of Pediatrics. 2004 Jul;71(7):583-6. | IND | 1999 | 1 | 0 | 17 |
| Risbud AR, Prasad SR, Mehendale SM, Mawar N, Shaikh N, Umrani UB, Bedekar SS, Banerjee K. Measles outbreak in a tribal population of Thane district, Maharashtra. Indian pediatrics. 1994 May 1;31(5):543-51. | IND | 1991 | 1 | 0 | 10 |
|  | IND | 1992 | 1 | 0 | 10 |
| Satpathy SK, Chakraborty AK. Epidemiological study of measles in Singur, West Bengal. The Journal of Communicable Diseases. 1990 Mar 1;22(1):23-6. | IND | 1986 | 1 | 0 | 99 |
| Sharma MK, Bhatia V, Swami H. Outbreak of measles amongst vaccinated children in a slum of Chandigarh. Indian Journal of Medical Sciences. 2004;58(2):47. | IND | 2003 | 1 | 0 | 14 |
| Sharma RS. An epidemiological study of measles epidemic in district Bhilwara, Rajasthan. Journal of communicable diseases. 1988;20(4):301-11. | IND | 1984 | 1 | 0 | 13 |
| Sharma RS, Kaushic VK, Johri SP, Ray SN. An epidemiological investigation of measles outbreak in Alwar-Rajasthan. Journal of communicable diseases. 1984;16(4):299-303. | IND | 1982 | 1 | 0 | 5 |
| Singh J, Kumar A, Rai RN, Khare S, Jain DC, Bhatia R, Datta KK. Widespread outbreaks of measles in rural Uttar Pradesh, India, 1996: high risk areas and groups. Indian pediatrics. 1999 Mar 1;36(3):249-56. | IND | 1996 | 1 | 0 | 99 |
| Singh J, Sharma RS, Verghese T. Measles mortality in India: a review of community based studies. The Journal of Communicable Diseases. 1994 Dec 1;26(4):203-14. | IND | 1980 | 1 | 0 | 99 |
|  | IND | 1985 | 1 | 0 | 99 |
|  | IND | 1992 | 1 | 0 | 14 |
| Swami SS, Chandra S, Dudani IU, Sharma R, Mathur MM. Epidemiology of measles in Western Rajasthan. Journal of communicable diseases. 1987;19(4):370-2. | IND | 1980 | 1 | 0 | 14 |
| Thakur JS, Ratho RK, Bhatia SP, Grover R, Issaivanan M, Ahmed B, Parmar V, Swami HM. Measles outbreak in a Periurban area of Chandigarh: need for improving vaccine coverage and strengthening surveillance. The Indian Journal of Pediatrics. 2002 Jan;69(1):33-7. | IND | 1998 | 1 | 0 | 99 |
| Vasudev JP, Nandan D, Chandra R, Srivastava BC. Post measles complications in a rural population. Journal of communicable diseases. 1983;15(4):249-52. | IND | 1980 | 1 | 0 | 17 |
| Janghorbani M, Parizi MH, Ghorbani K. Measles epidemics in Kerman city, Iran. Public Health. 1993 Mar 1;107(2):79-87. | IRN | 1990 | 1 | 0.4167 | 35 |
| Tollefson JE, Hospedales CJ, White FM. Epidemiological indicators and the epidemiology of measles in the English-speaking Caribbean and Suriname. The West Indian Medical Journal. 1992 Mar 1;41(1):2-7. | JAM | 1989 | 1 | 0 | 99 |
| World Health Organization. Measles surveillance: Measles in the Caribbean prior to the elimination campaign. Weekly Epidemiological Record= Relevé épidémiologique hebdomadaire. 1991;66(40):291-4. | JAM | 1989 | 1 | 0 | 99 |
| Alwar AJ. The effect of protein energy malnutrition on morbidity and mortality due to measles at Kenyatta National Hospital, Nairobi (Kenya). East African medical journal. 1992 Aug 1;69(8):415-8. | KEN | 1983 | 0 | 0.0385 | 20 |
| Borus PK, Cumberland P, Sonoiya S, Kombich J, Tukei PM, Cutts FT. Measles trends and vaccine effectiveness in Nairobi, Kenya. East African medical journal. 2003;80(7):361-4. | KEN | 1998 | 0 | 0 | 99 |
| Burström B, Abby P, Mutie DM. Validity of measles mortality data using hospital registers and community surveys. International journal of epidemiology. 1995 Jun 1;24(3):625-9. | KEN | 1986 | 0 | 0 | 17 |
|  | KEN | 1986 | 1 | 0 | 17 |
|  | KEN | 1988 | 0 | 0 | 17 |
|  | KEN | 1988 | 1 | 0 | 17 |
| Burström B, Aaby P, Mutie DM. Child mortality impact of a measles outbreak in a partially vaccinated rural African community. Scandinavian journal of infectious diseases. 1993 Jan 1;25(6):763-9. | KEN | 1987 | 1 | 0 | 99 |
| Burström B, Aaby P, Mutie DM, Kimani G, Bjerregaard P. Severe measles outbreak in western Kenya. East African medical journal. 1992 Aug 1;69(8):419-23. | KEN | 1985 | 1 | 0 | 17 |
| Kisangau N, Sergon K, Ibrahim Y, Yonga F, Langat D, Nzunza R, Borus P, Galgalo T, Lowther SA. Progress towards elimination of measles in Kenya, 2003-2016. Pan African Medical Journal. 2018 Sep 28;31(1). | KEN | 2009 | 1 | 0 | 99 |
| Mahamud A, Burton A, Hassan M, Ahmed JA, Wagacha JB, Spiegel P, Haskew C, Eidex RB, Shetty S, Cookson S, Navarro-Colorado C. Risk factors for measles mortality among hospitalized Somali refugees displaced by famine, Kenya, 2011. Clinical Infectious Diseases. 2013 Oct 15;57(8):e160-6. | KEN | 2011 | 0 | 0 | 99 |
| Menge I, Esamai F, Van Reken D, Anabwani G. Paediatric morbidity and mortality at the Eldoret District Hospital, Kenya. East African medical journal. 1995;72(3):165-9. | KEN | 1993 | 0 | 0 | 17 |
| Muller AS, Voorhoeve AM, T'mannetje W, Schulpen TW. The impact of measles in a rural area of Kenya. East African medical journal. 1977;54(7):364-72. | KEN | 1975 | 1 | 0 | 14 |
|  | KEN | 1976 | 1 | 0 | 14 |
| Navarro-Colorado C, Mahamud A, Burton A, Haskew C, Maina GK, Wagacha JB, Ahmed JA, Shetty S, Cookson S, Goodson JL, Schilperoord M. Measles outbreak response among adolescent and adult Somali refugees displaced by famine in Kenya and Ethiopia, 2011. The Journal of infectious diseases. 2014 Dec 15;210(12):1863-70. | KEN | 2011 | 0 | 0 | 99 |
| Centers for Disease Control and Prevention (CDC. Accelerated measles control--Cambodia, 1999-2002. MMWR. Morbidity and mortality weekly report. 2003 Jan 10;52(1):4-6. | KHM | 1999 | 1 | 0 | 99 |
| Oum S, Chandramohan D, Cairncross S. Community‐based surveillance: a pilot study from rural Cambodia. Tropical medicine & international health. 2005 Jul;10(7):689-97. | KHM | 2001 | 1 | 0 | 14 |
| Kuroiwa C, Vongphrachanh P, Xayyavong P, Southalack K, Hashizume M, Nakamura S. Measles epidemiology and outbreak investigation using IgM test in Laos. Journal of Epidemiology. 2001;11(6):255-62. | LAO | 1994 | 1 | 0 | 99 |
|  | LAO | 1995 | 1 | 0 | 99 |
|  | LAO | 1996 | 1 | 0 | 99 |
|  | LAO | 1997 | 1 | 0 | 99 |
|  | LAO | 1998 | 1 | 0 | 99 |
|  | LAO | 1999 | 1 | 0 | 99 |
|  | LAO | 2000 | 1 | 0 | 99 |
| Nagbe T, Williams GS, Rude JM, Flomo S, Yeabah T, Fallah M, Skrip L, Agbo C, Mahmoud N, Okeibunor JC, Yealue K. Lessons learned from detecting and responding to recurrent measles outbreak in Liberia post Ebola-Epidemic 2016-2017. The Pan African Medical Journal. 2019;33(Suppl 2). | LBR | 2016 | 1 | 0 | 99 |
| Lamabadusuriya SP, Jayantha UK. An outbreak of measles in the Southern Province. The Ceylon Medical Journal. 1992 Jun 1;37(2):46-8. | LKA | 1989 | 0 | 0 | 17 |
| Premaratna R, Luke N, Perera H, Gunathilake M, Amarasena P, Chandrasena TG. Sporadic cases of adult measles: a research article. BMC research notes. 2017 Dec;10(1):1-6. | LKA | 2015 | 0 | 18 | 99 |
| Puvimanasinghe JP, Arambepola CK, Abeysinghe NM, Rajapaksa LC, Kulatilaka TA. Measles outbreak in Sri Lanka, 1999–2000. Journal of Infectious Diseases. 2003 May 15;187(Supplement_1):S241-5. | LKA | 1999 | 1 | 0 | 99 |
| World Health Organization (WHO). Expanded programme on immunization. Public health importance of measles: Sri Lanka. Wkly Epidemiol Rec. 1985;60(13): 95-7. | LKA | 1983 | 1 | 0 | 99 |
| Nimpa MM, Andrianirinarison JC, Sodjinou VD, Douba A, Masembe YV, Randriatsarafara F, Ramamonjisoa CB, Rafalimanantsoa AS, Razafindratsimandresy R, Ndiaye CF, Rakotonirina J. Measles outbreak in 2018-2019, Madagascar: epidemiology and public health implications. The Pan African Medical Journal. 2020;35. | MDG | 2018 | 1 | 0 | 99 |
| Hyde TB, Dayan GH, Langidrik JR, Nandy R, Edwards R, Briand K, Konelios M, Marin M, Nguyen HQ, Khalifah AP, O'leary MJ. Measles outbreak in the Republic of the Marshall Islands, 2003. International journal of epidemiology. 2006 Apr 1;35(2):299-306. | MHL | 2003 | 1 | 0 | 49 |
| McIntyre RC, Preblud SR, Polloi AN, Korean MA. Measles and measles vaccine efficacy in a remote island population. Bulletin of the World Health Organization. 1982;60(5):767. | MHL | 1977 | 1 | 0 | 30 |
| World Health Organization. Measles mortality reduction in West Africa, 1996-2002. Weekly Epidemiological Record= Relevé épidémiologique hebdomadaire. 2003;78(45):390-2. | MLI | 2002 | 1 | 0 | 99 |
| Chin J, Thaung UM. The unchanging epidemiology and toll of measles in Burma. Bulletin of the World Health Organization. 1985;63(3):551. | MMR | 1983 | 0 | 0 | 9 |
|  | MMR | 1983 | 1 | 0 | 9 |
| Khin M, Win S, Aye SS. The impact of national measles immunization programme on measles admissions to the major children's hospital in Yangon. Tropical doctor. 1994 Jul;24(3):141-3. | MMR | 1985 | 0 | 0 | 12 |
|  | MMR | 1989 | 0 | 0 | 12 |
| Lee CT, Hagan JE, Jantsansengee B, Tumurbaatar OE, Altanchimeg S, Yadamsuren B, Demberelsuren S, Tserendorj C, Munkhtogoo O, Badarch D, Gunregjav N. Increase in infant measles deaths during a nationwide measles outbreak—Mongolia, 2015–2016. The Journal of infectious diseases. 2019 Oct 22;220(11):1771-9. | MNG | 2015 | 1 | 0 | 0.9999 |
|  | MNG | 2015 | 1 | 0 | 99 |
| Orsoo O, Saw YM, Sereenen E, Yadamsuren B, Byambaa A, Kariya T, Yamamoto E, Hamajima N. Epidemiological characteristics and trends of a Nationwide measles outbreak in Mongolia, 2015–2016. BMC Public Health. 2019 Dec;19(1):1-0. | MNG | 2015 | 1 | 0 | 99 |
| Cliff J, Simango A, Augusto O, Van der Paal L, Biellik R. Failure of targeted urban supplemental measles vaccination campaigns (1997–1999) to prevent measles epidemics in Mozambique (1998–2001). Journal of Infectious Diseases. 2003 May 15;187(Supplement_1):S51-7. | MOZ | 1993 | 1 | 0 | 99 |
|  | MOZ | 1998 | 1 | 0 | 99 |
| Mandomando I, Naniche D, Pasetti MF, Cuberos L, Sanz S, Vallès X, Sigauque B, Macete E, Nhalungo D, Kotloff KL, Levine MM. Assessment of the epidemiology and burden of measles in Southern Mozambique. The American journal of tropical medicine and hygiene. 2011 Jul 7;85(1):146. | MOZ | 2002 | 0 | 0 | 18 |
| Boushab BM, Savadogo M, Sow MS, Dao S. Epidemiological, clinical, and prognostic study of the measles in the Aioun regional hospital in Mauritania. Médecine et Santé Tropicales. 2015 Apr 1;25(2):180-3. | MRT | 2011 | 0 | 1 | 33 |
| Minetti A, Kagoli M, Katsulukuta A, Huerga H, Featherstone A, Chiotcha H, Noel D, Bopp C, Sury L, Fricke R, Iscla M. Lessons and challenges for measles control from unexpected large outbreak, Malawi. Emerging infectious diseases. 2013 Feb;19(2):202. | MWI | 2010 | 1 | 0 | 99 |
| Courtright P, Fine D, Broadhead RL, Misoya L, Vagh M. Abnormal vitamin A cytology and mortality in infants aged 9 months and less with measles. Annals of tropical paediatrics. 2002 Sep 1;22(3):239-43. | MWI | 1992 | 0 | 0.25 | 0.8333 |
| Yamaguchi S, Dunga A, Broadhead RL, Brabin BJ. Epidemiology of measles in Blantyre, Malawi: analyses of passive surveillance data from 1996 to 1998. Epidemiology & Infection. 2002 Oct;129(2):361-9. | MWI | 1997 | 1 | 0 | 99 |
| Grais RF, Dubray C, Gerstl S, Guthmann JP, Djibo A, Nargaye KD, Coker J, Alberti KP, Cochet A, Ihekweazu C, Nathan N. Unacceptably high mortality related to measles epidemics in Niger, Nigeria, and Chad. PLoS medicine. 2007 Jan;4(1):e16. | NER | 2003 | 1 | 0 | 99 |
| Kaninda AV, Legros D, Jataou IM, Malfait P, Maisonneuve M, Paquet C, Moren A. Measles vaccine effectiveness in standard and early immunization strategies, Niger, 1995. The Pediatric infectious disease journal. 1998 Nov 1;17(11):1034-9. | NER | 1995 | 1 | 0 | 4 |
| Malfait P, Jataou IM, Jollet MC, Margot A, DE BENOIST AC, Moren A. Measles epidemic in the urban community of Niamey: transmission patterns, vaccine efficacy and immunization strategies, Niger, 1990 to 1991. The Pediatric infectious disease journal. 1994 Jan 1;13(1):38-44. | NER | 1990 | 0 | 0.5 | 4.9167 |
| Nandy R, Handzel T, Zaneidou M, Biey J, Coddy RZ, Perry R, Strebel P, Cairns L. Case-fatality rate during a measles outbreak in eastern Niger in 2003. Clinical infectious diseases. 2006 Feb 1;42(3):322-8. | NER | 2003 | 1 | 0 | 99 |
| World Health Organization. Expanded Programme on Immunization: High measles case-fatality rates during an outbreak in a rural area. Weekly Epidemiological Record. 1993;68(20):142-5. | NER | 1991 | 1 | 0 | 99 |
| Adedoyin MA. The pattern of measles in Ilorin. West African Journal of Medicine. 1990 Apr 1;9(2):103-7. | NGA | 1982 | 0 | 0 | 5 |
|  | NGA | 1983 | 0 | 0 | 5 |
|  | NGA | 1984 | 0 | 0 | 5 |
| Ahmed PA, Babaniyi IB, Otuneye AT. Review of childhood measles admissions at the National Hospital, Abuja. Nigerian Journal of Clinical Practice. 2010;13(4). | NGA | 2003 | 0 | 0.5833 | 10 |
| Babalola OJ, Ibrahim IN, Kusfa IU, Gidado S, Nguku P, Olayinka A, Abubakar A. Measles outbreak investigation in an urban slum of Kaduna Metropolis, Kaduna State, Nigeria, March 2015. Pan African Medical Journal. 2019 Mar 28;32(1). | NGA | 2015 | 1 | 0 | 99 |
| Bamgboye EA, Familusi JB. Mortality pattern at a children's emergency ward, University College Hospital, Ibadan, Nigeria. African journal of medicine and medical sciences. 1990 Jun 1;19(2):127-32. | NGA | 1982 | 0 | 0 | 17 |
| Byass P, Adedeji MD, Mongdem JG, Zwandor AC, Brew-Graves SH, Clements CJ. Assessment and possible control of endemic measles in urban Nigeria. Journal of Public Health. 1995 Jun 1;17(2):140-5. | NGA | 1992 | 1 | 0 | 4 |
| Ekanem EE, Ochigbo SO, Kwagtsule JU. Unprecedented decline in measles morbidity and mortality in Calabar, south-eastern Nigeria. Tropical doctor. 2000 Oct;30(4):207-9. | NGA | 1994 | 0 | 0 | 11 |
| Fagbule D, Orifunmishe F. Measles and childhood mortality in semi-urban Nigeria. African journal of medicine and medical sciences. 1988 Sep 1;17(3):181-5. | NGA | 1984 | 0 | 0.25 | 7 |
| Faruk AS, Adebowale AS, Balogun MS, Taiwo L, Adeoye O, Mamuda S, Waziri NE. Temporal trend of measles cases and impact of vaccination on mortality in Jigawa State, Nigeria, 2013-2017: a secondary data analysis. The Pan African Medical Journal. 2020;35(Suppl 1). | NGA | 2015 | 1 | 0 | 99 |
| Fatiregun AA, Adebowale AS, Fagbamigbe AF. Epidemiology of measles in Southwest Nigeria: an analysis of measles case-based surveillance data from 2007 to 2012. Transactions of the Royal Society of Tropical Medicine and Hygiene. 2014 Mar 1;108(3):133-40. | NGA | 2009 | 1 | 0 | 99 |
| Fatiregun AA, Olowookere SA, Abubakar O, Aderibigbe A. Small-scale outbreak of measles in the Irewole local government area of Osun State in Nigeria. Asian Pacific Journal of Tropical Medicine. 2009;2(6):33-6. | NGA | 2008 | 1 | 0.6667 | 14 |
| Fetuga MB, Njakanna OF, Ongunfowora OB. A ten-year study of measles admissions in a Nigerian teaching hospital. Nigerian Journal of Clinical Practice. 2007 Sep 14;10(1):41-6. | NGA | 1999 | 0 | 0.3333 | 12 |
| Grais RF, Dubray C, Gerstl S, Guthmann JP, Djibo A, Nargaye KD, Coker J, Alberti KP, Cochet A, Ihekweazu C, Nathan N. Unacceptably high mortality related to measles epidemics in Niger, Nigeria, and Chad. PLoS medicine. 2007 Jan;4(1):e16. | NGA | 2004 | 1 | 0 | 99 |
| Ibia EO, Asindi AA. Measles in Nigerian children in Calabar during the era of expanded programme on immunization. Tropical and geographical medicine. 1990 Jul 1;42(3):226-32. | NGA | 1985 | 0 | 0.375 | 11 |
| Ibrahim BS, Usman R, Mohammed Y, Datti Z, Okunromade O, Abubakar AA, Nguku PM. Burden of measles in Nigeria: a five-year review of casebased surveillance data, 2012-2016. The Pan African Medical Journal. 2019;32(Suppl 1). | NGA | 2012 | 1 | 0 | 99 |
|  | NGA | 2013 | 1 | 0 | 99 |
|  | NGA | 2014 | 1 | 0 | 99 |
|  | NGA | 2015 | 1 | 0 | 99 |
|  | NGA | 2016 | 1 | 0 | 99 |
| Lagunju IA, Orimadegun AE, Oyedemi DG. Measles in Ibadan: a continuous scourge. African journal of medicine and medical sciences. 2005 Dec 1;34(4):383-7. | NGA | 2002 | 0 | 0.3333 | 10 |
| Olugbade OT, Adeyemi AS, Adeoti AH, Ilesanmi OS, Gidado SO, Waziri NE, Aworh MK. Measles outbreaks and Supplemental Immunization Activities (SIAs): the Gwagwalada experience, Abuja 2015. The Pan African Medical Journal. 2019;32(Suppl 1). | NGA | 2015 | 1 | 0 | 33 |
| Weldegebriel GG, Gasasira A, Harvey P, Masresha B, Goodson JL, Pate MA, Abanida E, Chevez A. Measles resurgence following a nationwide measles vaccination campaign in Nigeria, 2005–2008. The Journal of infectious diseases. 2011 Jul 1;204(suppl_1):S226-31. | NGA | 2008 | 1 | 0 | 99 |
| Joshi AB, Luman ET, Nandy R, Subedi BK, Liyanage JB, Wierzba TF. Measles deaths in Nepal: estimating the national case-fatality ratio. Bulletin of the World Health Organization. 2009;87:456-65. | NPL | 2004 | 1 | 0 | 99 |
| Sitaula S, Awasthi GR, Thapa JB, Ramaiya A. Measles outbreak among unvaccinated children in Bajura. Journal of Nepal Medical Association. 2010 Oct 1;50(180). | NPL | 2010 | 1 | 0.75 | 25 |
| Aurangzeb B, Fatmee A, Waris R, Haider N, Berjees A, Raza SH. Risk factors for mortality among admitted children with complications of measles in Pakistan–an observational. Journal of the Pakistan Medical Association. 2020 Nov 3:1-4. | PAK | 2015 | 0 | 0.75 | 18 |
| Aurangzeb B, Nisar YB, Hazir T, Burki F, Hassan M. Clinical outcome in children hospitalized with complicated measles. J Coll Physicians Surg Pak. 2005 Sep 1;15(9):547-1. | PAK | 2003 | 0 | 0.5 | 12 |
| Murray M, Rasmussen Z. Measles outbreak in a northern Pakistani village: epidemiology and vaccine effectiveness. American journal of epidemiology. 2000 Apr 15;151(8):811-9. | PAK | 1990 | 1 | 0 | 13 |
| Anis-ur-Rehman ST, Idris M. Clinical outcome in measles patients hospitalized with complications. J Ayub Med Coll Abbottabad. 2008;20(2):14-6. | PAK | 2004 | 0 | 0.5 | 12 |
| Rehman IU, Bukhsh A, Khan TM. Measles in Pakistan: time to make steps towards eradication. Travel medicine and infectious disease. 2017 Jul 1;18:67-9. | PAK | 2012 | 1 | 0 | 18 |
|  | PAK | 2013 | 1 | 0 | 18 |
| Saeed A, Butt ZA, Malik T. Investigation of measles outbreak in a district of Balochistan province, Pakistan. Journal of Ayub Medical College Abbottabad. 2015 Dec 15;27(4):900-3. | PAK | 2014 | 1 | 0 | 11 |
| Sniadack DH, Moscoso B, Aguilar R, Heath J, Bellini W, Chiu MC. Measles epidemiology and outbreak response immunization in a rural community in Peru. Bulletin of the World Health Organization. 1999;77(7):545. | PER | 1993 | 1 | 0 | 40 |
| Almoradie-Javonillo I, Javonillo T. Profile of a measles epidemic in a remote Philippine barrio. Journal of the Philippine Medical Association. 1984 Apr 30;60(3). | PHL | 1983 | 1 | 0 | 14 |
| Bronzwaer SL, De Groot CJ. Risk factors for a complicated disease course in children with measles admitted to a Philippine university hospital. Nederlands Tijdschrift Voor Geneeskunde. 1997 Dec 1;141(51):2492-5. | PHL | 1994 | 0 | 0 | 15 |
| Benjamin AL, Dramoi V. Outbreak of measles in the National Capital District, Papua New Guinea in 2001. Papua and New Guinea Medical Journal. 2002 Sep 1;45(3-4):178-84. | PNG | 2001 | 0 | 0 | 17 |
| Coakley KJ, Coakley CA, Spooner V, Smith TA, Javati A, Kajoi M. A review of measles admissions and deaths in the paediatric ward of Goroka Base Hospital during 1989. Papua and New Guinea Medical Journal. 1991 Mar 1;34(1):6-12. | PNG | 1989 | 0 | 0 | 17 |
| Mgone JM, Mgone CS, Duke TR, Frank DA, Yeka WI. Control measures and the outcome of the measles epidemic of 1999 in the Eastern Highlands Province. Papua and New Guinea medical journal. 2000 Mar 1;43(1-2):91-7. | PNG | 1999 | 0 | 0 | 13 |
| Centers for Disease Control and Prevention (CDC. Emergency measles control activities--Darfur, Sudan, 2004. MMWR. Morbidity and mortality weekly report. 2004 Oct 1;53(38):897-9. | SDN | 2004 | 1 | 0 | 99 |
| Coronado F, Musa N, Ahmed El Tayeb ES, Haithami S, Dabbagh A, Mahoney F, Nandy R, Cairns L. Retrospective measles outbreak investigation: Sudan, 2004. Journal of tropical pediatrics. 2006 Oct 1;52(5):329-34. | SDN | 2003 | 1 | 0 | 99 |
| El Karim O, Salih MA. Morbidity and mortality from measles in an urban community of the Sudan. Annals of Tropical Medicine & Parasitology. 1981 Apr 1;75(2):227-30. | SDN | 1975 | 0 | 0 | 18 |
| Ibrahim SA, Mustafa O, Mukhtar MM, Saleh EA, El Mubarak HS, Abdallah A, El‐Hassan AM, Osterhaus AD, Groen J, De Swart RL, Zijlstra EE. Measles in suburban Khartoum: an epidemiological and clinical study. Tropical Medicine & International Health. 2002 May;7(5):442-9. | SDN | 1998 | 1 | 0.4167 | 14 |
| Sulaiman AA, Elmadhoun WM, Noor SK, Almobarak AO, Bushara SO, Osman MM, Awadalla H, Ahmed MH. An outbreak of measles in gold miners in River Nile State, Sudan, 2011. Eastern Mediterranean Health Journal. 2020 Feb 1;26(2). | SDN | 2011 | 1 | 0 | 99 |
| Aaby P, Whittle H, Cisse B, Samb B, Jensen H, Simondon F. The frailty hypothesis revisited: mainly weak children die of measles. Vaccine. 2001 Dec 12;20(5-6):949-53. | SEN | 1984 | 1 | 0 | 99 |
|  | SEN | 1988 | 1 | 0 | 99 |
|  | SEN | 1992 | 1 | 0 | 99 |
| Aaby P. Influence of cross-sex transmission on measles mortality in rural Senegal. The Lancet. 1992 Aug 15;340(8816):388-91. | SEN | 1984 | 1 | 0 | 18 |
| Cisse B, Aaby P, Simondon F, Samb B, Soumare M, Whittle H. Role of schools in the transmission of measles in rural Senegal: implications for measles control in developing countries. American journal of epidemiology. 1999 Feb 15;149(4):295-301. | SEN | 1994 | 1 | 0.4167 | 30 |
| Pison G, Bonneuil N. Increased risk of measles mortality for children with siblings among the Fula Bande, Senegal. Clinical Infectious Diseases. 1988 Mar 1;10(2):468-70. | SEN | 1985 | 1 | 0 | 11 |
| Pison G. Dynamique d'une population traditionelle: les Peul Bande (Senegal oriental). Institut national d'etudes demographiques. Cahier no. 99. Paris: Presses Universitaires de France, 1982. 1982. | SEN | 1977 | 1 | 0 | 19 |
| Samb B, Aaby P, Whittle H, Seck AM, Simondon F. Decline in measles case fatality ratio after the introduction of measles immunization in rural Senegal. American journal of epidemiology. 1997 Jan 1;145(1):51-7. | SEN | 1984 | 1 | 0 | 99 |
|  | SEN | 1988 | 1 | 0 | 99 |
| Sesay T, Denisiuk O, Zachariah R. Paediatric morbidity and mortality in Sierra Leone. Have things changed after the 2014/2015 Ebola outbreak?. F1000Research. 2019;8. | SLE | 2013 | 0 | 0 | 5 |
|  | SLE | 2014 | 0 | 0 | 5 |
|  | SLE | 2016 | 0 | 0 | 5 |
| Sugerman DE, Fall A, Guigui MT, N'dolie M, Balogun T, Wurie A, Goodson JL. Preplanned national measles vaccination campaign at the beginning of a measles outbreak—Sierra Leone, 2009–2010. The Journal of infectious diseases. 2011 Jul 1;204(suppl_1):S260-9. | SLE | 2009 | 1 | 0 | 99 |
| World Health Organization (WHO). Expanded Programme on Immunization: Epidemiology of Measles in a Rural Community. Wkly Epidemiol Rec. 1980;55(12): 85-7. | SOM | 1978 | 1 | 0 | 99 |
| Grais RF, Dubray C, Gerstl S, Guthmann JP, Djibo A, Nargaye KD, Coker J, Alberti KP, Cochet A, Ihekweazu C, Nathan N. Unacceptably high mortality related to measles epidemics in Niger, Nigeria, and Chad. PLoS medicine. 2007 Jan;4(1):e16. | TCD | 2003 | 1 | 0 | 99 |
| Ndikuyeze A, Cook A, Cutts FT, Bennett S. Priorities in global measles control: report of an outbreak in N'Djamena, Chad. Epidemiology & Infection. 1995 Oct;115(2):309-14. | TCD | 1990 | 1 | 0 | 5 |
| World Health Organization. Measles mortality reduction in West Africa, 1996-2002. Weekly Epidemiological Record= Relevé épidémiologique hebdomadaire. 2003;78(45):390-2. | TGO | 2002 | 1 | 0 | 99 |
| Ariyasriwatana C, Kalayanarooj S. Severity of measles: a study at the Queen Sirikit National Institute of Child Health. Journal of the Medical Association of Thailand. 2004 Jun 1;87(6):581. | THA | 2000 | 0 | 0 | 14 |
| World Health Organization (WHO). Expanded programme on immunization. Measles outbreak among the hill tribes. Wkly Epidemiol Rec. 1985; 60(11): 79. | THA | 1984 | 1 | 0.5833 | 25 |
| Burgess W, Mduma B, Josephson GV. Measles in Mbeya, Tanzania-1981-1983. Journal of tropical pediatrics. 1986;32(4):148-53. | TZA | 1982 | 0 | 0 | 99 |
| Mafigiri R, Nsubuga F, Ario AR. Risk factors for measles death: Kyegegwa District, western Uganda, February–September, 2015. BMC Infectious Diseases. 2017 Dec;17(1):1-7. | UGA | 2015 | 1 | 0.3 | 36 |
| Weeks RM, Barenzi JF, Wayira JR. A low-cost, community-based measles outbreak investigation with follow-up action. Bulletin of the World Health Organization. 1992;70(3):317. | UGA | 1990 | 1 | 0.4167 | 12 |
| Espinosa L, Mirinaviciute G. Health crisis in Venezuela: status of communicable diseases and implications for the European Union and European Economic Area, May 2019. Eurosurveillance. 2019 May 30;24(22):1900308. | VEN | 2017 | 1 | 0 | 99 |
|  | VEN | 2018 | 1 | 0 | 99 |
| Pan American Health Organization / World Health Organization. Epidemiological Update: Measles. 18 January 2019, Washington, D.C.: PAHO/WHO; 2019 | VEN | 2017 | 1 | 0 | 99 |
| Karim SA, Karim QA, Dilraj A, Chamane M. Unsustainability of a measles immunisation campaign-rise in measles incidence within 2 years of the campaign. South African Medical Journal. 1993;83(5). | ZAF | 1989 | 0 | 0 | 99 |
|  | ZAF | 1990 | 0 | 0 | 99 |
|  | ZAF | 1991 | 0 | 0 | 99 |
| Coetzee S, Morrow BM, Argent AC. Measles in a S outh A frican paediatric intensive care unit: Again!. Journal of Paediatrics and Child Health. 2014 May;50(5):379-85. | ZAF | 2010 | 0 | 0.42 | 0.75 |
| Dramowski A, Aucamp M, Bekker A, Mehtar S. Infectious disease exposures and outbreaks at a South African neonatal unit with review of neonatal outbreak epidemiology in Africa. International Journal of Infectious Diseases. 2017 Apr 1;57:79-85. | ZAF | 2010 | 0 | 0 | 0.9999 |
| Gibson, IHN, Carmichael, TR & Kustner HG. Measles notifications-the first year. South African Medical Journal. 1982 Jan 1;61(3):84-8. | ZAF | 1979 | 1 | 0 | 99 |
| Hussey GD, Klein M. Routine high-dose vitamin A therapy for children hospitalized with measles. Journal of tropical pediatrics. 1993 Dec 1;39(6):342-5. | ZAF | 1985 | 0 | 0 | 18 |
| Jeena PM, Wesley AG, Coovadia HM. Infectious diseases at the paediatric isolation units of Clairwood and King Edward VIII hospitals, Durban. South African Medical Journal. 1998;88(7). | ZAF | 1985 | 0 | 0 | 18 |
|  | ZAF | 1986 | 0 | 0 | 18 |
|  | ZAF | 1987 | 0 | 0 | 18 |
|  | ZAF | 1988 | 0 | 0 | 18 |
|  | ZAF | 1989 | 0 | 0 | 18 |
|  | ZAF | 1990 | 0 | 0 | 18 |
|  | ZAF | 1991 | 0 | 0 | 18 |
|  | ZAF | 1992 | 0 | 0 | 18 |
|  | ZAF | 1993 | 0 | 0 | 18 |
|  | ZAF | 1994 | 0 | 0 | 18 |
|  | ZAF | 1995 | 0 | 0 | 18 |
|  | ZAF | 1996 | 0 | 0 | 18 |
| Le Roux DM, Le Roux SM, Nuttall JJ, Eley BS. South African measles outbreak 2009-2010 as experienced by a paediatric hospital. South African Medical Journal. 2012;102(9):760-4. | ZAF | 2009 | 0 | 0 | 18 |
| Loening WE, Coovadia HM. Age-specific occurrence rates of measles in urban, peri-urban, and rural environments: implications for time of vaccination. The Lancet. 1983 Aug 6;322(8345):324-6. | ZAF | 1980 | 0 | 0 | 17 |
| McMorrow ML, Gebremedhin G, Van den Heever J, Kezaala R, Harris BN, Nandy R. Measles outbreak in South Africa, 2003-2005. South African Medical Journal. 2009;99(5). | ZAF | 2004 | 1 | 0 | 99 |
|  | ZAF | 2005 | 1 | 0 | 99 |
| Uzicanin A, Eggers R, Webb E, Harris B, Durrheim D, Ogunbanjo G, Isaacs V, Hawkridge A, Biellik R, Strebel P. Impact of the 1996–1997 supplementary measles vaccination campaigns in South Africa. International journal of epidemiology. 2002 Oct 1;31(5):968-76. | ZAF | 1989 | 1 | 0 | 99 |
| Centers for Disease Control and Prevention (CDC. Measles incidence before and after supplementary vaccination activities--Lusaka, Zambia, 1996-2000. MMWR. Morbidity and mortality weekly report. 2001 Jun 22;50(24):513-6. | ZMB | 1996 | 0 | 0 | 99 |
|  | ZMB | 1997 | 0 | 0 | 99 |
|  | ZMB | 1998 | 0 | 0 | 99 |
|  | ZMB | 1999 | 0 | 0 | 99 |
| Moss WJ, Monze M, Ryon JJ, Quinn TC, Griffin DE, Cutts F. Prospective Study of Measles in Hospitalized, Human Immunodeficiency Virus (HIV)—Infected and HIV—Uninfected Children in Zambia. Clinical infectious diseases. 2002 Jul 15;35(2):189-96. | ZMB | 1999 | 0 | 0 | 18 |
| Oshitani H, Mpabalwani M, Kasolo F, Mizuta K, Luo NP, Bhat GJ, Suzuki H, Numazaki Y. Measles infection in hospitalized children in Lusaka, Zambia. Annals of tropical paediatrics. 1995 Jun 1;15(2):167-72. | ZMB | 1992 | 0 | 0 | 15 |
| Rolfe M. Measles immunization in the Zambian Copperbelt: cause for concern. Transactions of the Royal Society of Tropical Medicine and Hygiene. 1982 Jan 1;76(4):529-30. | ZMB | 1980 | 1 | 0 | 4 |
| Kambarami RA, Nathoo KJ, Nkrumah FK, Pirie DJ. Measles epidemic in Harare, Zimbabwe, despite high measles immunization coverage rates. Bulletin of the World Health Organization. 1991;69(2):213. | ZWE | 1988 | 1 | 0.1154 | 30 |
| Marufu T, Siziya S, Tshimanga M, Murugasampillay S, Mason E, Manyume B. Factors associated with measles complications in Gweru, Zimbabwe. East African medical journal. 2001;78(3):135-8. | ZWE | 1984 | 1 | 0 | 99 |
| Nsungu M. Measles vaccination status, delay in recognizing measles outbreaks and outbreak outcome. The Central African journal of medicine. 1995 Nov 1;41(11):336-9. | ZWE | 1994 | 0 | 0 | 24 |
| Uyirwoth GP. Measles in Mashonaland Central Province: Zimbabwe. East African medical journal. 1993 Jul 1;70(7):455-9. | ZWE | 1987 | 0 | 0 | 99 |
|  | ZWE | 1988 | 0 | 0 | 99 |
|  | ZWE | 1989 | 0 | 0 | 99 |

#### **Supplementary Table 3.** Second step model fitted meta-regression coefficients.

| **Variable** | **Coefficient values** | **p-values** |
| --- | --- | --- |
| Intercept | -2.1084 | <0.0001 |
| Community indicator | -1.4597 | <0.0001 |
| Incidence | 0.2513 | <0.0001 |
| Mortality rate due to war and terrorism | 0 | 1.0000 |
| Maternal education | -0.6915 | <0.0001 |
| GDP per capita | 0 | 1.0000 |
| HIV prevalence | 0 | 1.0000 |
| MCV1 coverage | -0.1581 | <0.0001 |
| Total fertility | 0 | 1.0000 |
| Under 5 mortality rate | 0.1140 | <0.0001 |
| Proportion living in urban setting | 0.4161 | <0.0001 |
| Vitamin A deficiency prevalence | 0.0652 | <0.0001 |
| Wasting prevalence | 0 | 1.0000 |

#### **Supplementary Table 4.** In-sample validation metrics from first stage decomposition analysis.

| IS | Model 0 | Model 1.A | Model 1.B | Model 1.C |
| --- | --- | --- | --- | --- |
| Correlation | 0.29 | 0.2785 | 0.2776 | 0.3415 |
| RMSE | 0.0616 | 0.0629 | 0.0631 | 0.0598 |
| Mean Error | 0.0104 | 0.0218 | 0.0202 | 0.0075 |
| Mean Abs. Error | 0.0356 | 0.0354 | 0.0358 | 0.0354 |

#### **Supplementary Table 5.** Out-of-sample validation metrics from first stage decomposition analysis.

| OOS | Model 0 | Model 1.A | Model 1.B | Model 1.C |
| --- | --- | --- | --- | --- |
| Correlation | 0.2897 | 0.2783 | 0.2775 | 0.3376 |
| RMSE | 0.0624 | 0.0635 | 0.0641 | 0.0608 |
| Mean Error | 0.0100 | 0.0200 | 0.0182 | 0.0052 |
| Mean Abs. Error | 0.0357 | 0.0357 | 0.0362 | 0.0364 |

#### **Supplementary Table 6.** In-sample validation metrics from first stage decomposition analysis.

| IS | Model 0 | Model 1 | Model 2 |
| --- | --- | --- | --- |
| Correlation | 0.29 | 0.3415 | 0.3929 |
| RMSE | 0.0616 | 0.0598 | 0.0591 |
| Mean Error | 0.0104 | 0.0075 | -0.0010 |
| Mean Abs. Error | 0.0356 | 0.0354 | 0.0349 |

#### **Supplementary Table 7.** Out-of-sample validation metrics from first and second stage decomposition analysis.

| OOS | Model 0 | Model 1 | Model 2 |
| --- | --- | --- | --- |
| Correlation | 0.2897 | 0.3376 | 0.3833 |
| RMSE | 0.0624 | 0.0608 | 0.0616 |
| Mean Error | 0.0100 | 0.0052 | -0.0022 |
| Mean Abs. Error | 0.0357 | 0.0364 | 0.0362 |

#### **Supplementary Table 8.** In-sample (IS) and out-of-sample (OOS) validation metrics from final model using age split input data for comparison.

|  | IS | OOS |
| --- | --- | --- |
| Correlation | 0.5002 | 0.4517 |
| RMSE | 0.0451 | 0.0270 |
| Mean Error | 0.0035 | 0.0011 |
| Mean Abs. Error | 0.0175 | 0.0110 |

#### **Supplementary Table 9.** In-sample (IS) and out-of-sample (OOS) validation metrics from final model using original (pre-age split) data for comparison.

|  | IS | OOS |
| --- | --- | --- |
| Correlation | 0.4667 | 0.2754 |
| RMSE | 0.0934 | 0.0820 |
| Mean Error | 0.0156 | 0.0027 |
| Mean Abs. Error | 0.0343 | 0.0325 |

### ***Section 7.*** *Supplementary Figures*

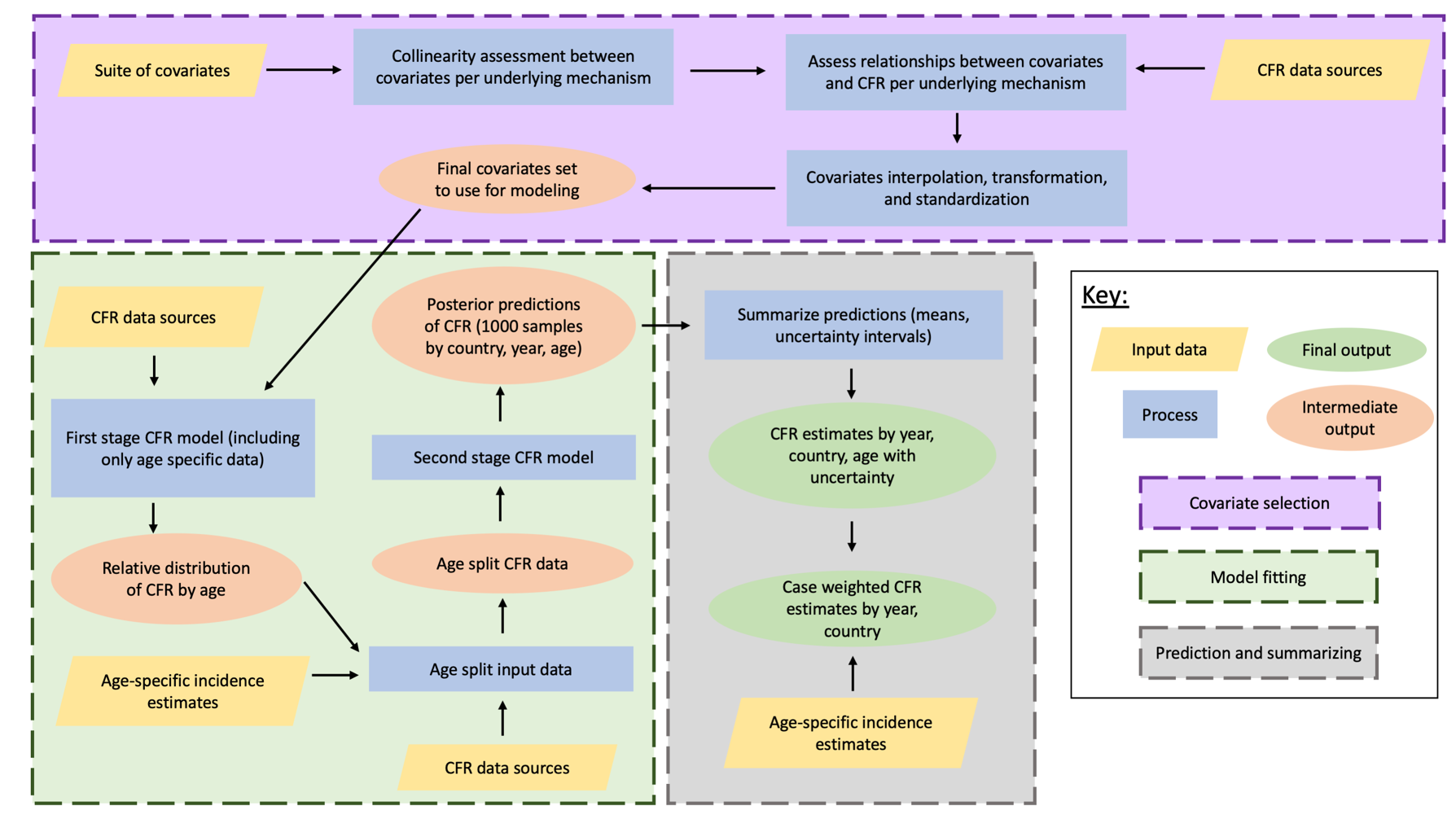

#### **Supplementary Figure 1.** Overview of modeling process.

#### **Supplementary Figure 2.** Relative age pattern from first-stage model with 4 knots (with 2 internal).

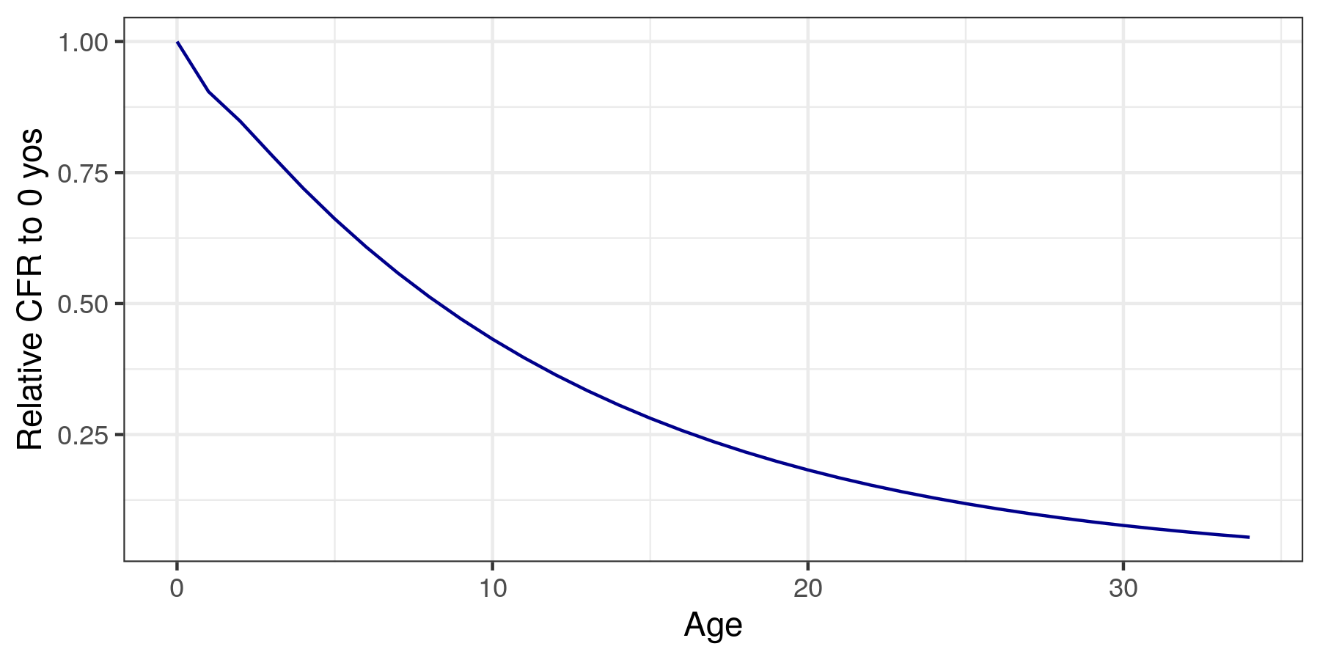

#### **Supplementary Figure 3.** Relative age pattern from first-stage model with 5 knots (with 3 internal).

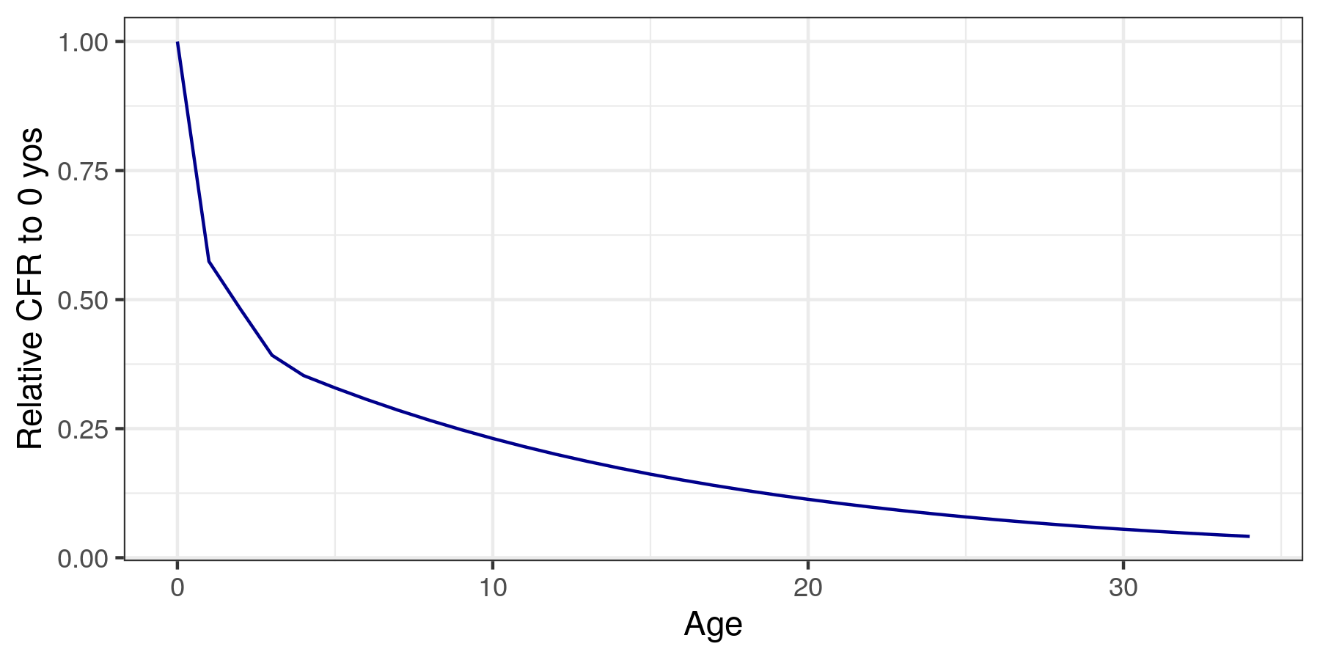

#### **Supplementary Figure 4.** Relationship between age of input data and standardized measles incidence from country-year input data was collected.

Grey lines represent a smooth loess curve, and black lines represent a loess curve weighted on standard error of each input data.

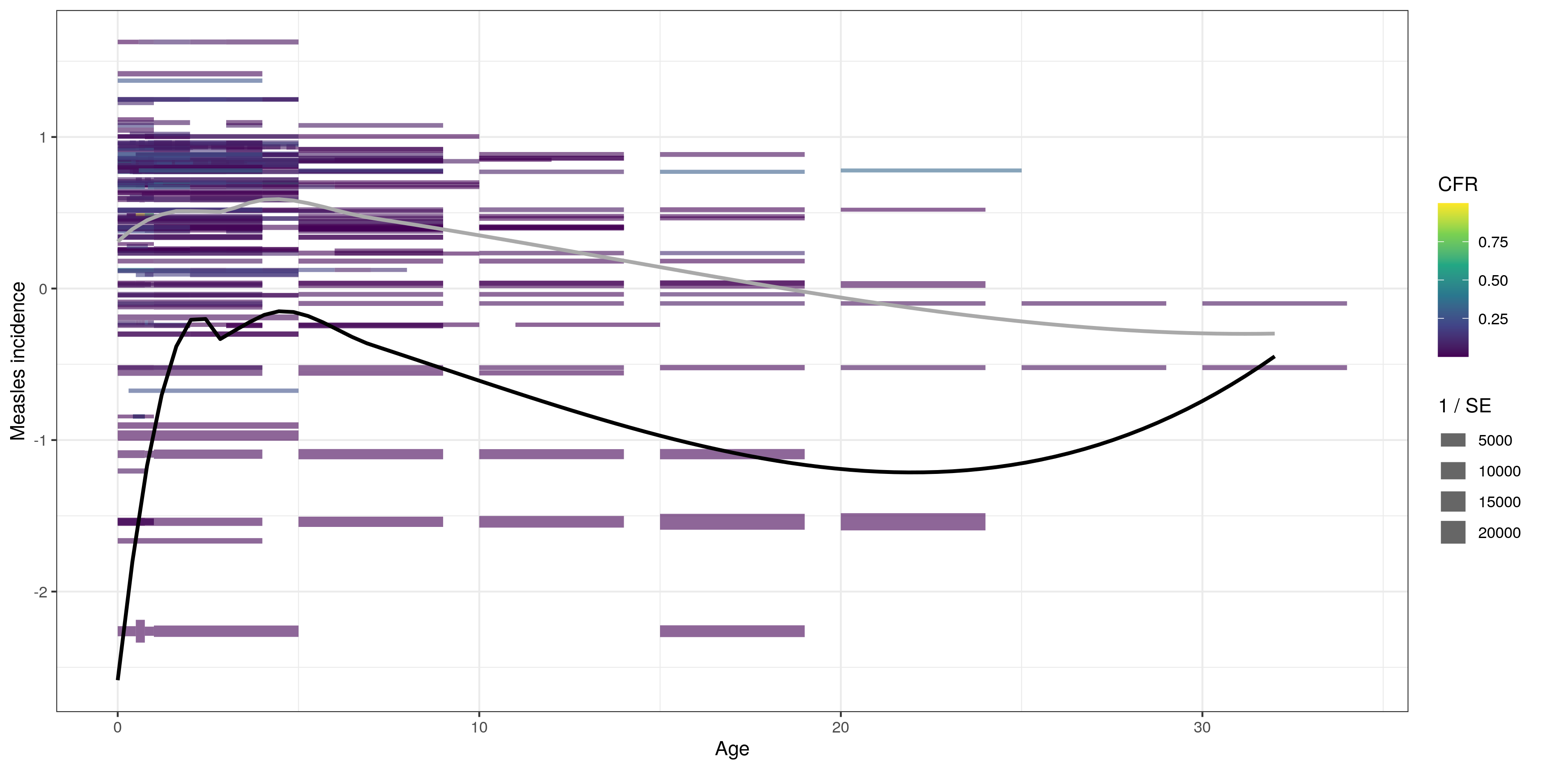

#### **Supplementary Figure 5.** Relationship between age of input data and standardized first-dose measles-containing vaccine (MCV1) coverage from country-year input data was collected.

Grey lines represent a smooth loess curve, and black lines represent a loess curve weighted on standard error of each input data.

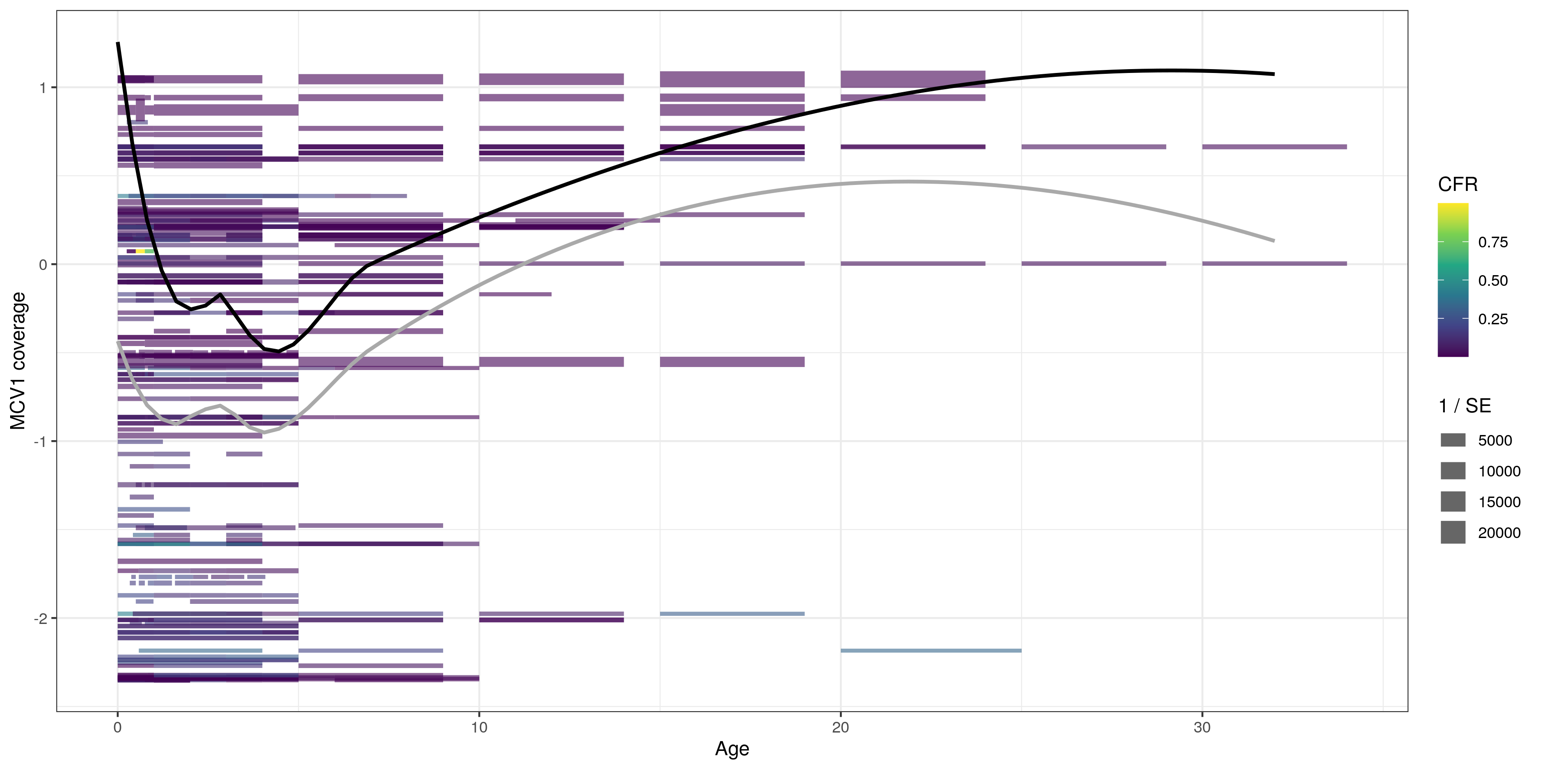

#### **Supplementary Figure 6.** Average age of measles cases by country and year.

Grey lines represent the average age of cases by year for each country included in analysis, and the red line is a smoothed LOESS curve through individual country lines.

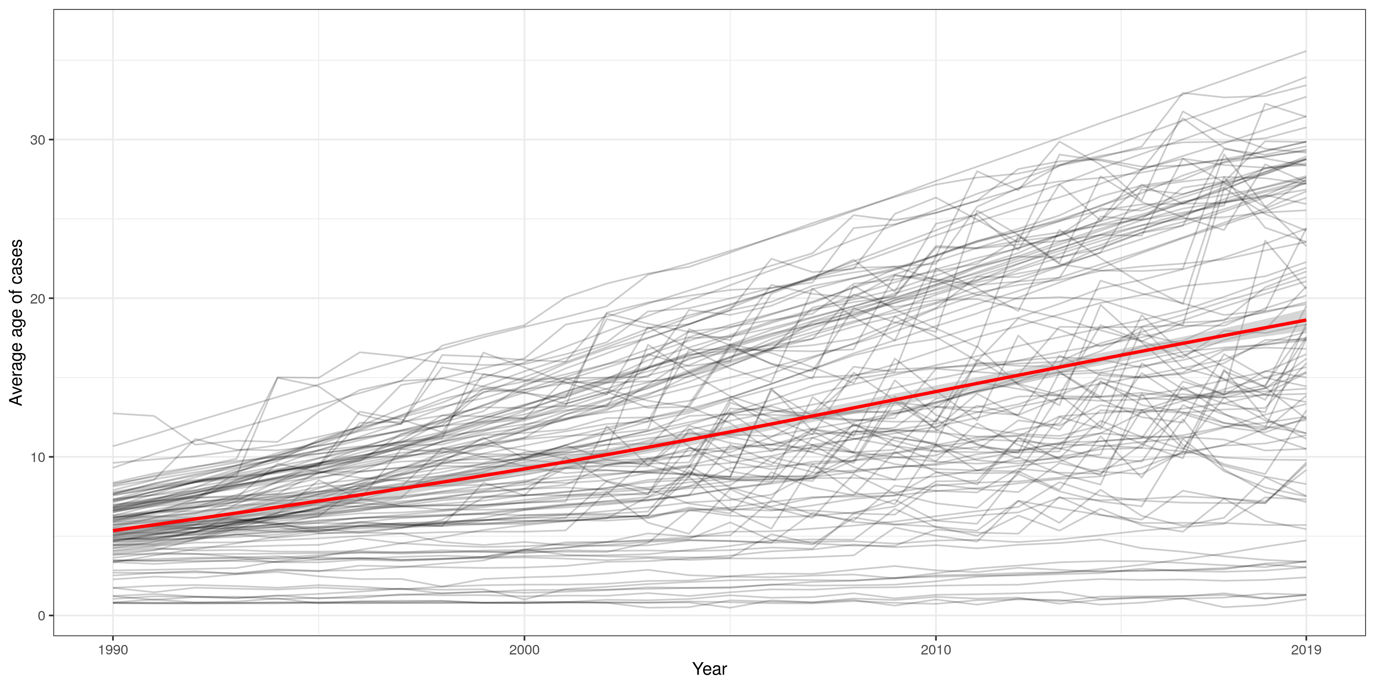

#### **Supplementary Figure 7.** Standardized and unstandardized estimates of case-weighted measles CFR across all countries from 1990 to 2019.

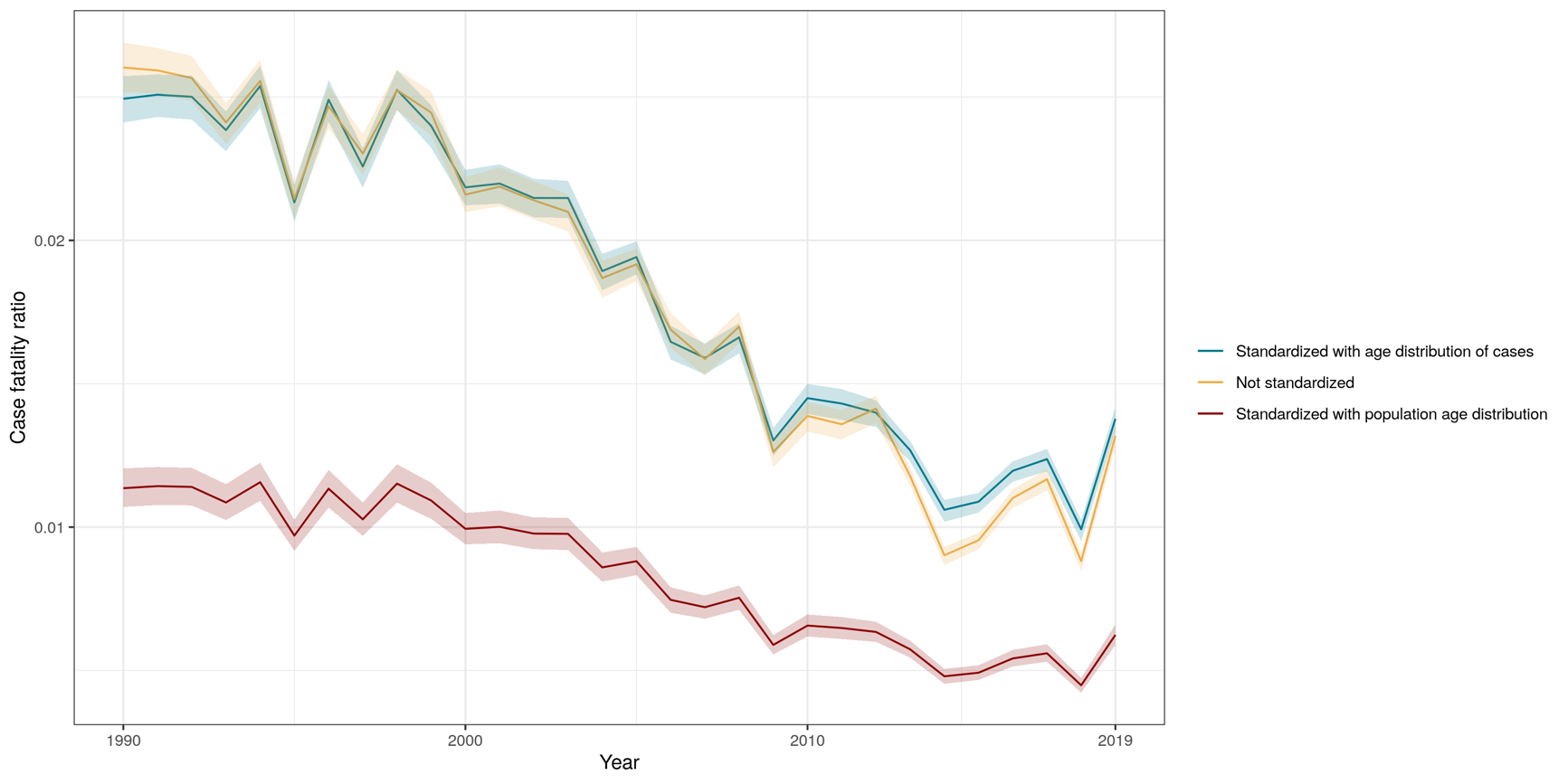
Case-weighted mean CFR across LMICs is presented in yellow, by year. Using the UN standard population from 2019, we age-standardized case-weighted mean CFR estimates for LMICs (shown in red). In blue, we additionally age-standardized case-weighted mean CFR estimates using the age distribution across cases in LMICs in 1990 as our “standard population”.

#### **Supplementary Figure 8.** Distribution of CFR values for studies providing information on laboratory confirmation of cases (1) versus not providing information on laboratory confirmation of cases (0).

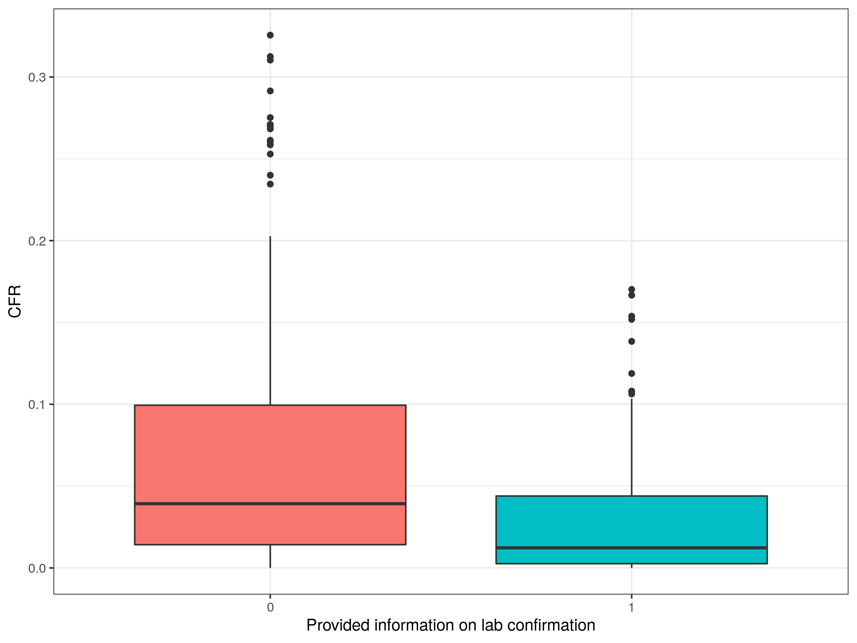

#### **Supplementary Figure 9.** Distribution measles incidence values used for covariates of country-years for studies providing information on laboratory confirmation of cases (1) versus not providing information on laboratory confirmation of cases (0).

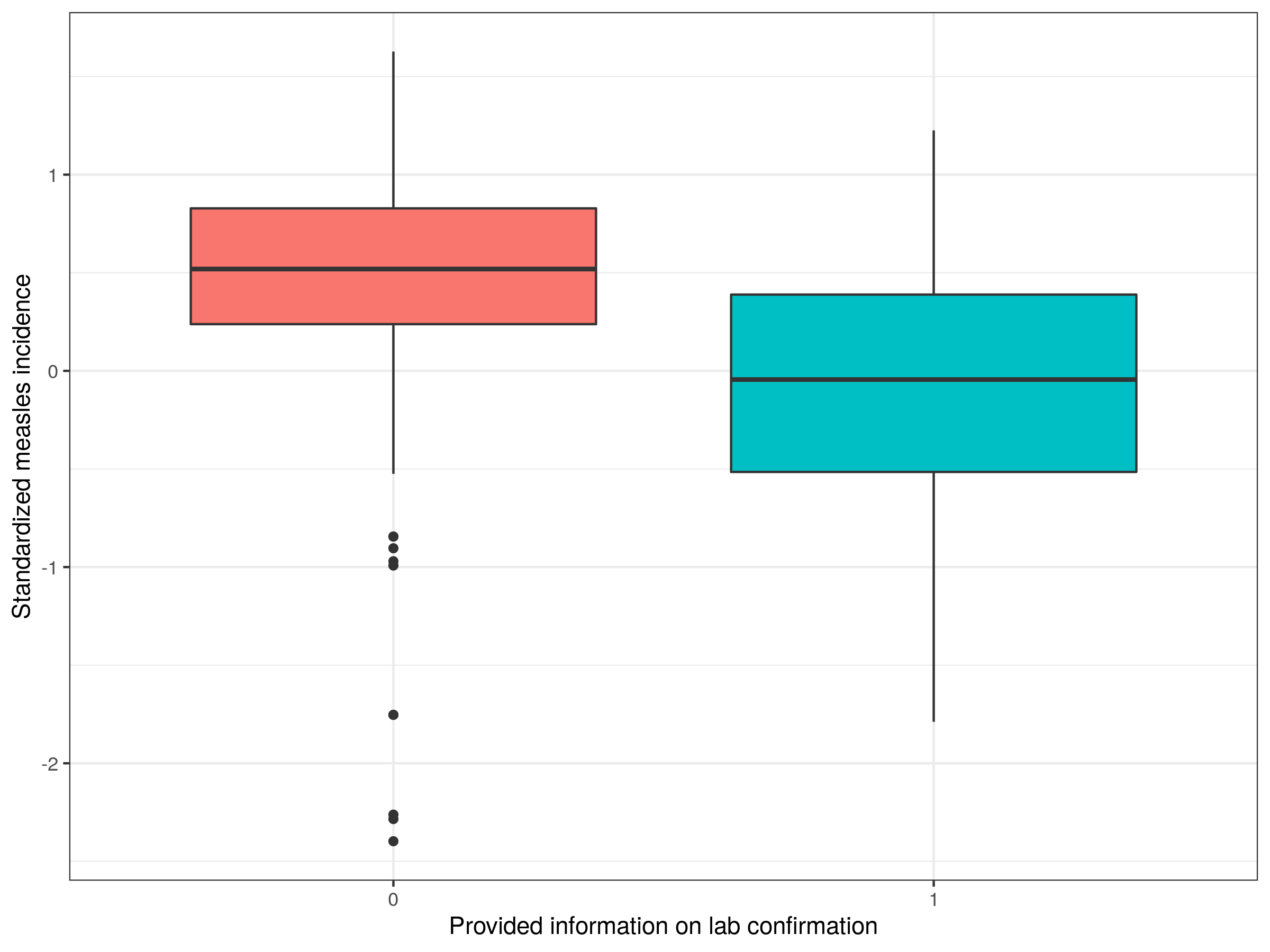

#### **Supplementary Figure 10.** Distribution MCV1 coverage values used for covariates of country-years for studies providing information on laboratory confirmation of cases (1) versus not providing information on laboratory confirmation of cases (0).

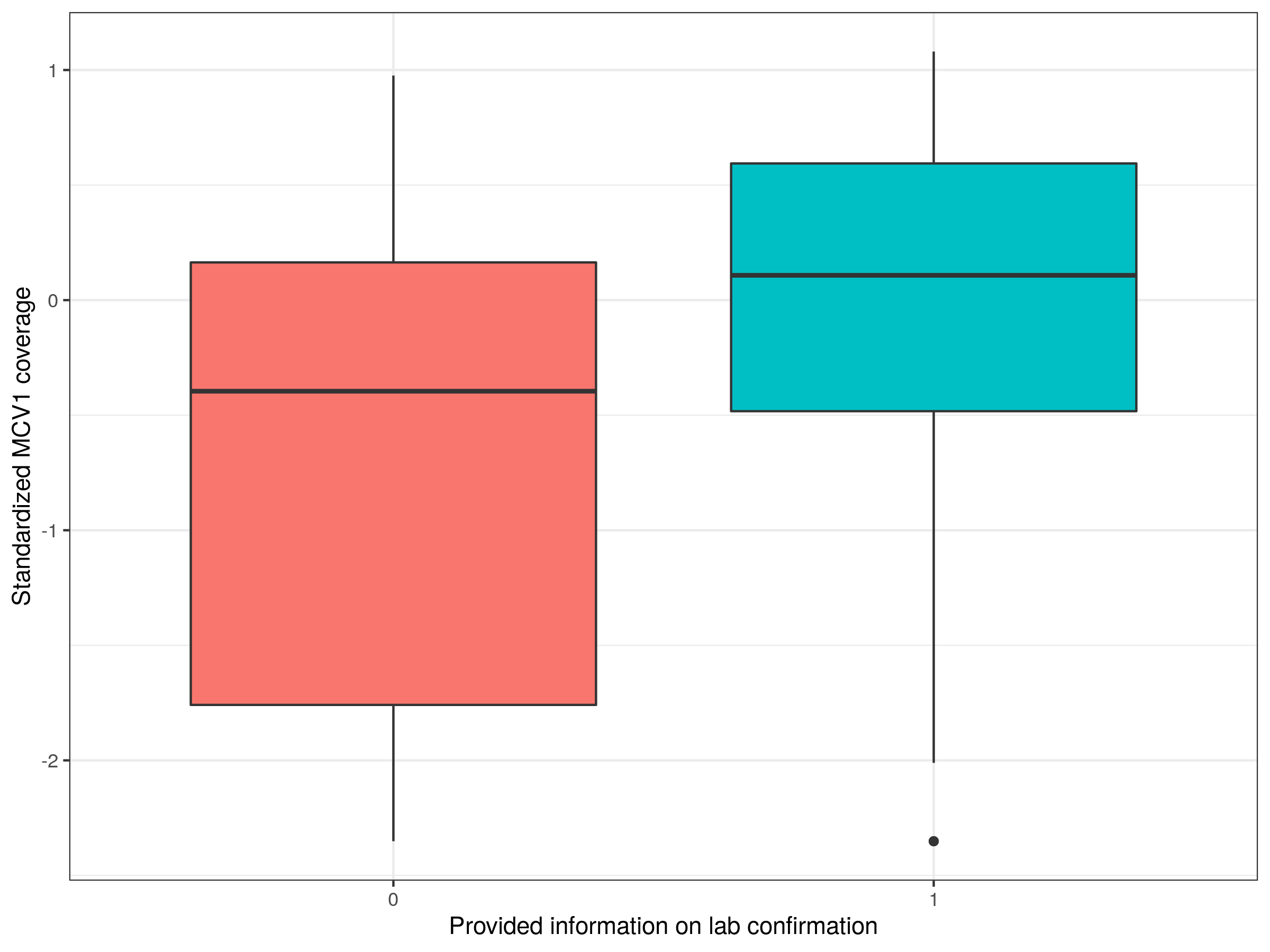

#### **Supplementary Figure 11.** CFR estimates from framework using all studies versus only studies providing information on laboratory confirmation of cases, for select years.

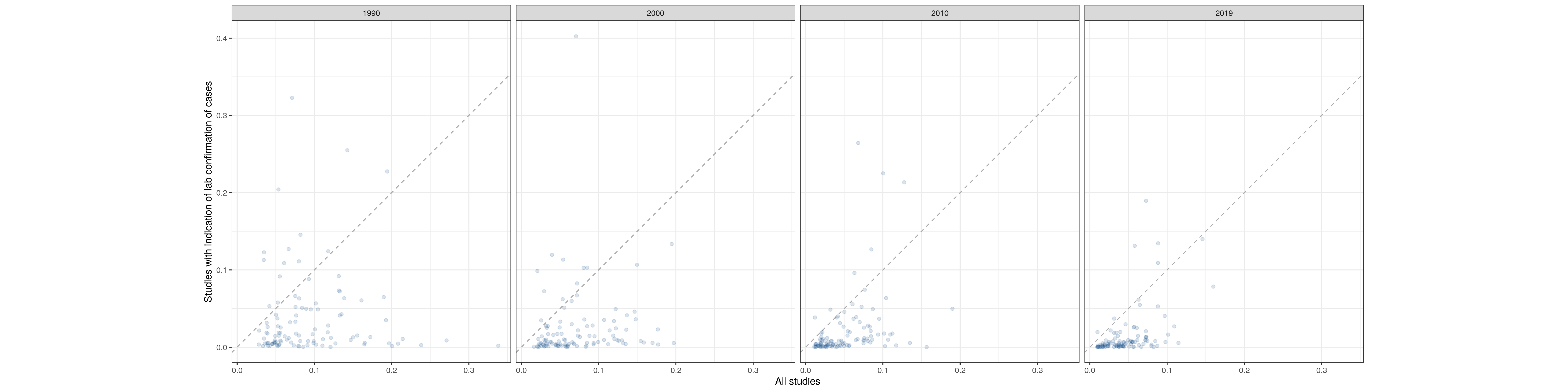

#### **Supplementary Figure 12.** CFR estimates from framework using all studies versus only studies providing definition of death attributable to measles, for select years.

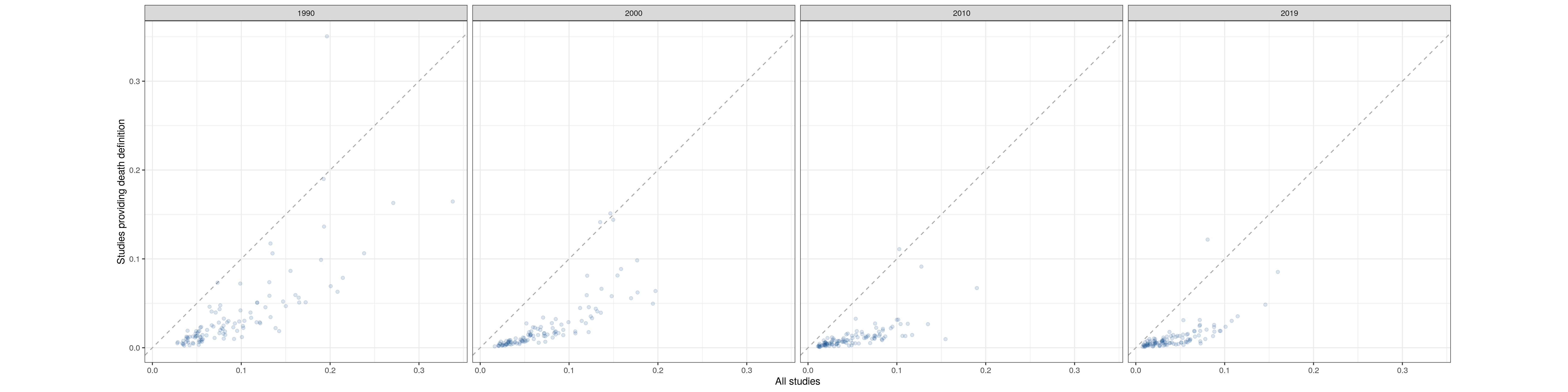

### **References**

1.	Zheng P, Barber R, Sorensen RJ, Murray CJ. Trimmed constrained mixed effects models: formulations and algorithms. *Journal of Computational and Graphical Statistics* 2021; **30**(3): 1-13.

2.	Portnoy A, Jit M, Ferrari M, Hanson M, Brenzel L, Verguet S. Estimates of case-fatality ratios of measles in low-income and middle-income countries: a systematic review and modelling analysis. *Lancet Glob Health* 2019; **7**(4): E472-E81.
