## Supplementary Information 2 for "Estimating national-level measles case fatality ratios: an updated systematic review and modelling study"

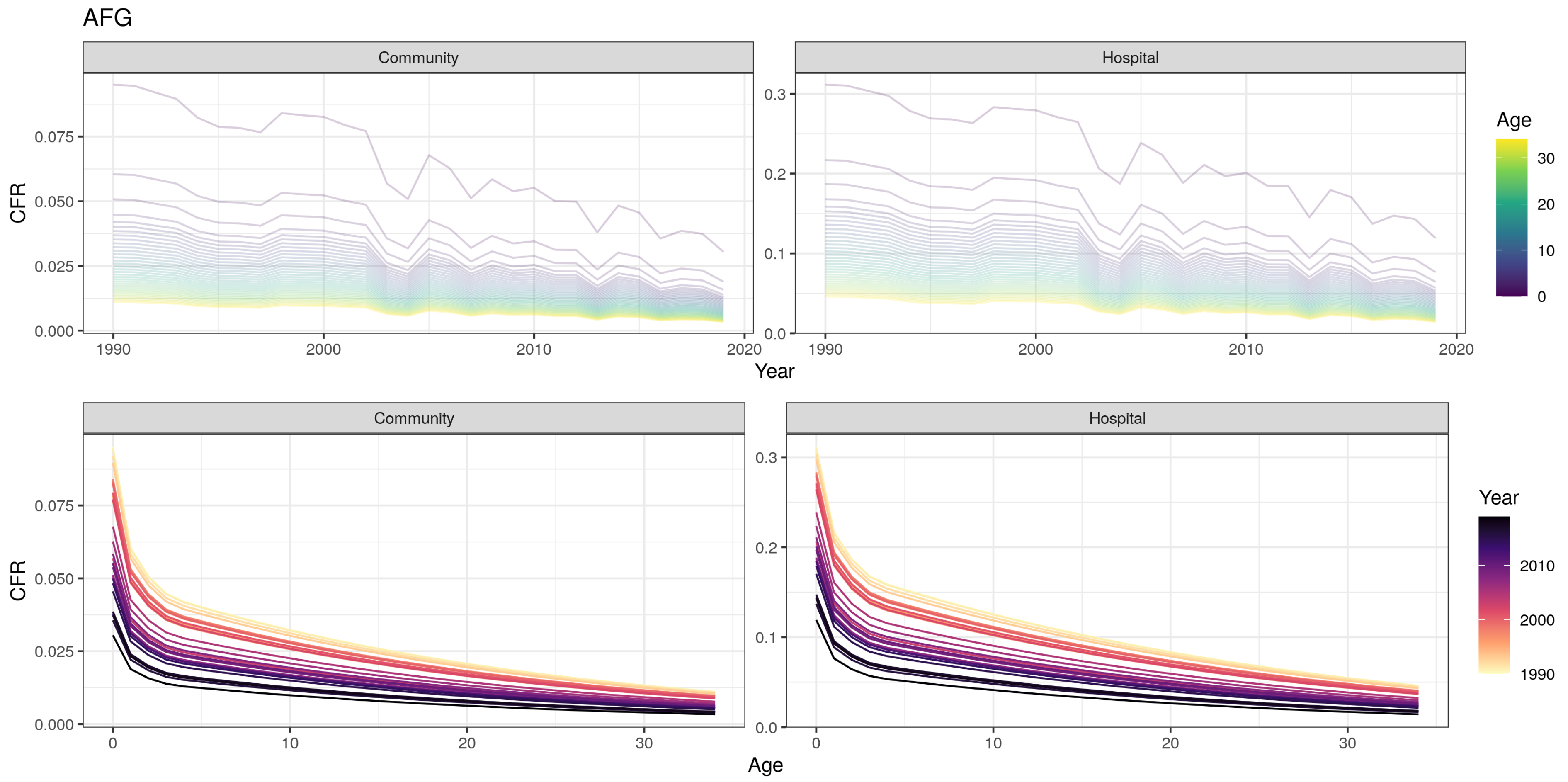
AFG
Community
Hospital
CFR
Year
Age
Community
Hospital
CFR
Age
Year
0.000
0.025
0.050
0.075
0.0
0.1
0.2
0.3
1990
2000
2010
2020
0
10
20
30
1990
2000
2010

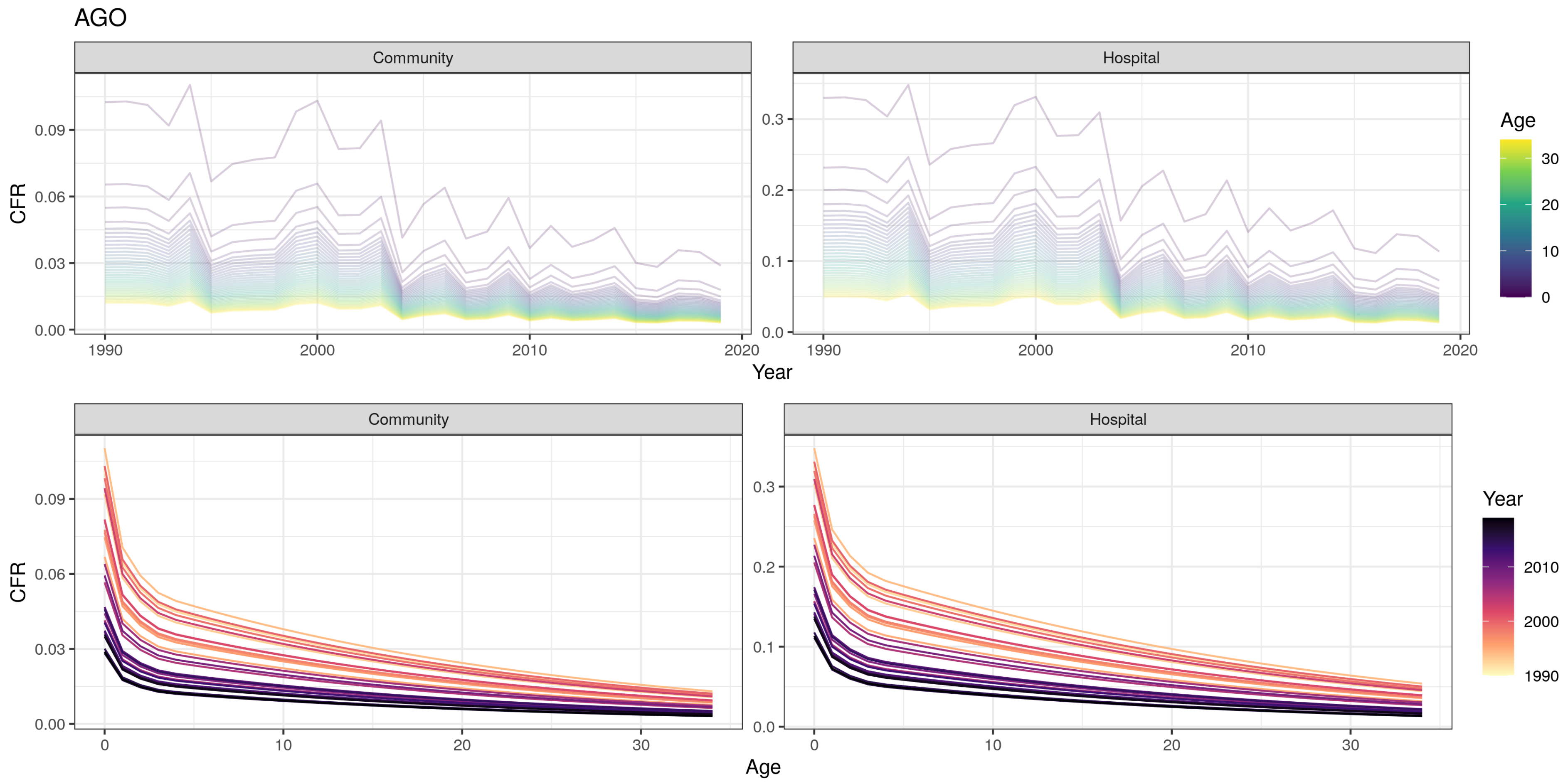

AGO
Community
Hospital
Age
CFR
Year
Community
Hospital
CFR
Age
Year

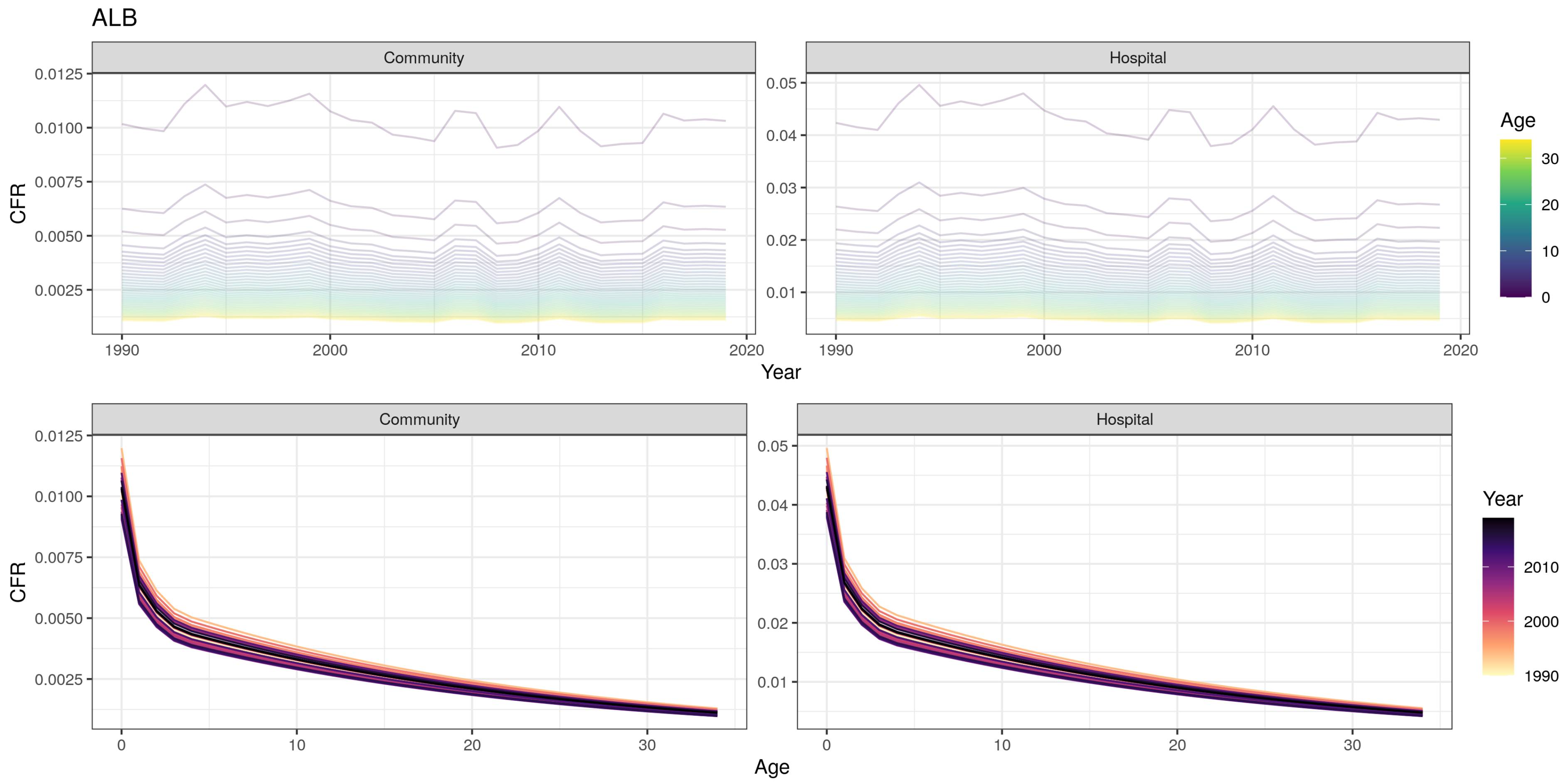
ALB
Community
Hospital
CFR
Year
Age
Community
Hospital
CFR
Age
Year
0.0125
0.0100
0.0075
0.0050
0.0025
0.05
0.04
0.03
0.02
0.01
1990
2000
2010
2020
0
10
20
30

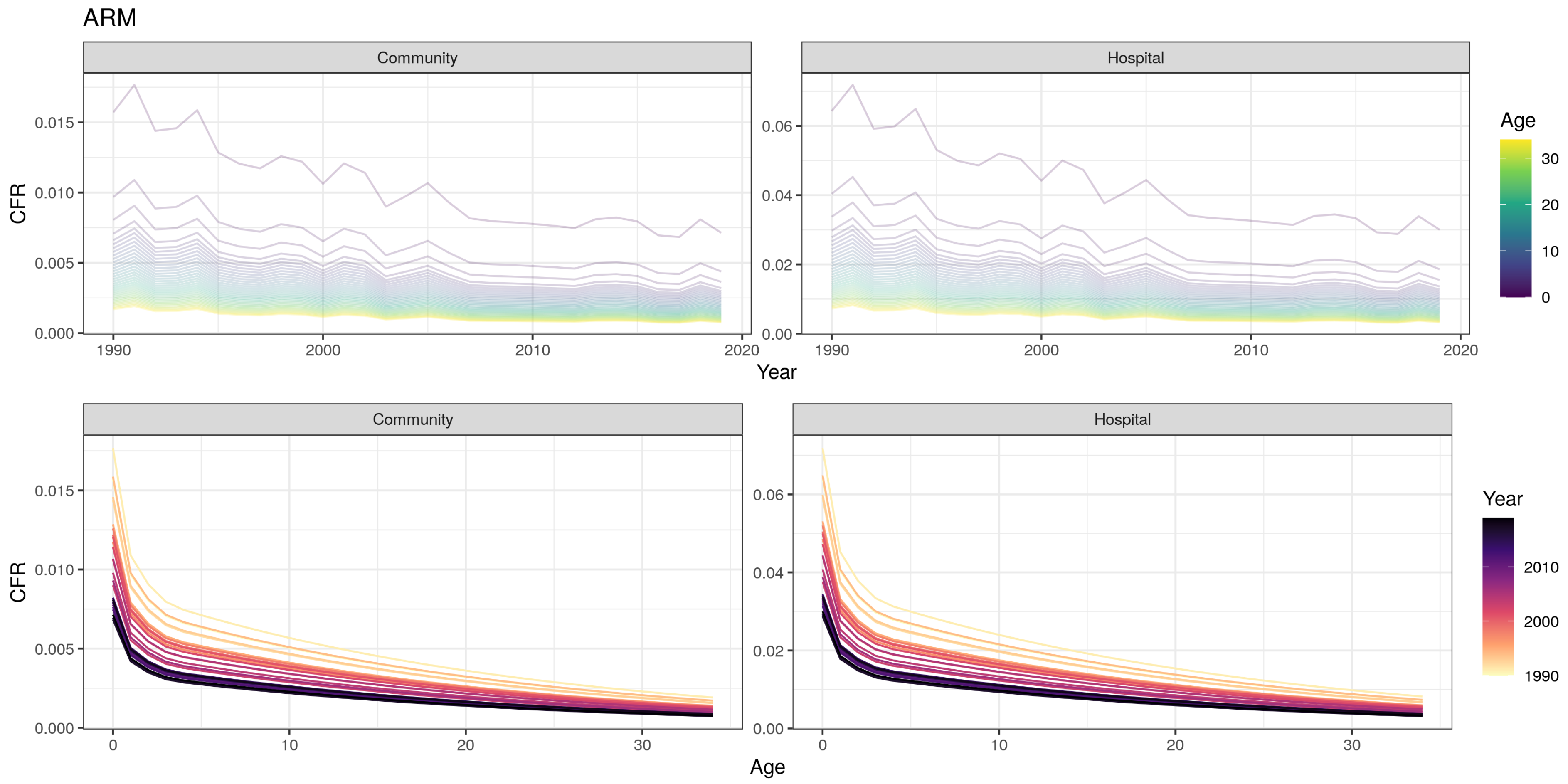

ARM
Community
Hospital
CFR
Year
Age
Community
Hospital
CFR
Age
Year
0.000
0.005
0.010
0.015
0.00
0.02
0.04
0.06
1990
2000
2010
2020
0
10
20
30
1990
2000
2010

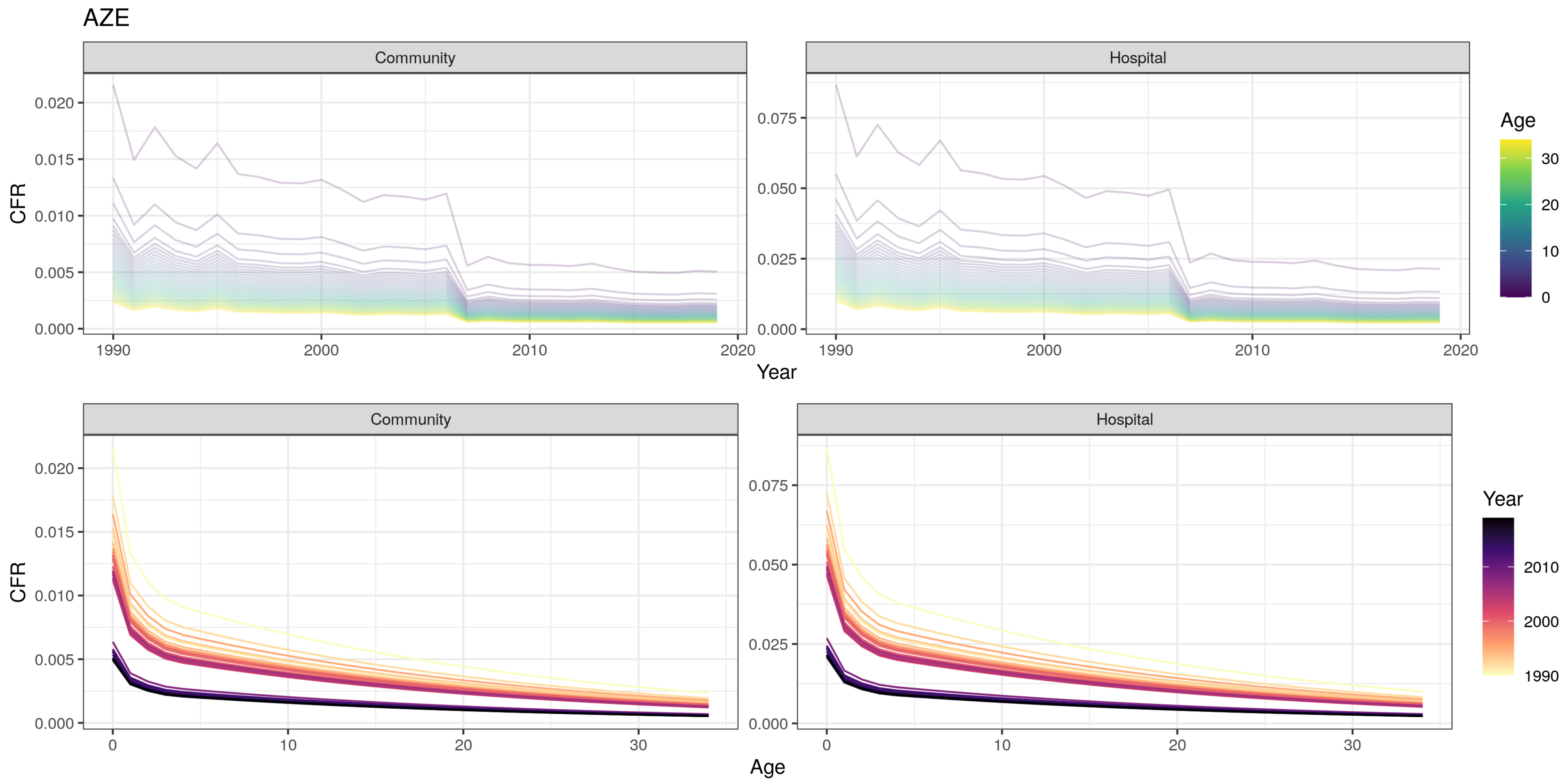
AZE
Community
Hospital
CFR
Year
Age
Community
Hospital
CFR
Age
Year
0.000
0.005
0.010
0.015
0.020
0.025
0.050
0.075
1990
2000
2010
2020
0
10
20
30

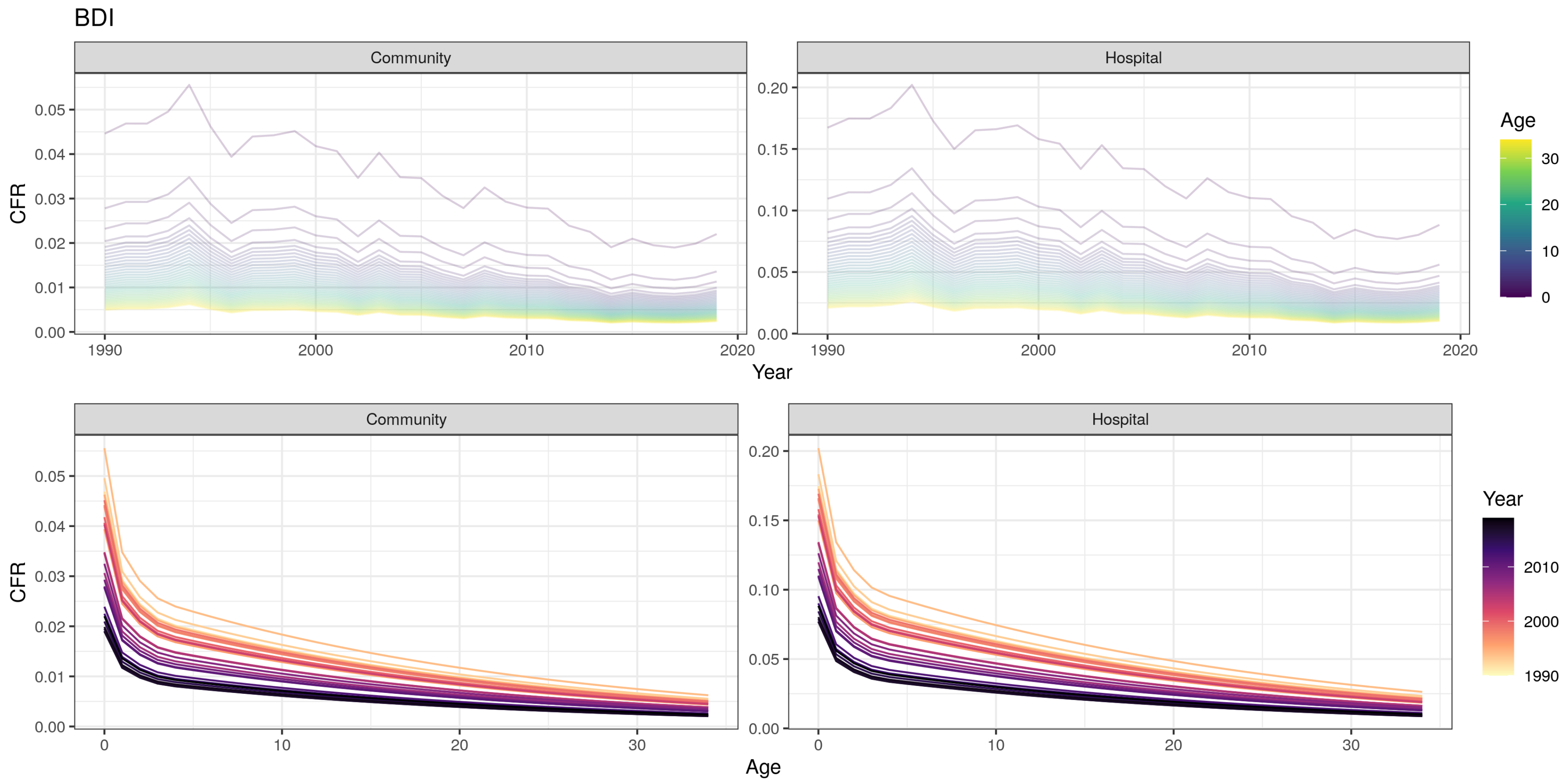

BDI
Community
Hospital
CFR
Year
Age
0.00
0.01
0.02
0.03
0.04
0.05
0.10
0.15
0.20
1990
2000
2010
2020
0
10
20
30

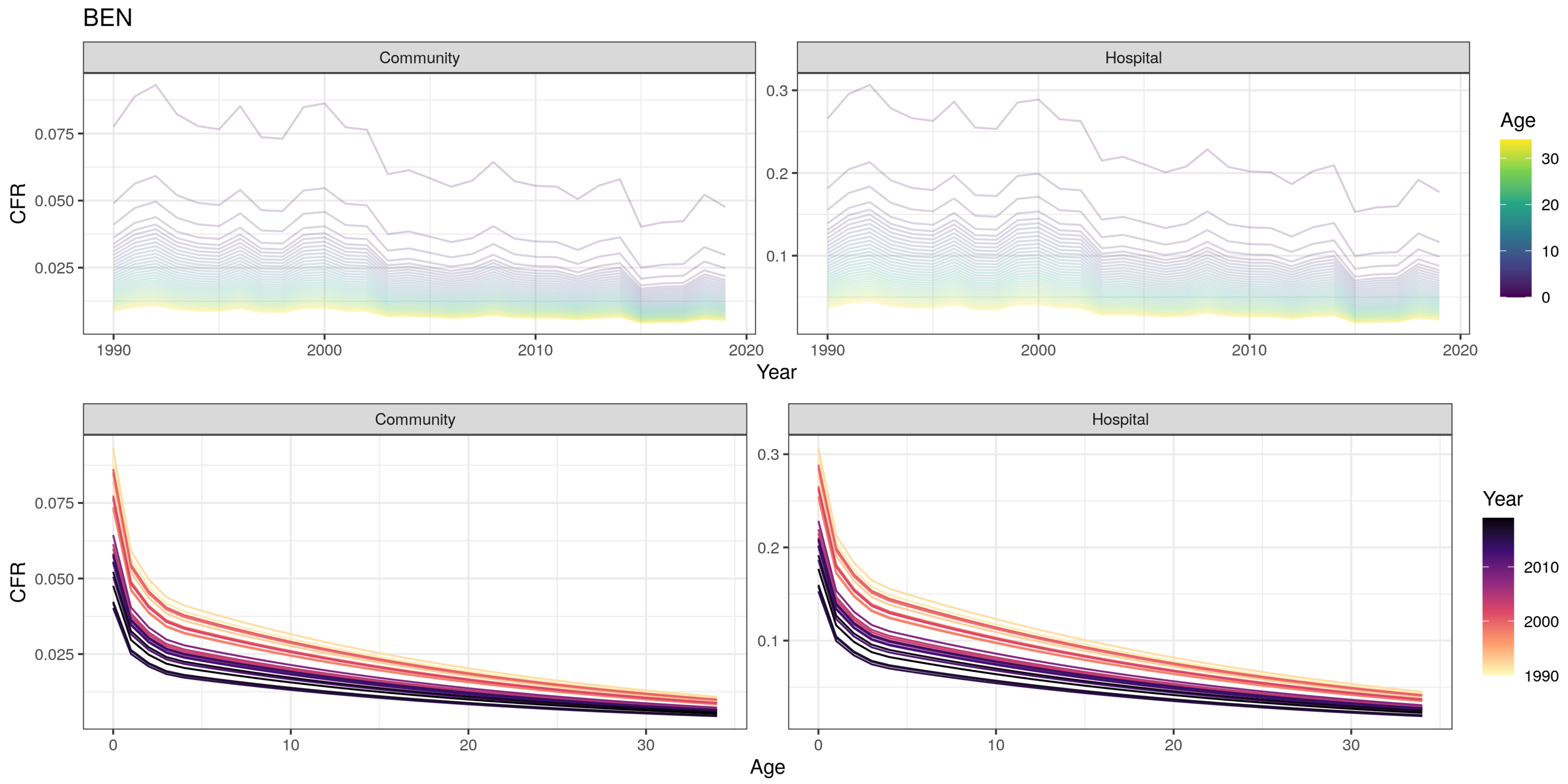

BEN
Community
Hospital
CFR
0.075
0.050
0.025
0.3
0.2
0.1
1990
2000
2010
2020
Year
Age
30
20
10
0
Community
Hospital
CFR
0.075
0.050
0.025
0.3
0.2
0.1
0
10
20
30
Age
Year
2010
2000
1990

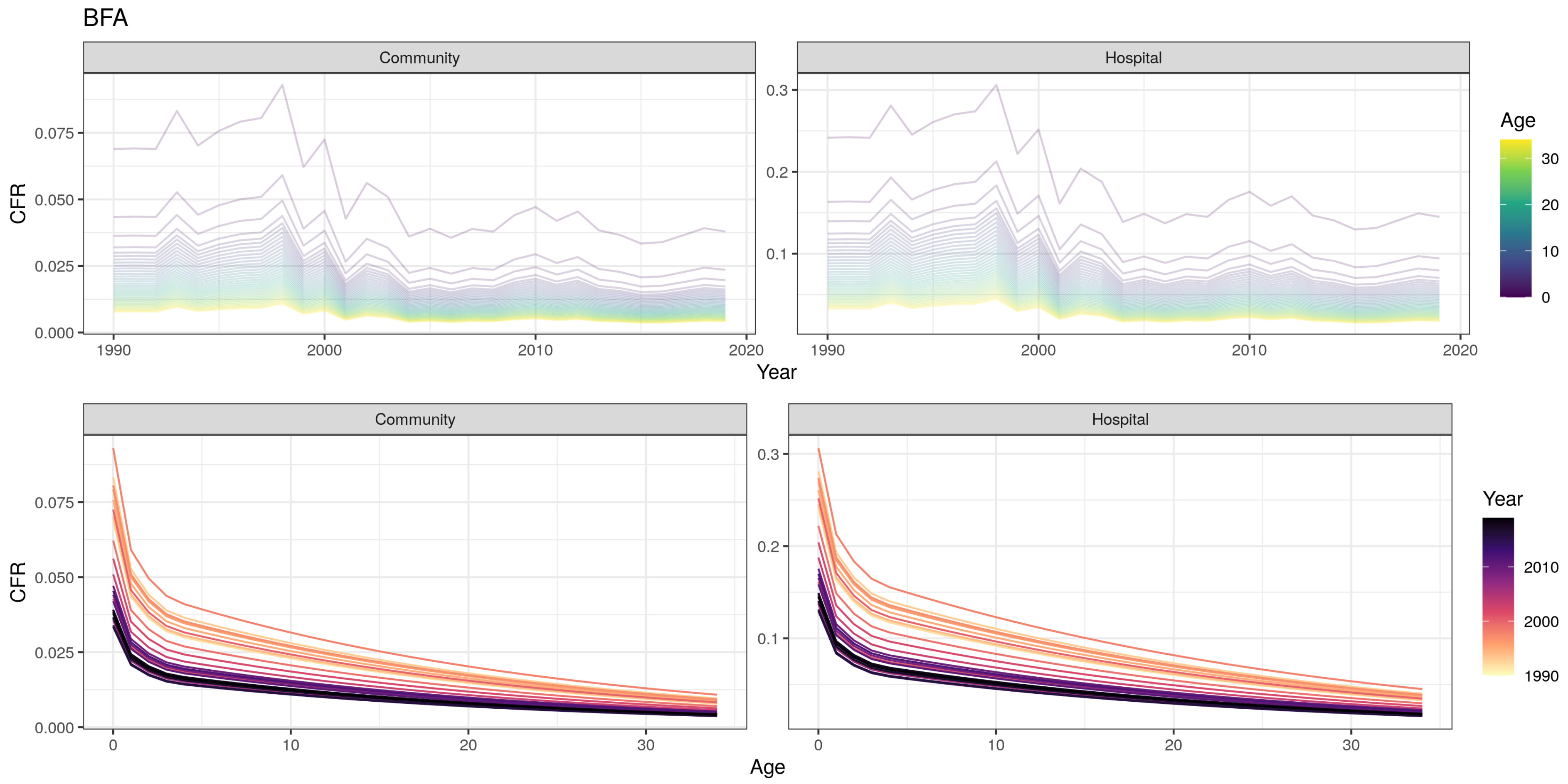
BFA
Community
Hospital
CFR
Year
Age
0.000
0.025
0.050
0.075
0.1
0.2
0.3
1990
2000
2010
2020
0
10
20
30

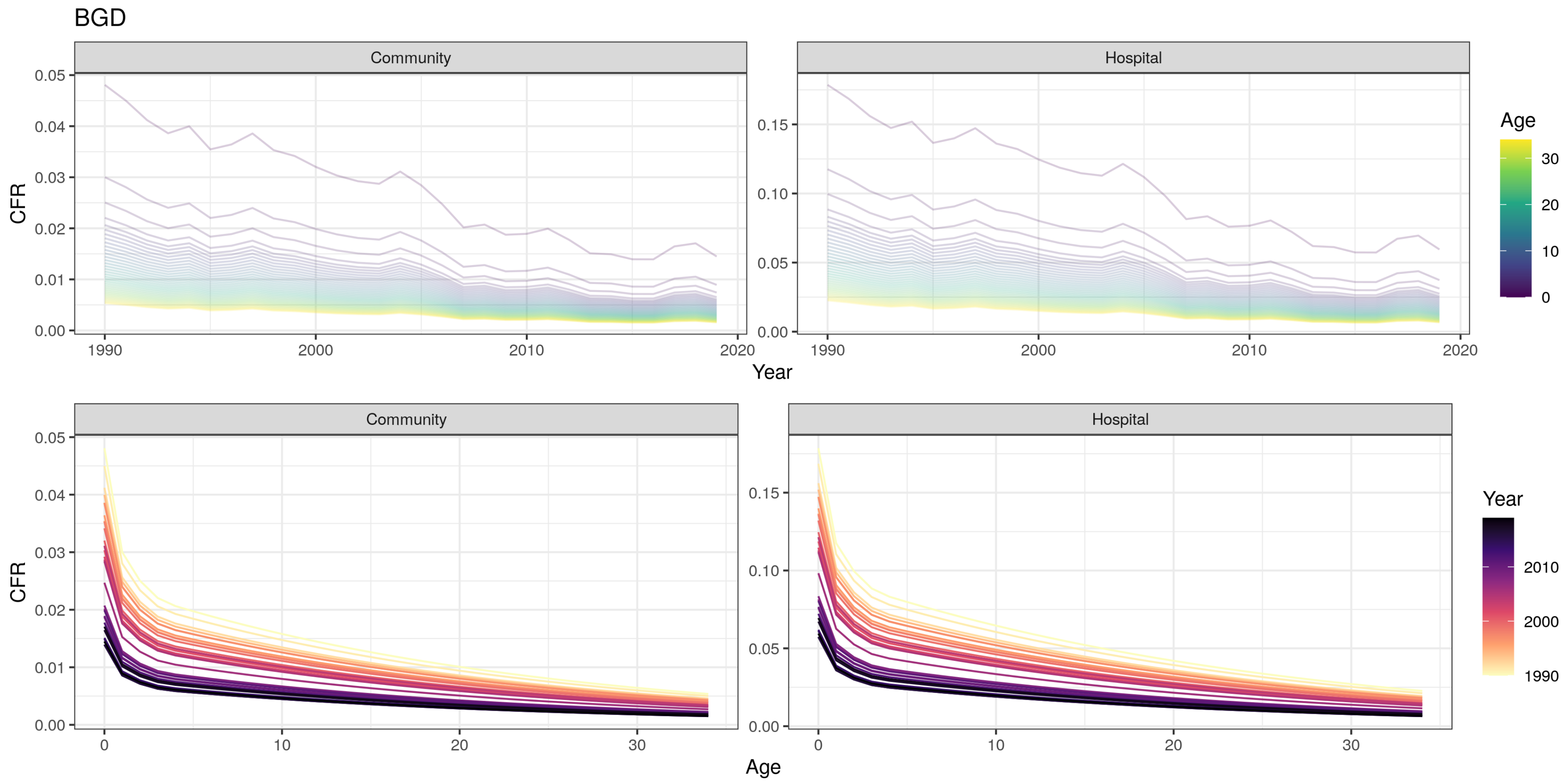
BGD
Community
Hospital
CFR
Year
Age
0.00
0.01
0.02
0.03
0.04
0.05
0.10
0.15
1990
2000
2010
2020
0
10
20
30
Community
Hospital
CFR
Age
Year
1990
2000
2010
0
10
20
30

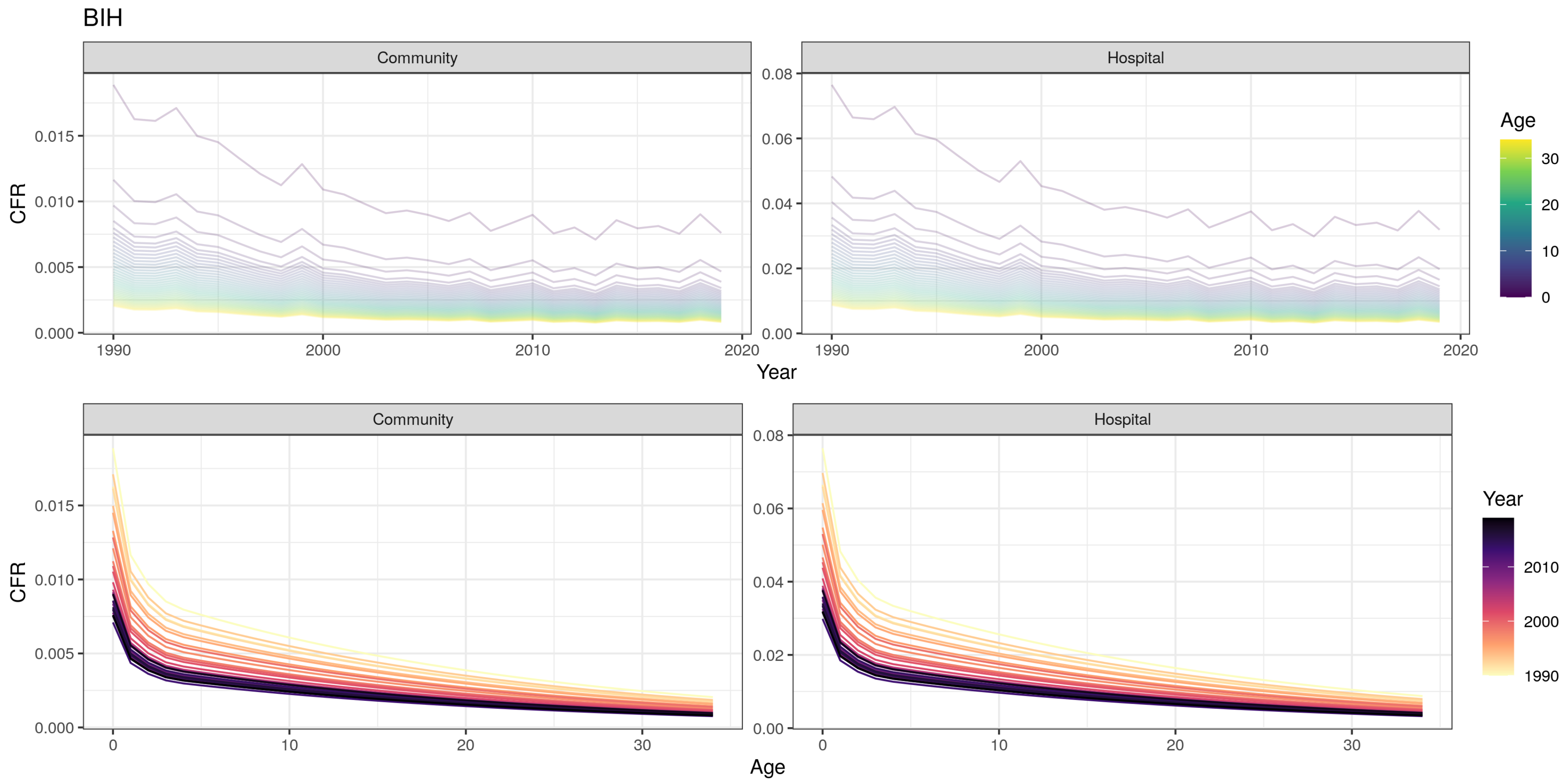
BIH
Community
Hospital
CFR
Year
Age
Age
Year
1990
2000
2010
2020
0
10
20
30

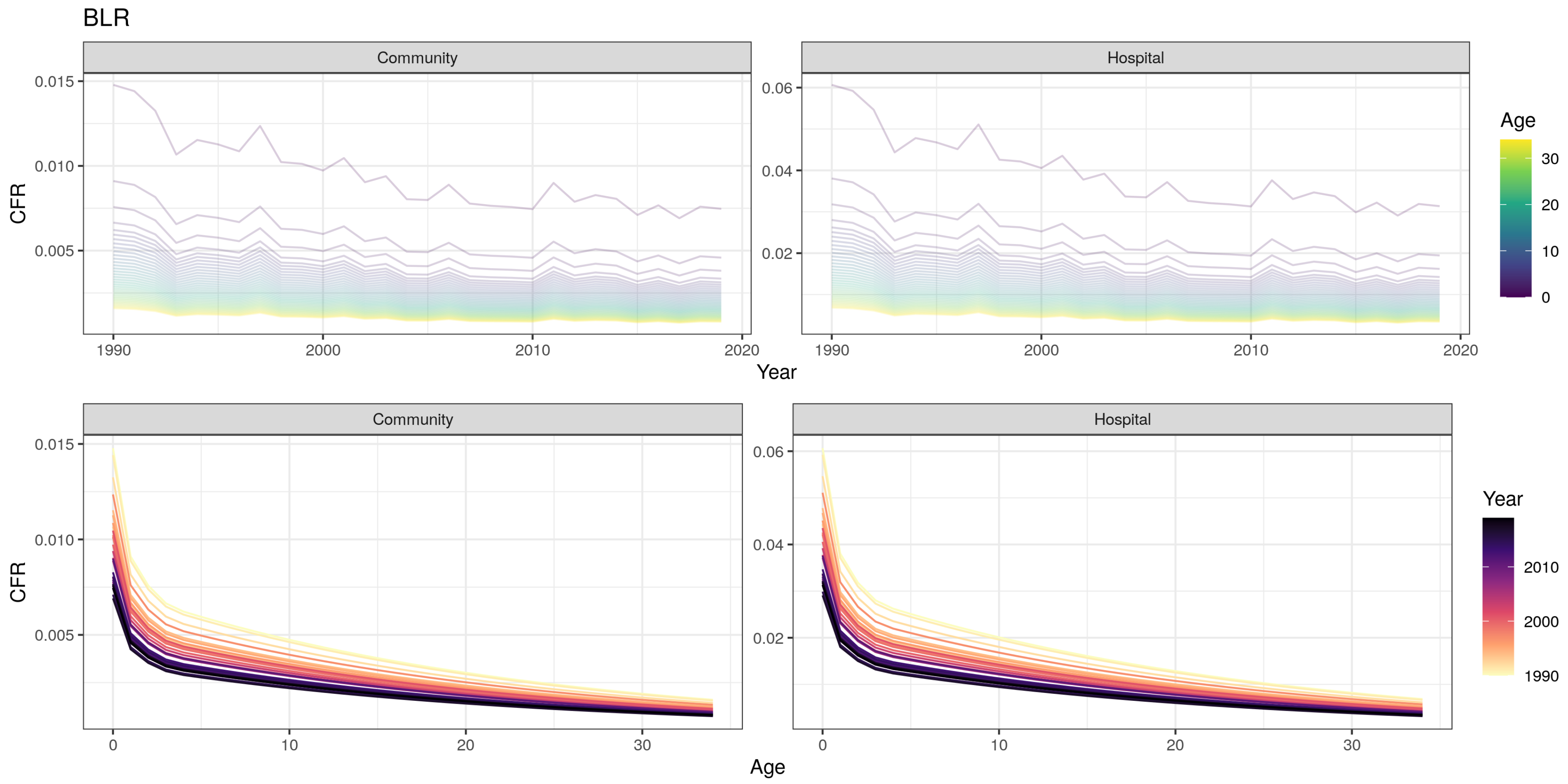
BLR
Community
Hospital
CFR
Year
Age
0.015
0.010
0.005
0.06
0.04
0.02
1990
2000
2010
2020
30
20
10
0
Community
Hospital
CFR
Age
Year
2010
2000
1990

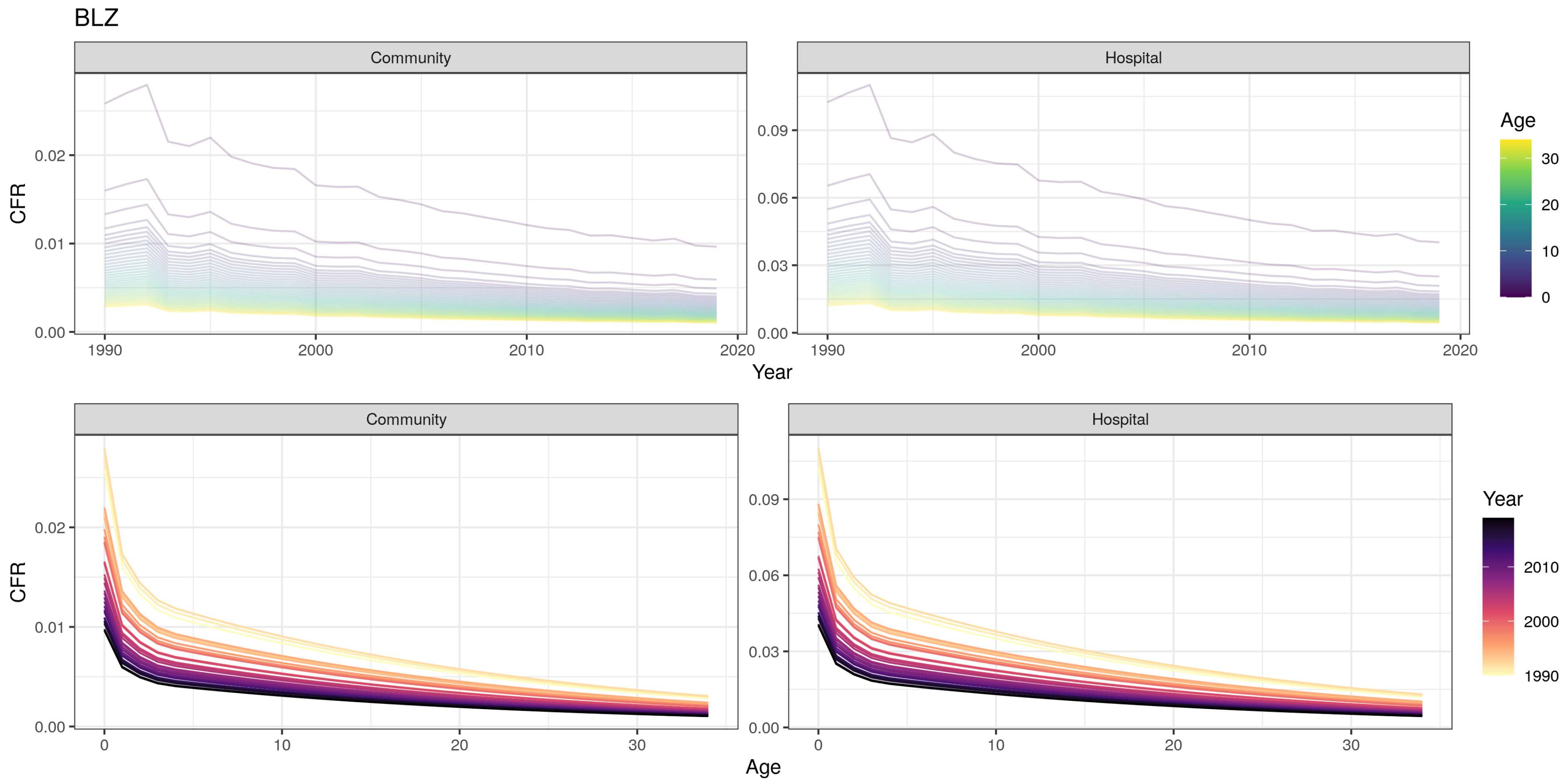

BLZ
Community
Hospital
CFR
0.00
0.01
0.02
0.03
0.06
0.09
1990
2000
2010
2020
Year
Age
0
10
20
30
Age
Year
1990
2000
2010

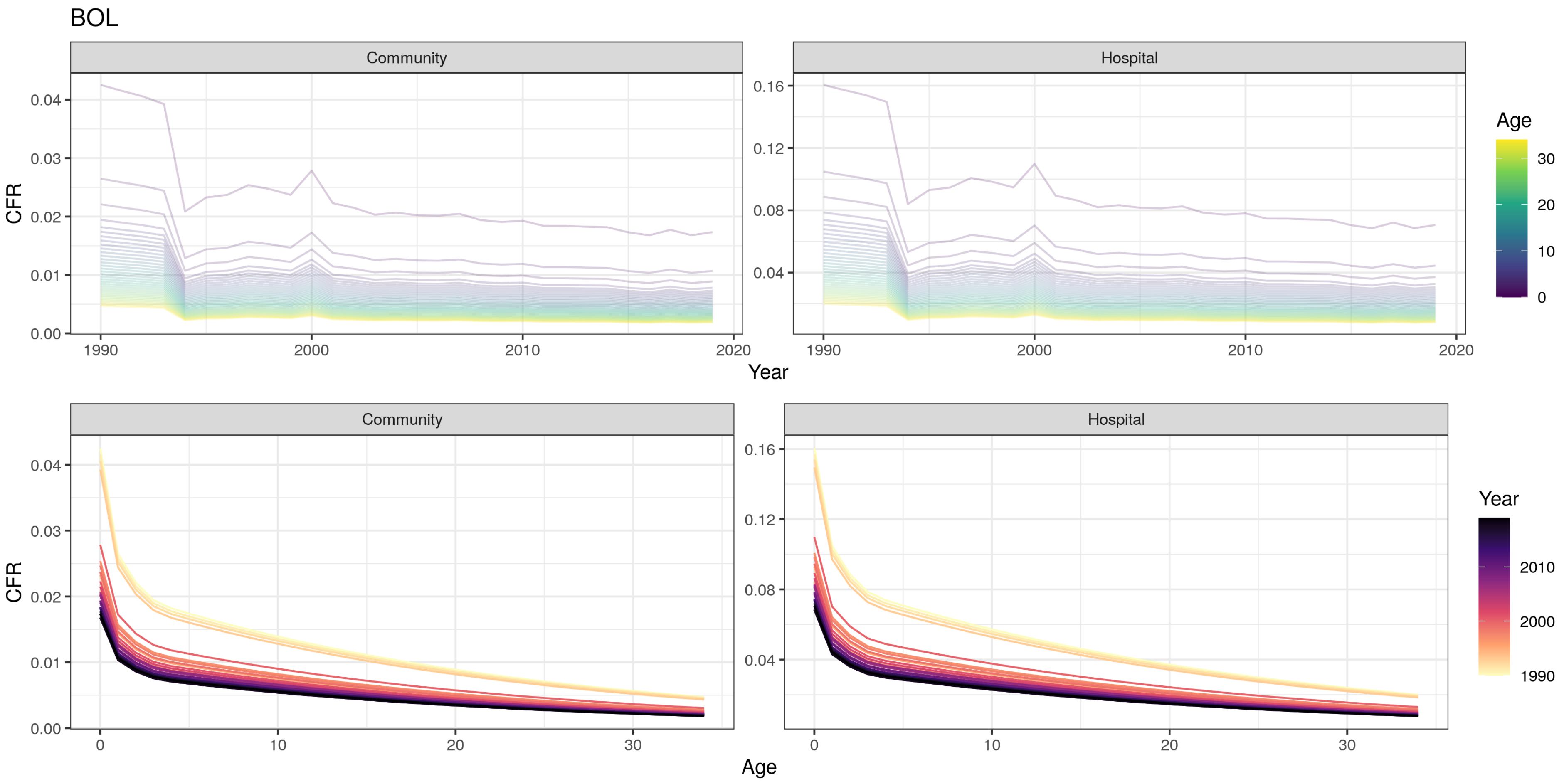

BOL
Community
Hospital
CFR
Year
Age
Community
Hospital
CFR
Age
Year
0.00
0.01
0.02
0.03
0.04
0.04
0.08
0.12
0.16
1990
2000
2010
2020
0
10
20
30

BTN

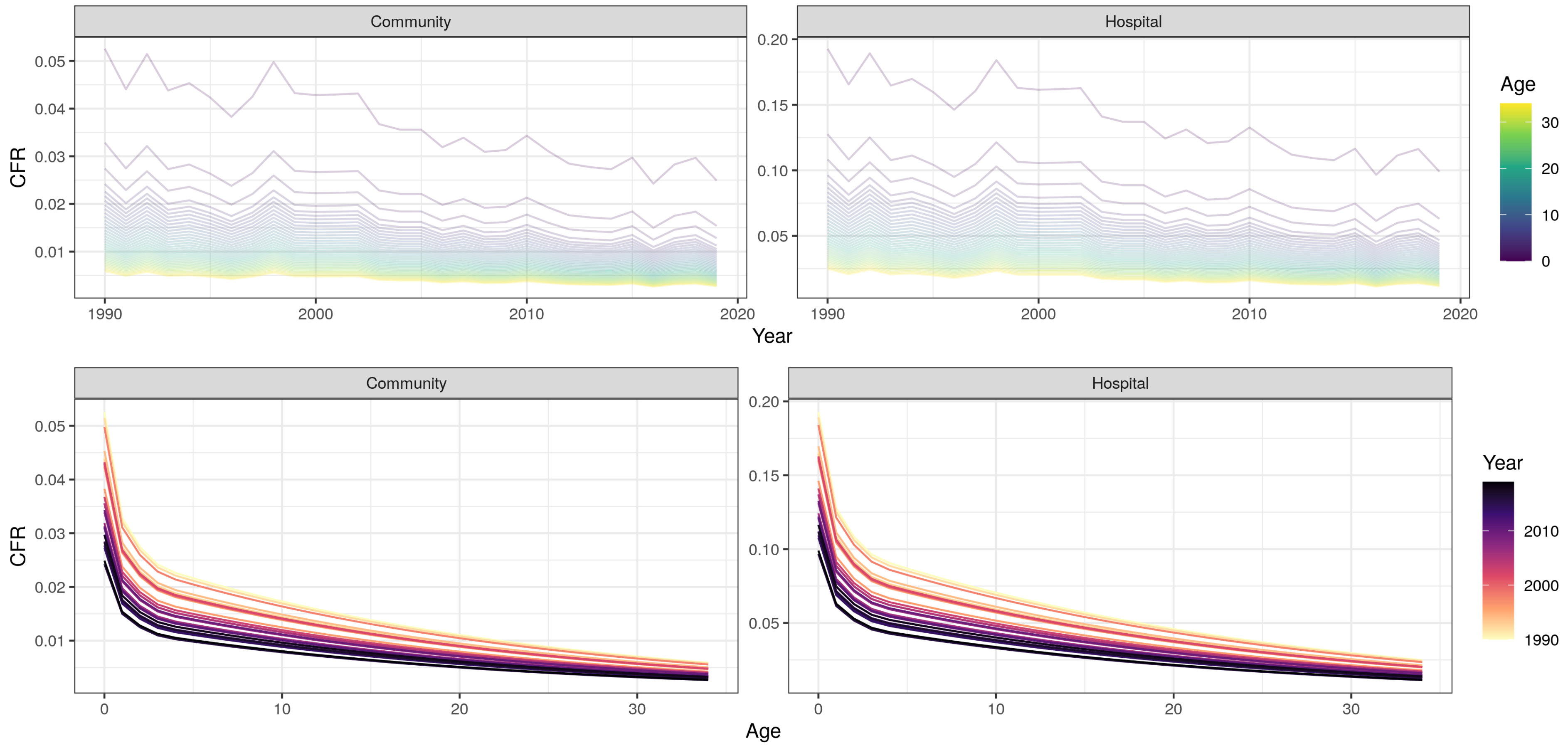
Community
Hospital
CFR
Year
Age
Community
Hospital
CFR
Age
Year
0.05
0.04
0.03
0.02
0.01
0.20
0.15
0.10
0.05
1990
2000
2010
2020
0
10
20
30

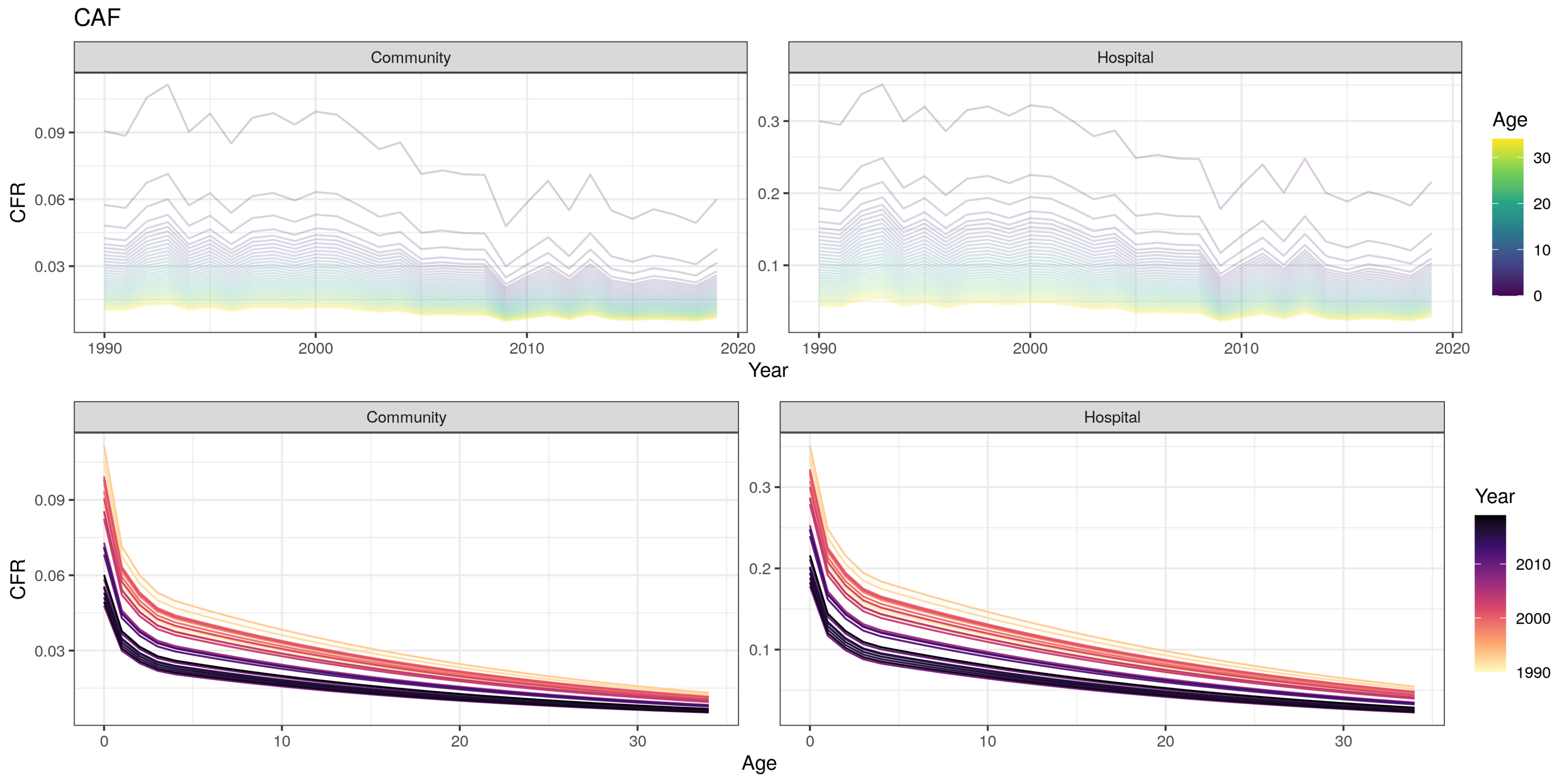
CAF
Community
Hospital
CFR
0.09
0.06
0.03
0.3
0.2
0.1
1990
2000
2010
2020
Year
Age
30
20
10
0
Community
Hospital
0
10
20
30
Age
Year
2010
2000
1990

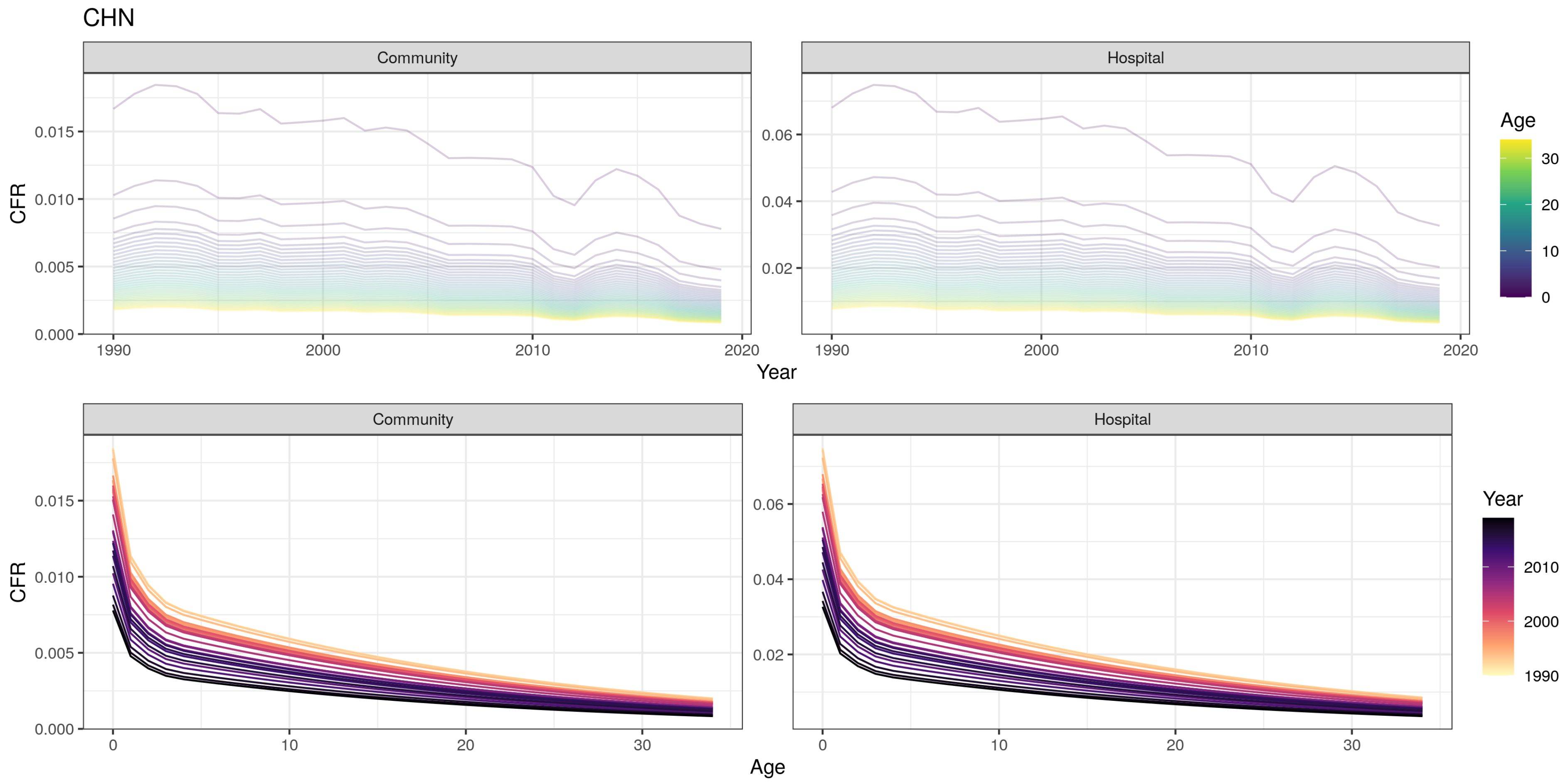

CHN
Community
Hospital
CFR
Year
Age
Community
Hospital
CFR
Age
Year

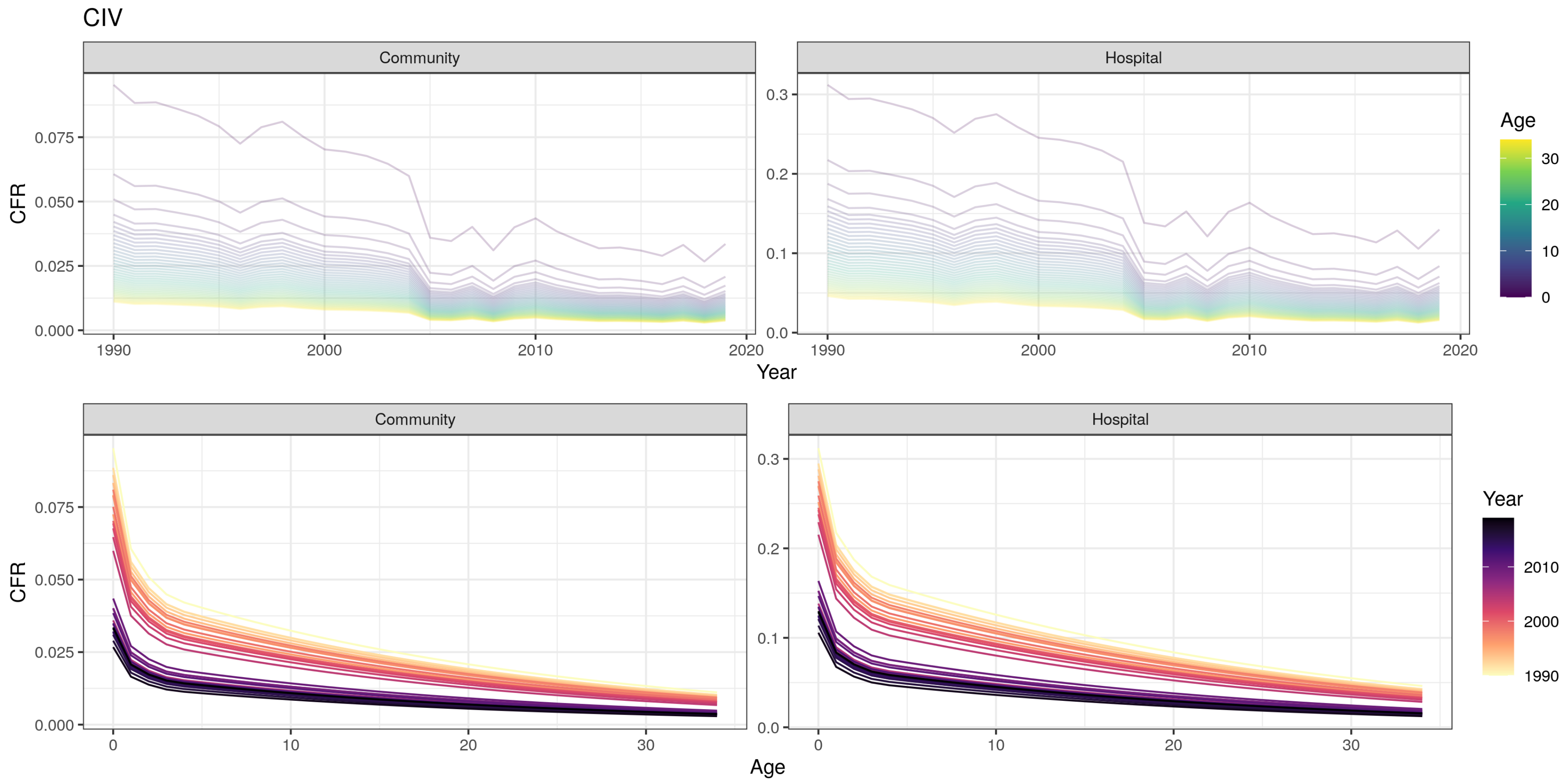

CIV
Community
Hospital
CFR
Year
Age
0.000
0.025
0.050
0.075
0.0
0.1
0.2
0.3
1990
2000
2010
2020
0
10
20
30
Year
1990
2000
2010

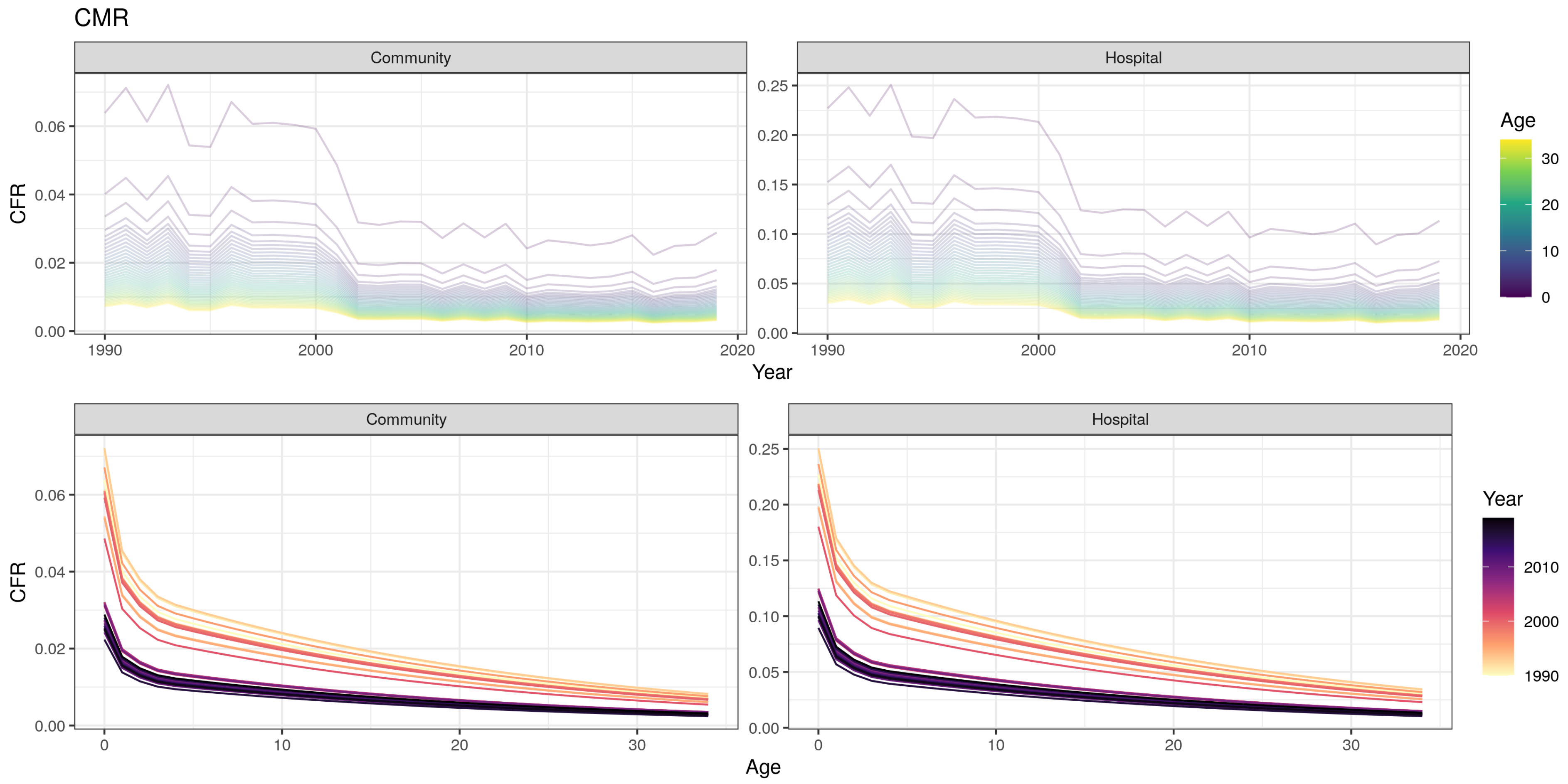
CMR
Community
Hospital
CFR
Year
Age
Age
0
10
20
30
Year
1990
2000
2010
0.00
0.02
0.04
0.06
0.05
0.10
0.15
0.20
0.25
1990
2000
2010
2020
0
10
20
30

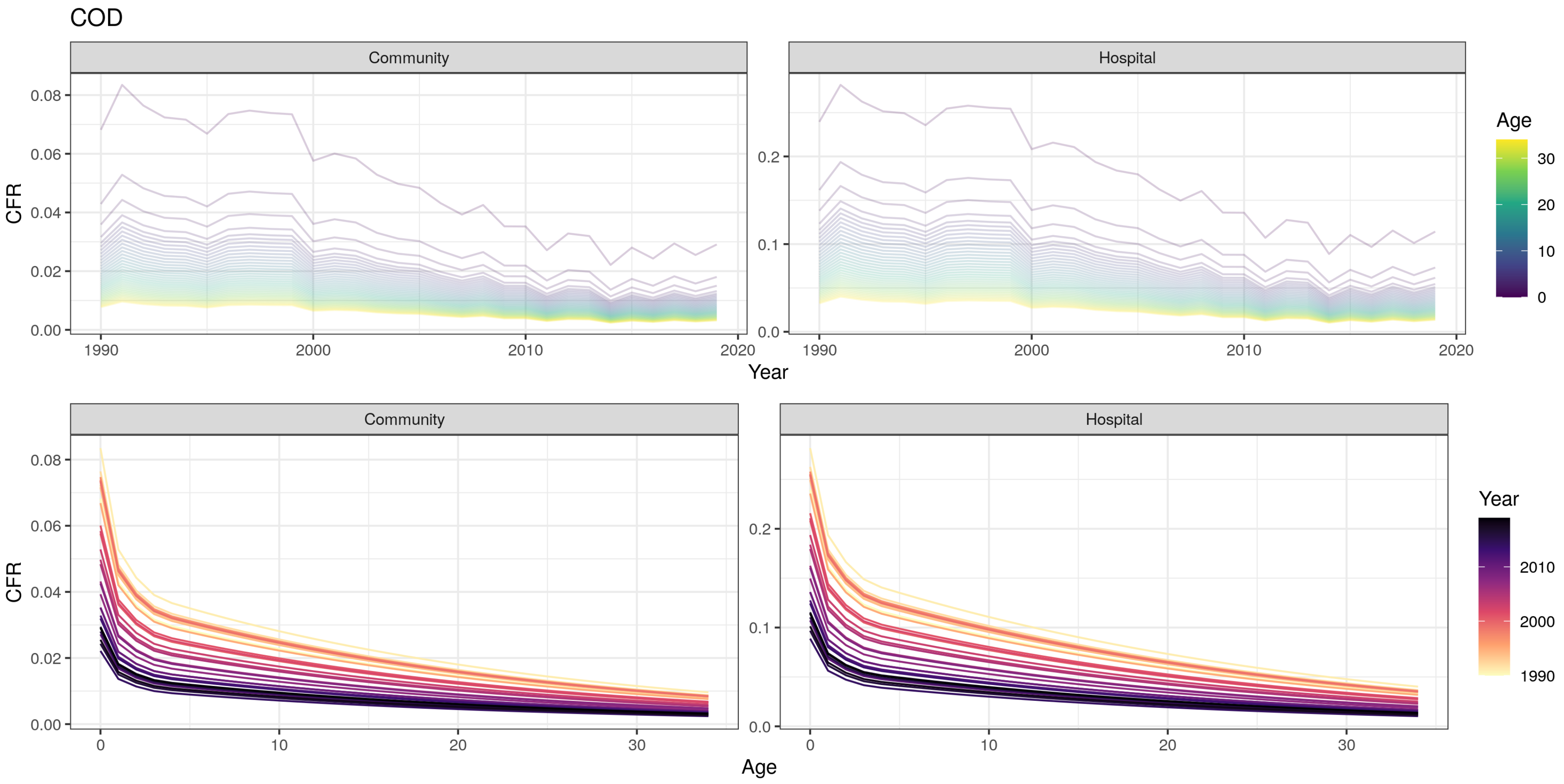
COD
Community
Hospital
CFR
Year
Age
Community
Hospital
CFR
Age
Year
0.08
0.06
0.04
0.02
0.00
0.2
0.1
0.0
1990
2000
2010
2020
0
10
20
30
1990
2000
2010

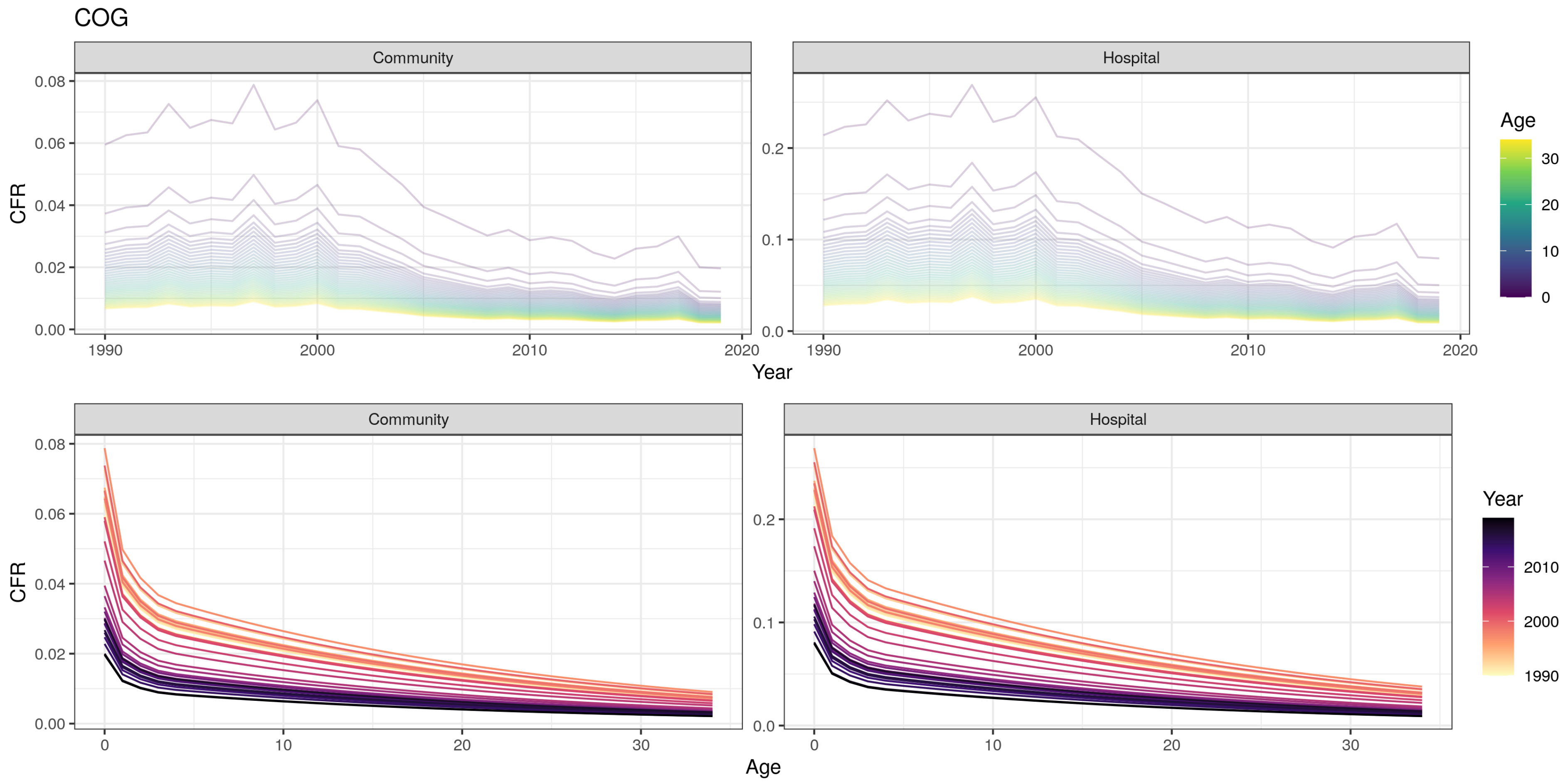

COG
Community
Hospital
CFR
Year
Age
0.00
0.02
0.04
0.06
0.08
0.0
0.1
0.2
1990
2000
2010
2020
0
10
20
30

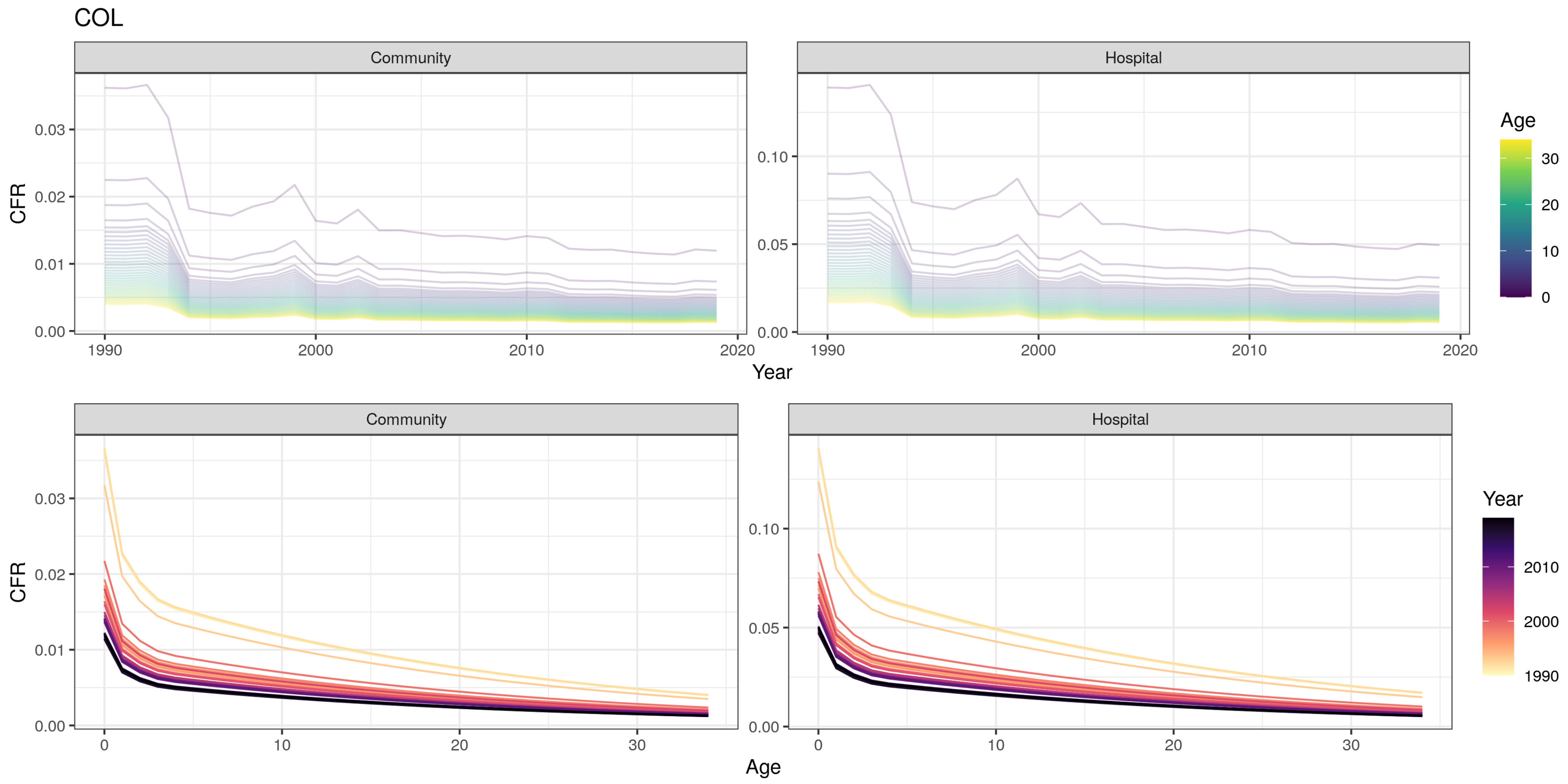

COL
Community
Hospital
CFR
Year
Age
0.00
0.01
0.02
0.03
0.05
0.10
1990
2000
2010
2020
0
10
20
30

COM

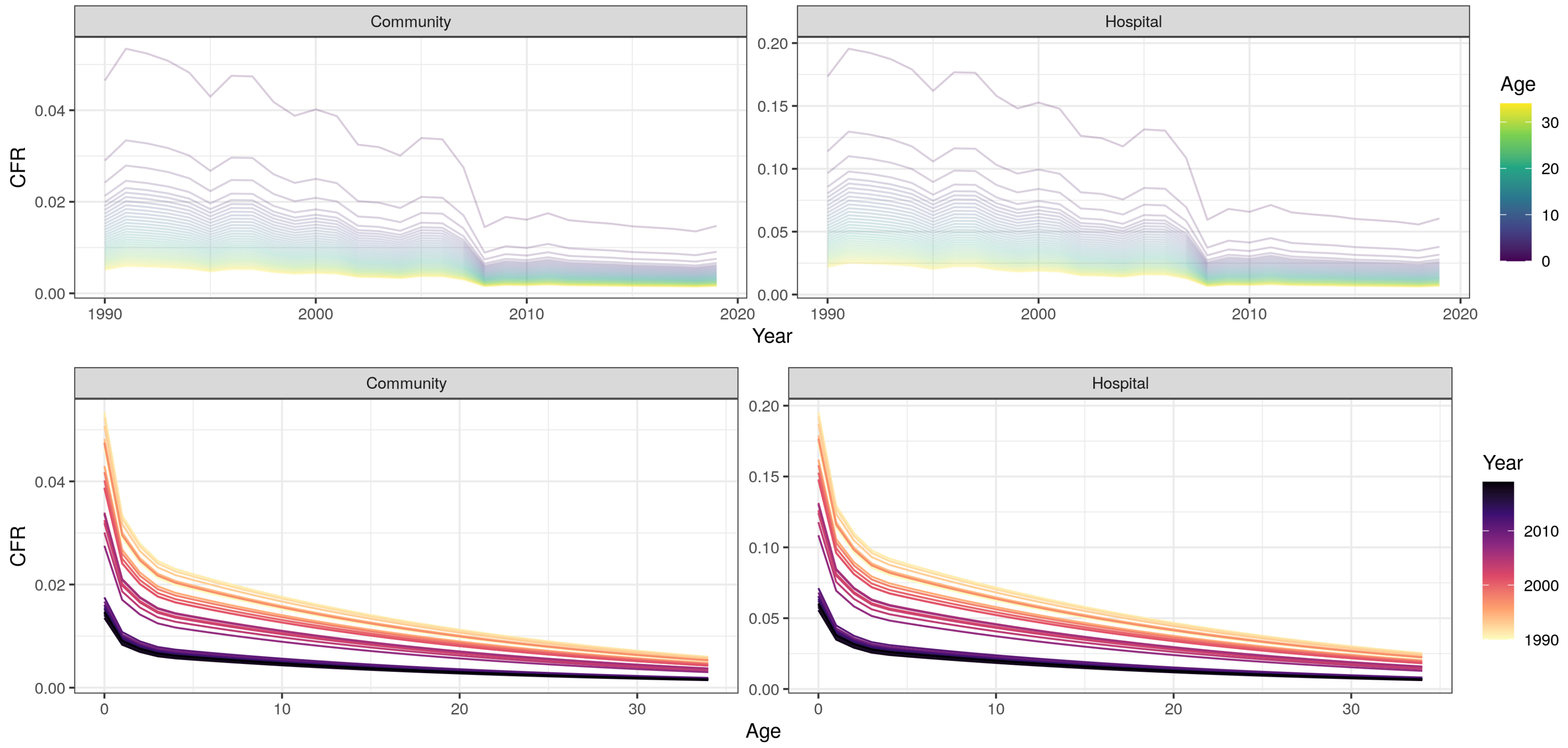
Community
Hospital
CFR
Year
Age
0.00
0.02
0.04
0.05
0.10
0.15
0.20
1990
2000
2010
2020
0
10
20
30

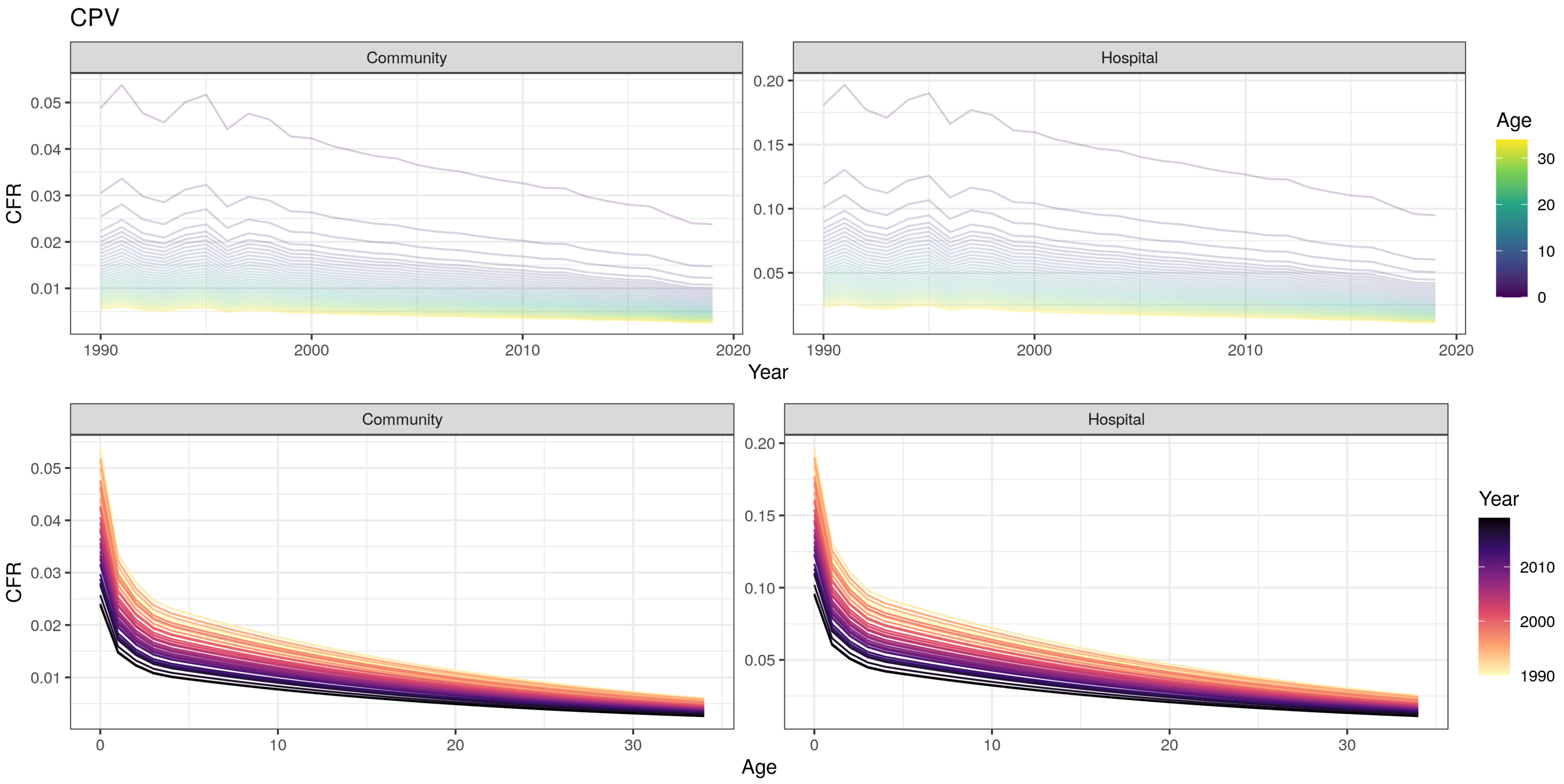
CPV
Community
Hospital
CFR
0.05
0.04
0.03
0.02
0.01
0.20
0.15
0.10
0.05
1990
2000
2010
2020
Year
Age
30
20
10
0
Community
Hospital
CFR
0
10
20
30
Age
Year
2010
2000
1990

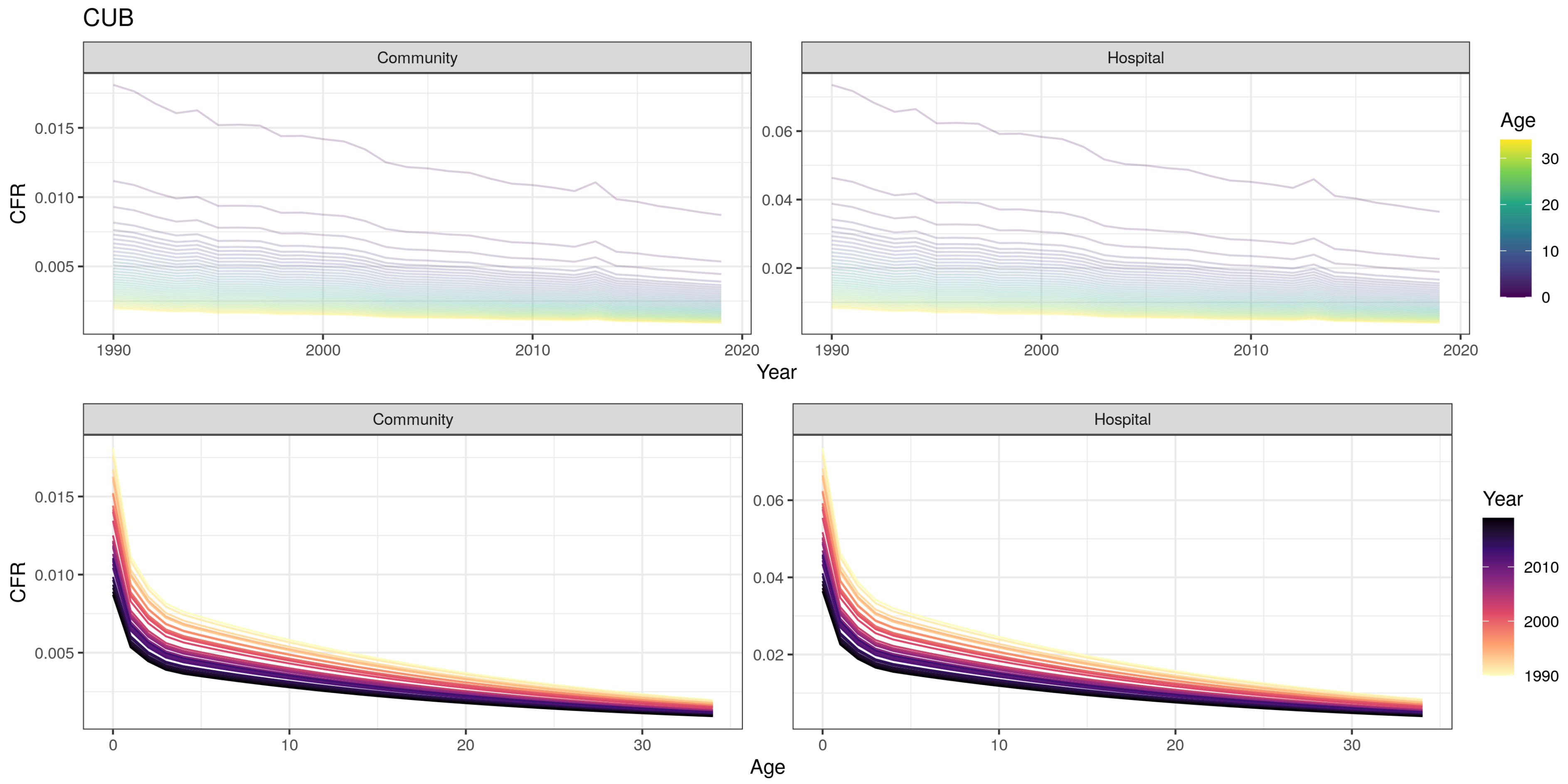

CUB
Community
Hospital
CFR
0.015
0.010
0.005
0.06
0.04
0.02
1990
2000
2010
2020
Year
Age
30
20
10
0
Community
Hospital
CFR
0.015
0.010
0.005
0.06
0.04
0.02
0
10
20
30
Age
Year
2010
2000
1990

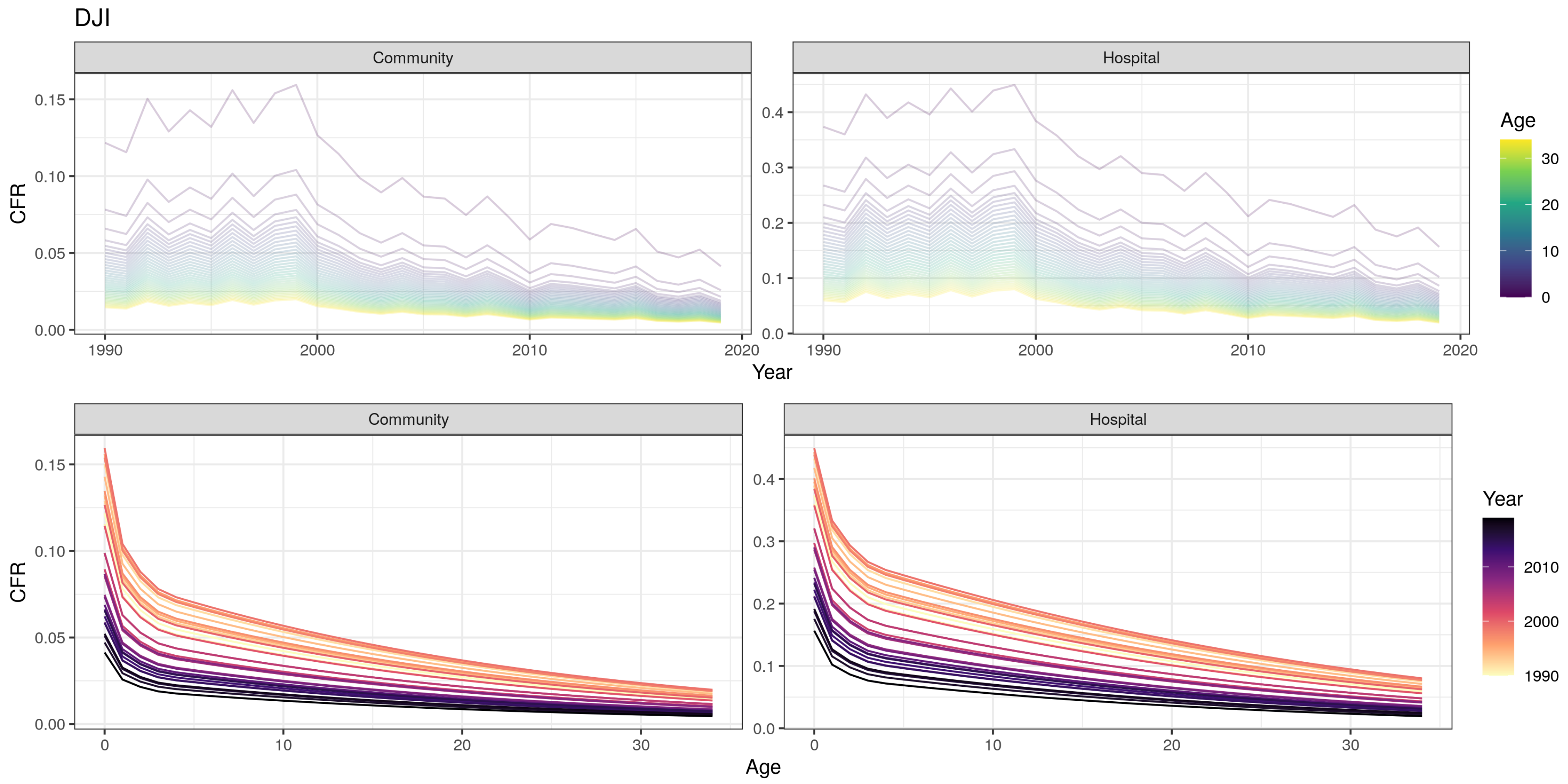

DJI
Community
Hospital
CFR
Year
Age
Community
Hospital
CFR
Age
Year
0.15
0.10
0.05
0.00
0.4
0.3
0.2
0.1
0.0
1990
2000
2010
2020
0
10
20
30
1990
2000
2010

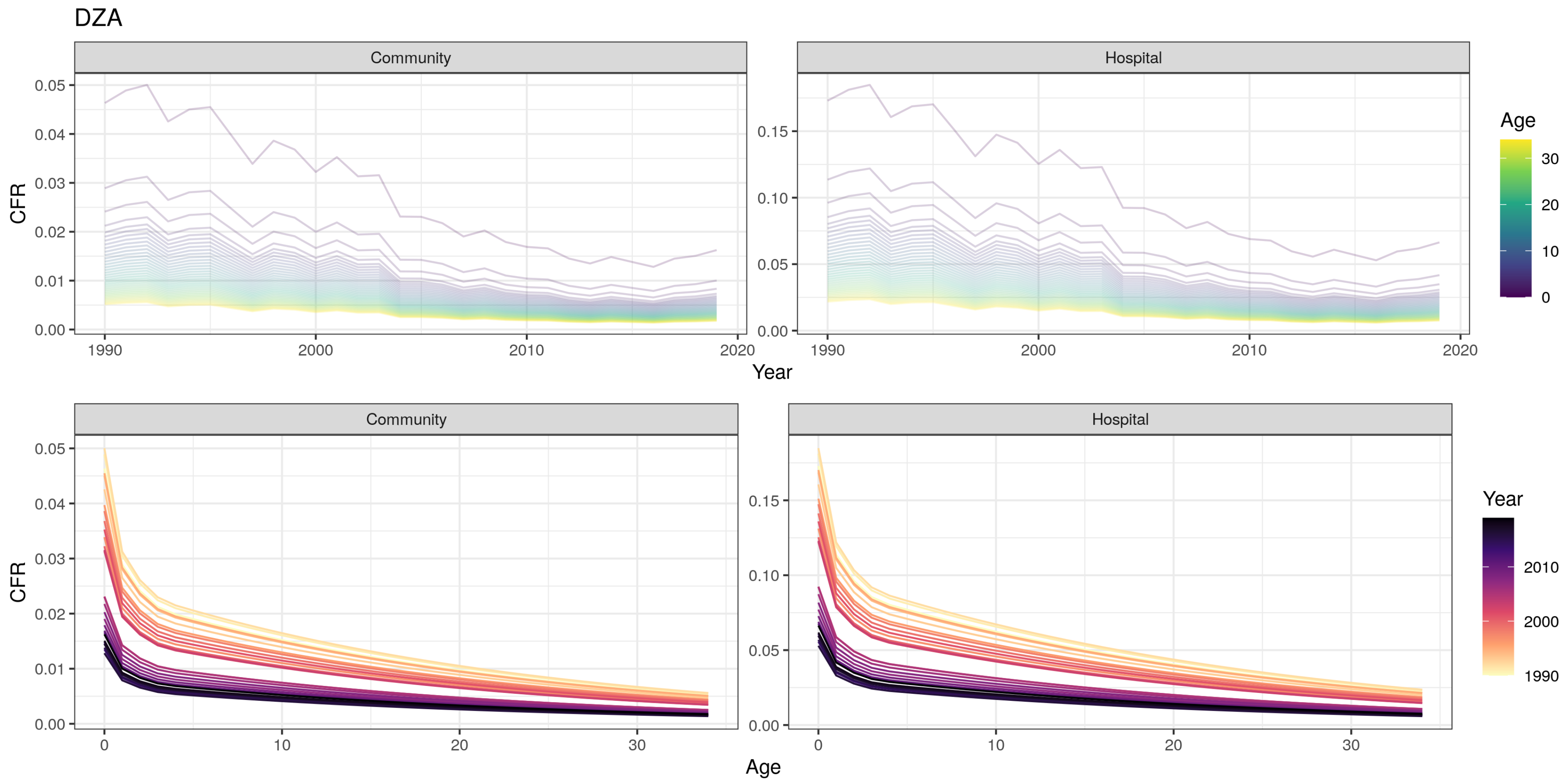
DZA
Community
Hospital
CFR
Year
Age
Age
Year
0.05
0.04
0.03
0.02
0.01
0.00
0.15
0.10
0.05
0.00
1990
2000
2010
2020
0
10
20
30

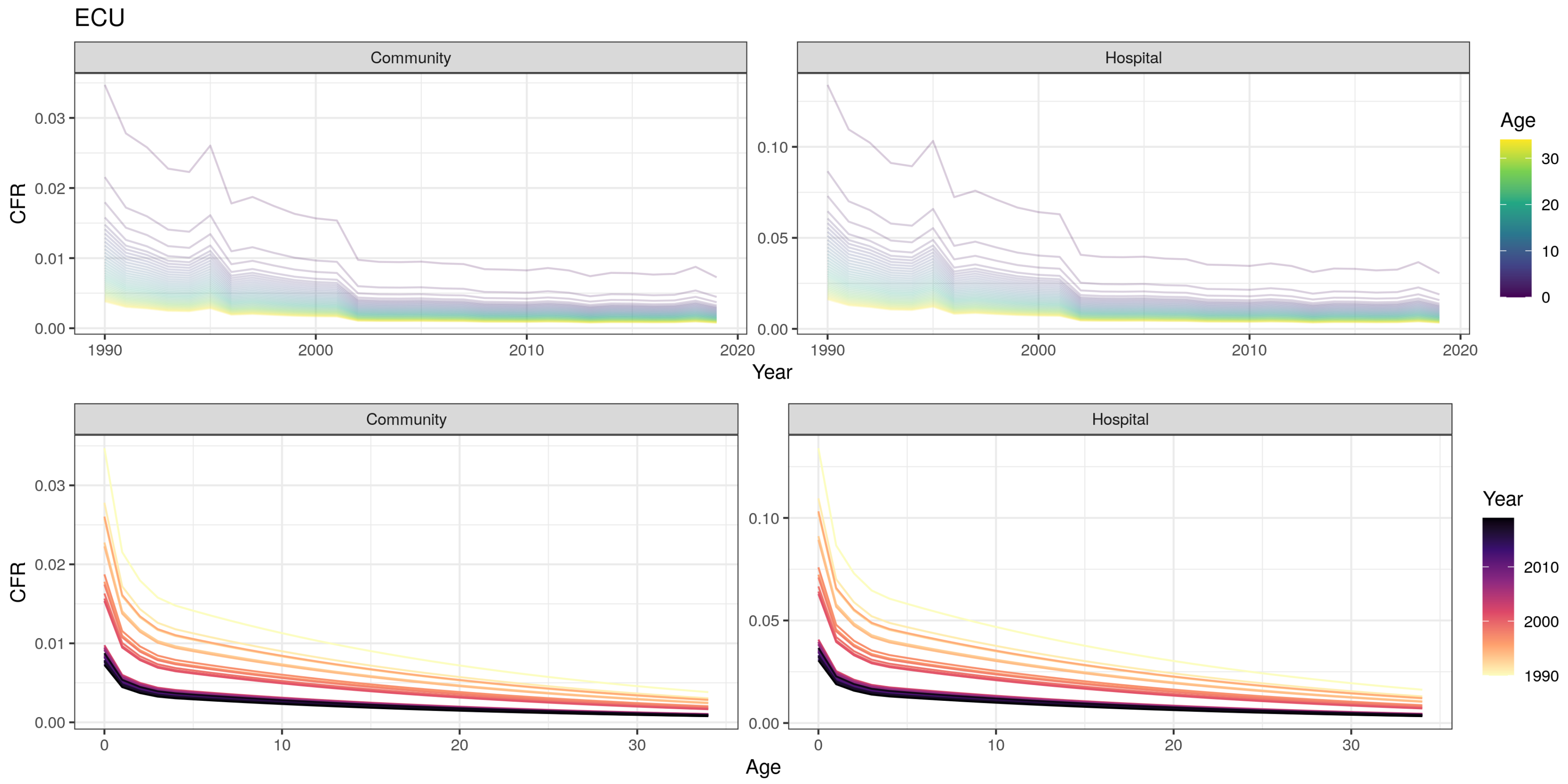
ECU
Community
Hospital
CFR
Year
Age
0.00
0.01
0.02
0.03
0.05
0.10
1990
2000
2010
2020
0
10
20
30

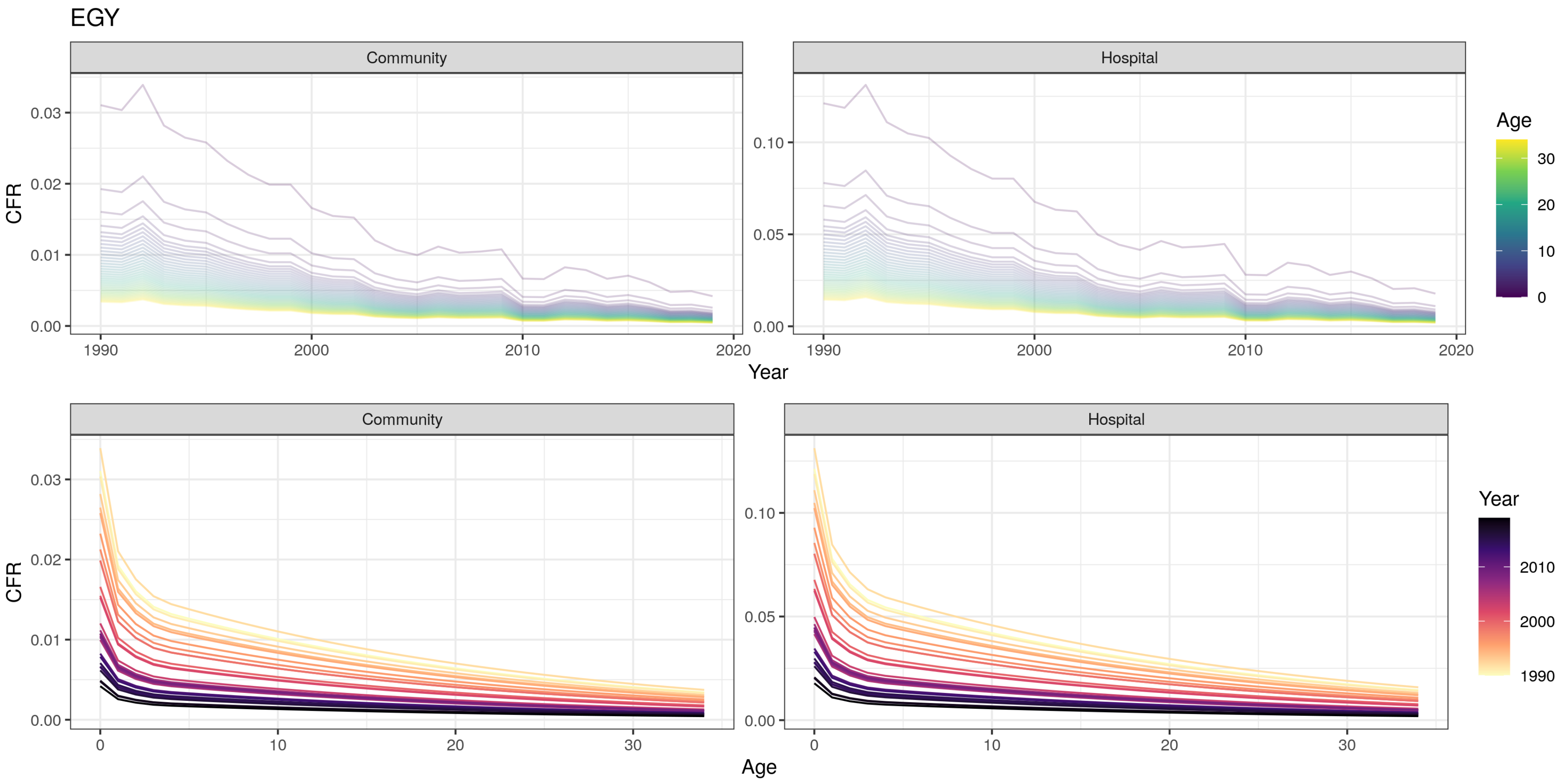

EGY
Community
Hospital
CFR
Year
Age
0.00
0.01
0.02
0.03
0.05
0.10
1990
2000
2010
2020
0
10
20
30

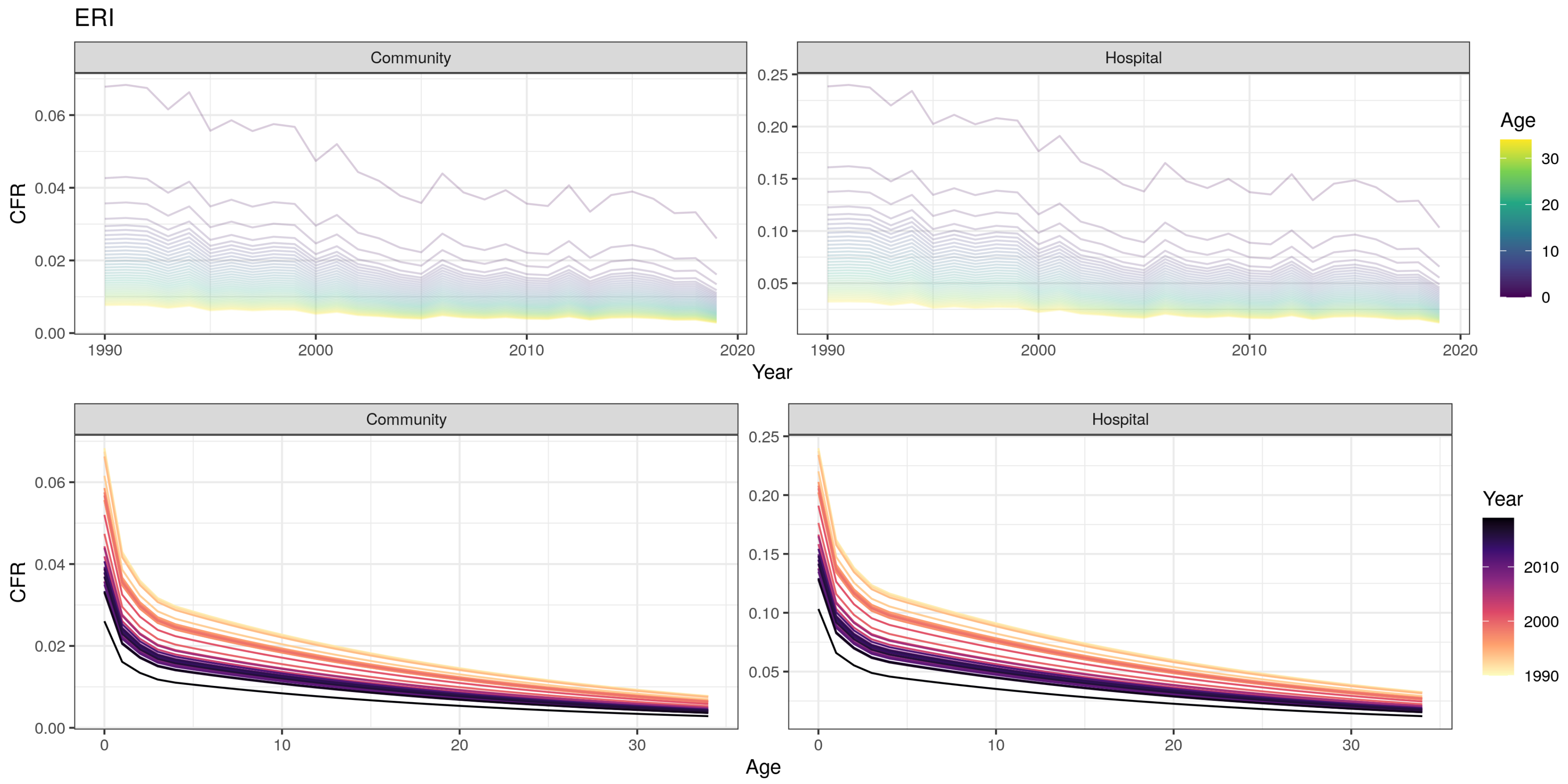
ERI
Community
Hospital
CFR
Year
Age
0.00
0.02
0.04
0.06
0.05
0.10
0.15
0.20
0.25
1990
2000
2010
2020
0
10
20
30
Age
Year
1990
2000
2010

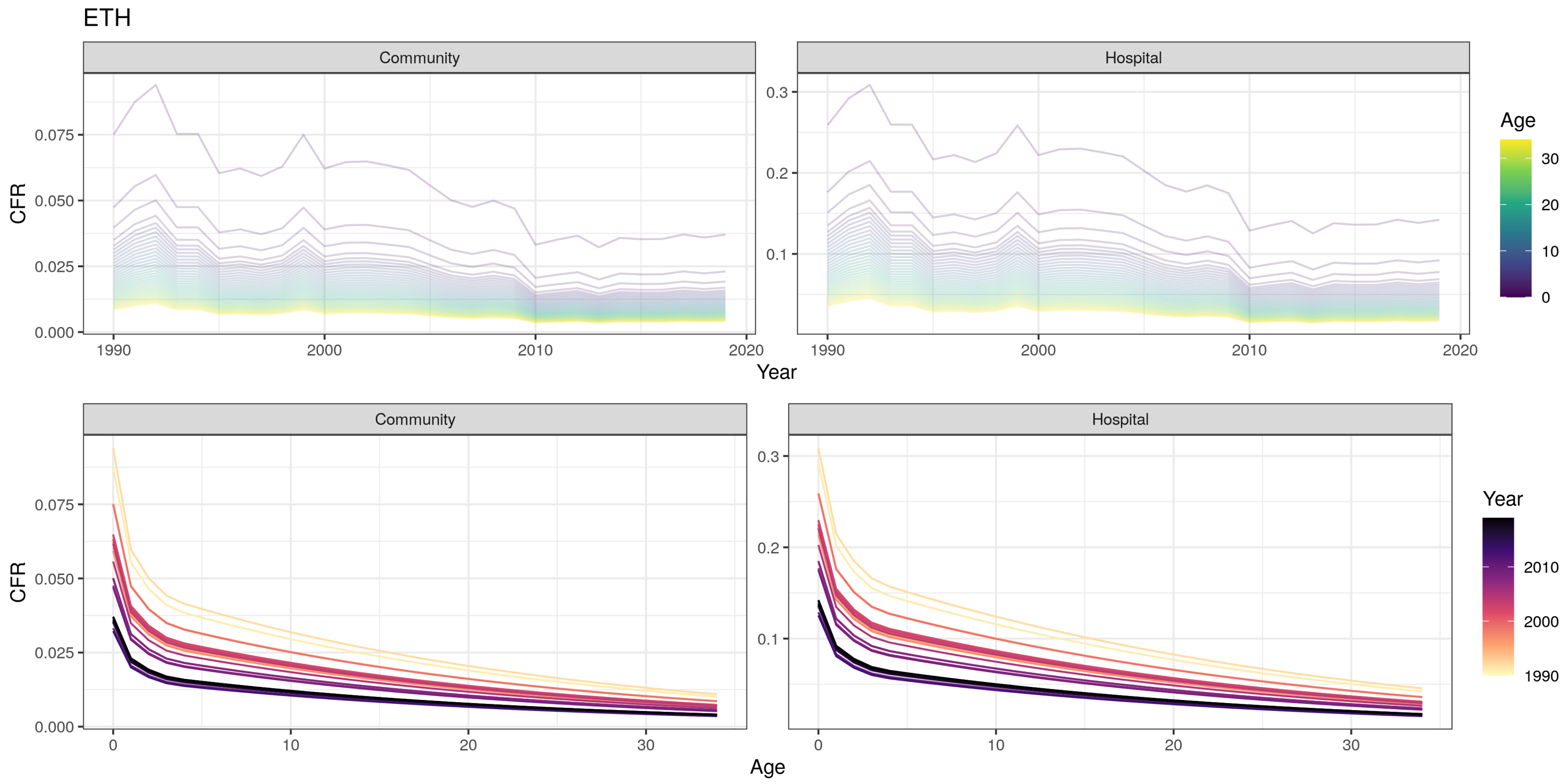
ETH
Community
Hospital
CFR
Year
Age
Community
Hospital
CFR
Age
Year

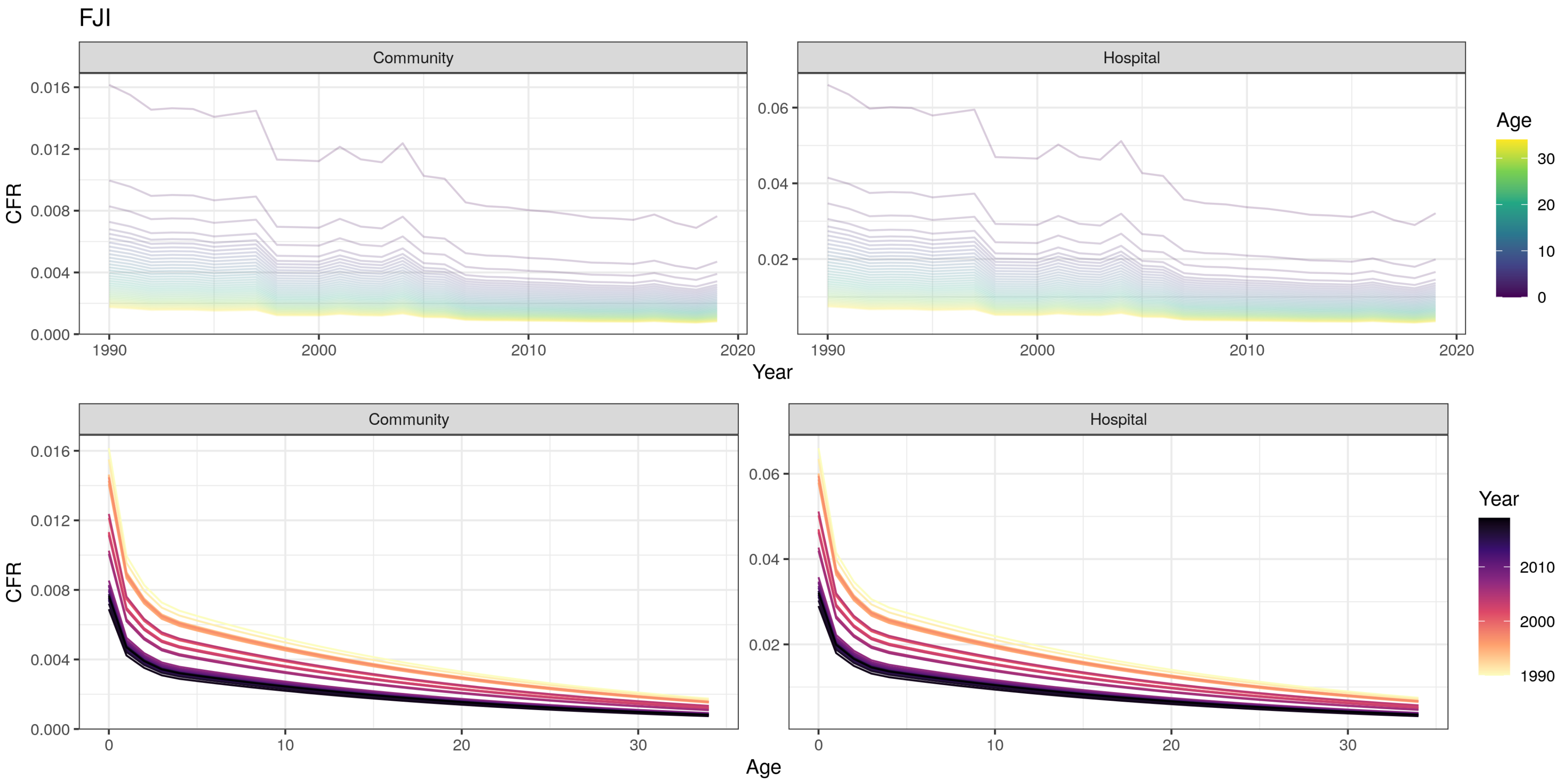
FJI
Community
Hospital
CFR
Year
Age
Community
Hospital
CFR
Age
Year

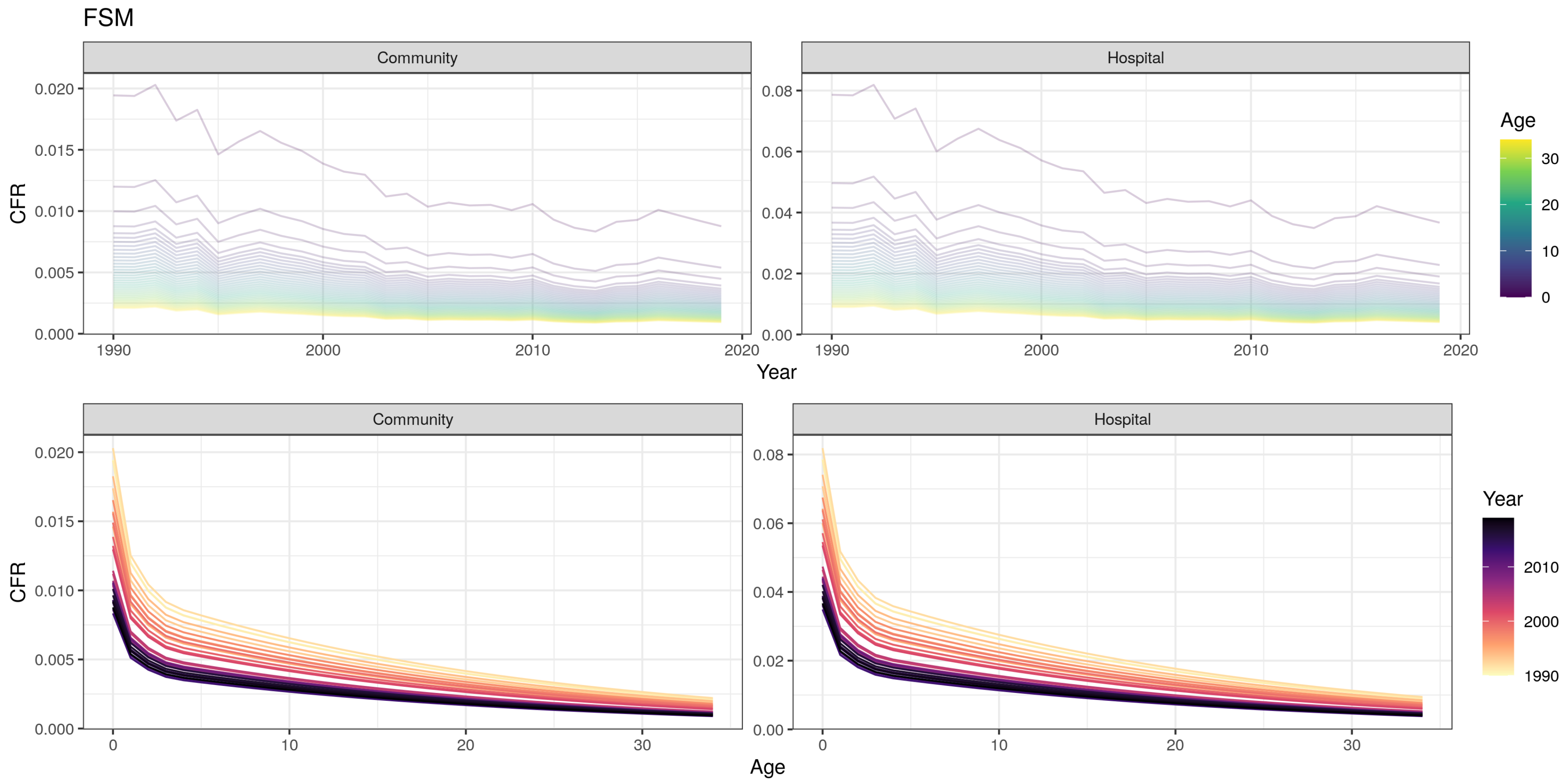

FSM
Community
Hospital
CFR
Year
Age
Community
Hospital
CFR
Age
Year
0.000
0.005
0.010
0.015
0.020
0.00
0.02
0.04
0.06
0.08
1990
2000
2010
2020
0
10
20
30

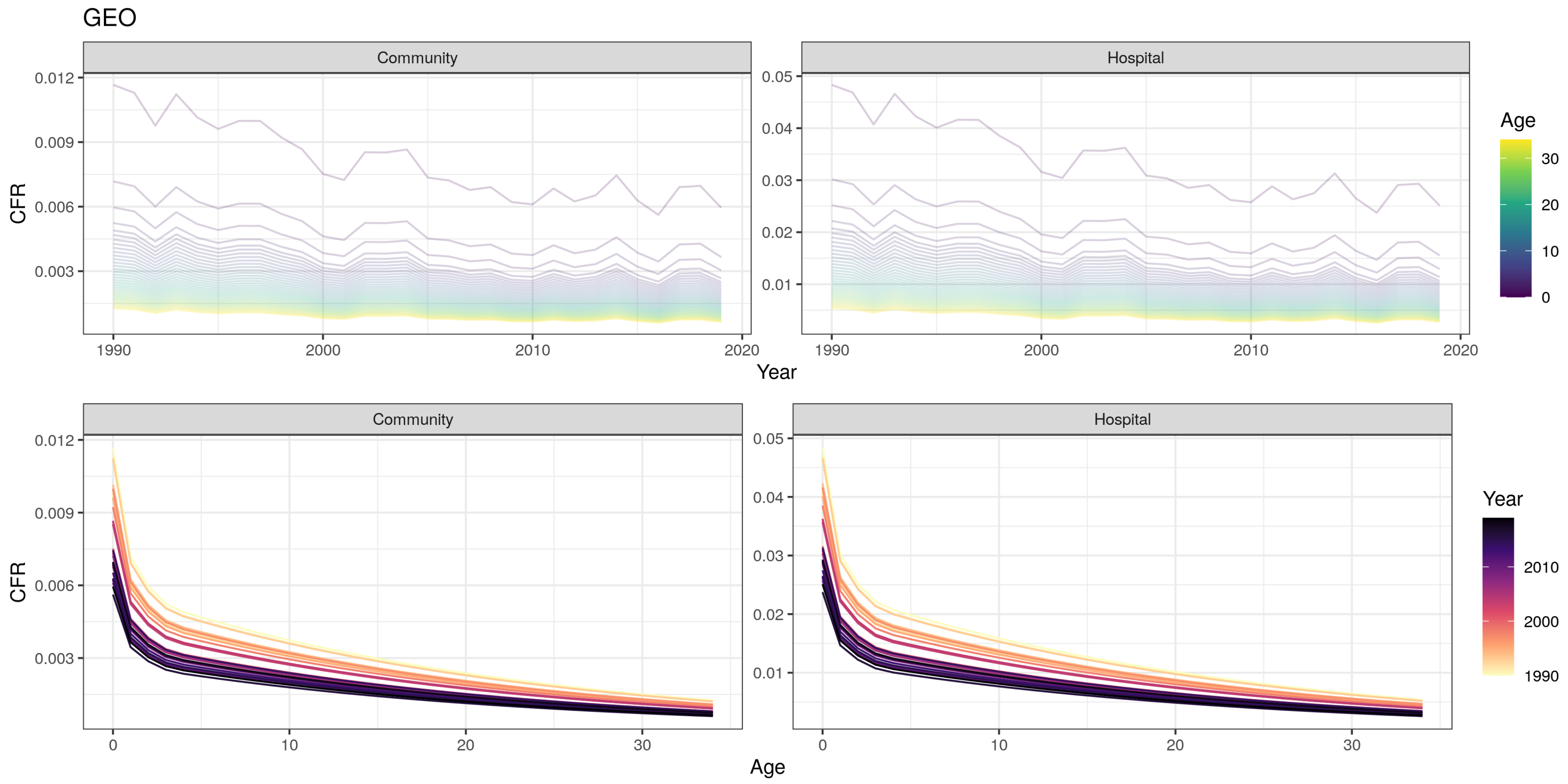
GEO
Community
Hospital
CFR
0.012
0.009
0.006
0.003
0.05
0.04
0.03
0.02
0.01
1990
2000
2010
2020
Year
Age
30
20
10
0
Community
Hospital
CFR
0
10
20
30
Age
Year
2010
2000
1990

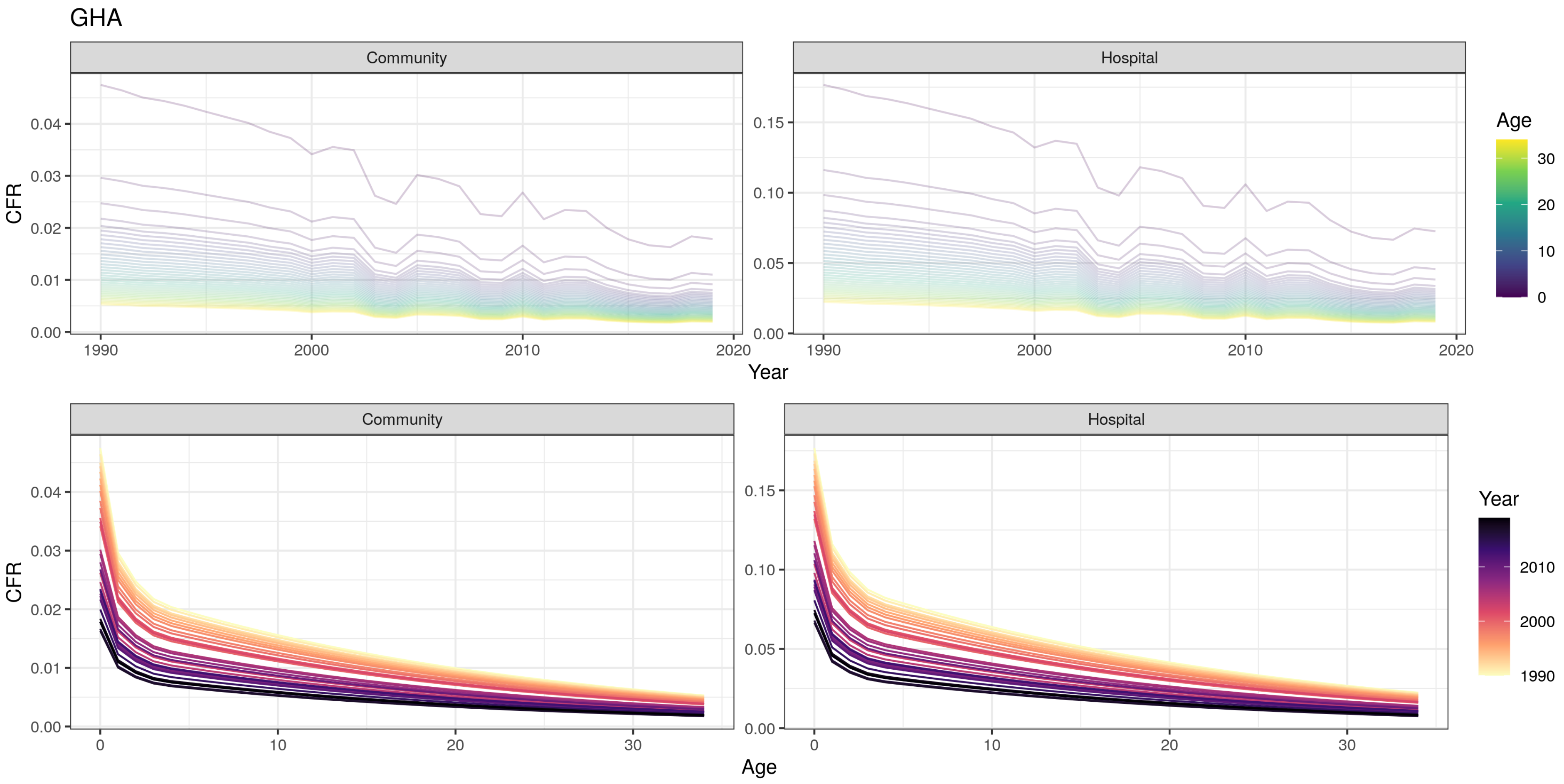
GHA
Community
Hospital
CFR
Year
Age
0.00
0.01
0.02
0.03
0.04
0.05
0.10
0.15
1990
2000
2010
2020
0
10
20
30
Age
Year

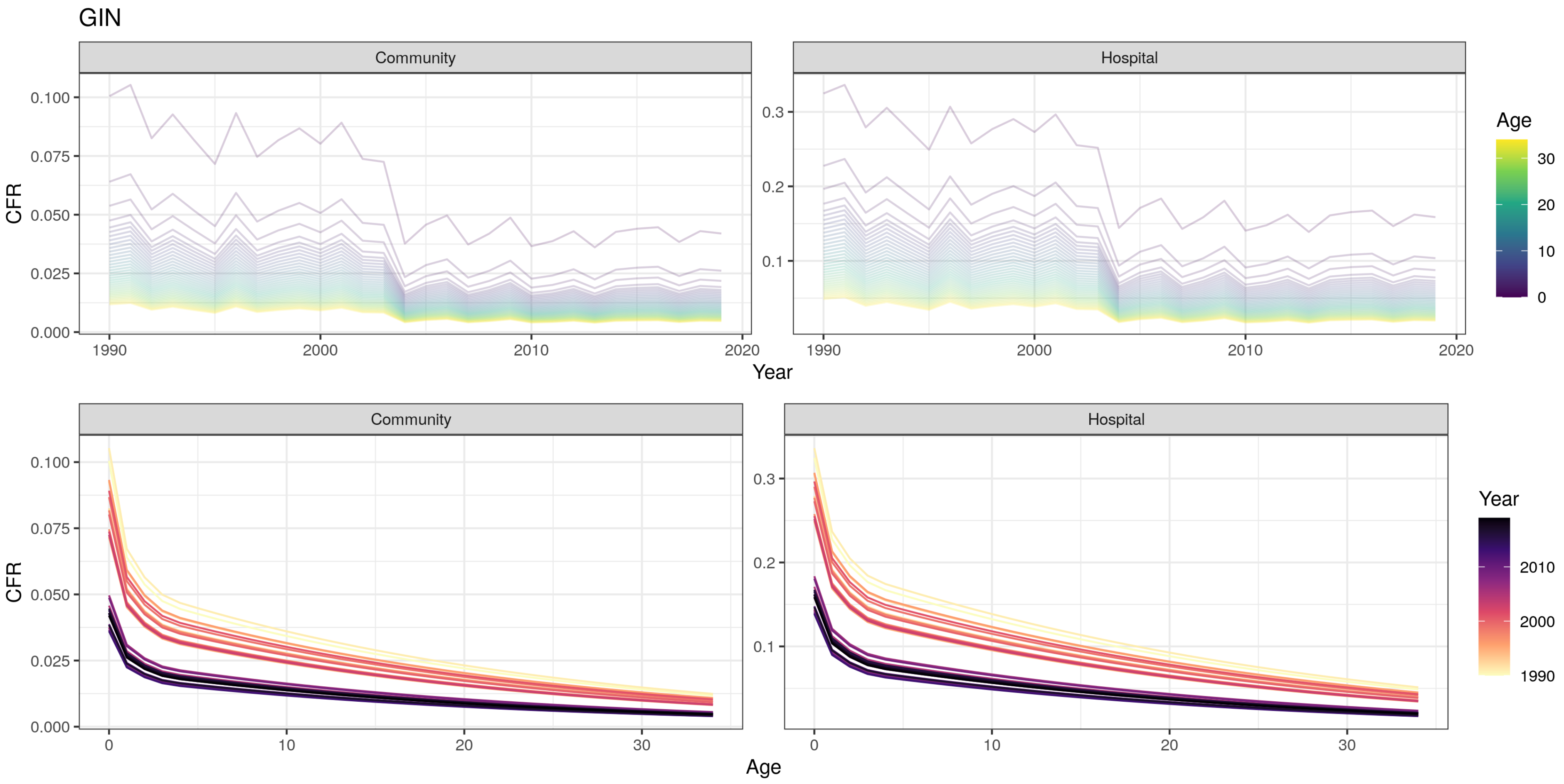

GIN
Community
Hospital
CFR
Year
Age
0.000
0.025
0.050
0.075
0.100
0.1
0.2
0.3
1990
2000
2010
2020
0
10
20
30
Age
Year

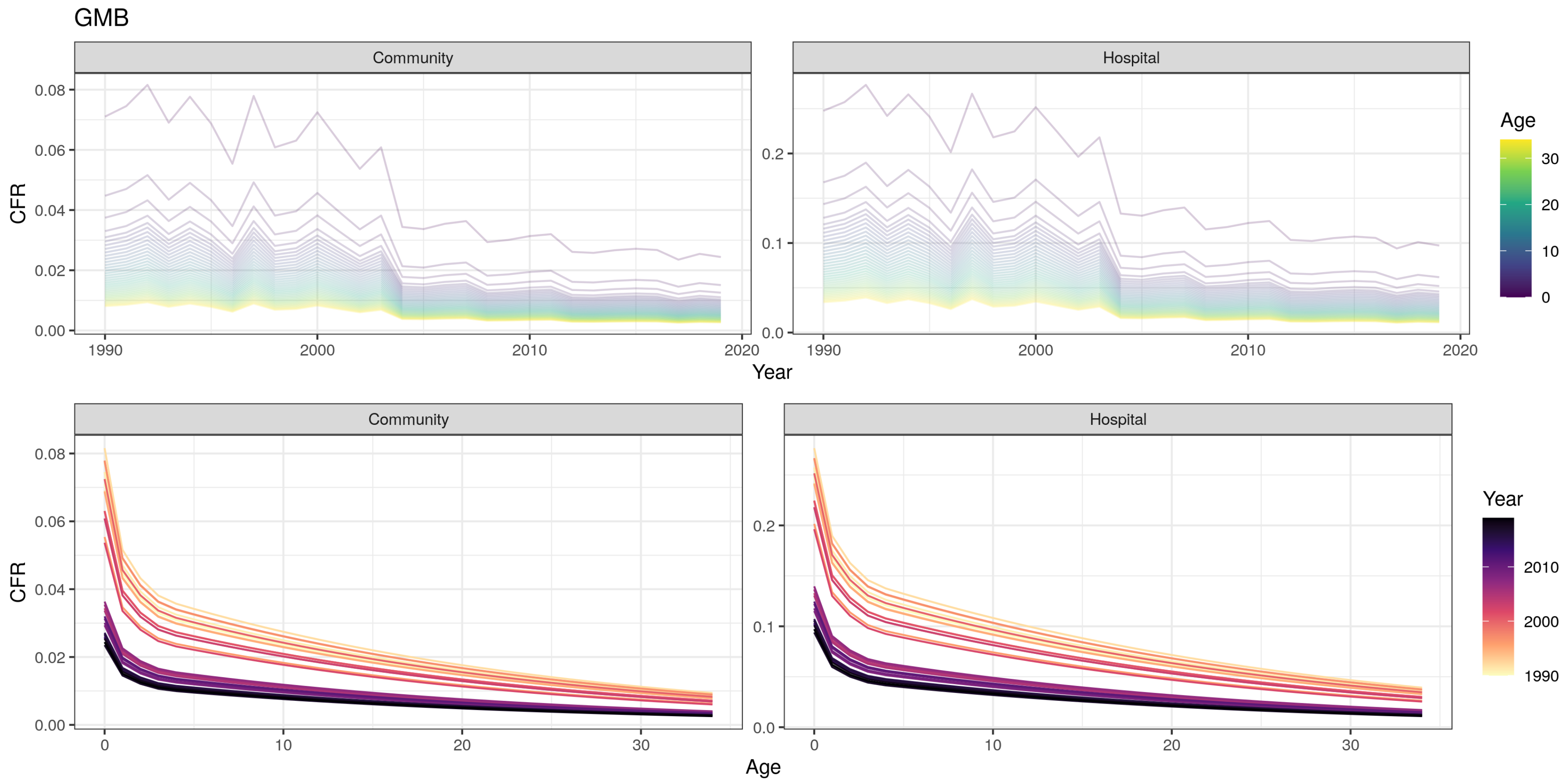

GMB
Community
Hospital
CFR
Year
Age
0.00
0.02
0.04
0.06
0.08
0.0
0.1
0.2
1990
2000
2010
2020
0
10
20
30
Age
Year

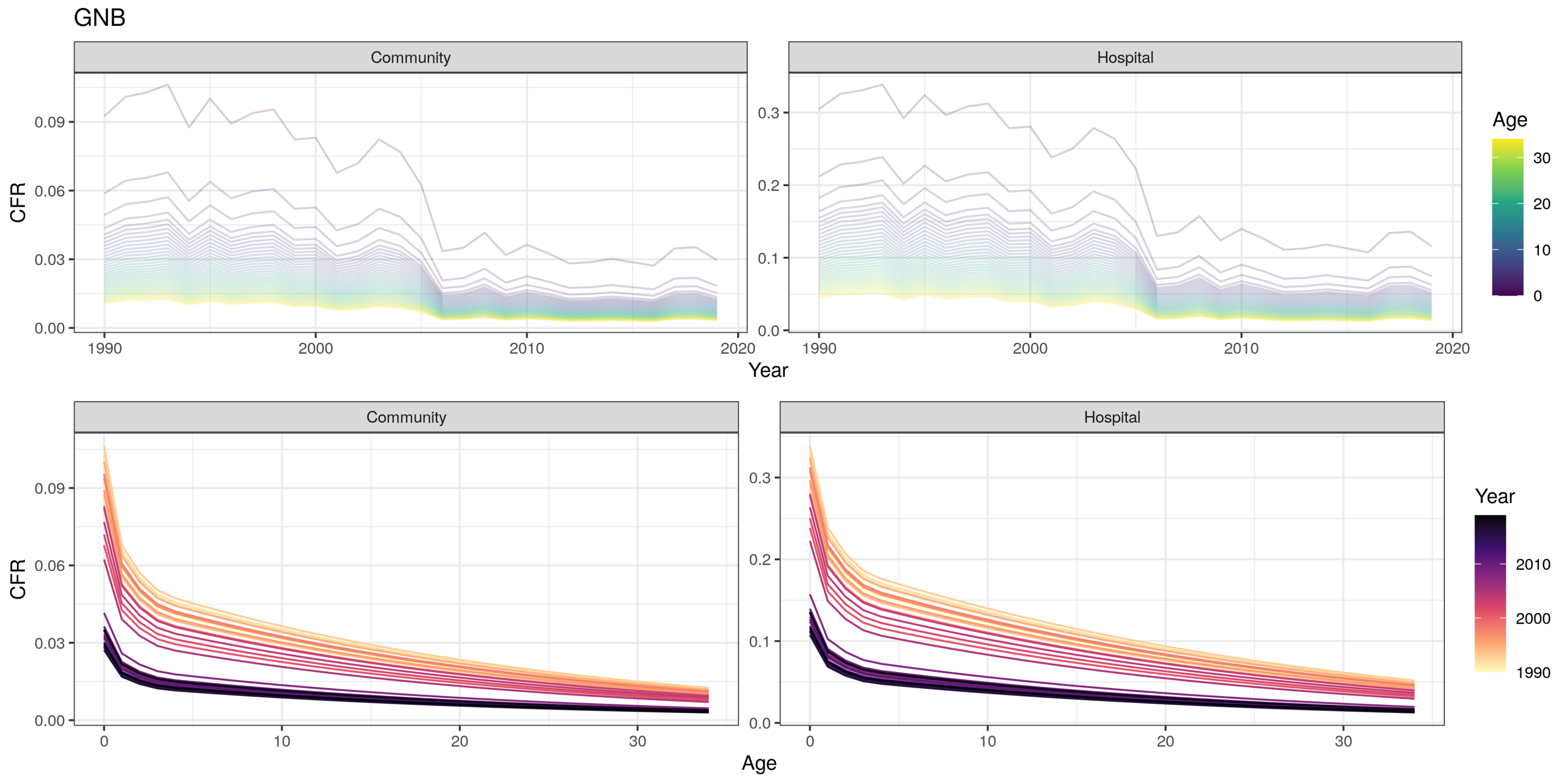
GNB
Community
Hospital
CFR
Year
Age
Community
Hospital
CFR
Age
Year
0.09
0.06
0.03
0.00
0.3
0.2
0.1
0.0
1990
2000
2010
2020
0
10
20
30

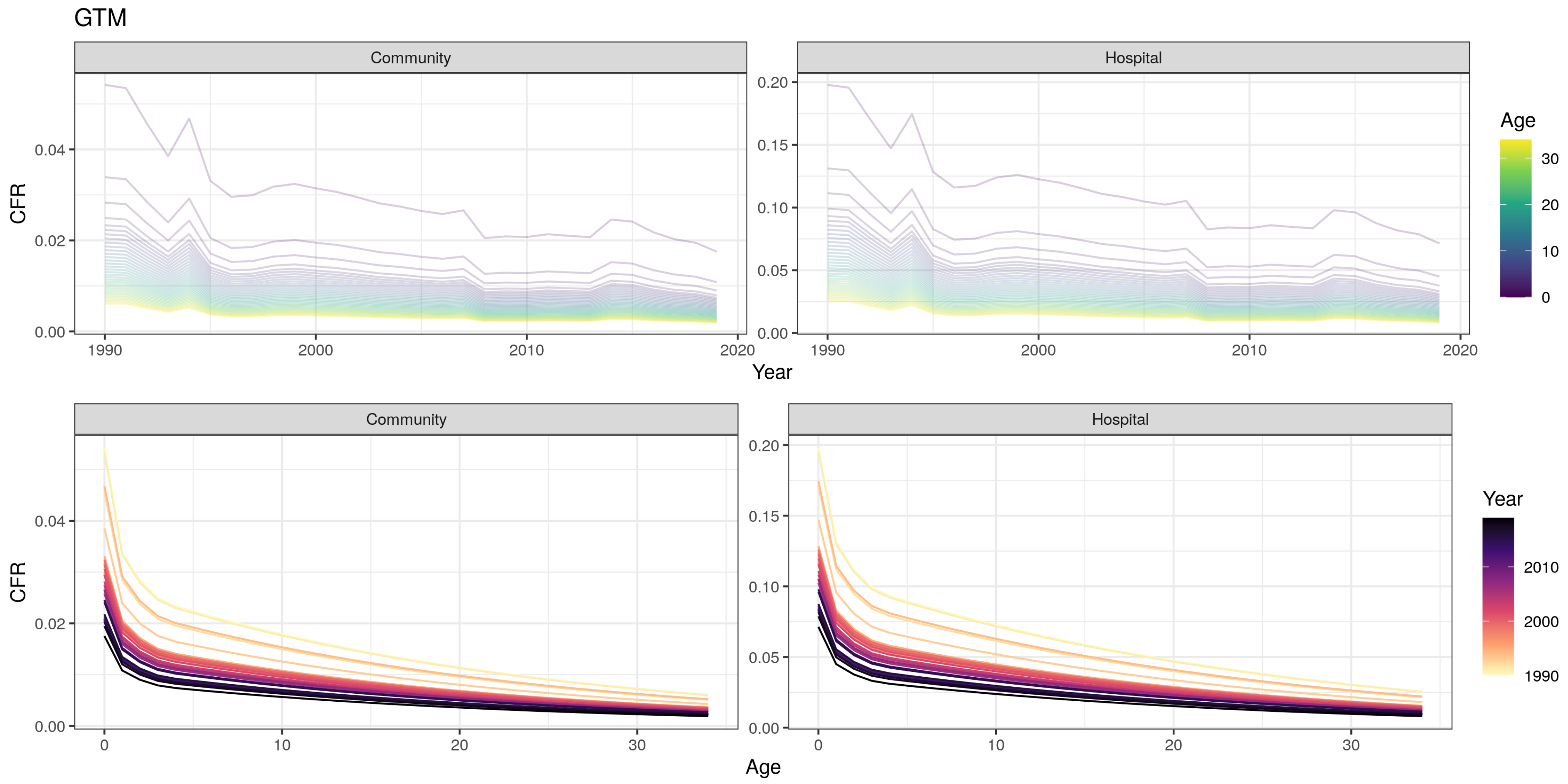
GTM
Community
Hospital
CFR
Year
Age
Community
Hospital
CFR
Age
Year

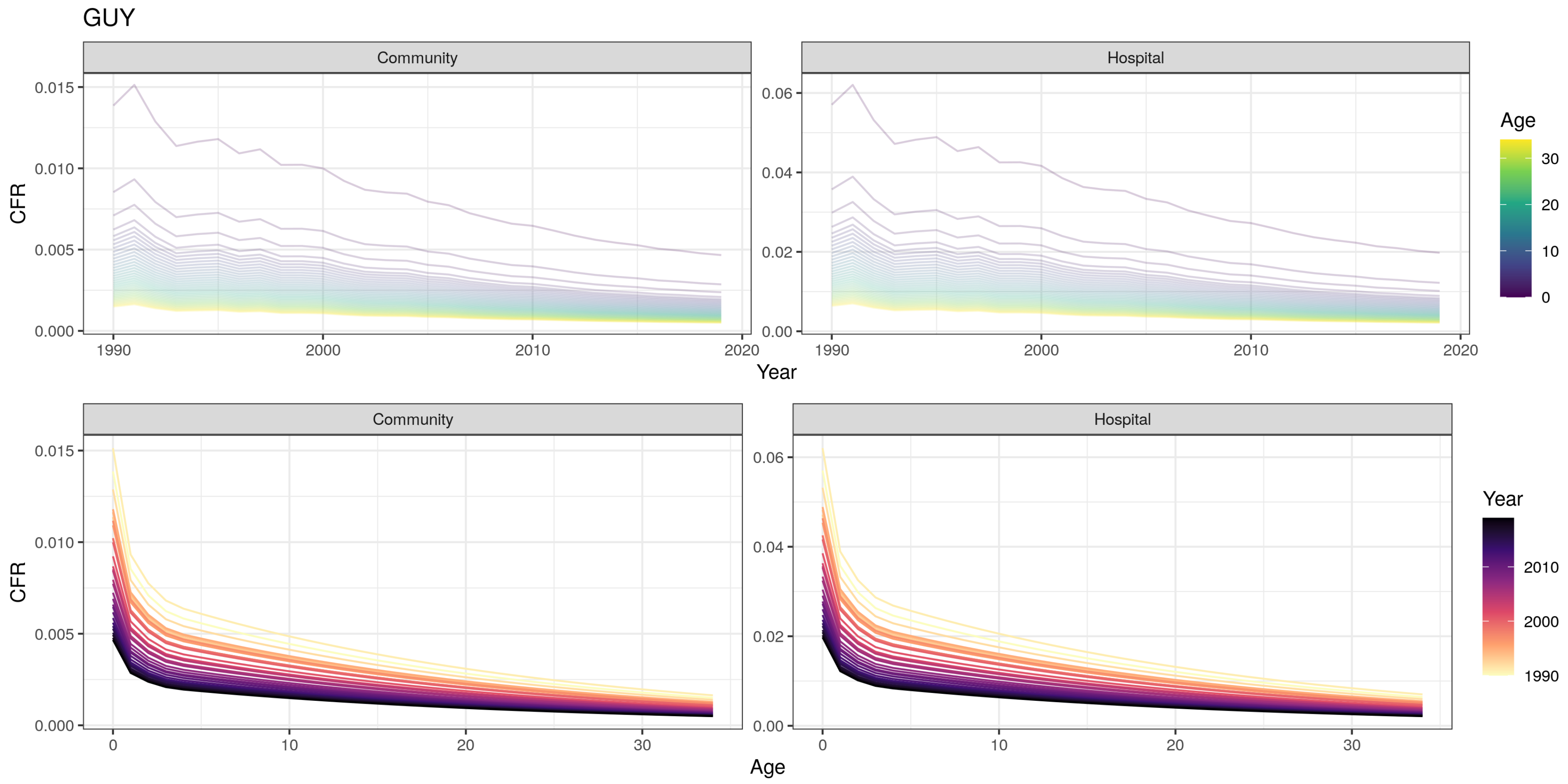

GUY
Community
Hospital
CFR
Year
Age
Community
Hospital
CFR
Age
Year

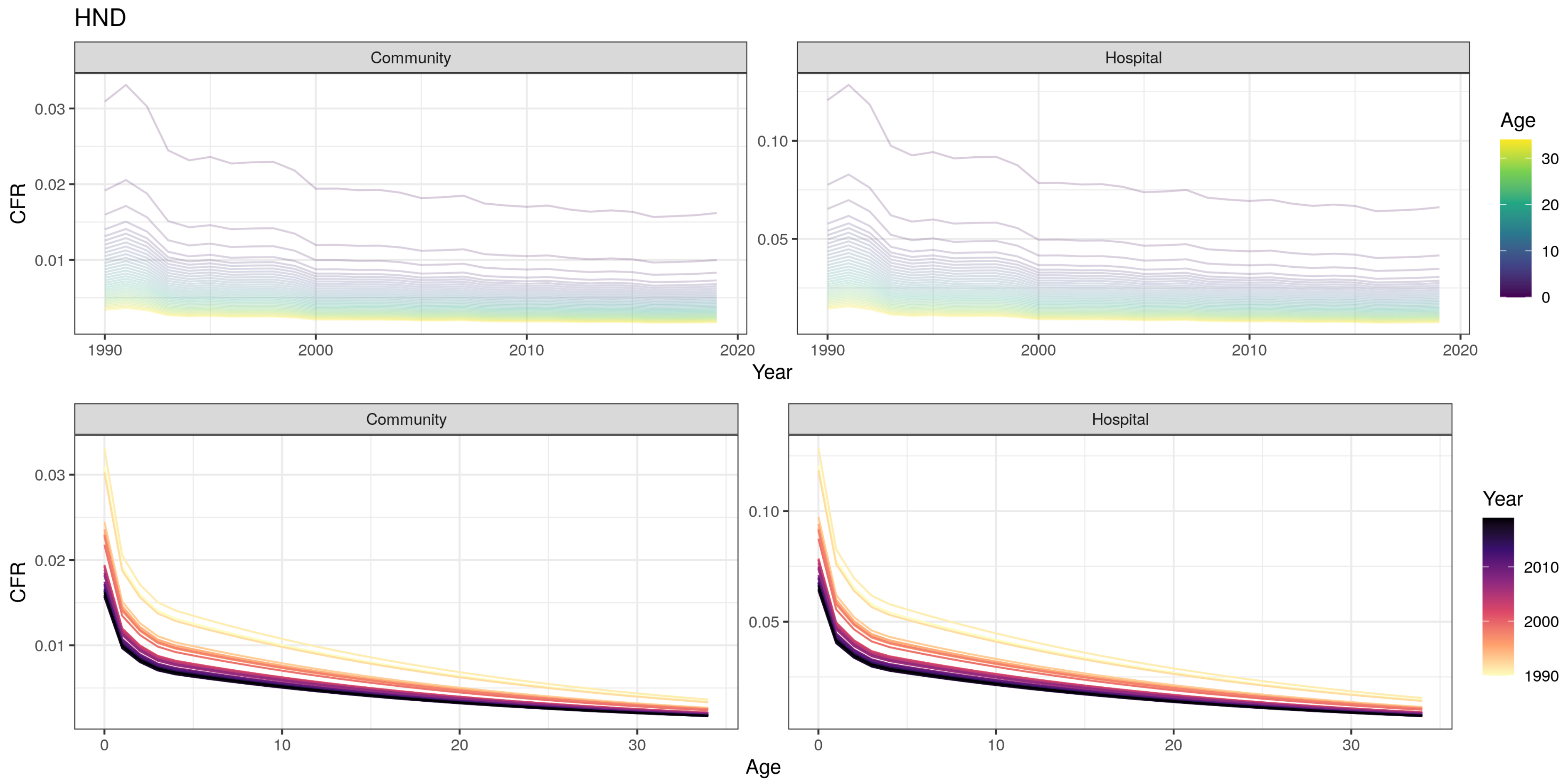
HND
Community
Hospital
CFR
Year
Age
0.03
0.02
0.01
0.10
0.05
1990
2000
2010
2020
0
10
20
30
Year
2010
2000
1990

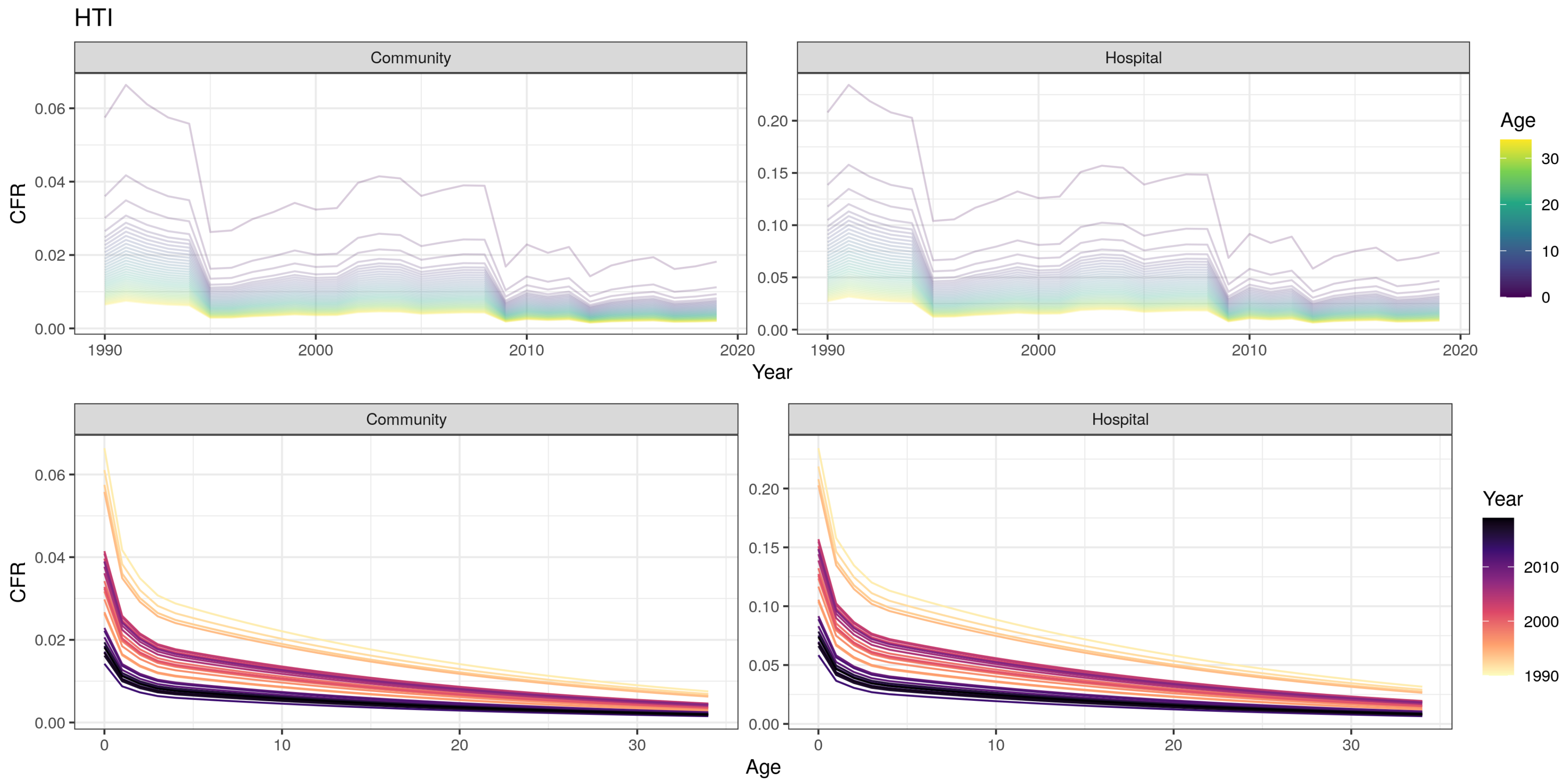
HTI
Community
Hospital
CFR
Year
Age
0.06
0.04
0.02
0.00
0.20
0.15
0.10
0.05
1990
2000
2010
2020
30
20
10
0
Community
Hospital
CFR
Age
Year
2010
2000
1990

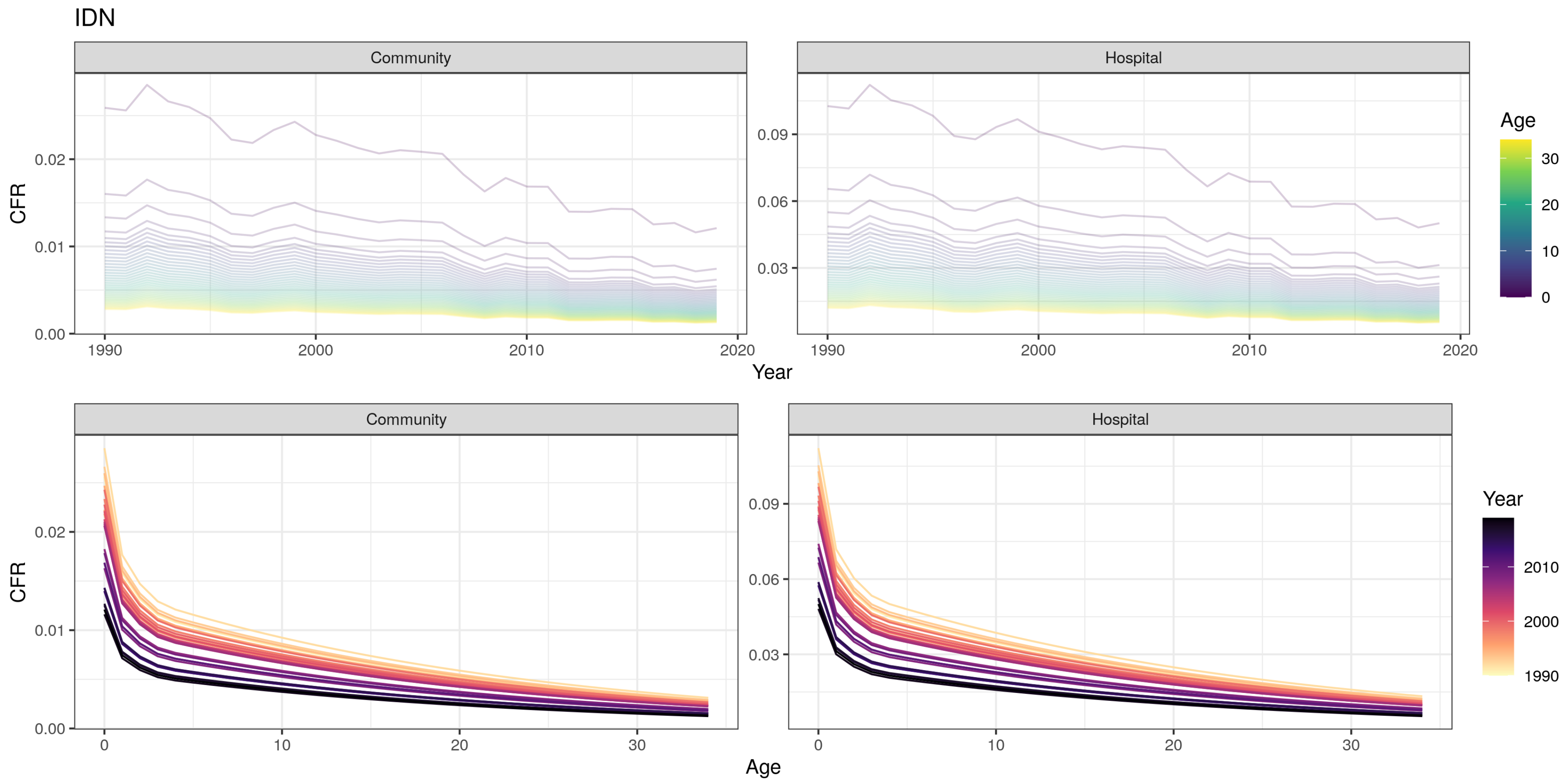
IDN
Community
Hospital
CFR
0.00
0.01
0.02
0.03
0.06
0.09
1990
2000
2010
2020
Year
Age
0
10
20
30
Community
Hospital
CFR
0.00
0.01
0.02
0.03
0.06
0.09
0
10
20
30
Age
Year
1990
2000
2010

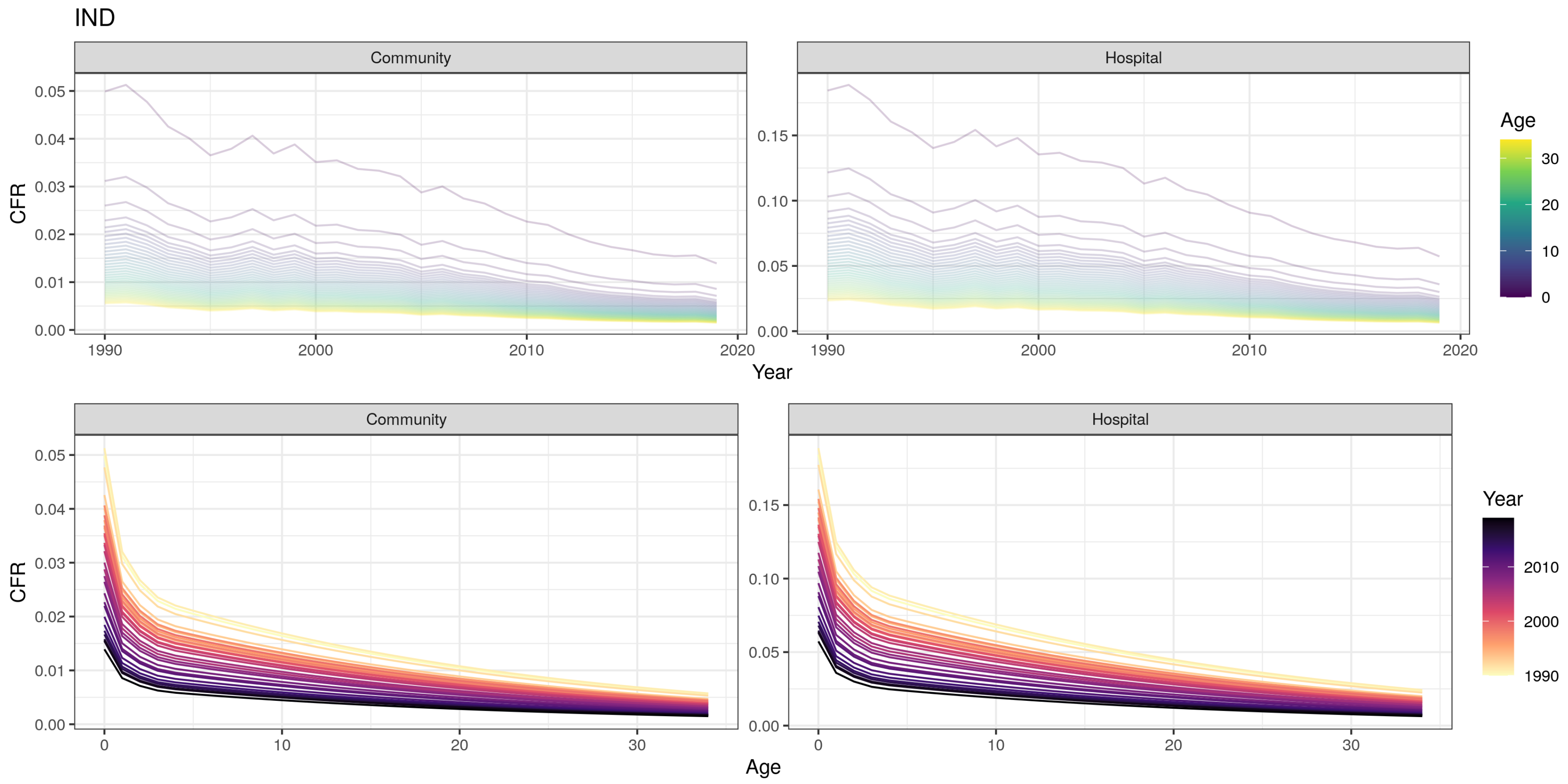
IND
Community
Hospital
CFR
0.05
0.04
0.03
0.02
0.01
0.00
0.15
0.10
0.05
0.00
1990
2000
2010
2020
Year
Age
30
20
10
0
Community
Hospital
CFR
0
10
20
30
Age
Year
2010
2000
1990

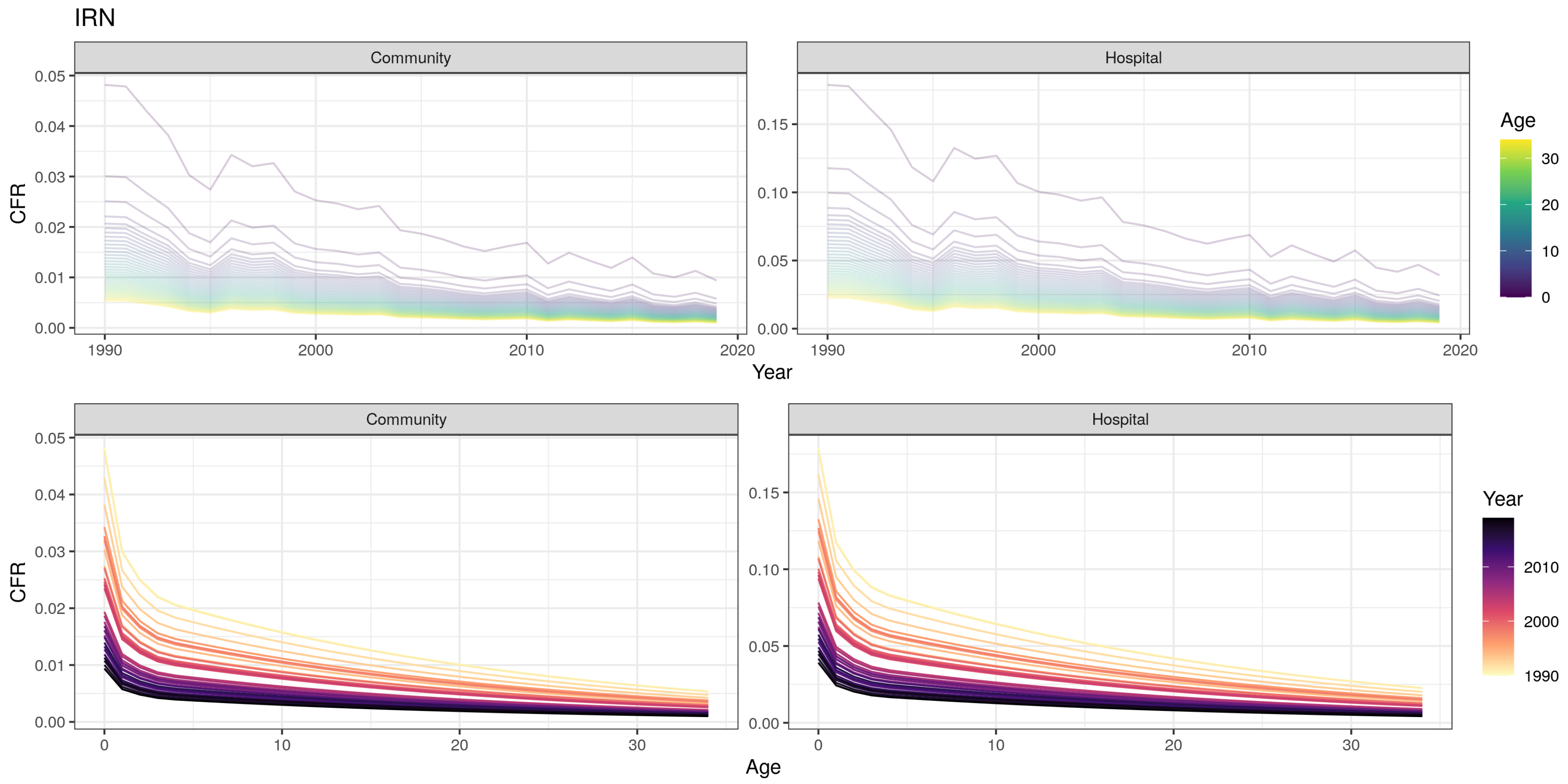

IRN
Community
Hospital
CFR
Year
Age
Community
Hospital
CFR
Age
Year

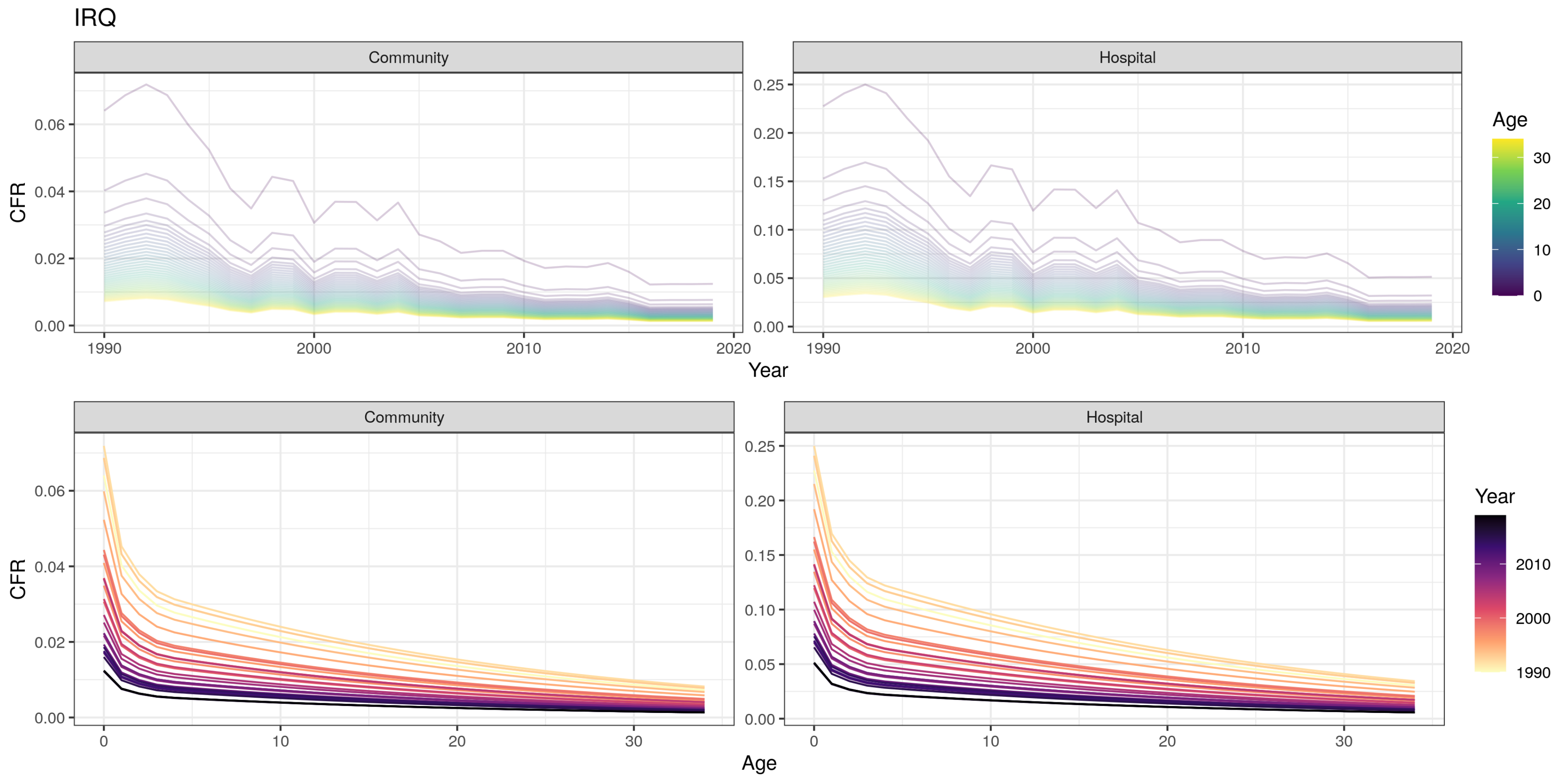
IRQ
Community
Hospital
CFR
Year
Age
0.00
0.02
0.04
0.06
0.05
0.10
0.15
0.20
0.25
1990
2000
2010
2020
0
10
20
30

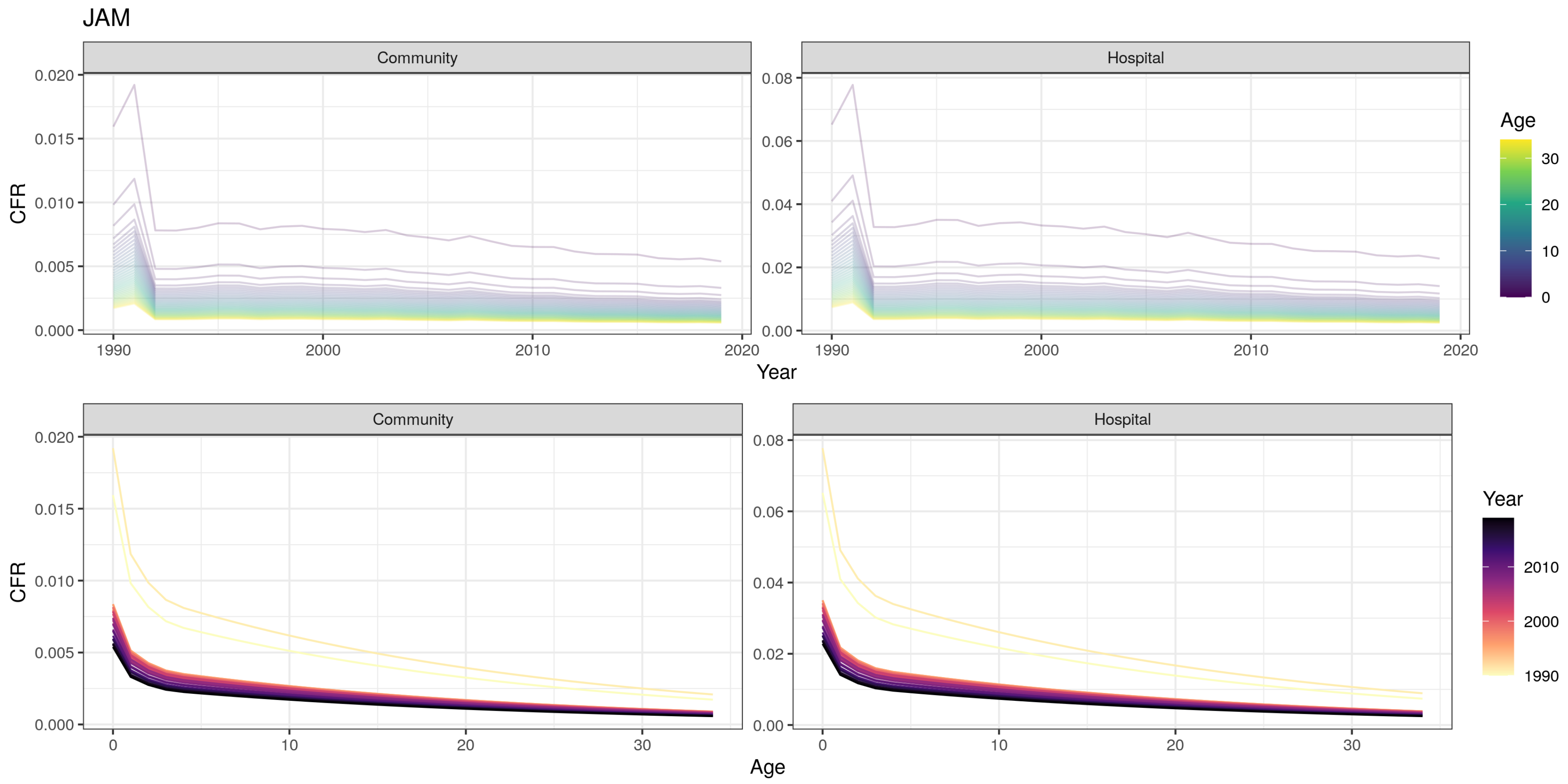
JAM
Community
Hospital
CFR
Year
Age
Community
Hospital
CFR
Age
Year

JOR

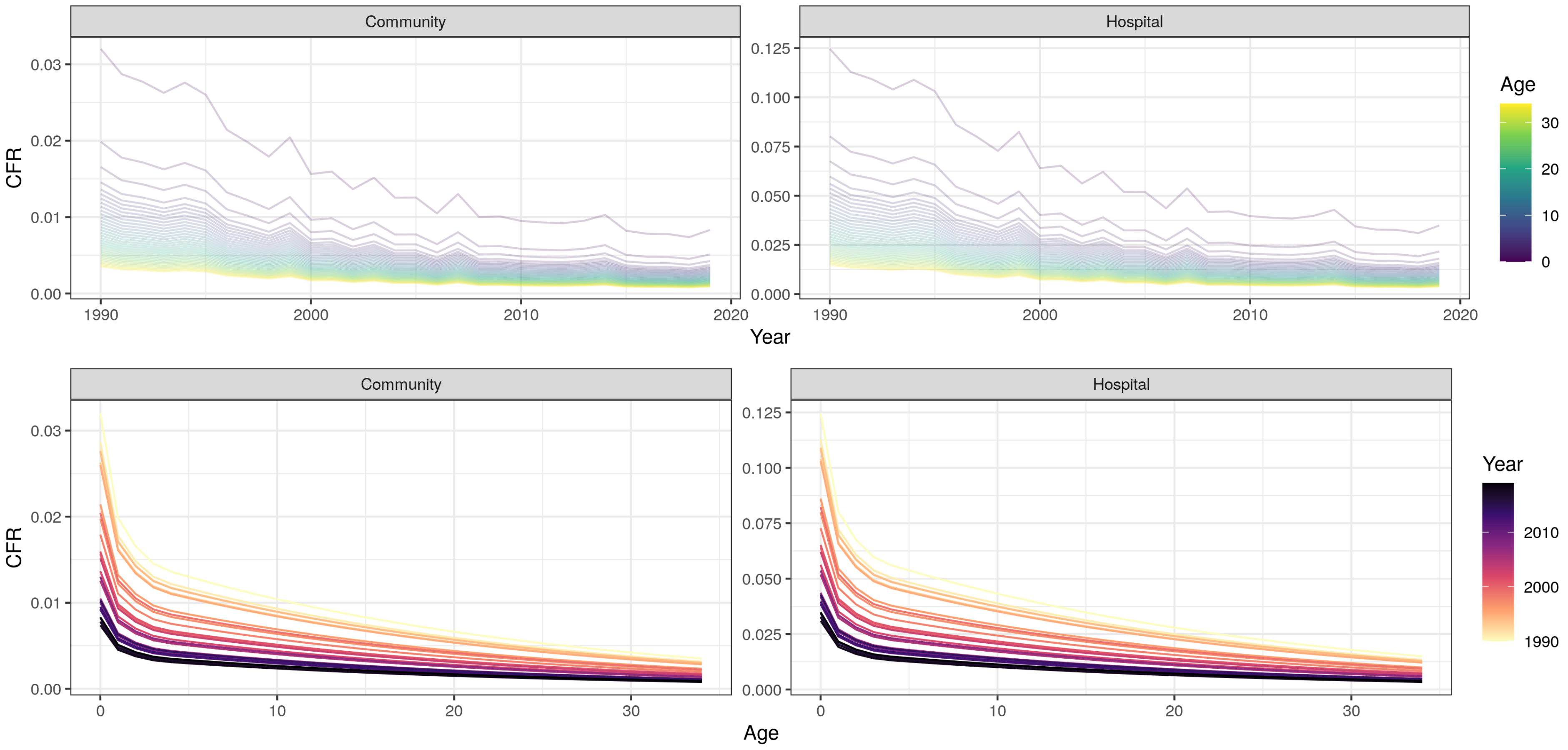

Community
Hospital
CFR
Year
Age
0.00
0.01
0.02
0.03
0.000
0.025
0.050
0.075
0.100
0.125
1990
2000
2010
2020
0
10
20
30

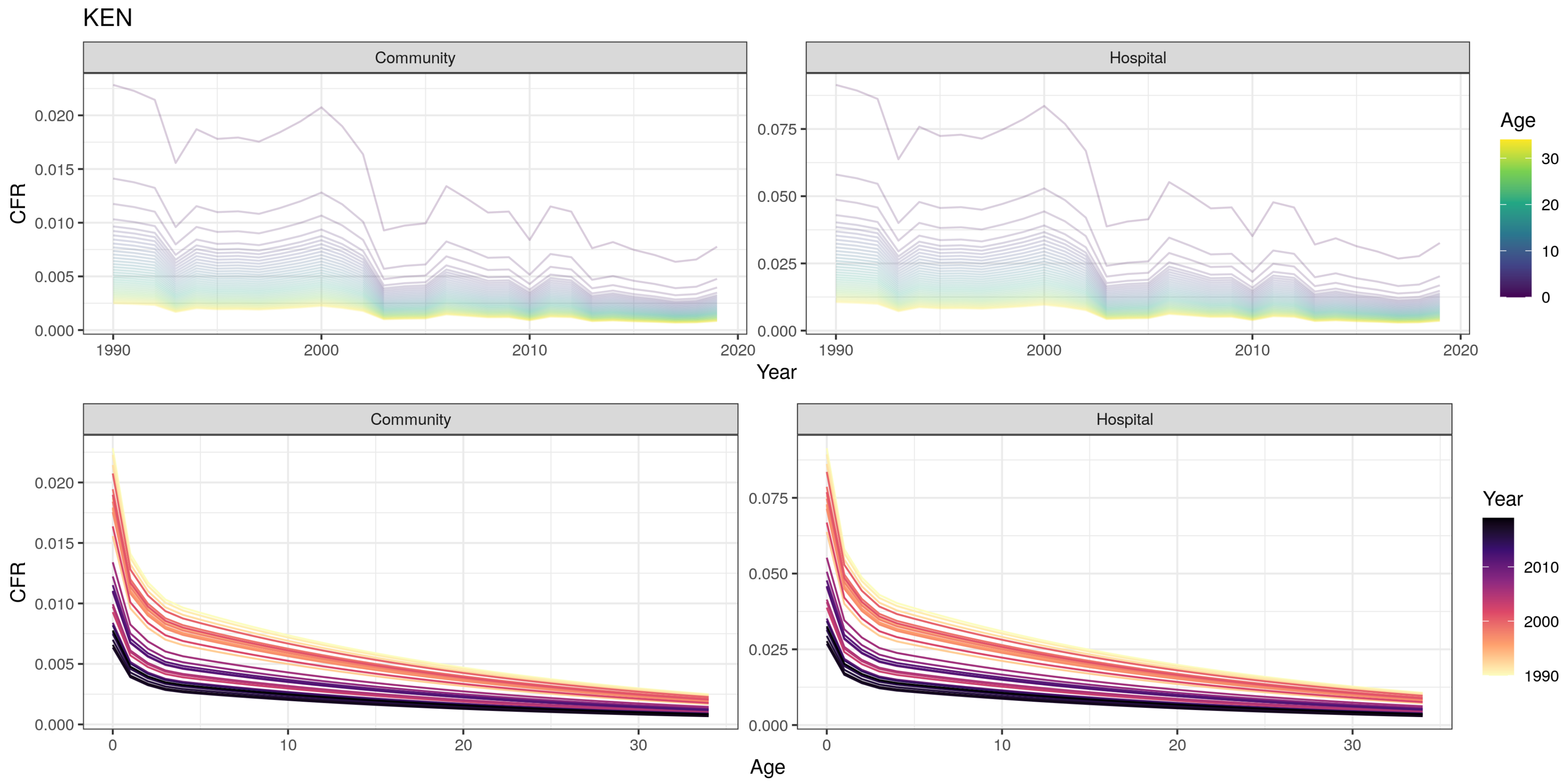
KEN
Community
Hospital
CFR
0.000
0.005
0.010
0.015
0.020
0.025
0.050
0.075
1990
2000
2010
2020
Year
Age
30
20
10
0
Community
Hospital
CFR
0
10
20
30
Age
Year
2010
2000
1990

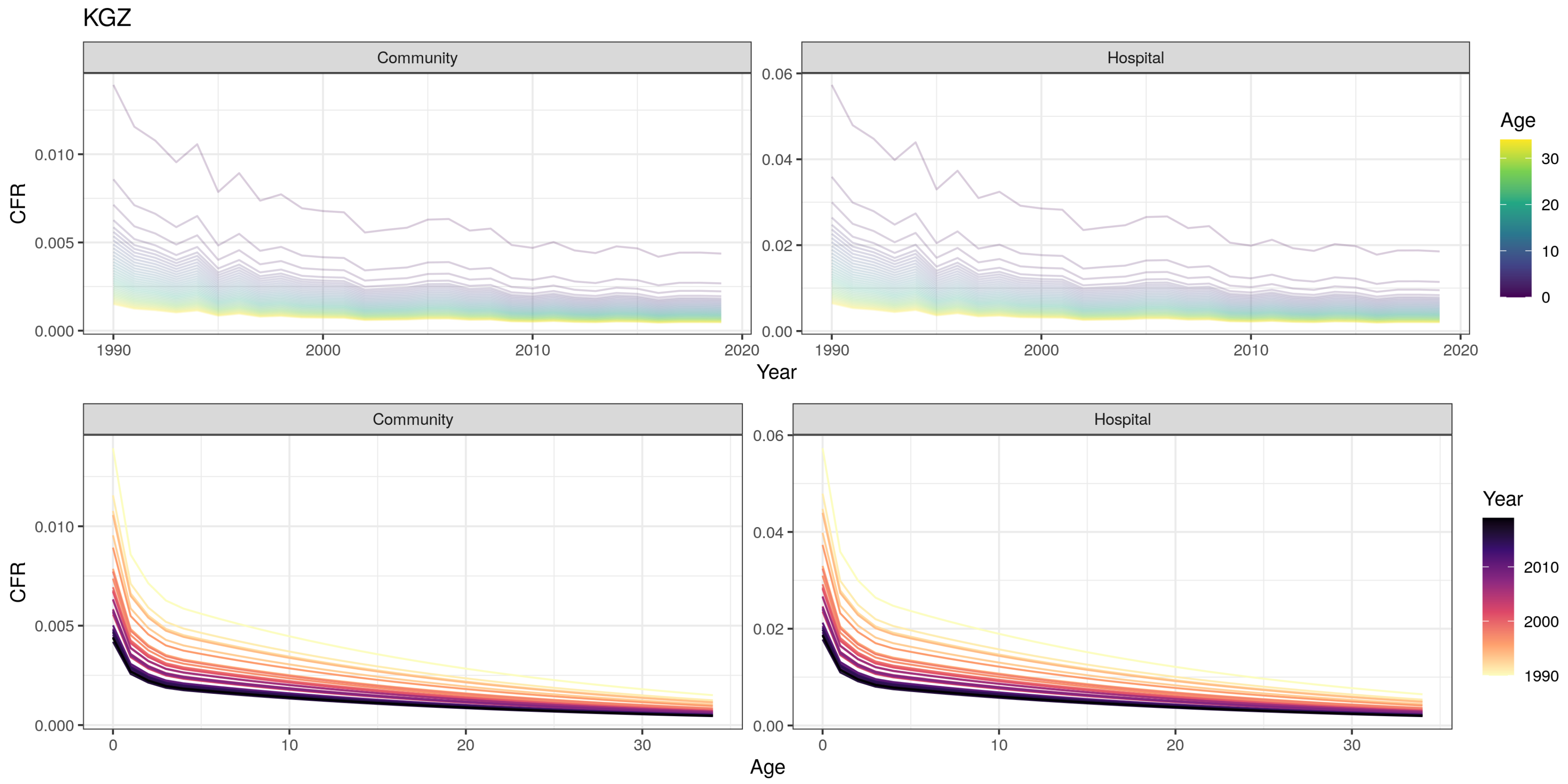
KGZ
Community
Hospital
CFR
Year
Age
1990
2000
2010
2020
0.000
0.005
0.010
0.00
0.02
0.04
0.06
0
10
20
30

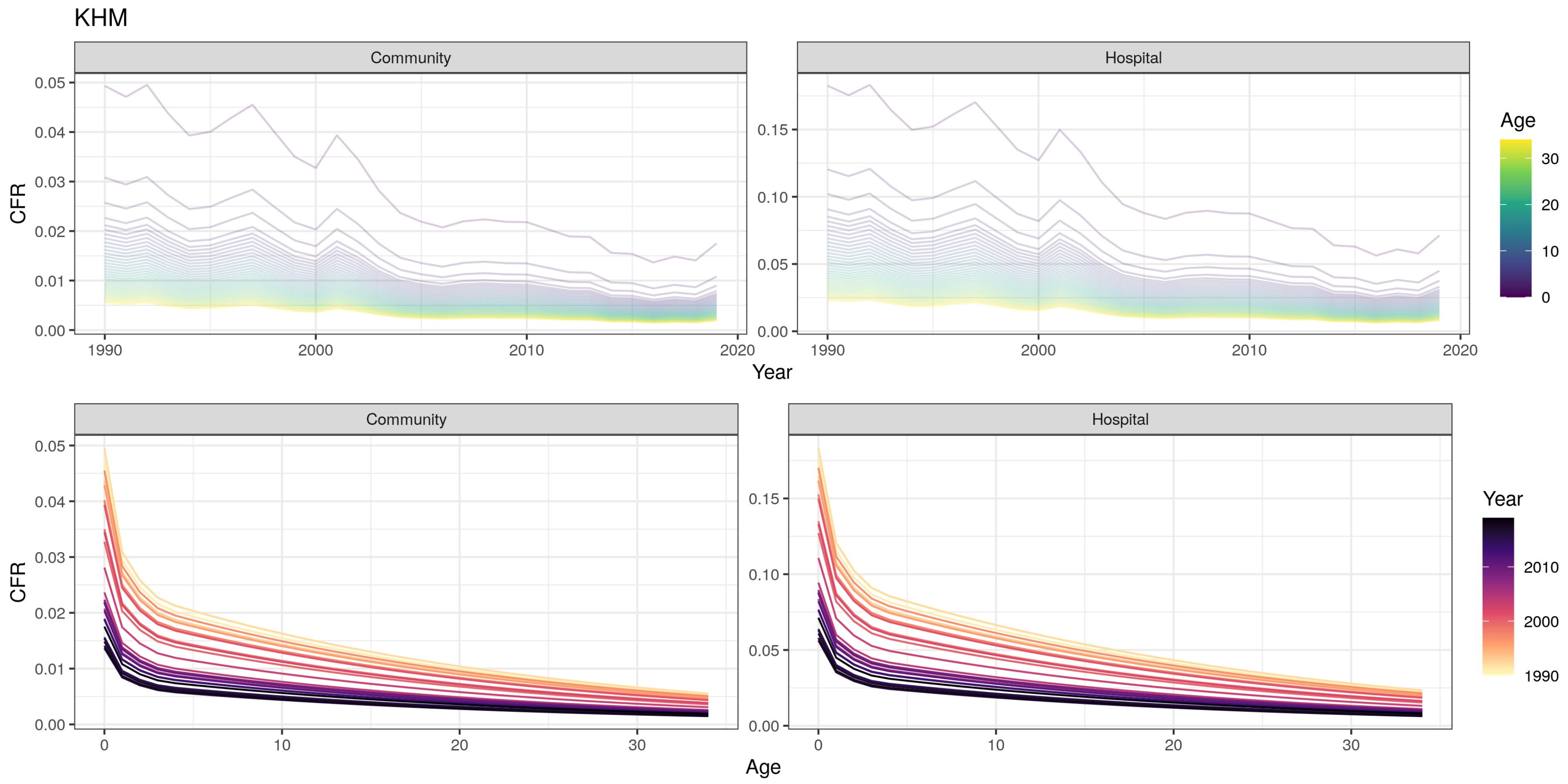

KHM
Community
Hospital
CFR
Year
Age
0.00
0.01
0.02
0.03
0.04
0.05
0.10
0.15
1990
2000
2010
2020
0
10
20
30

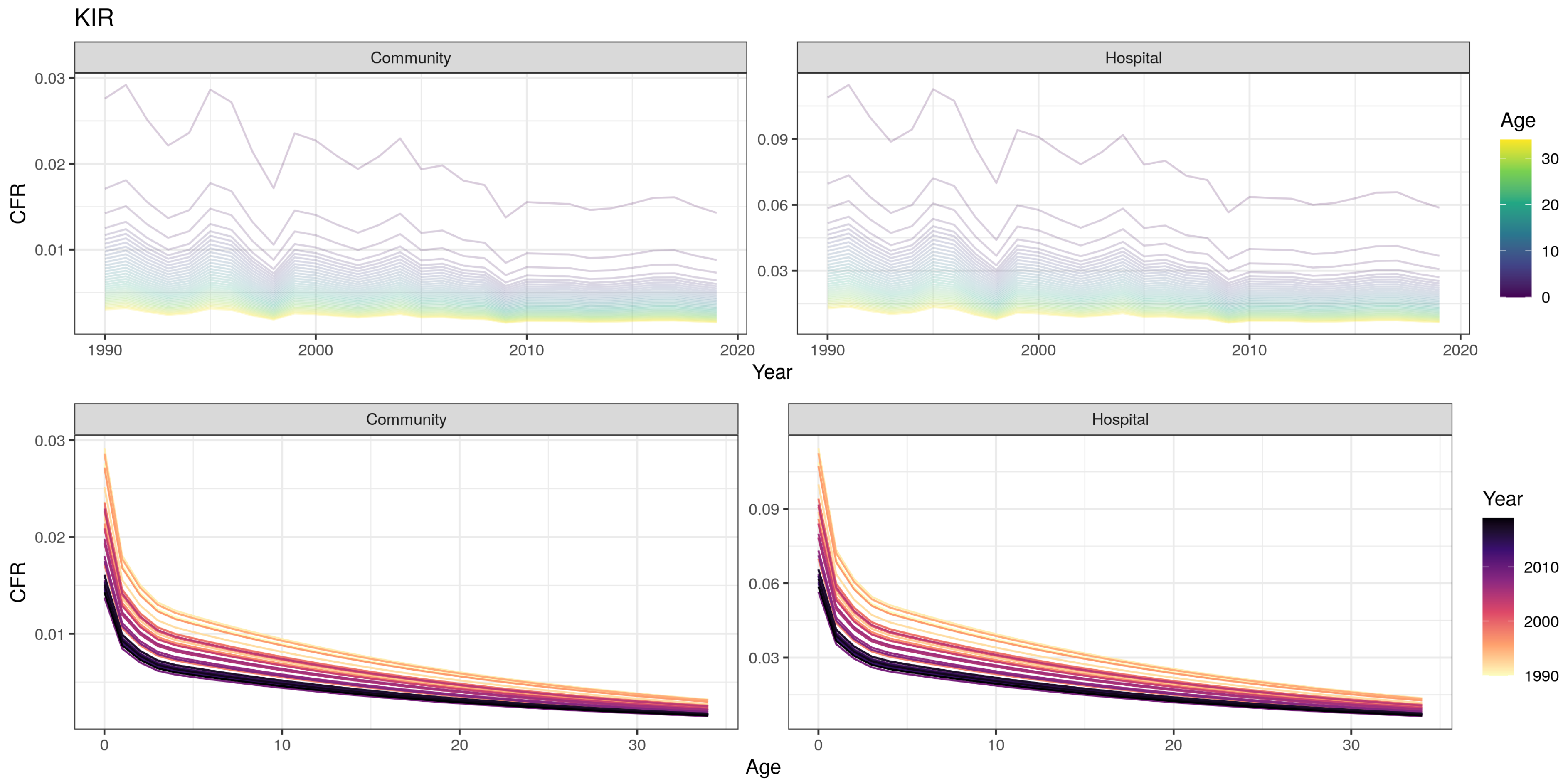

KIR
Community
Hospital
CFR
0.03
0.02
0.01
0.09
0.06
0.03
1990
2000
2010
2020
Year
Age
30
20
10
0
Community
Hospital
Age
Year
2010
2000
1990

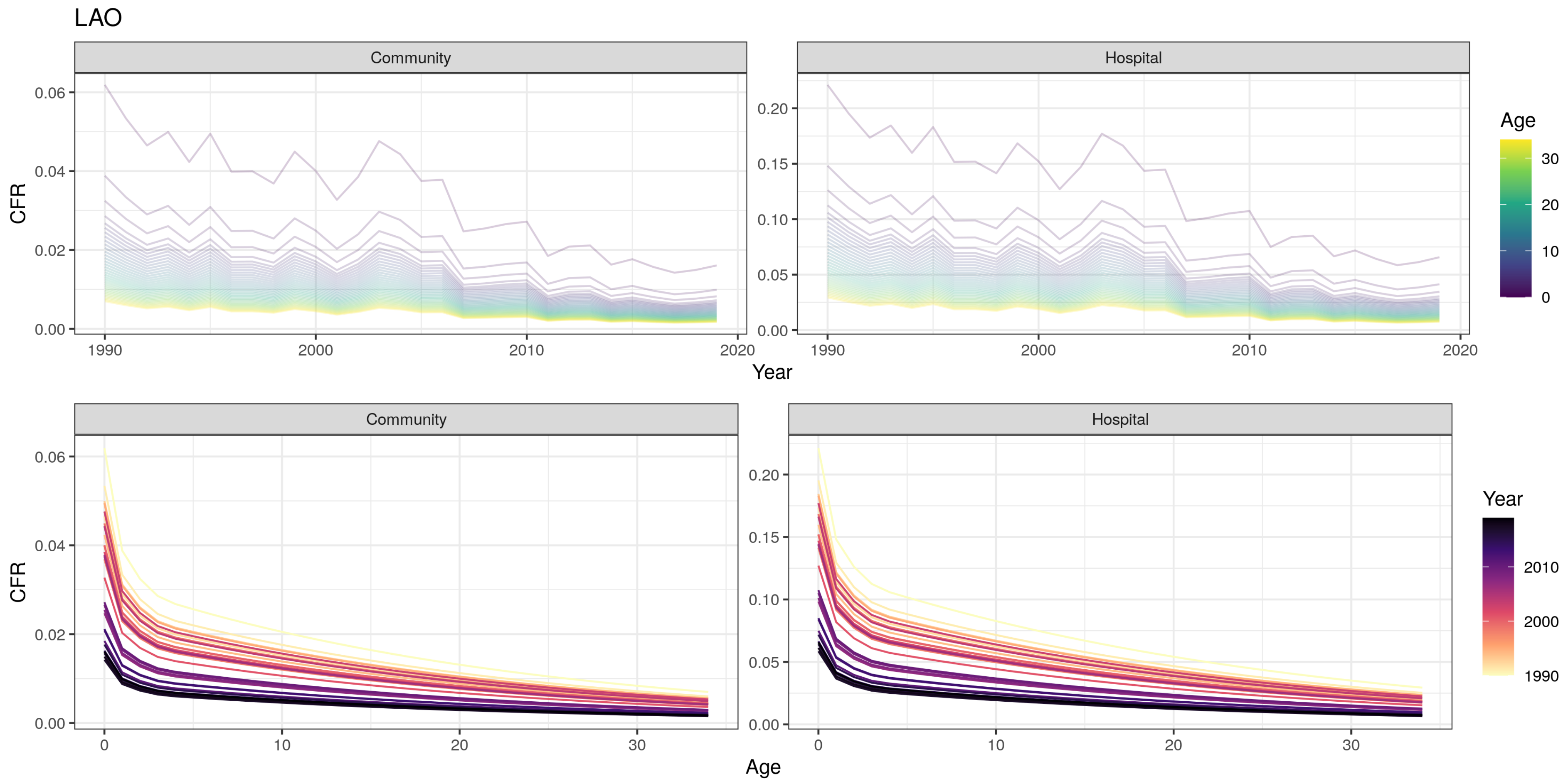

LAO
Community
Hospital
CFR
Year
Age
0.00
0.02
0.04
0.06
0.05
0.10
0.15
0.20
1990
2000
2010
2020
0
10
20
30
Age
Year

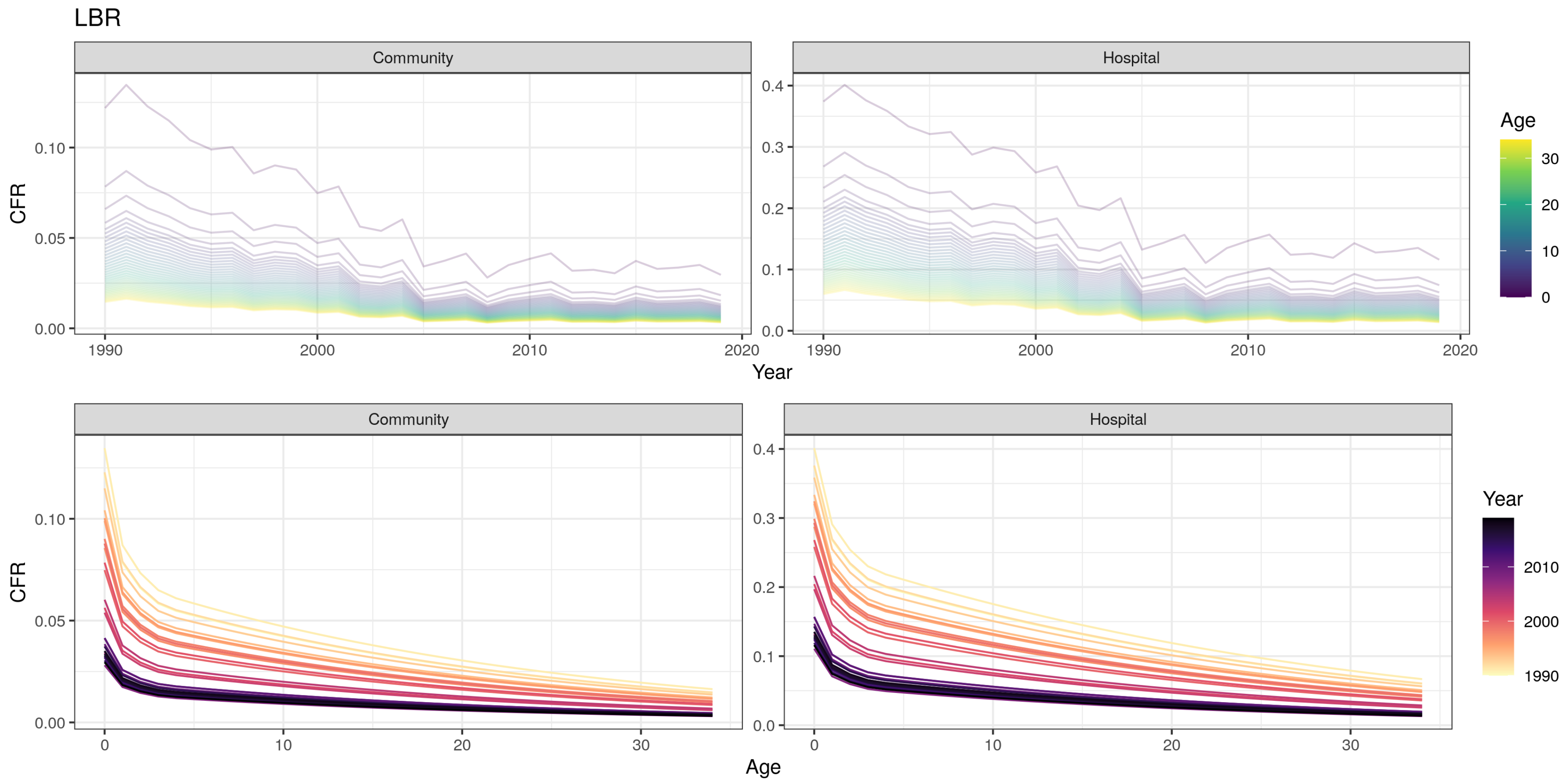
LBR
Community
Hospital
CFR
0.00
0.05
0.10
0.0
0.1
0.2
0.3
0.4
1990
2000
2010
2020
Year
Age
0
10
20
30
Age
Year
1990
2000
2010

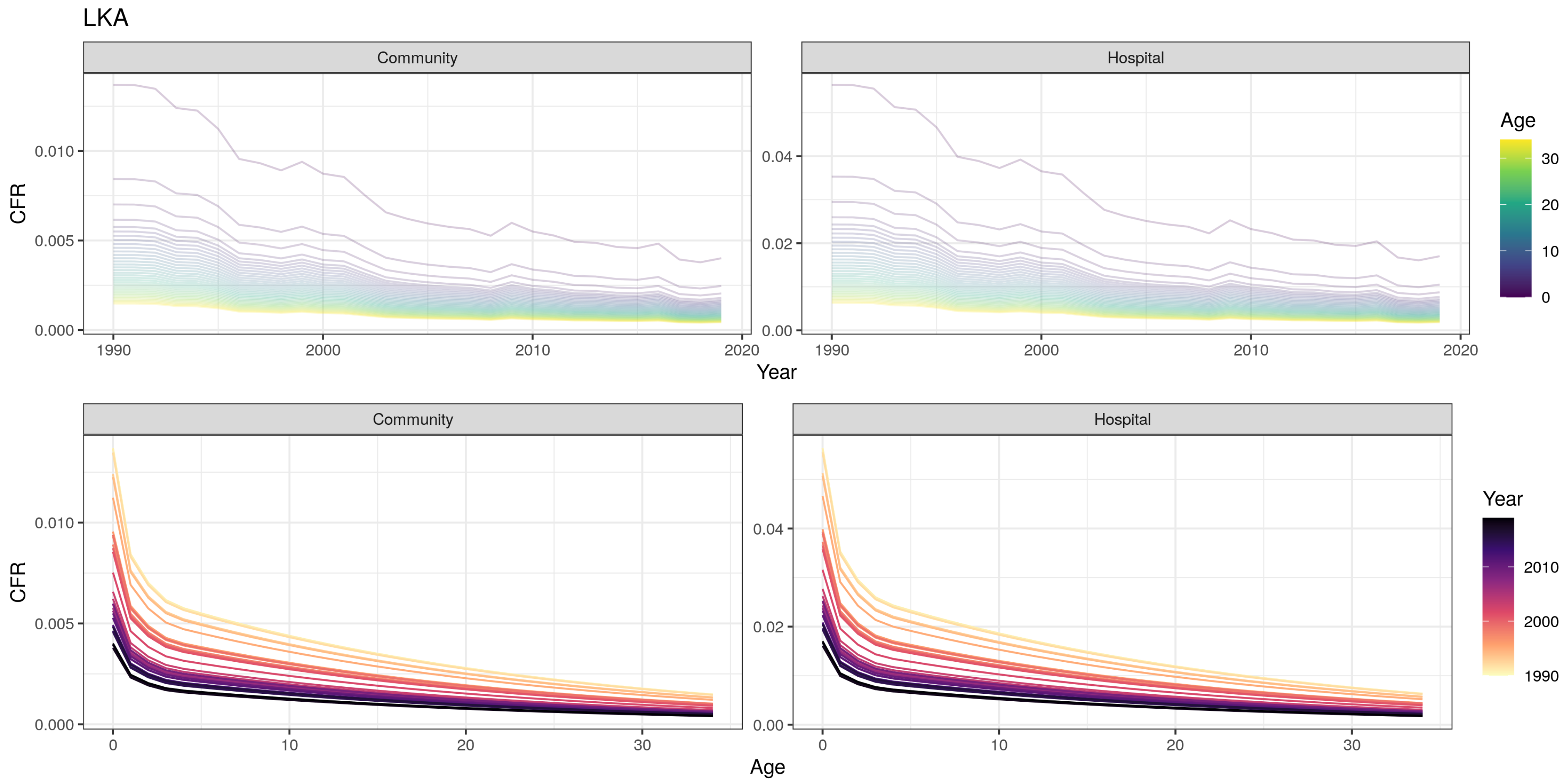

LKA
Community
Hospital
CFR
0.010
0.005
0.000
0.04
0.02
0.00
1990
2000
2010
2020
Year
Age
30
20
10
0
Community
Hospital
CFR
0.010
0.005
0.000
0.04
0.02
0.00
0
10
20
30
Age
Year
2010
2000
1990

LSO

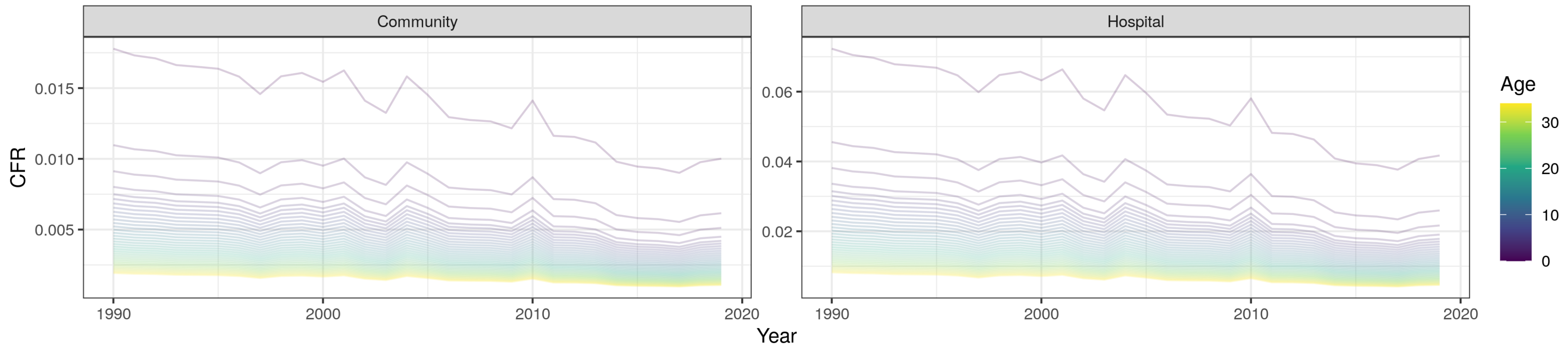

Community
Hospital
CFR
0.015
0.010
0.005
0.06
0.04
0.02
1990
2000
2010
2020
Year
Age
30
20
10
0


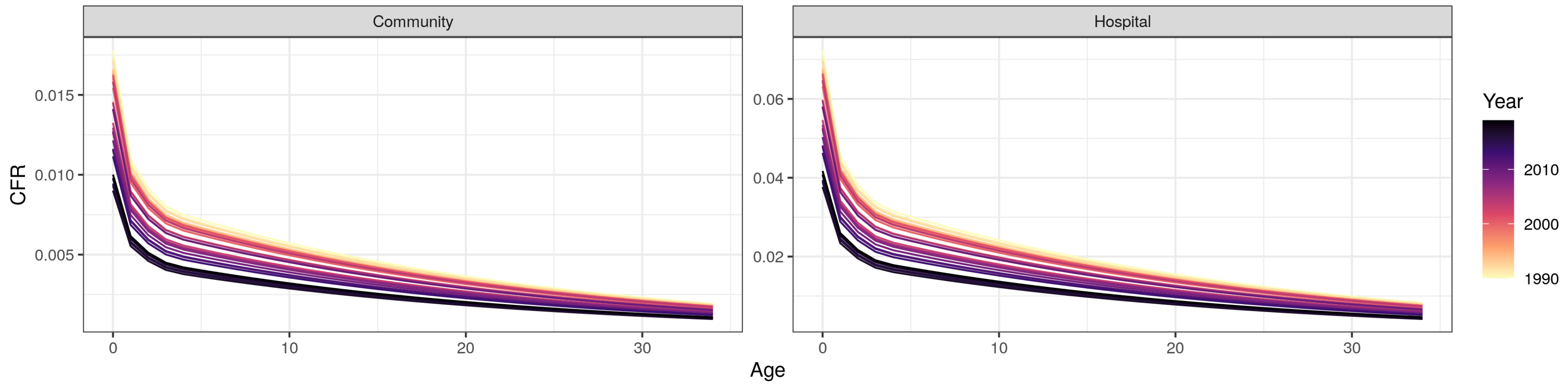

Community
Hospital
CFR
0.015
0.010
0.005
0.06
0.04
0.02
0
10
20
30
Age
Year
2010
2000
1990

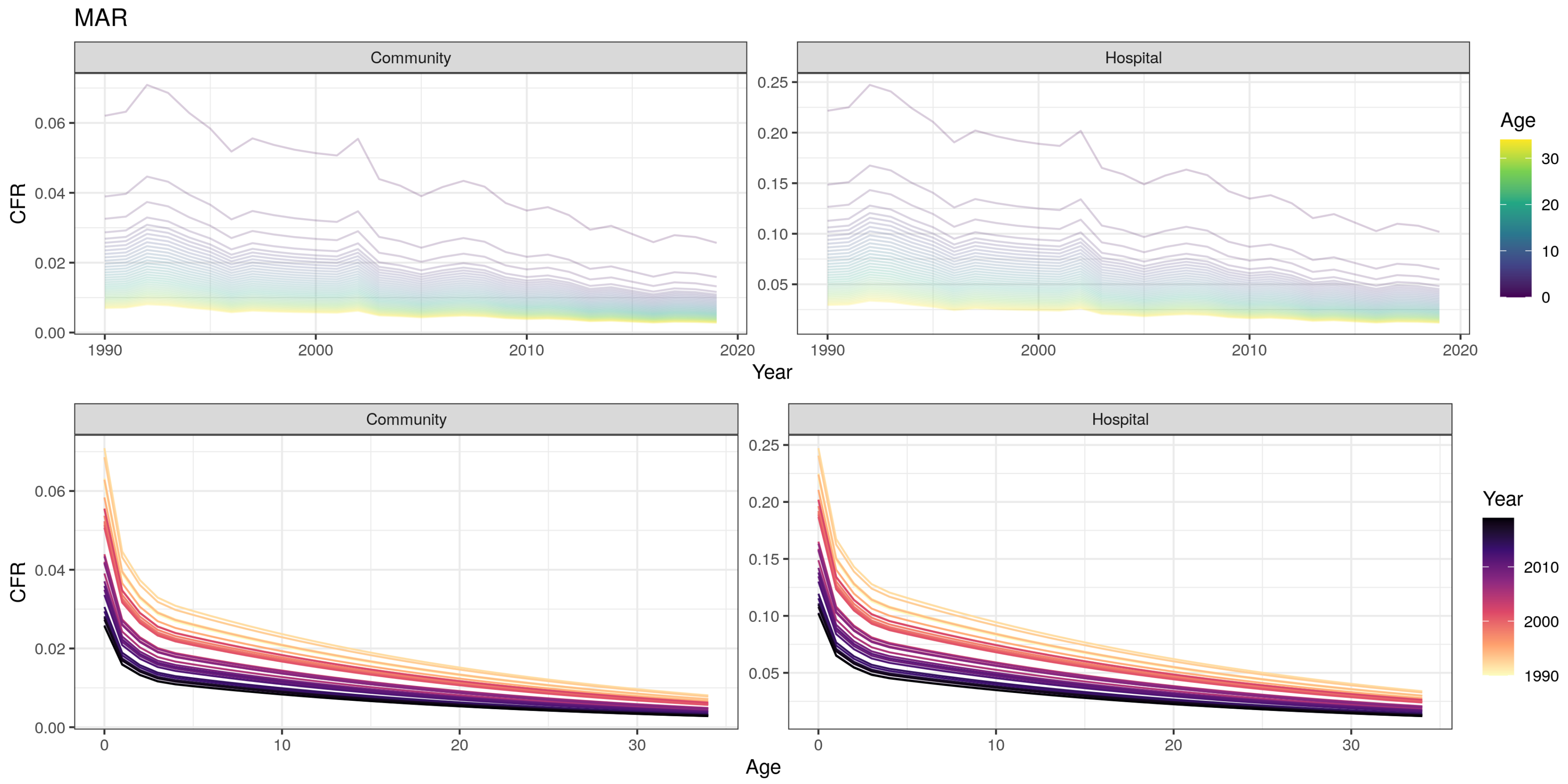
MAR
Community
Hospital
CFR
0.00
0.02
0.04
0.06
0.05
0.10
0.15
0.20
0.25
1990
2000
2010
2020
Year
Age
0
10
20
30
Community
Hospital
CFR
0.00
0.02
0.04
0.06
0.05
0.10
0.15
0.20
0.25
0
10
20
30
Age
Year
1990
2000
2010

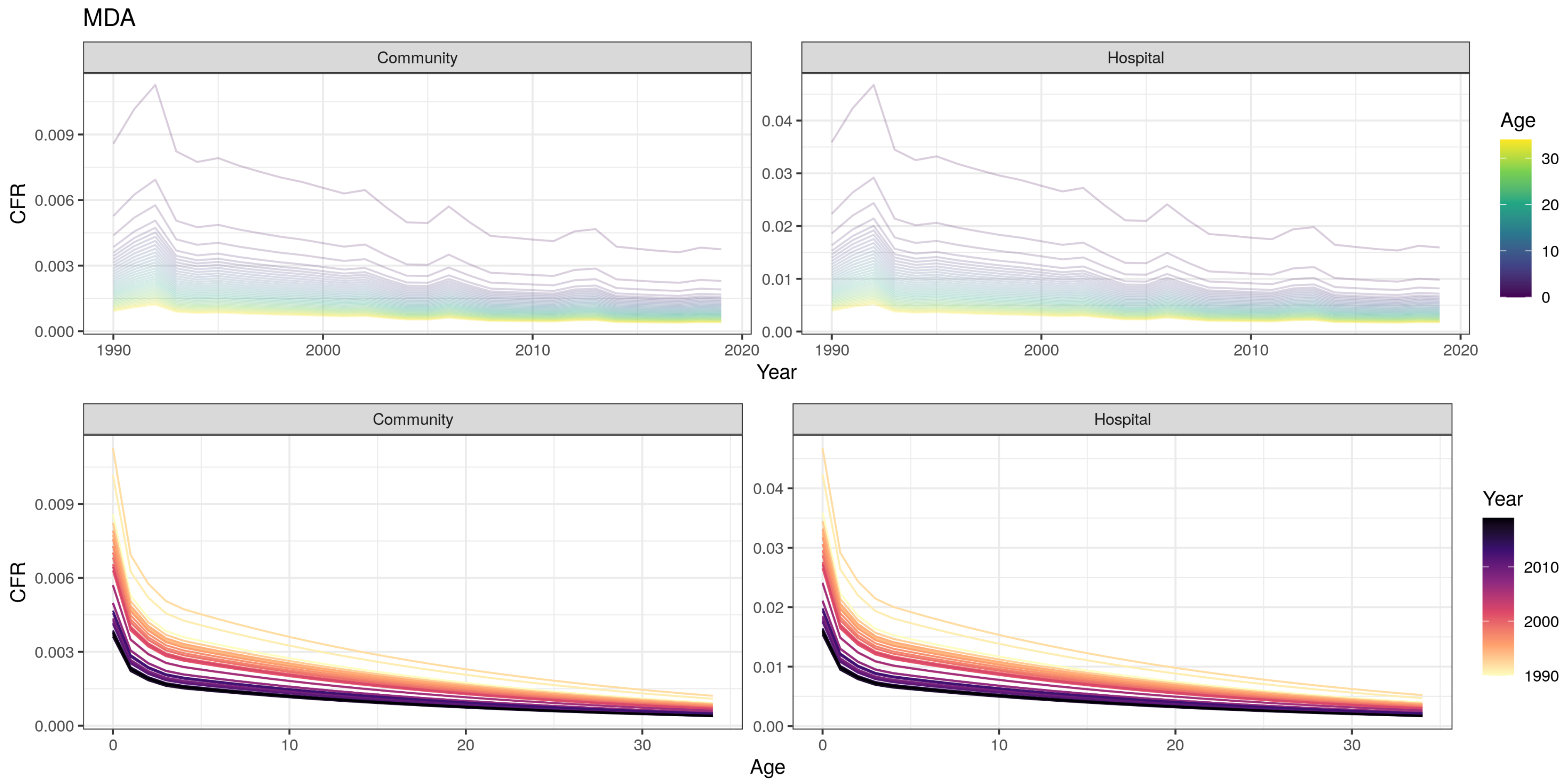

MDA
Community
Hospital
CFR
0.009
0.006
0.003
0.000
0.04
0.03
0.02
0.01
0.00
1990
2000
2010
2020
Year
Age
30
20
10
0
Community
Hospital
CFR
0
10
20
30
Age
Year
2010
2000
1990

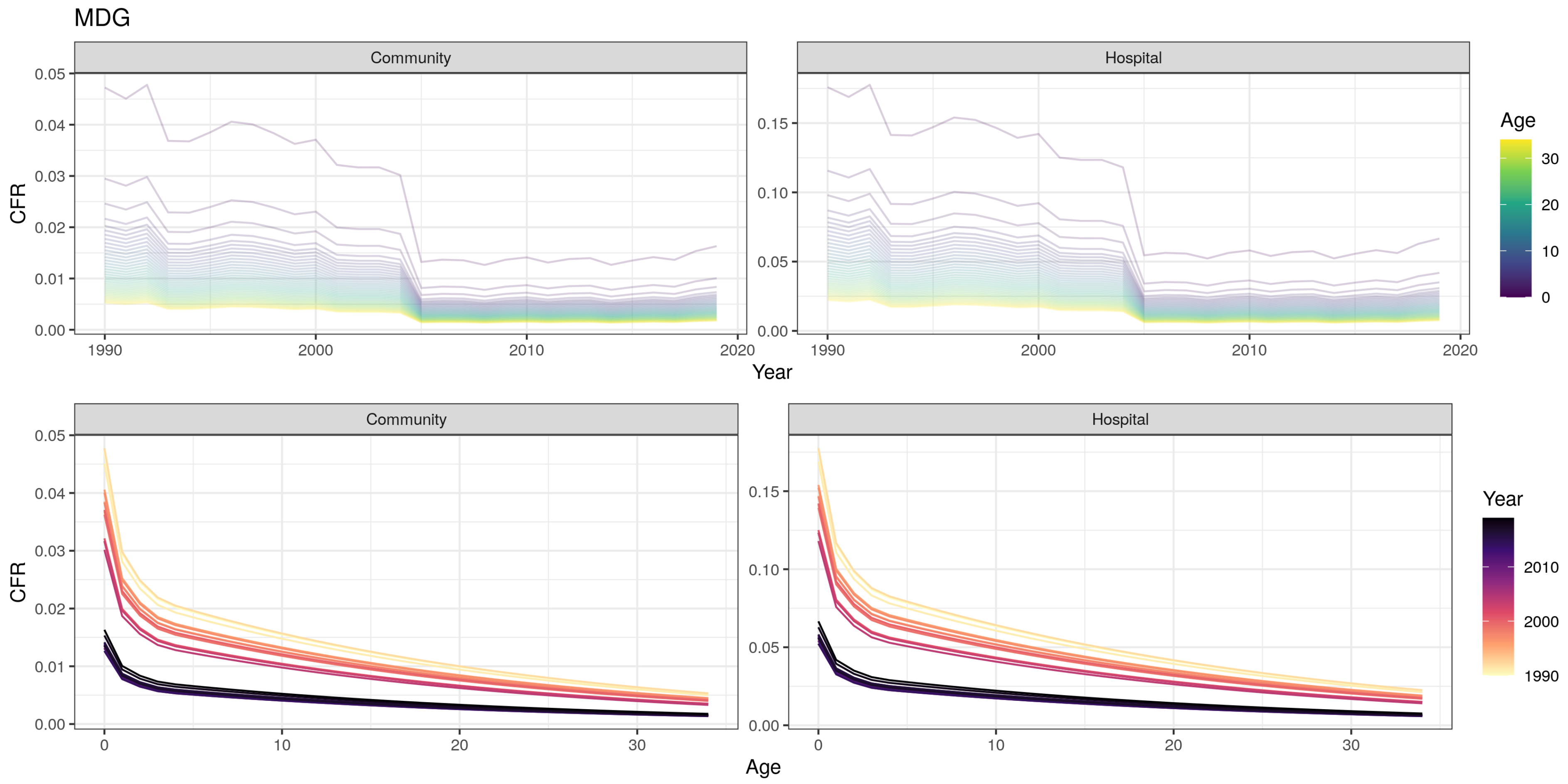

MDG
Community
Hospital
CFR
Year
Age
0.00
0.01
0.02
0.03
0.04
0.05
0.05
0.10
0.15
1990
2000
2010
2020
0
10
20
30

MHL

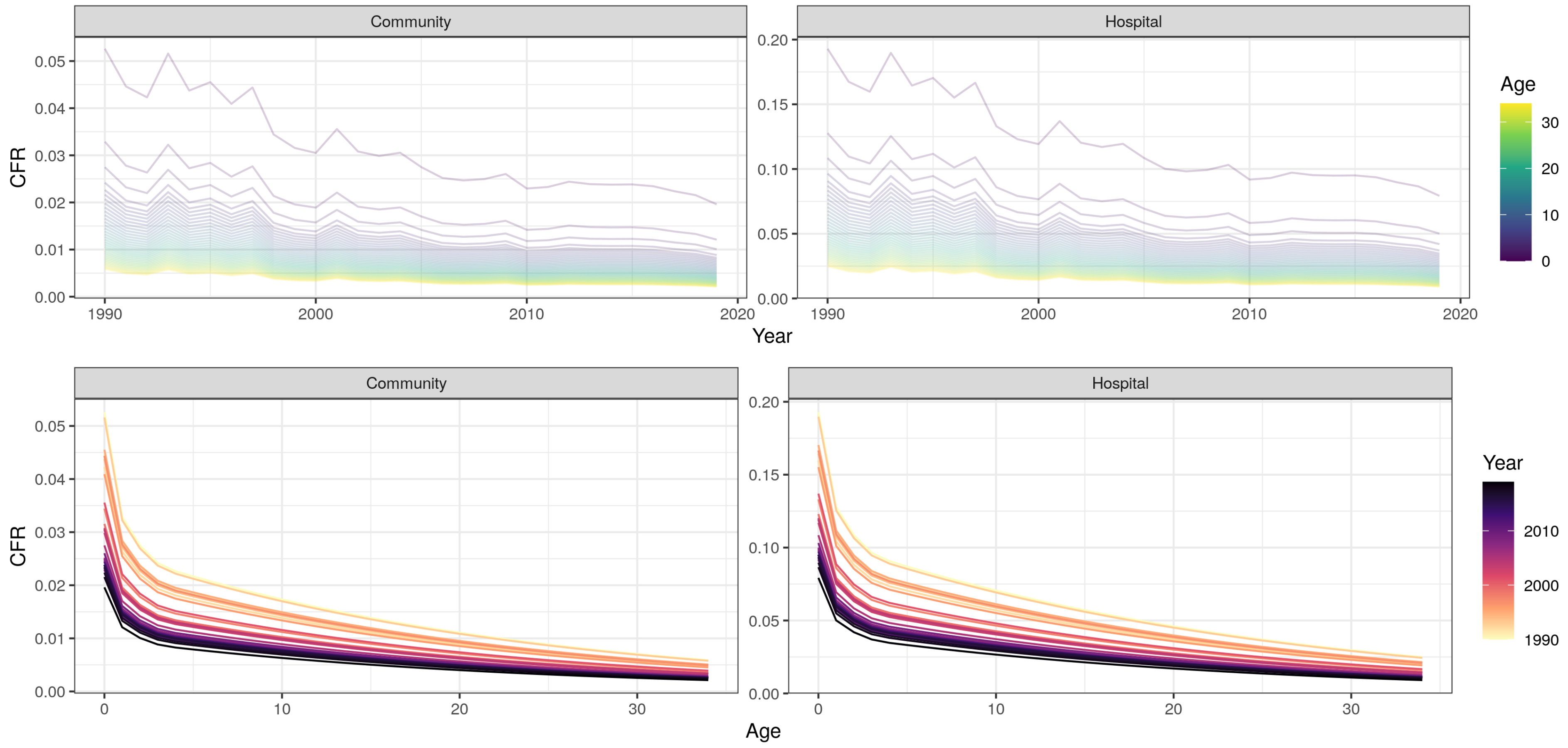

Community
Hospital
CFR
0.05
0.04
0.03
0.02
0.01
0.00
0.20
0.15
0.10
0.05
0.00
1990
2000
2010
2020
Year
Age
30
20
10
0
Community
Hospital
CFR
0
10
20
30
Age
Year
2010
2000
1990

MKD

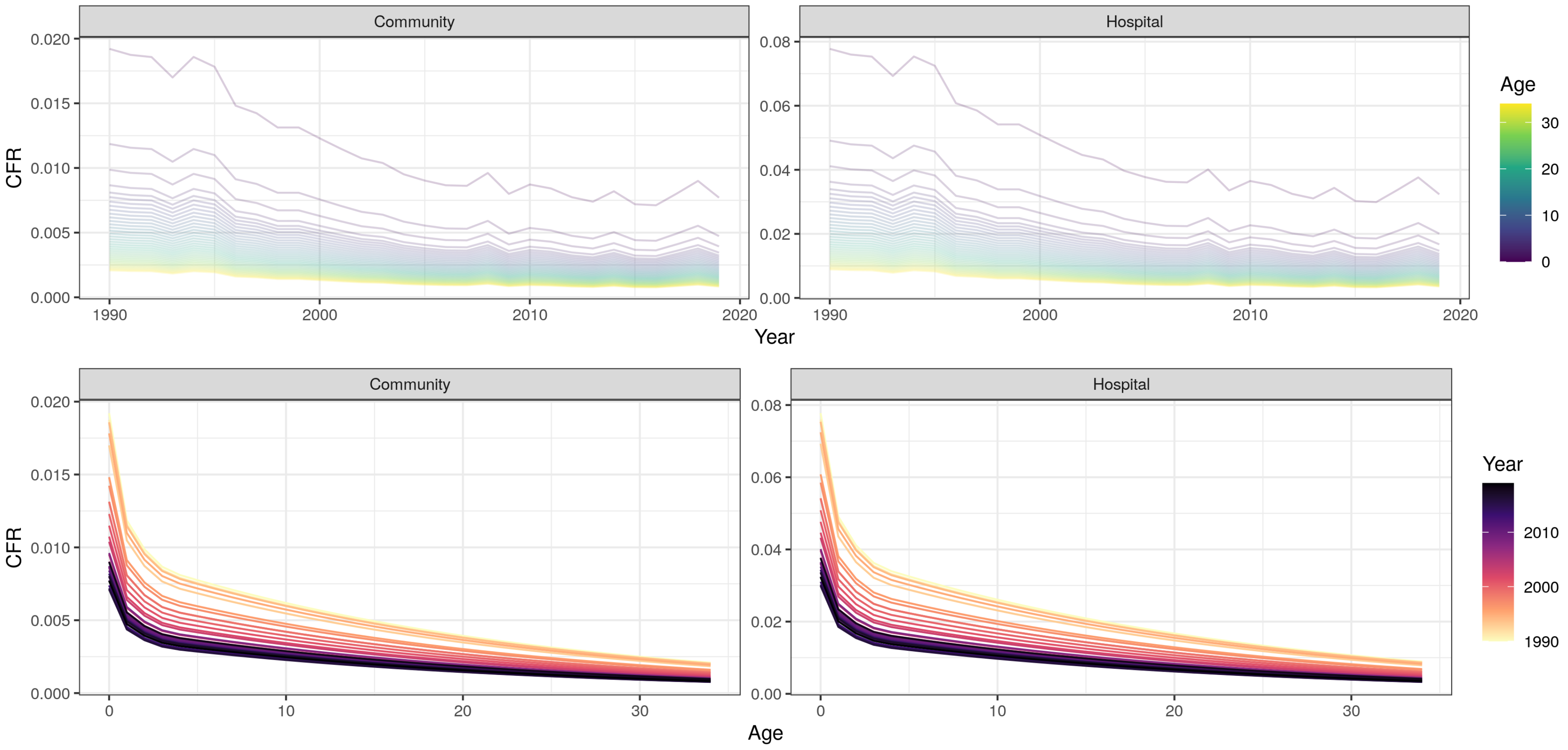

Community
Hospital
CFR
0.020
0.015
0.010
0.005
0.000
0.08
0.06
0.04
0.02
0.00
1990
2000
2010
2020
Year
Age
30
20
10
0
Community
Hospital
0
10
20
30
Age
Year
2010
2000
1990

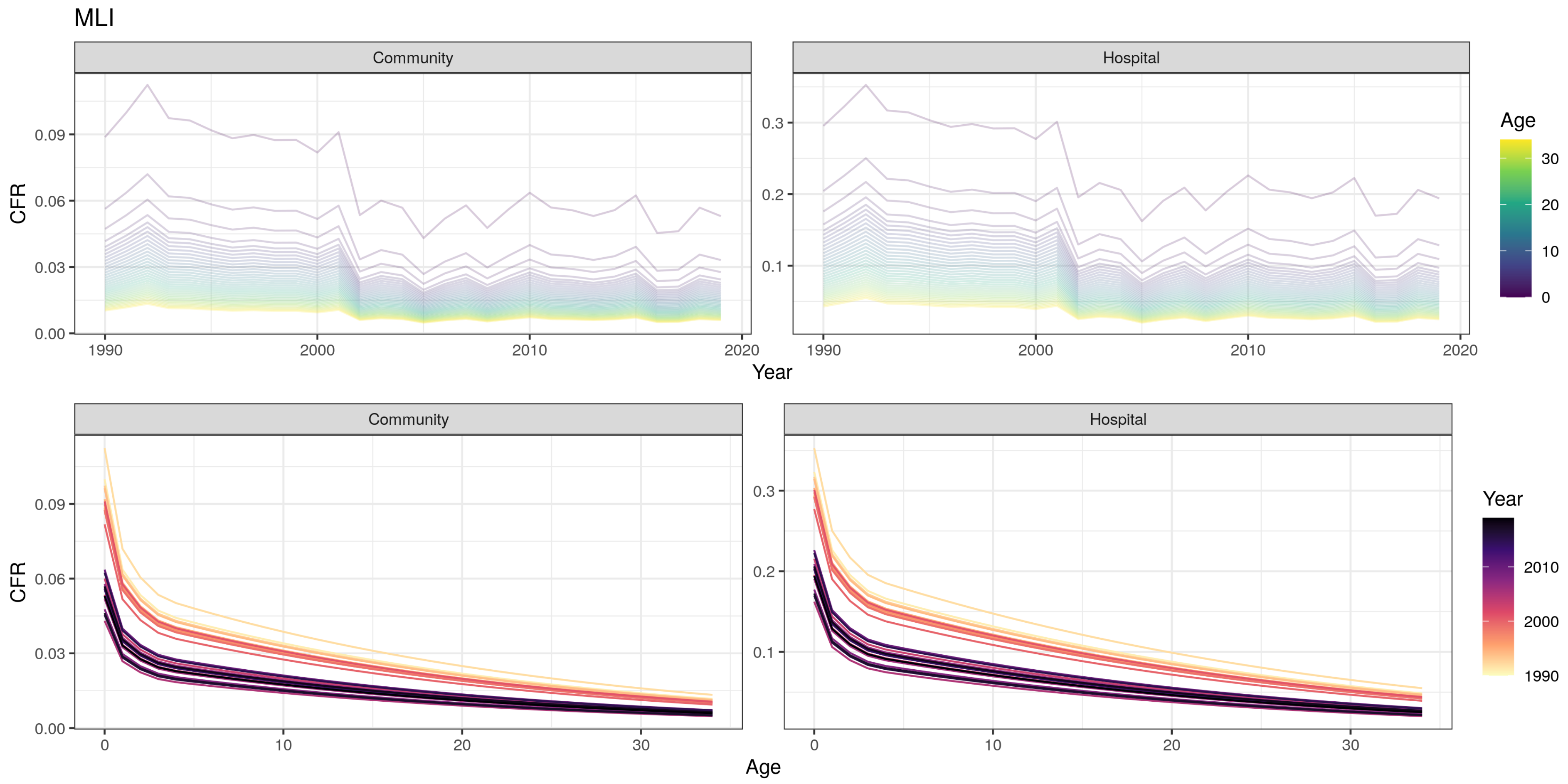
MLI
Community
Hospital
CFR
Year
Age
0.00
0.03
0.06
0.09
0.1
0.2
0.3
1990
2000
2010
2020
0
10
20
30

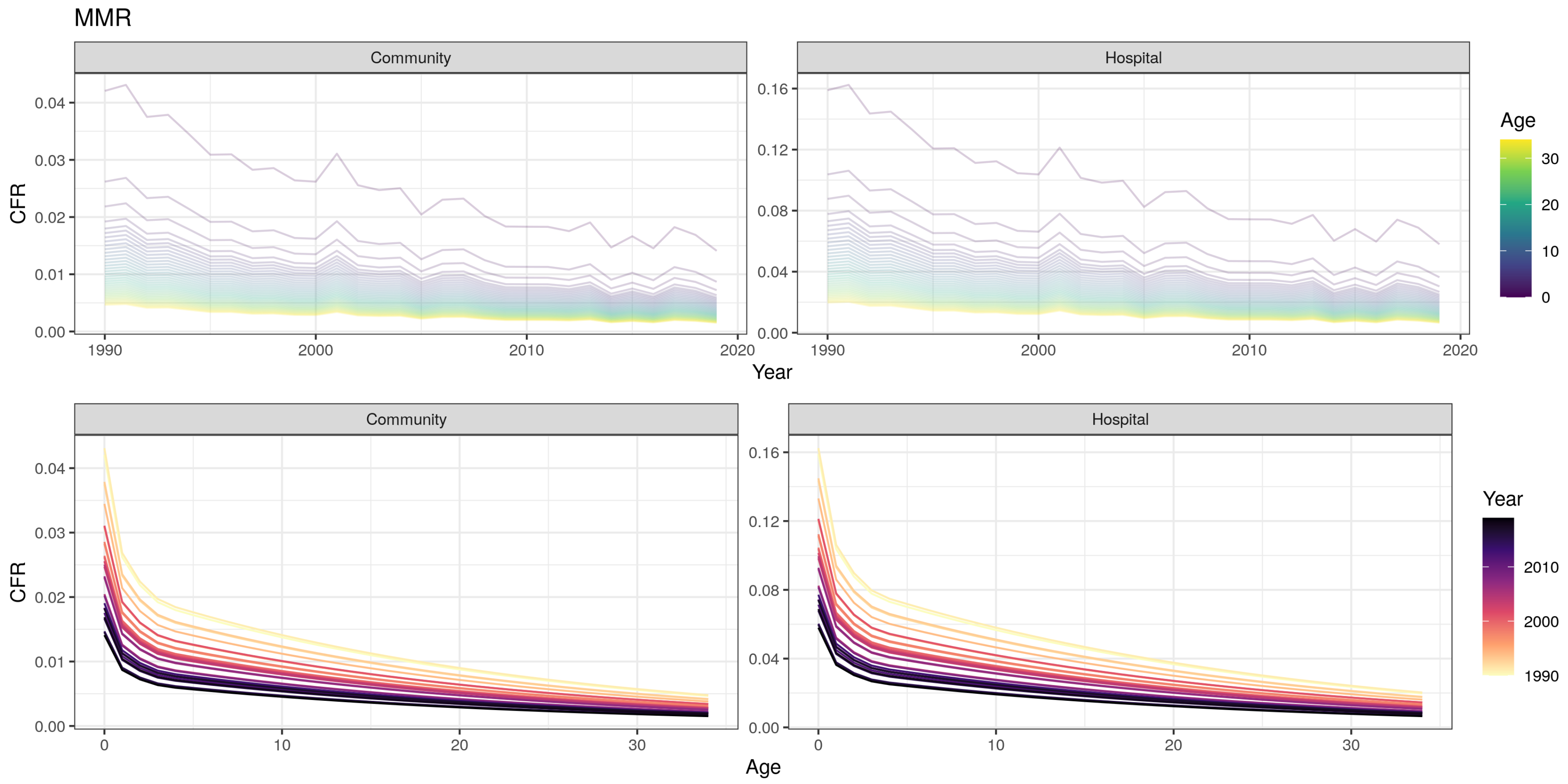
MMR
Community
Hospital
CFR
Year
Age
1990
2000
2010
2020
0.00
0.01
0.02
0.03
0.04
0.04
0.08
0.12
0.16
0
10
20
30

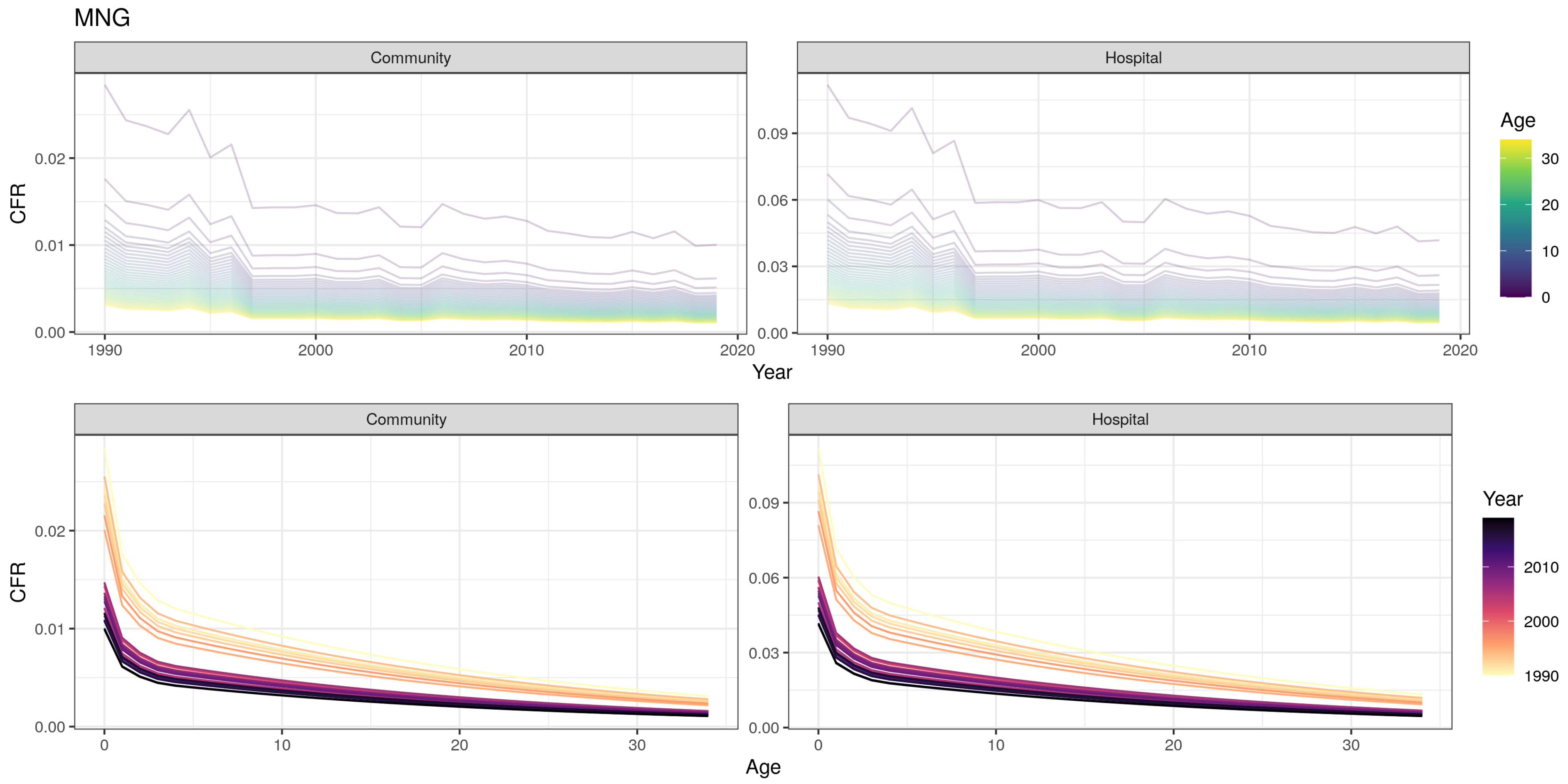

MNG
Community
Hospital
CFR
0.02
0.01
0.00
0.09
0.06
0.03
0.00
1990
2000
2010
2020
Year
Age
30
20
10
0
Community
Hospital
CFR
0.02
0.01
0.00
0.09
0.06
0.03
0.00
0
10
20
30
Age
Year
2010
2000
1990

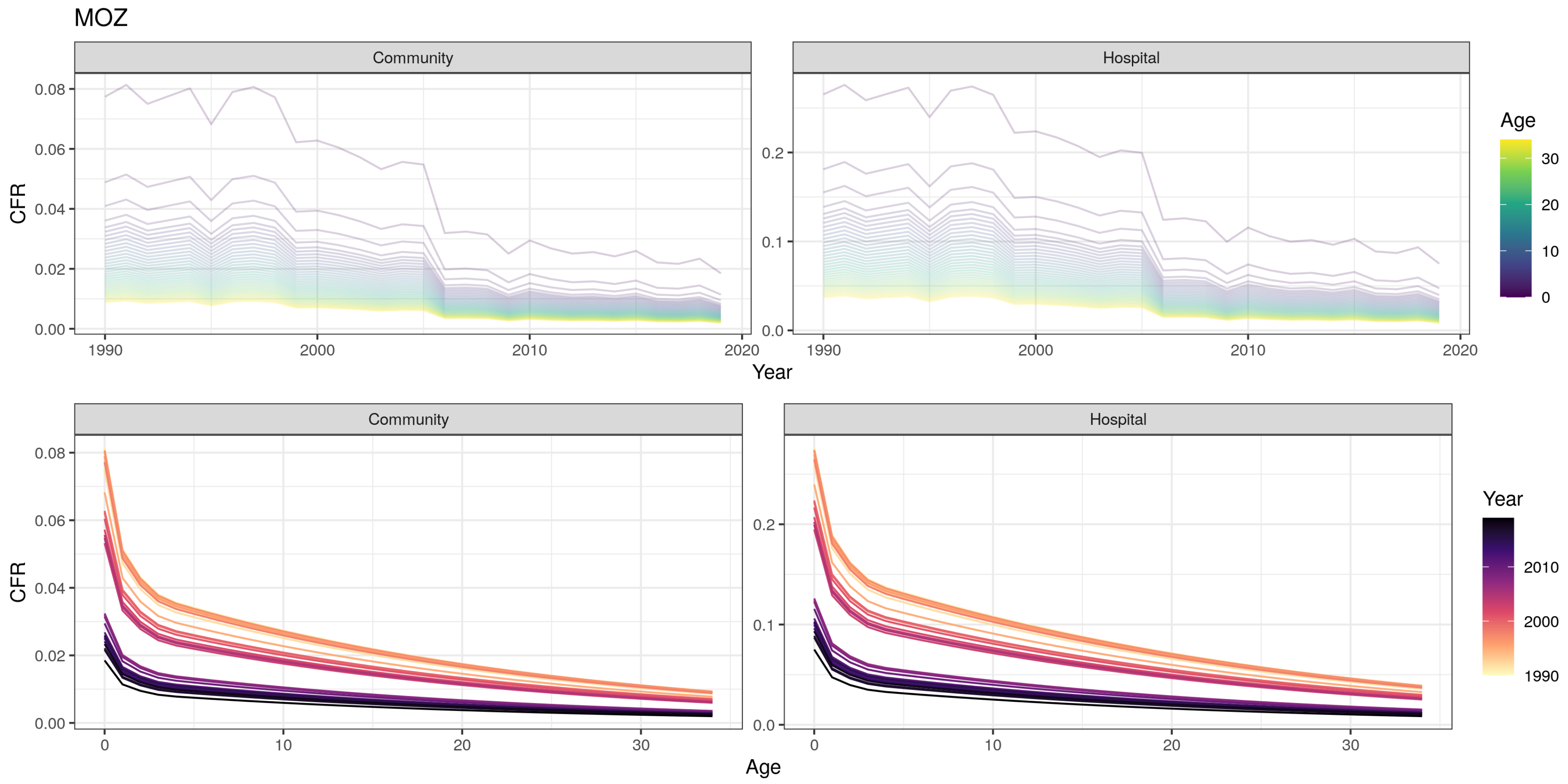
MOZ
Community
Hospital
CFR
Year
Age
Community
Hospital
CFR
Age
Year

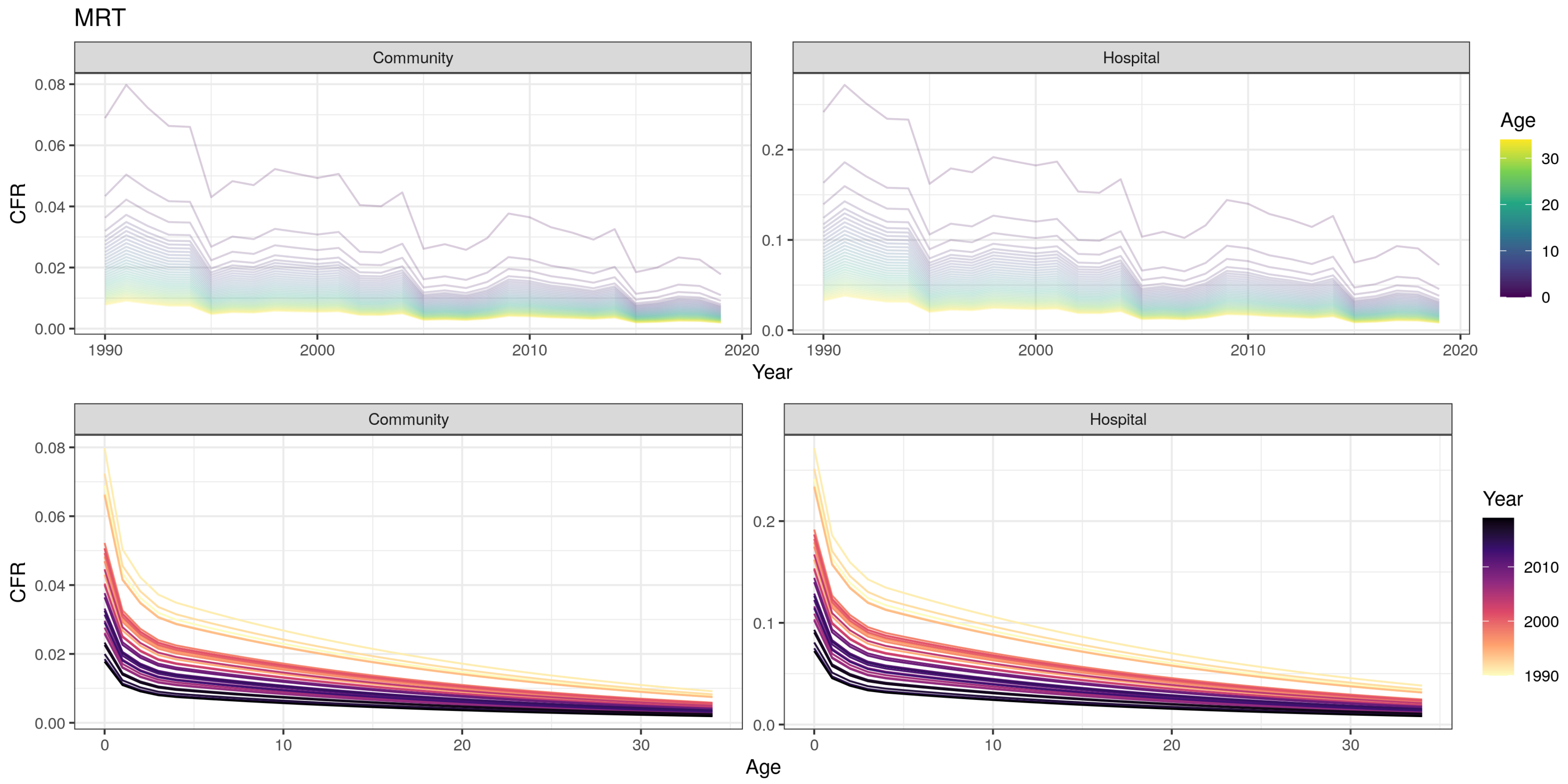
MRT
Community
Hospital
CFR
Year
Age
0.08
0.06
0.04
0.02
0.00
0.2
0.1
0.0
1990
2000
2010
2020
30
20
10
0
Community
Hospital
Year
2010
2000
1990

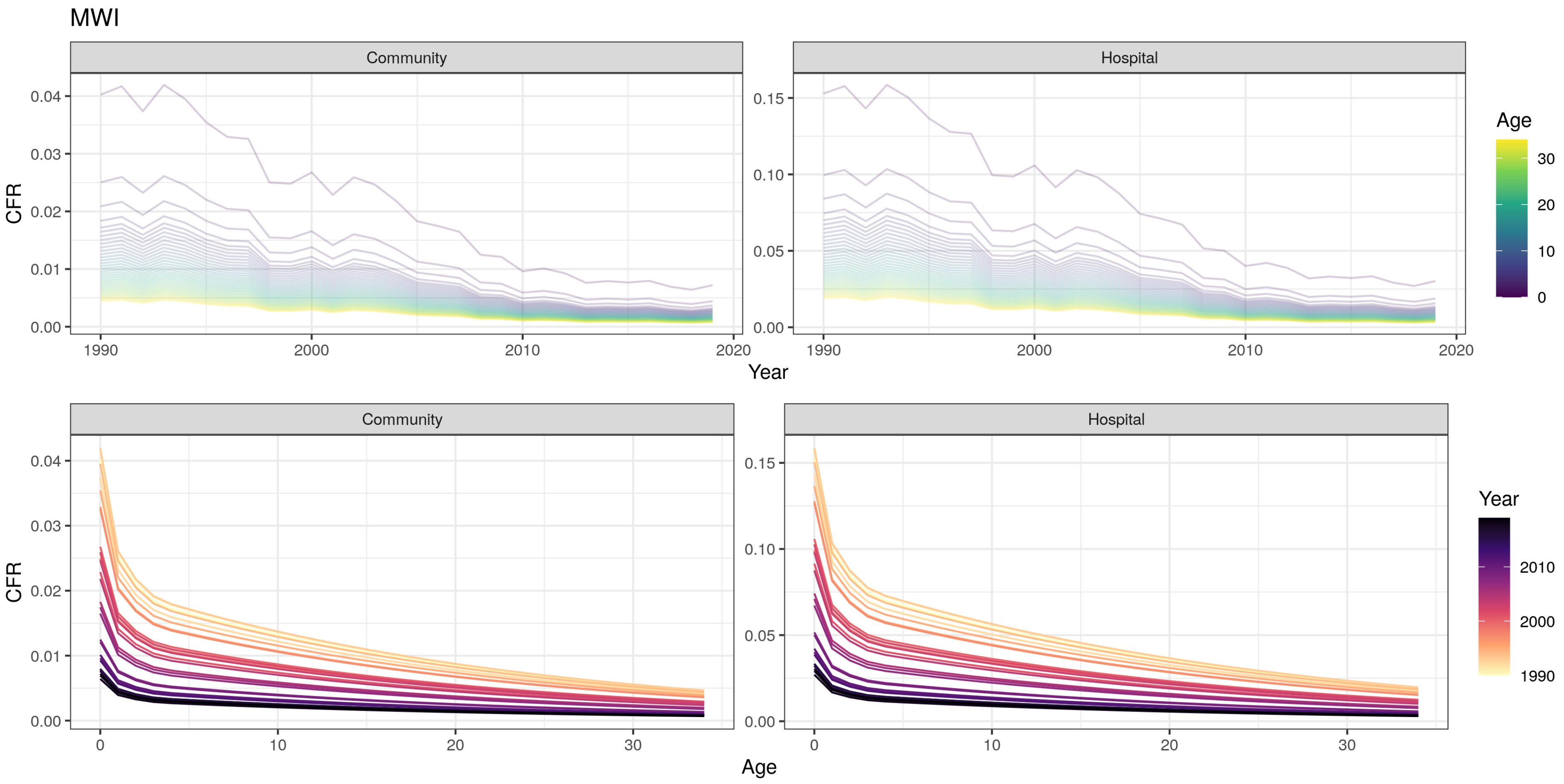
MWI
Community
Hospital
CFR
Year
Age
0.00
0.01
0.02
0.03
0.04
0.05
0.10
0.15
1990
2000
2010
2020
0
10
20
30

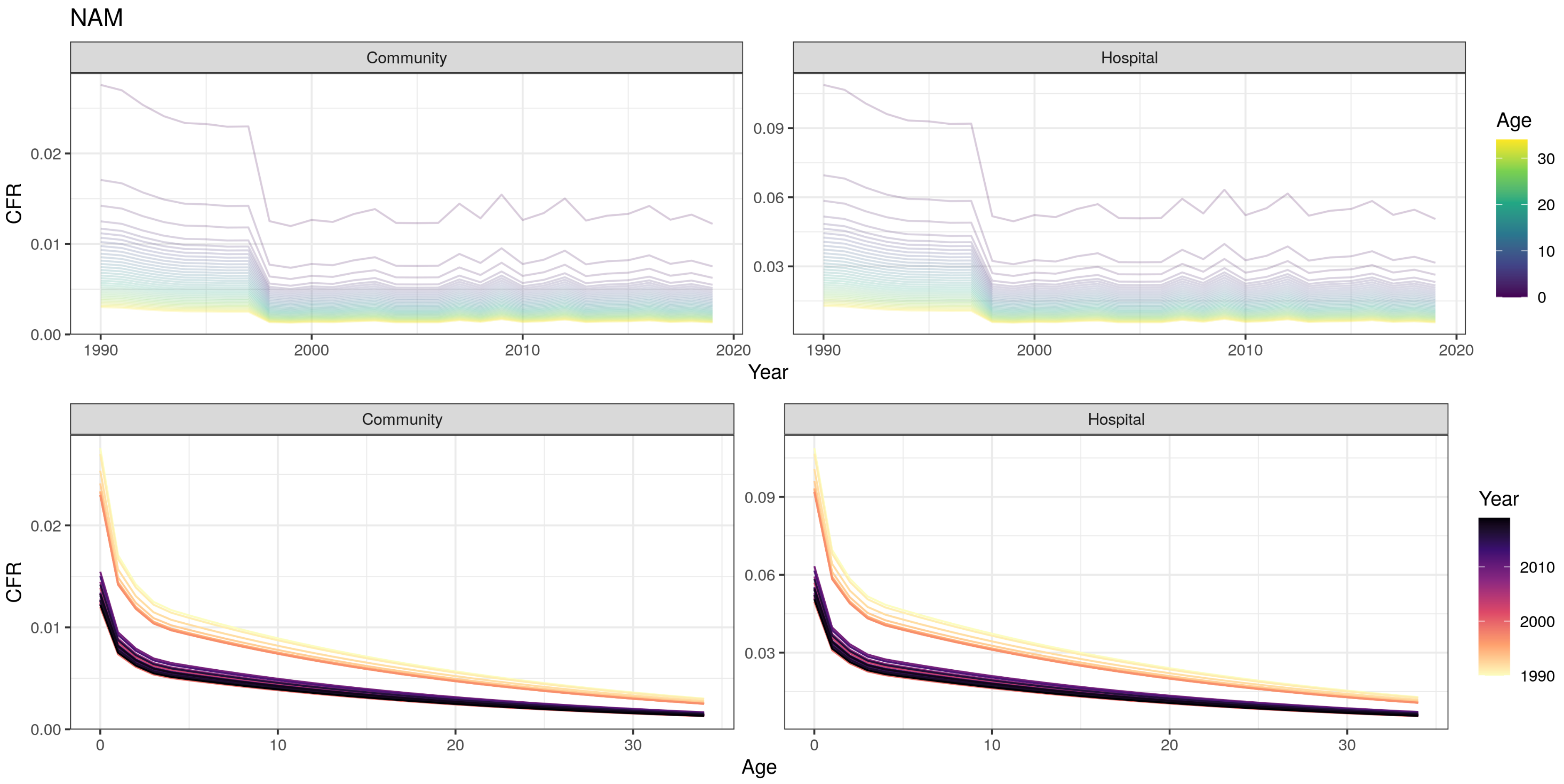
NAM
Community
Hospital
CFR
Year
Age
0.00
0.01
0.02
0.03
0.06
0.09
1990
2000
2010
2020
0
10
20
30
Year
2010
2000
1990

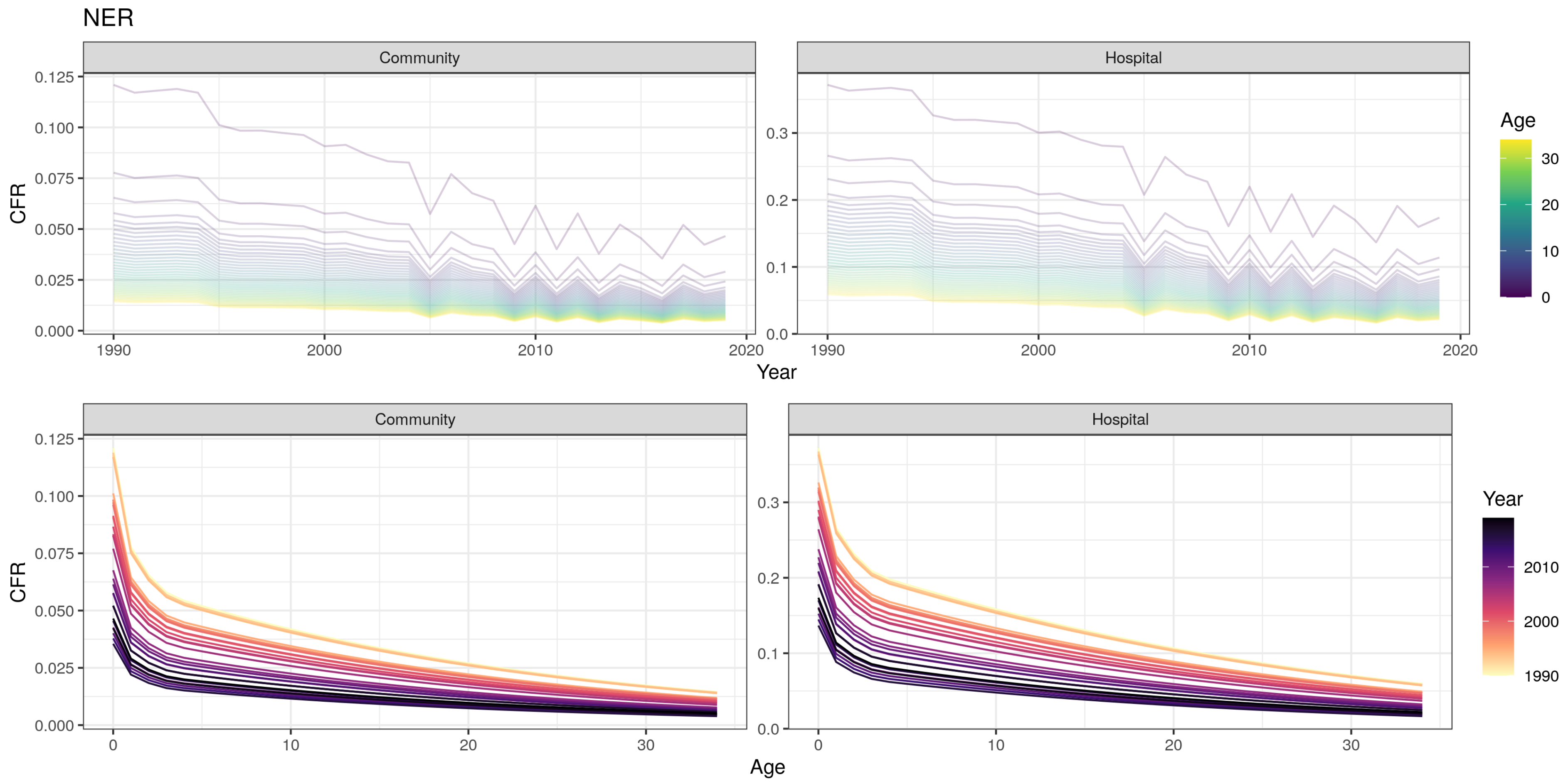
NER
Community
Hospital
CFR
Year
Age
0.125
0.100
0.075
0.050
0.025
0.000
0.3
0.2
0.1
0.0
1990
2000
2010
2020
0
10
20
30
Age
Year
1990
2000
2010

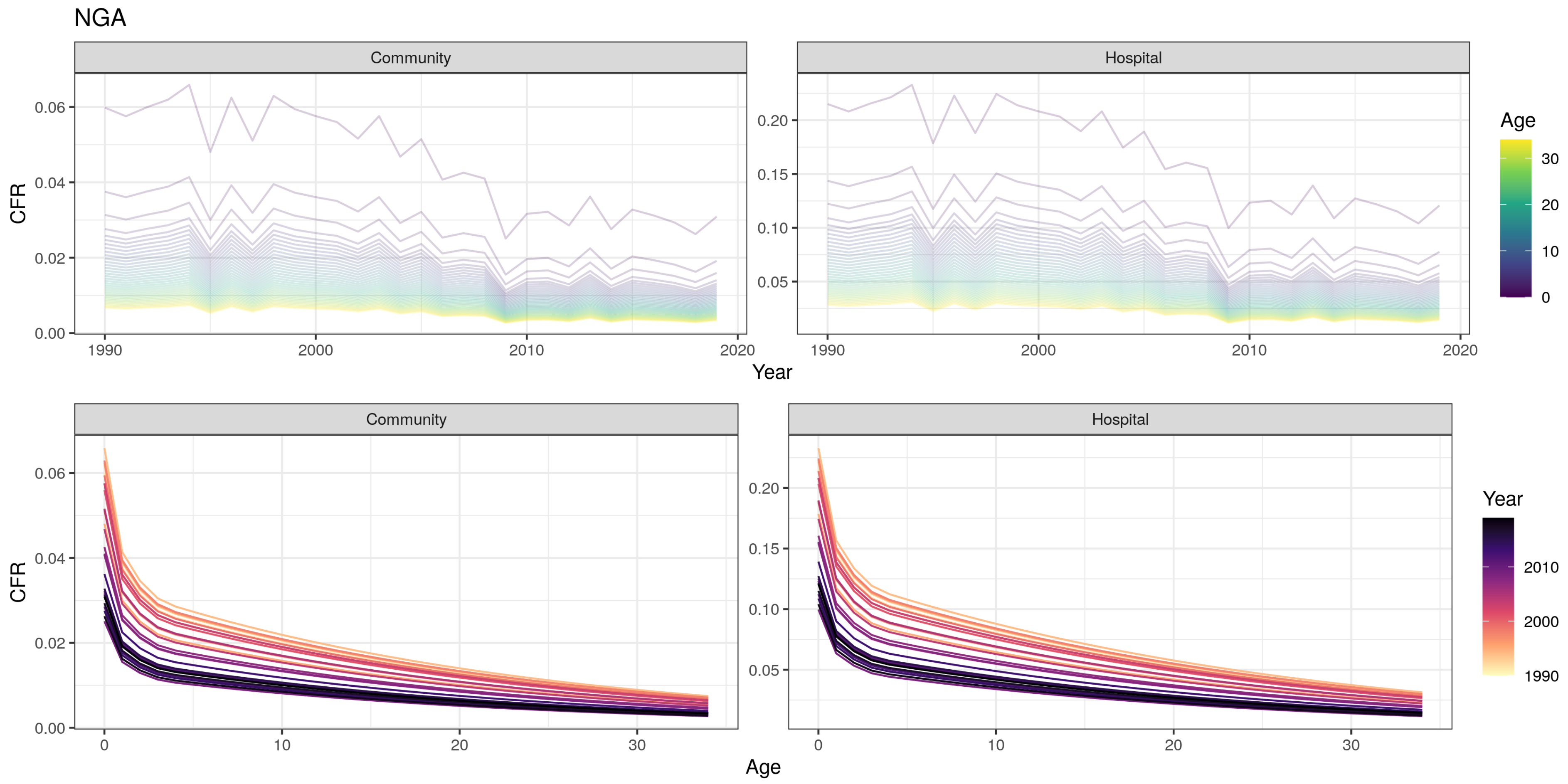

NGA
Community
Hospital
CFR
Year
Age
Community
Hospital
CFR
Age
Year

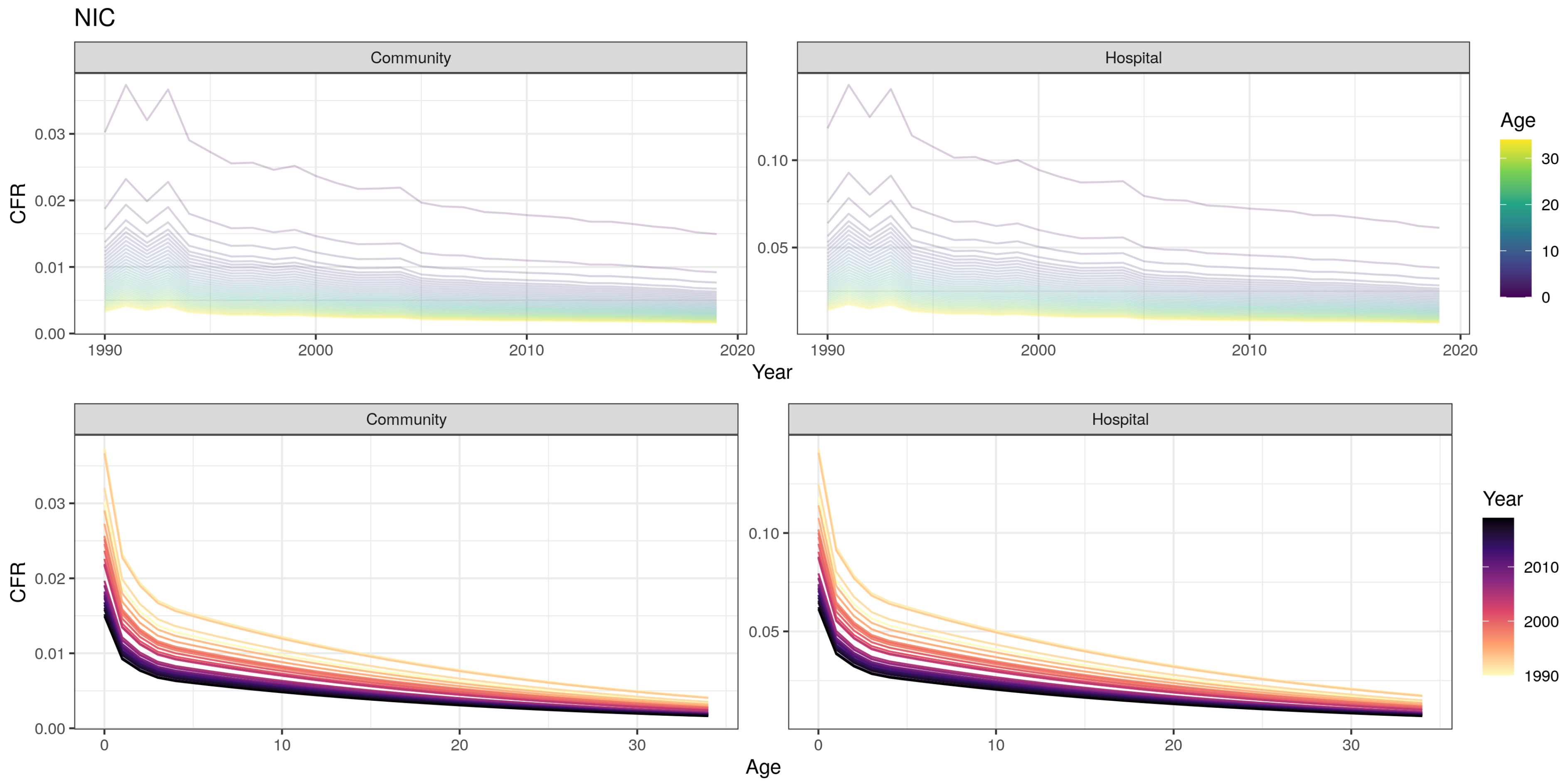

NIC
Community
Hospital
CFR
0.03
0.02
0.01
0.00
0.10
0.05
1990
2000
2010
2020
Year
Age
30
20
10
0
Community
Hospital
CFR
0.03
0.02
0.01
0.00
0.10
0.05
0
10
20
30
Age
Year
2010
2000
1990

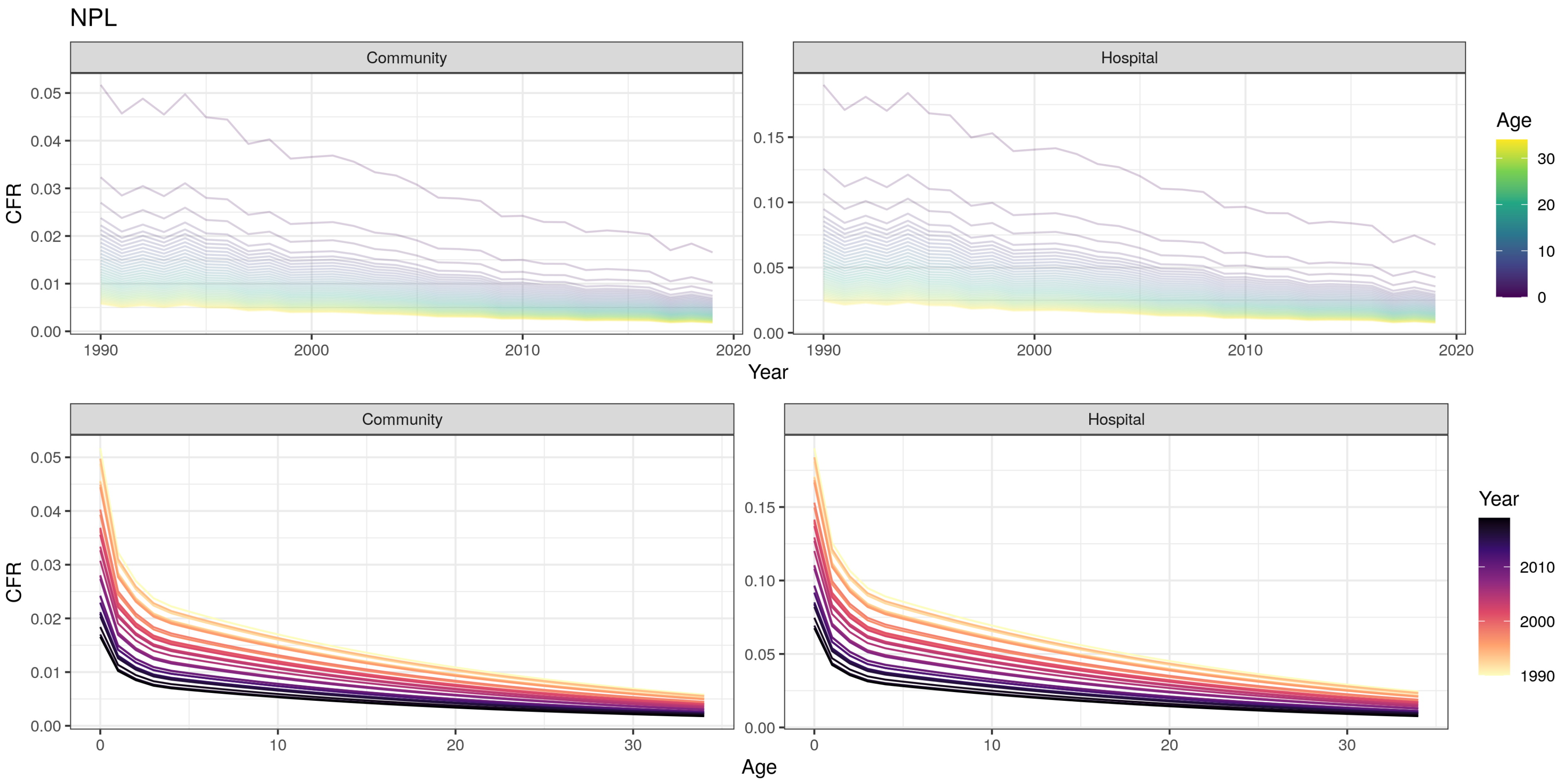
NPL
Community
Hospital
CFR
Year
Age
Community
Hospital
CFR
Age
Year
0.05
0.04
0.03
0.02
0.01
0.00
0.15
0.10
0.05
0.00
1990
2000
2010
2020
0
10
20
30

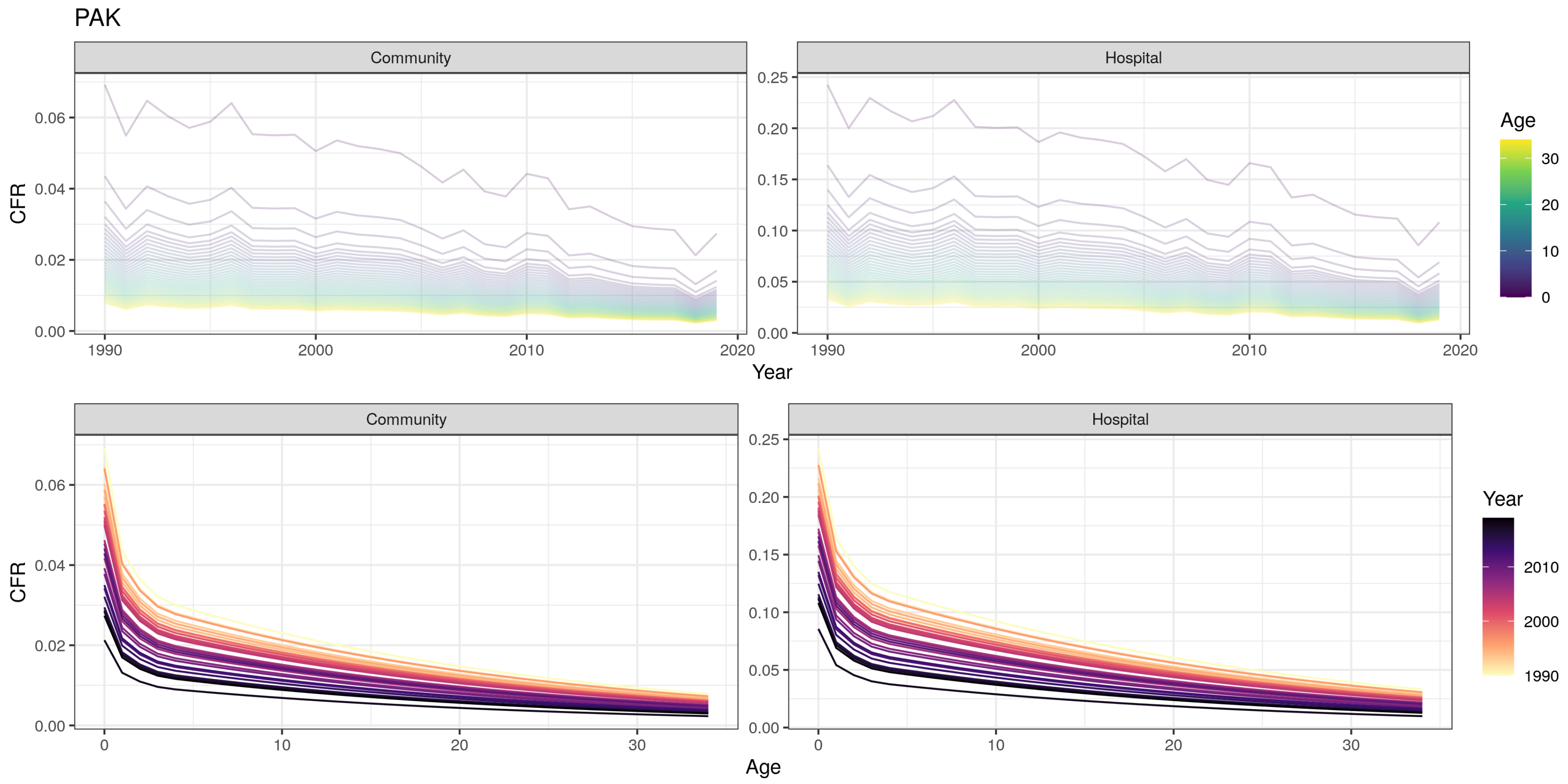

PAK
Community
Hospital
CFR
Year
Age
0.00
0.02
0.04
0.06
0.05
0.10
0.15
0.20
0.25
1990
2000
2010
2020
0
10
20
30

PER

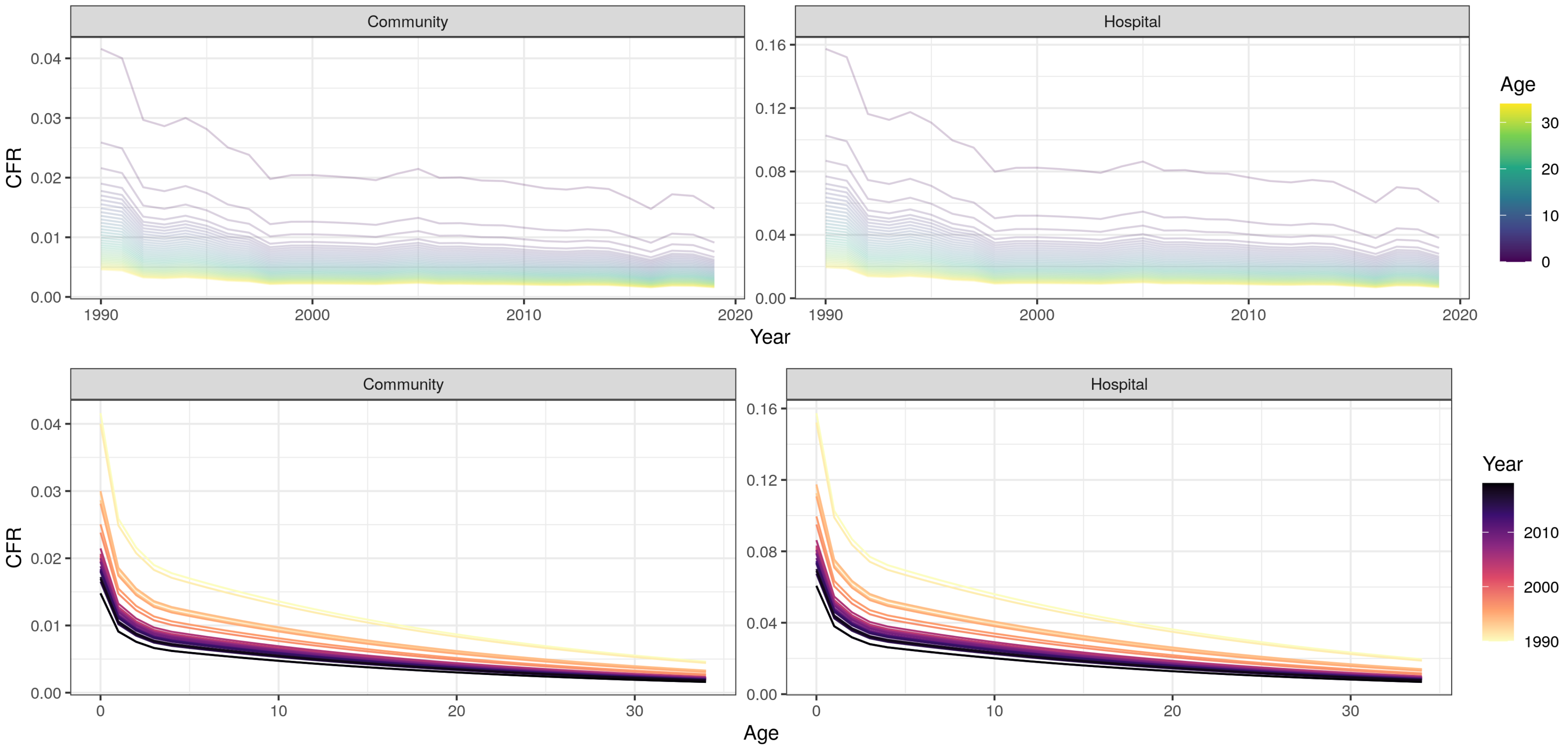

Community
Hospital
CFR
Year
Age
0.00
0.01
0.02
0.03
0.04
0.00
0.04
0.08
0.12
0.16
1990
2000
2010
2020
0
10
20
30
Community
Hospital
CFR
Age
Year
1990
2000
2010

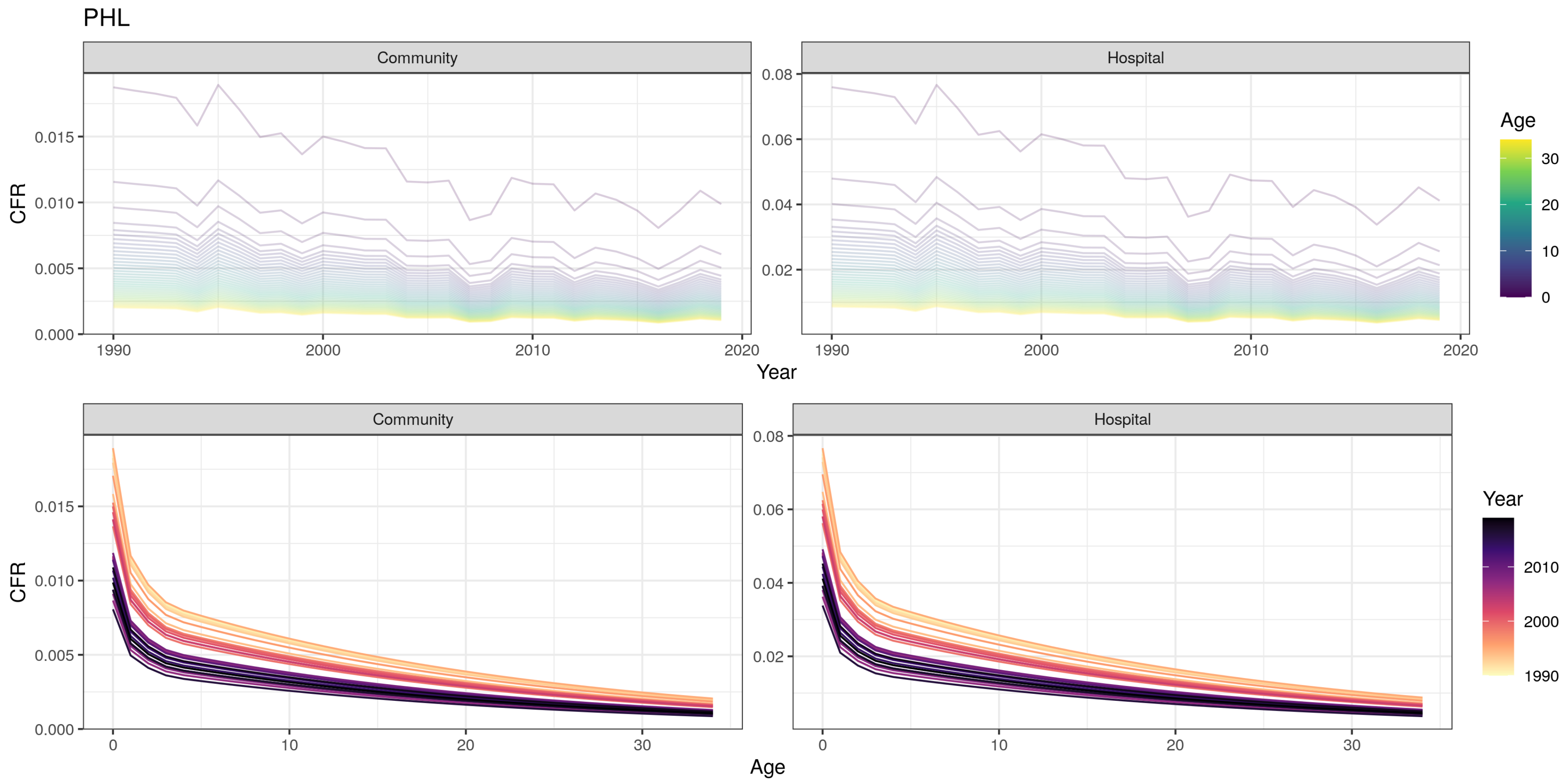
PHL
Community
Hospital
CFR
Year
Age
0.000
0.005
0.010
0.015
0.02
0.04
0.06
0.08
1990
2000
2010
2020
0
10
20
30
Age
Year
1990
2000
2010

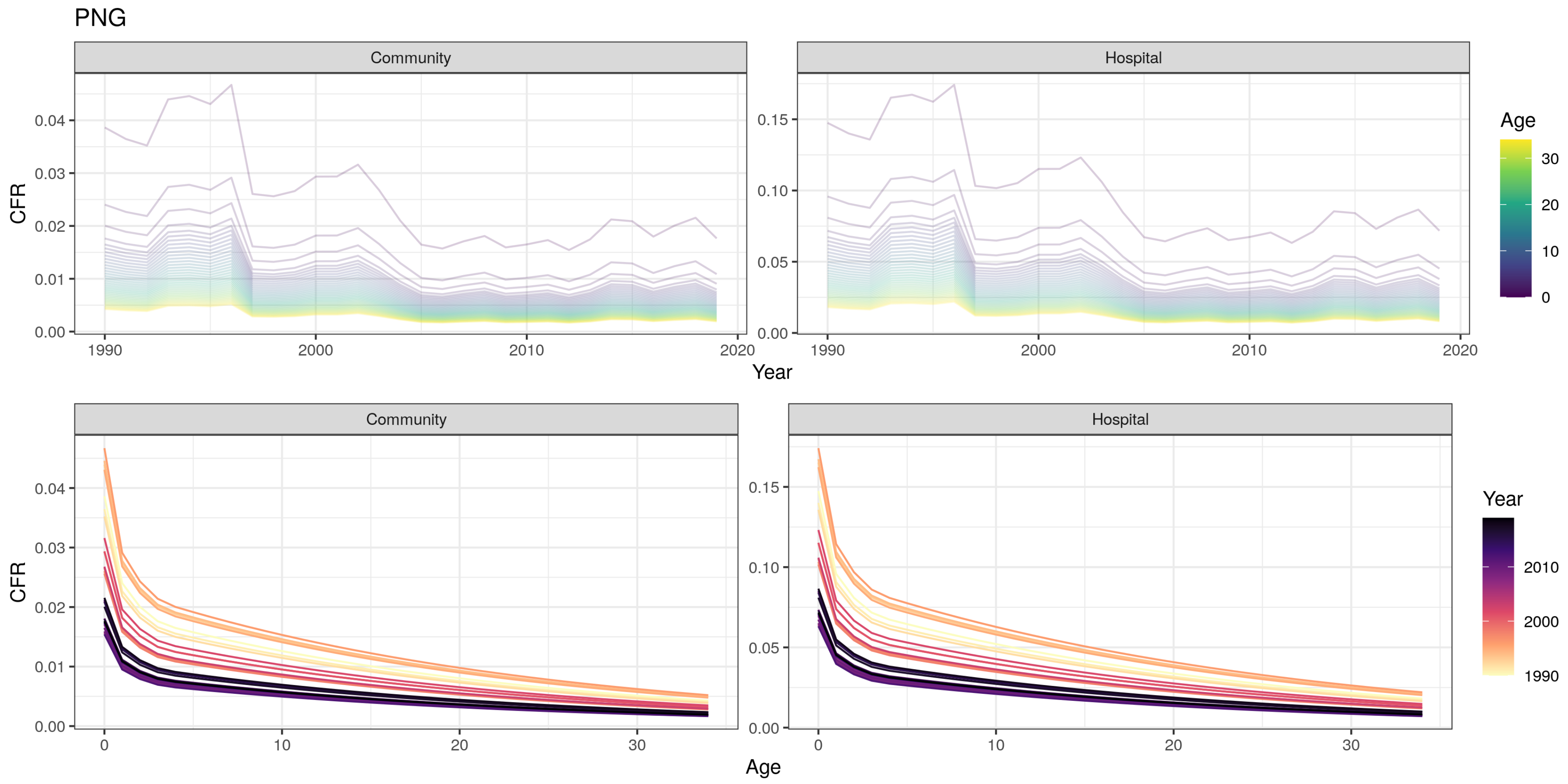
PNG
Community
Hospital
CFR
Year
Age
Community
Hospital
CFR
Age
Year
0.00
0.01
0.02
0.03
0.04
0.05
0.10
0.15
1990
2000
2010
2020
0
10
20
30

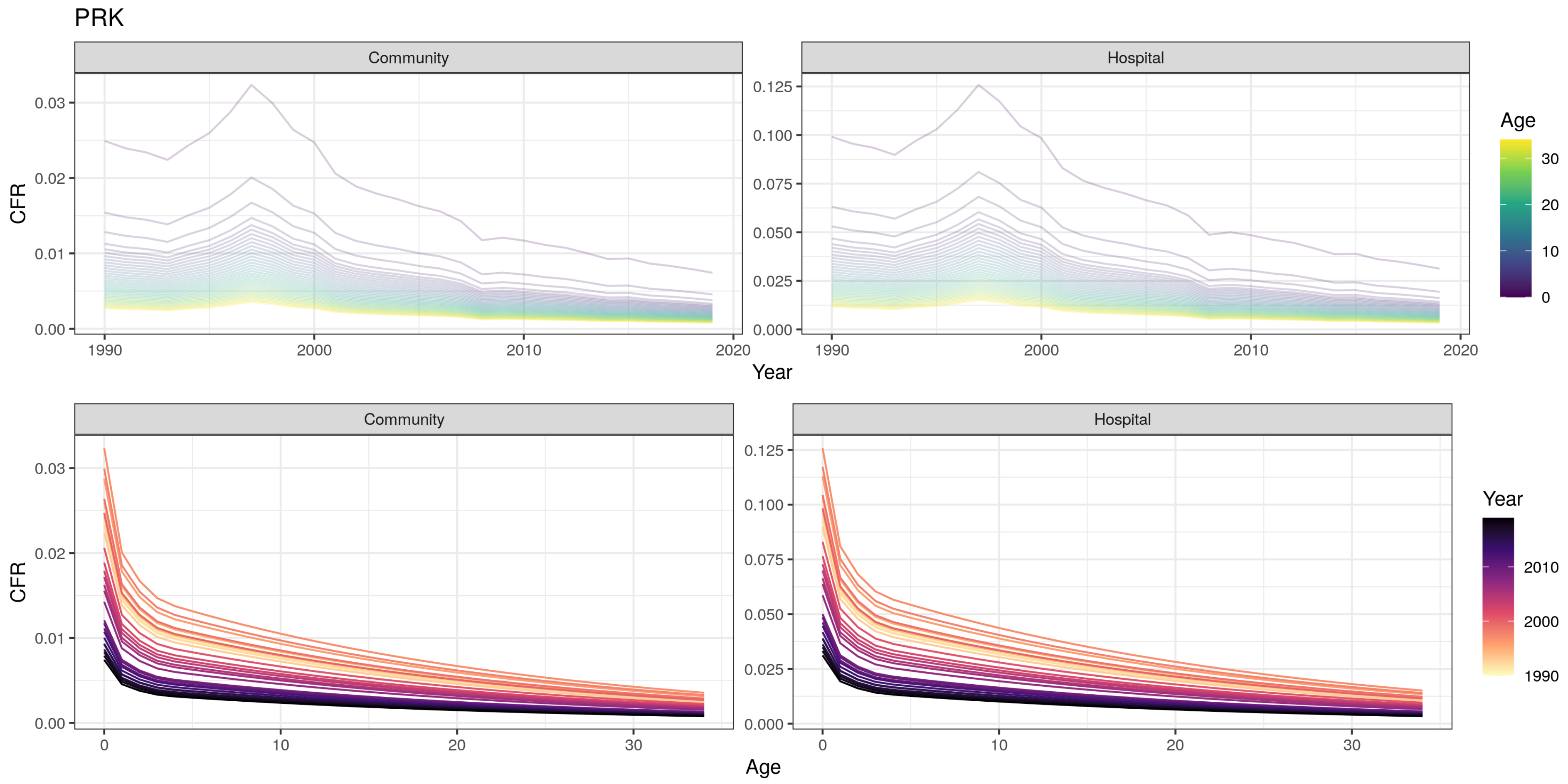
PRK
Community
Hospital
CFR
Year
Age
Community
Hospital
CFR
Age
Year
0.00
0.01
0.02
0.03
0.000
0.025
0.050
0.075
0.100
0.125
1990
2000
2010
2020
0
10
20
30

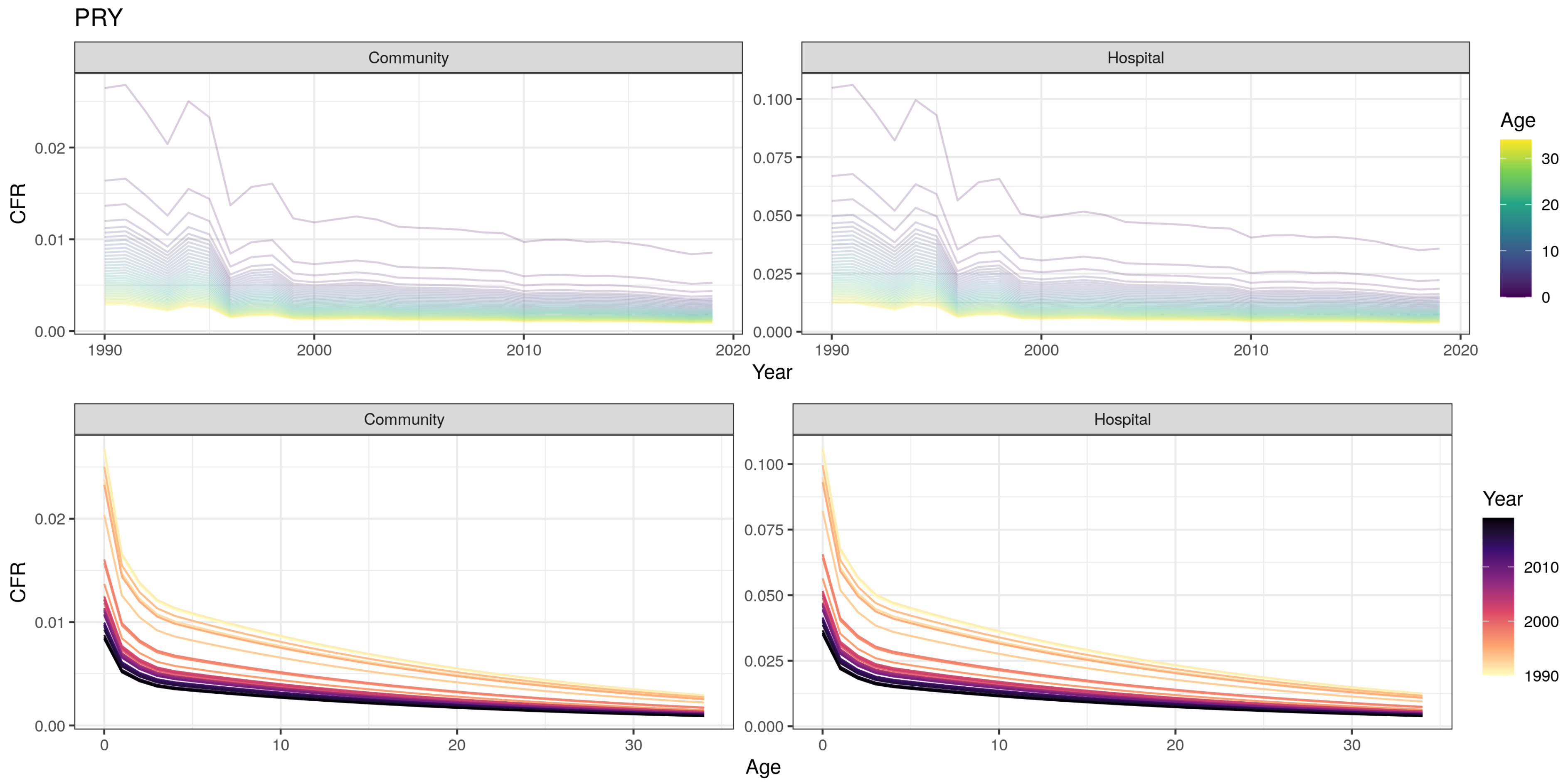

PRY
Community
Hospital
CFR
Year
Age
0.00
0.01
0.02
0.000
0.025
0.050
0.075
0.100
1990
2000
2010
2020
0
10
20
30
Year
2010
2000
1990

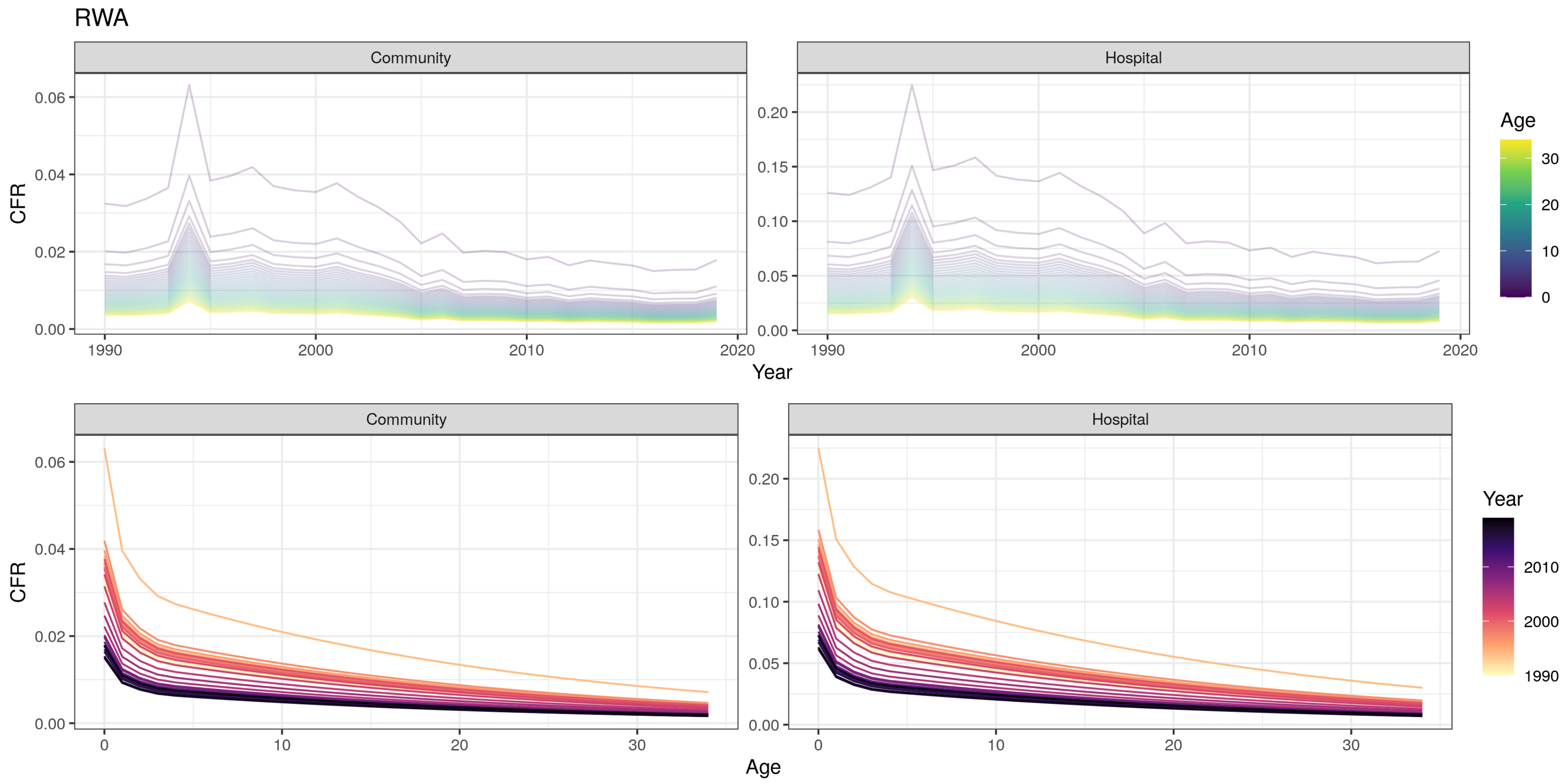

RWA
Community
Hospital
CFR
Year
Age
Community
Hospital
CFR
Age
Year

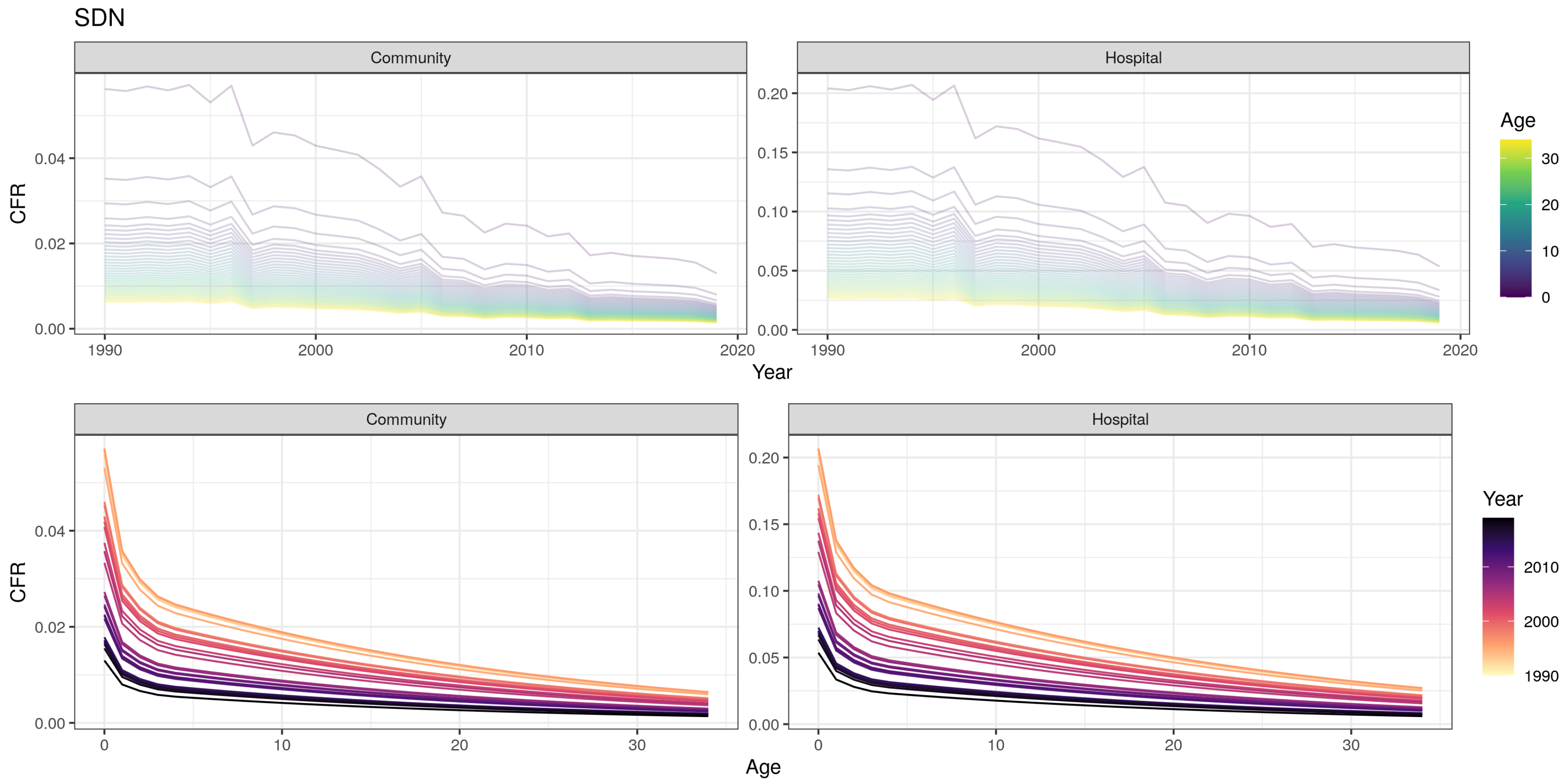

SDN
Community
Hospital
CFR
Year
Age
0.00
0.02
0.04
0.05
0.10
0.15
0.20
1990
2000
2010
2020
0
10
20
30

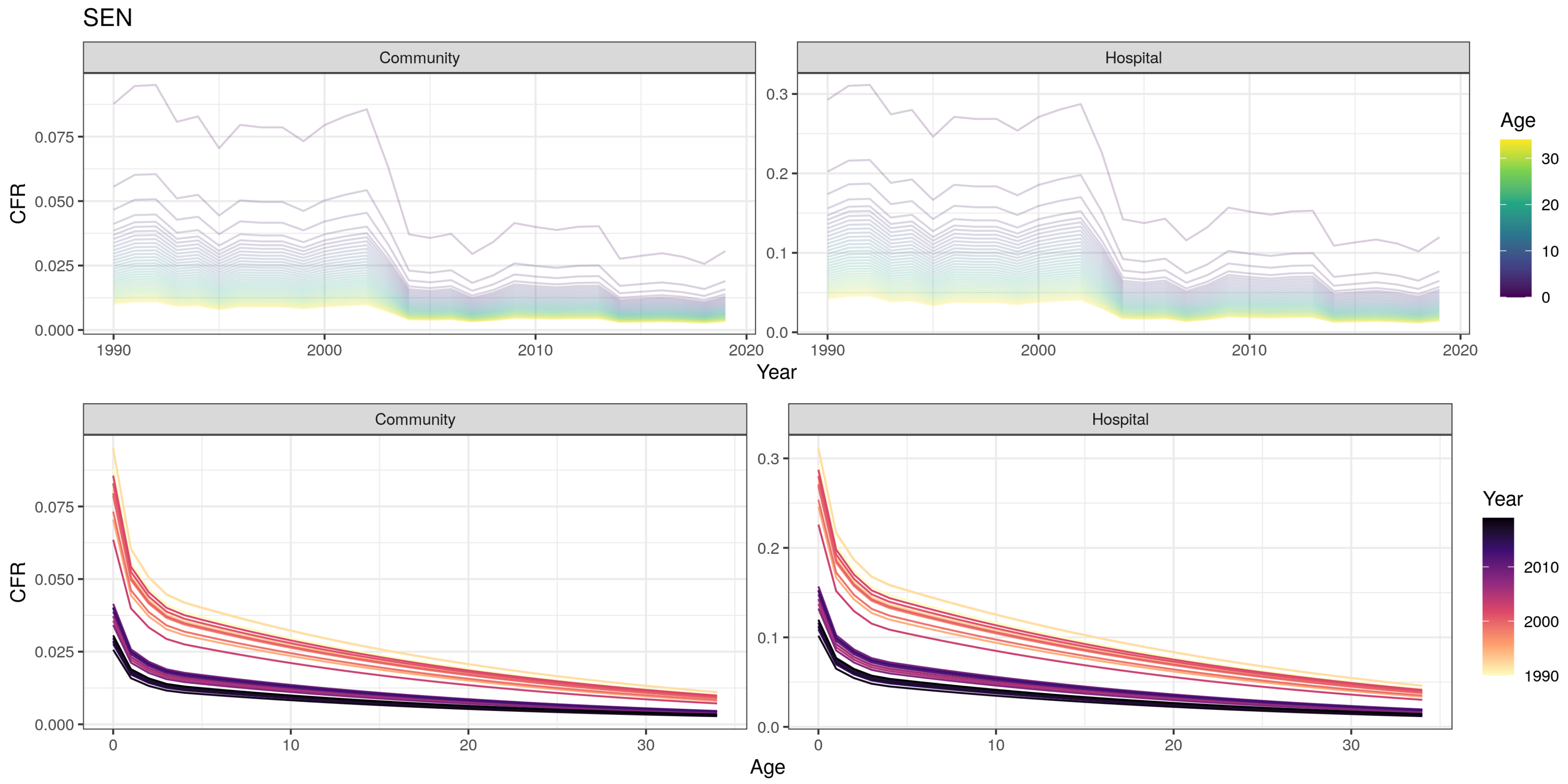
SEN
Community
Hospital
CFR
Year
Age
Community
Hospital
CFR
Age
Year

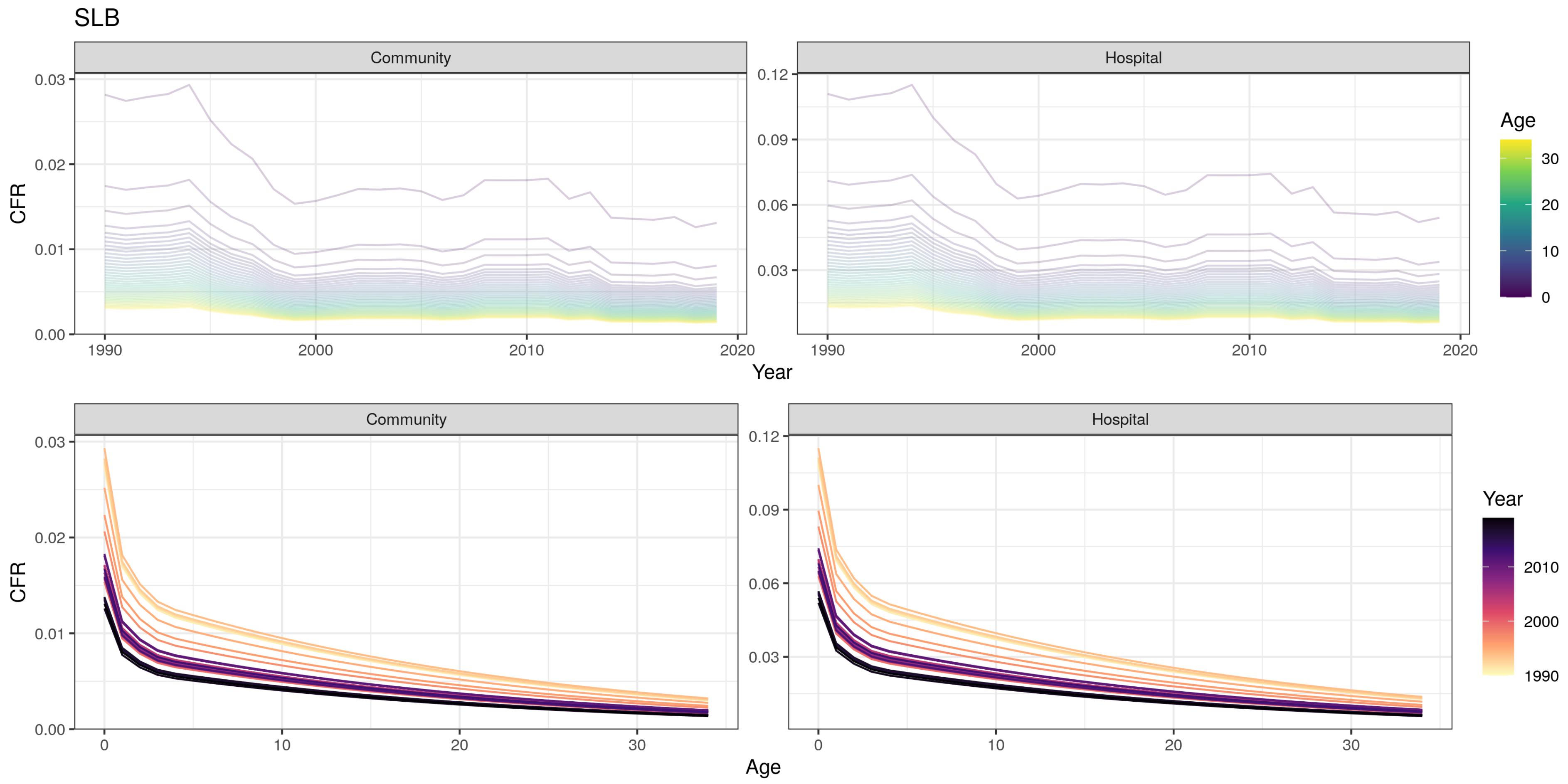
SLB
Community
Hospital
CFR
Year
Age
1990
2000
2010
2020
0.00
0.01
0.02
0.03
0.03
0.06
0.09
0.12
0
10
20
30
Age
Year
1990
2000
2010

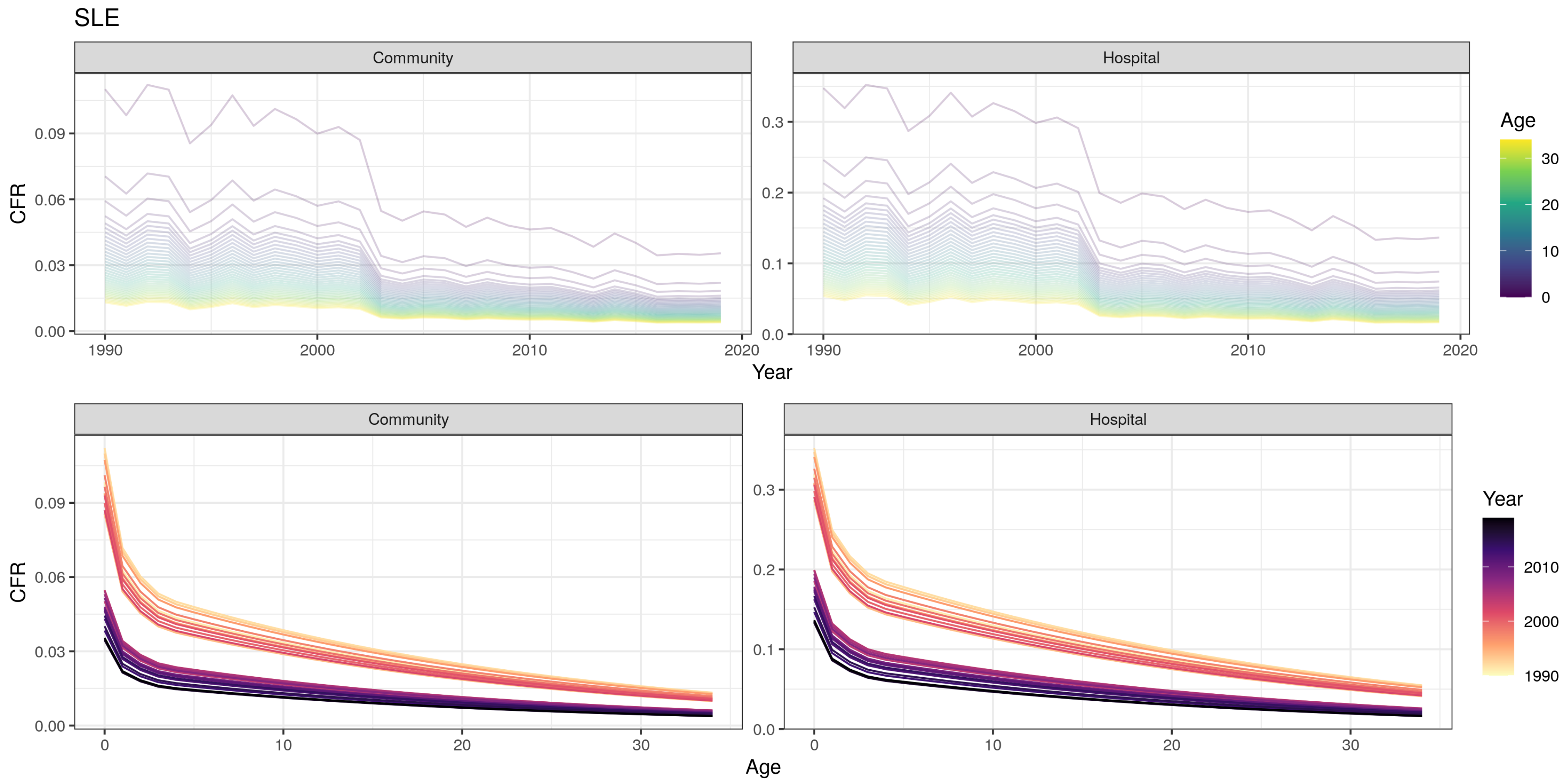

SLE
Community
Hospital
CFR
Year
Age
0.00
0.03
0.06
0.09
0.0
0.1
0.2
0.3
1990
2000
2010
2020
0
10
20
30

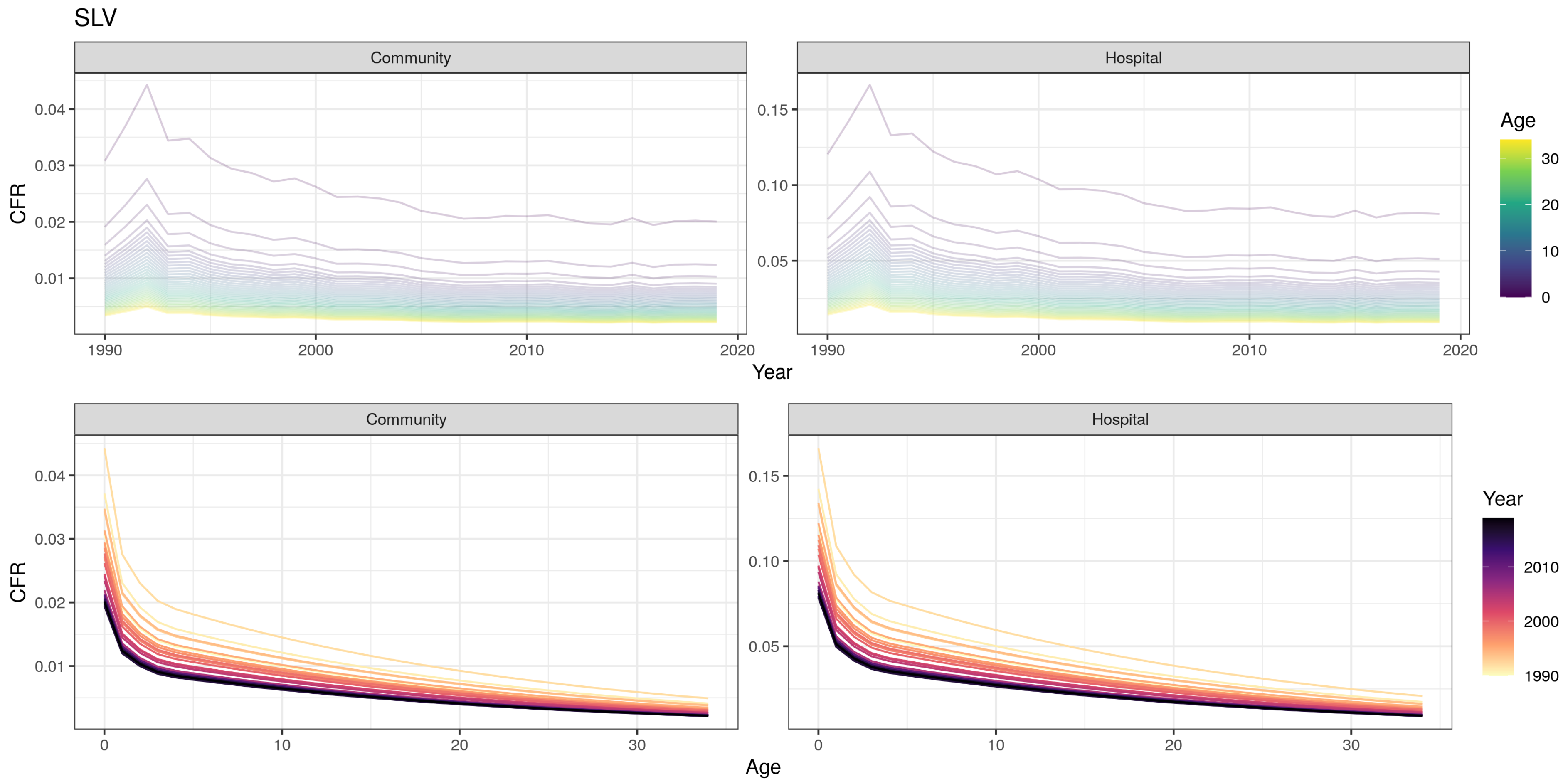

SLV
Community
Hospital
CFR
Year
Age
Community
Hospital
CFR
Age
Year

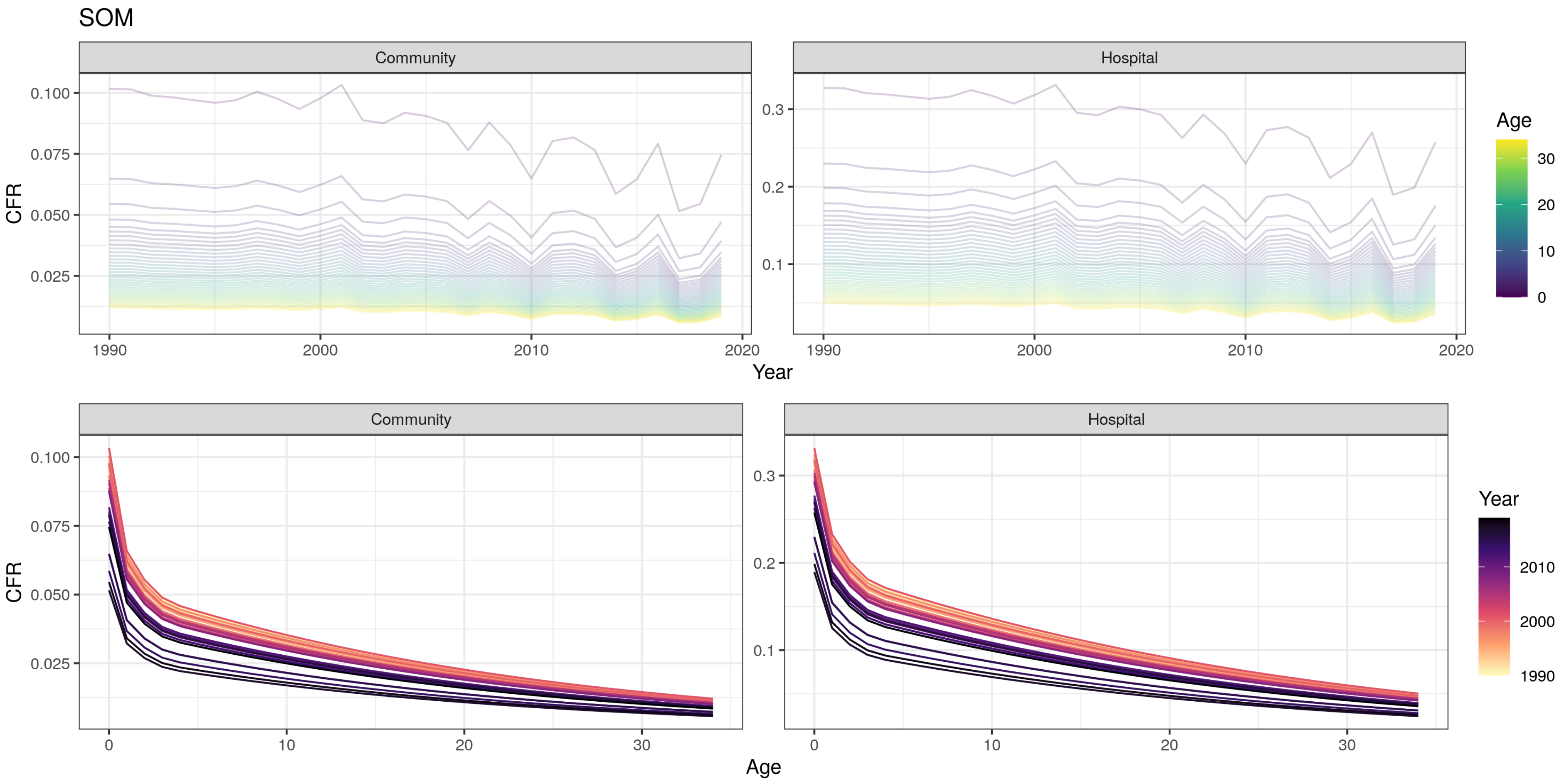

SOM
Community
Hospital
CFR
0.100
0.075
0.050
0.025
0.3
0.2
0.1
1990
2000
2010
2020
Year
Age
30
20
10
0
Community
Hospital
CFR
0.100
0.075
0.050
0.025
0.3
0.2
0.1
0
10
20
30
Age
Year
2010
2000
1990

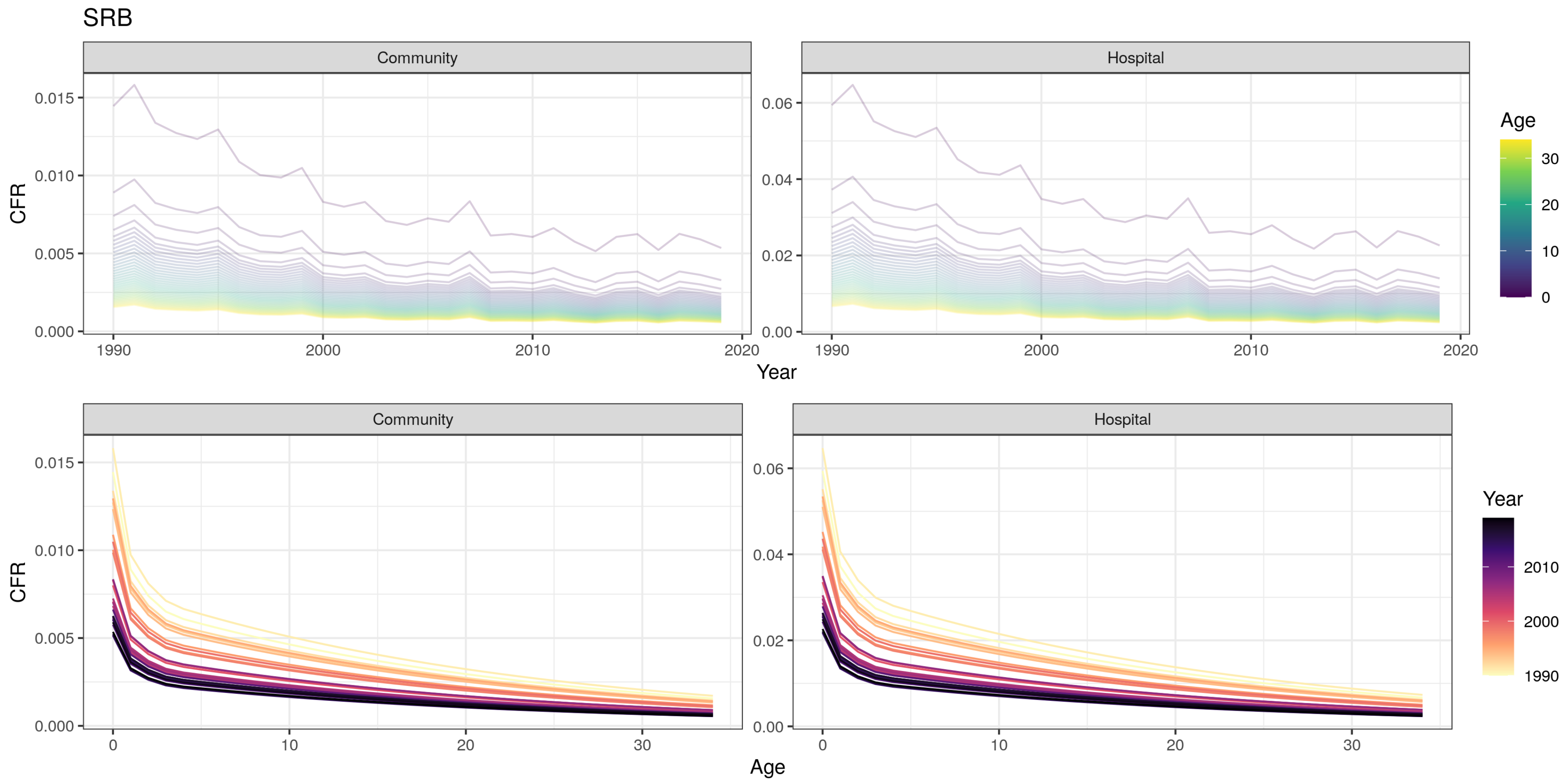
SRB
Community
Hospital
CFR
Year
Age
0.015
0.010
0.005
0.000
0.06
0.04
0.02
0.00
1990
2000
2010
2020
30
20
10
0
Year
2010
2000
1990

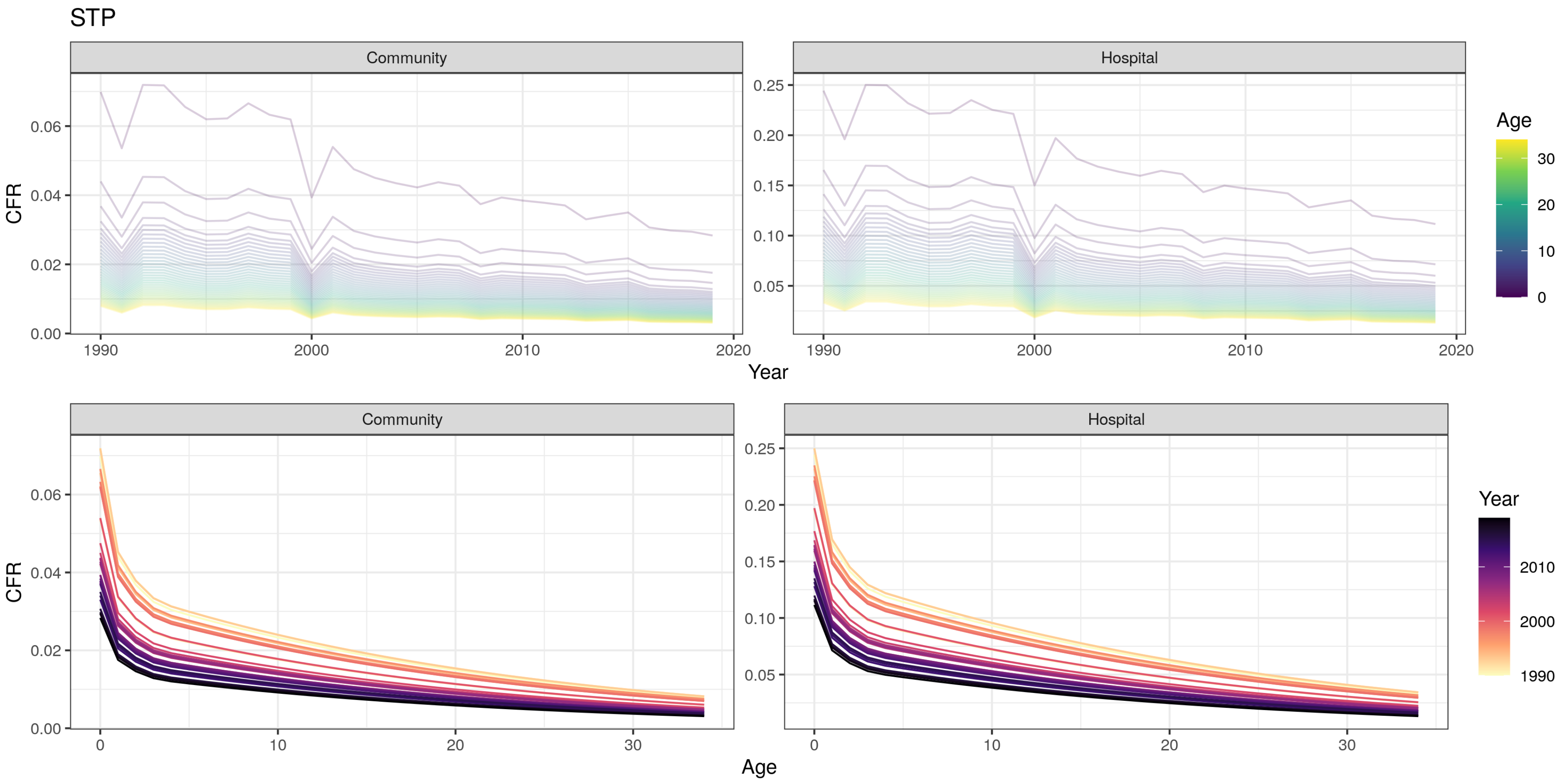

STP
Community
Hospital
CFR
Year
Age
0.00
0.02
0.04
0.06
0.05
0.10
0.15
0.20
0.25
1990
2000
2010
2020
0
10
20
30
Age
Year

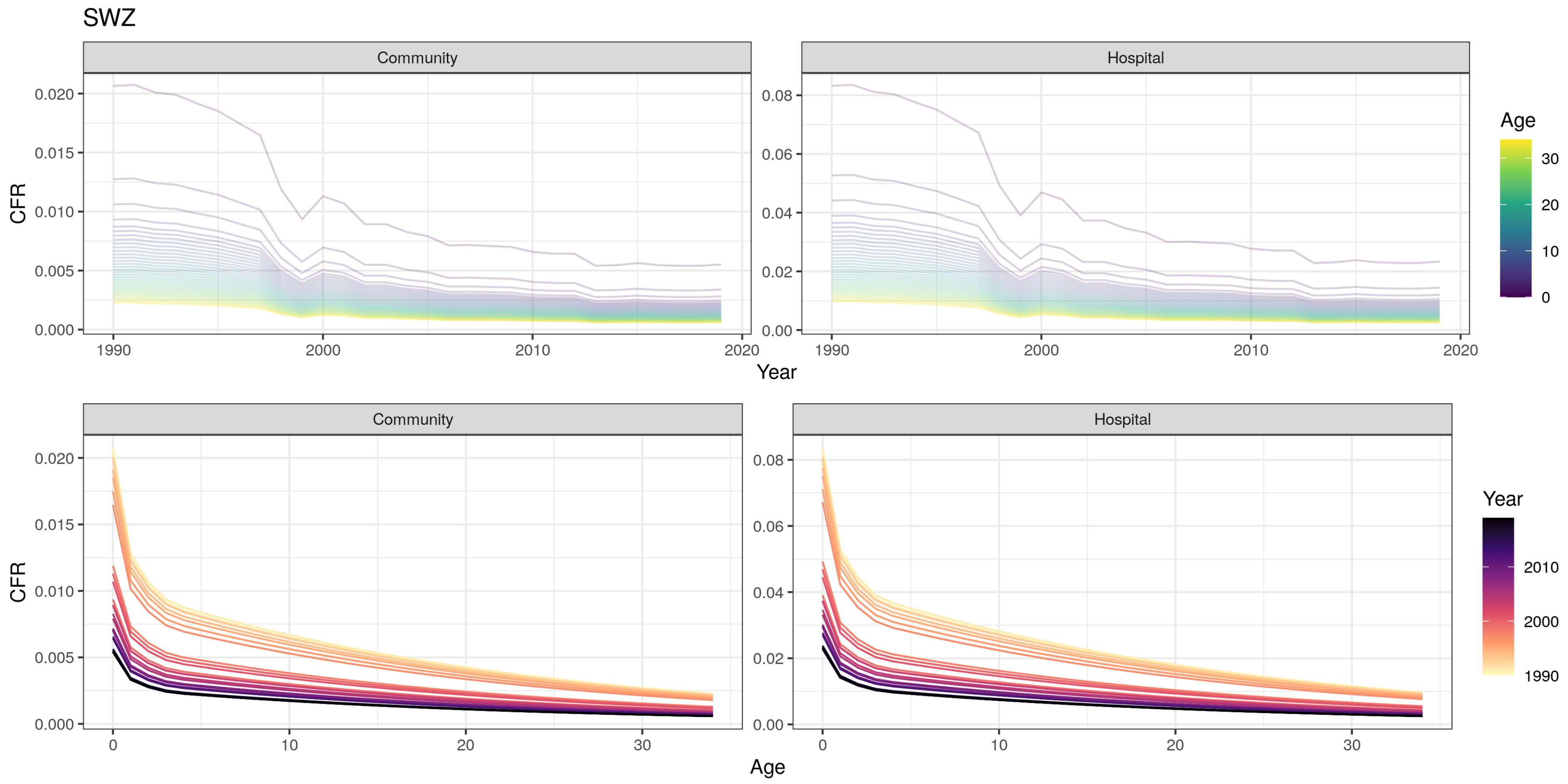
SWZ
Community
Hospital
CFR
Year
Age
0.000
0.005
0.010
0.015
0.020
0.00
0.02
0.04
0.06
0.08
1990
2000
2010
2020
0
10
20
30
Year
1990
2000
2010

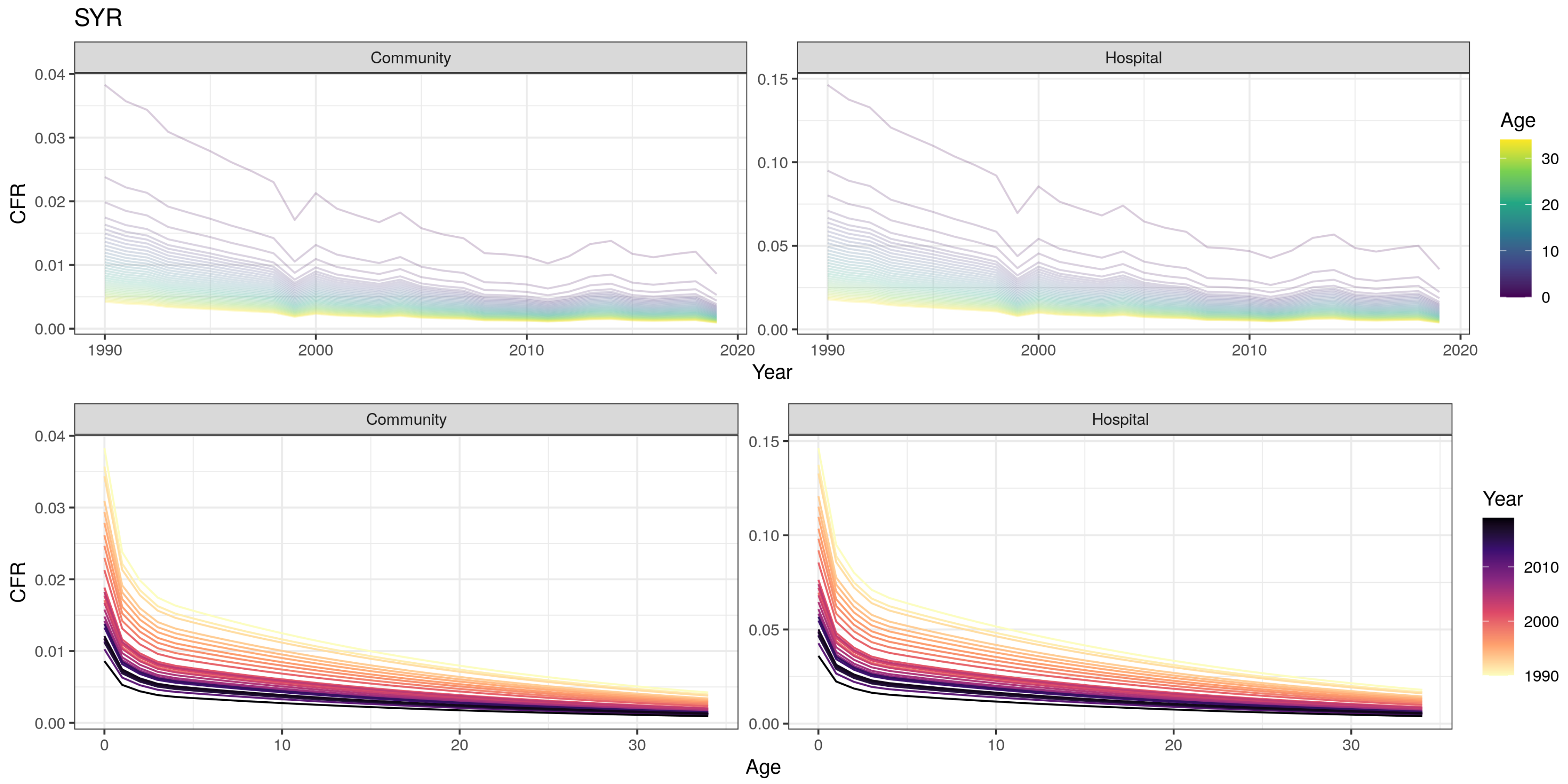
SYR
Community
Hospital
CFR
0.04
0.03
0.02
0.01
0.00
0.15
0.10
0.05
1990
2000
2010
2020
Year
Age
30
20
10
0
Community
Hospital
CFR
Age
Year
2010
2000
1990

TCD

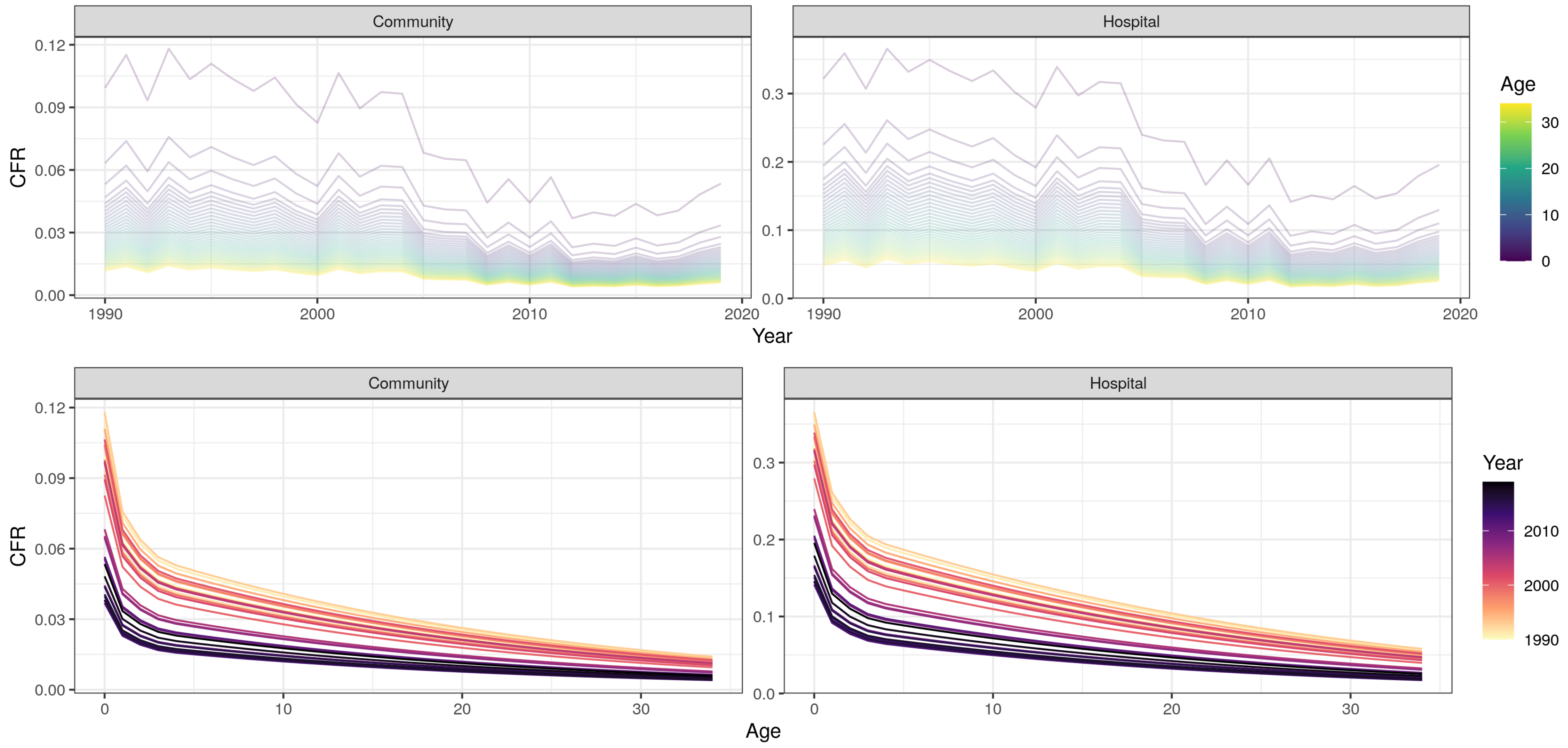

Community
Hospital
0.12
0.09
0.06
0.03
0.00
CFR
0.3
0.2
0.1
0.0
1990
2000
2010
2020
Year
Age
30
20
10
0
Community
Hospital
0.12
0.09
0.06
0.03
0.00
CFR
0.3
0.2
0.1
0.0
0
10
20
30
Age
Year
2010
2000
1990

TGO

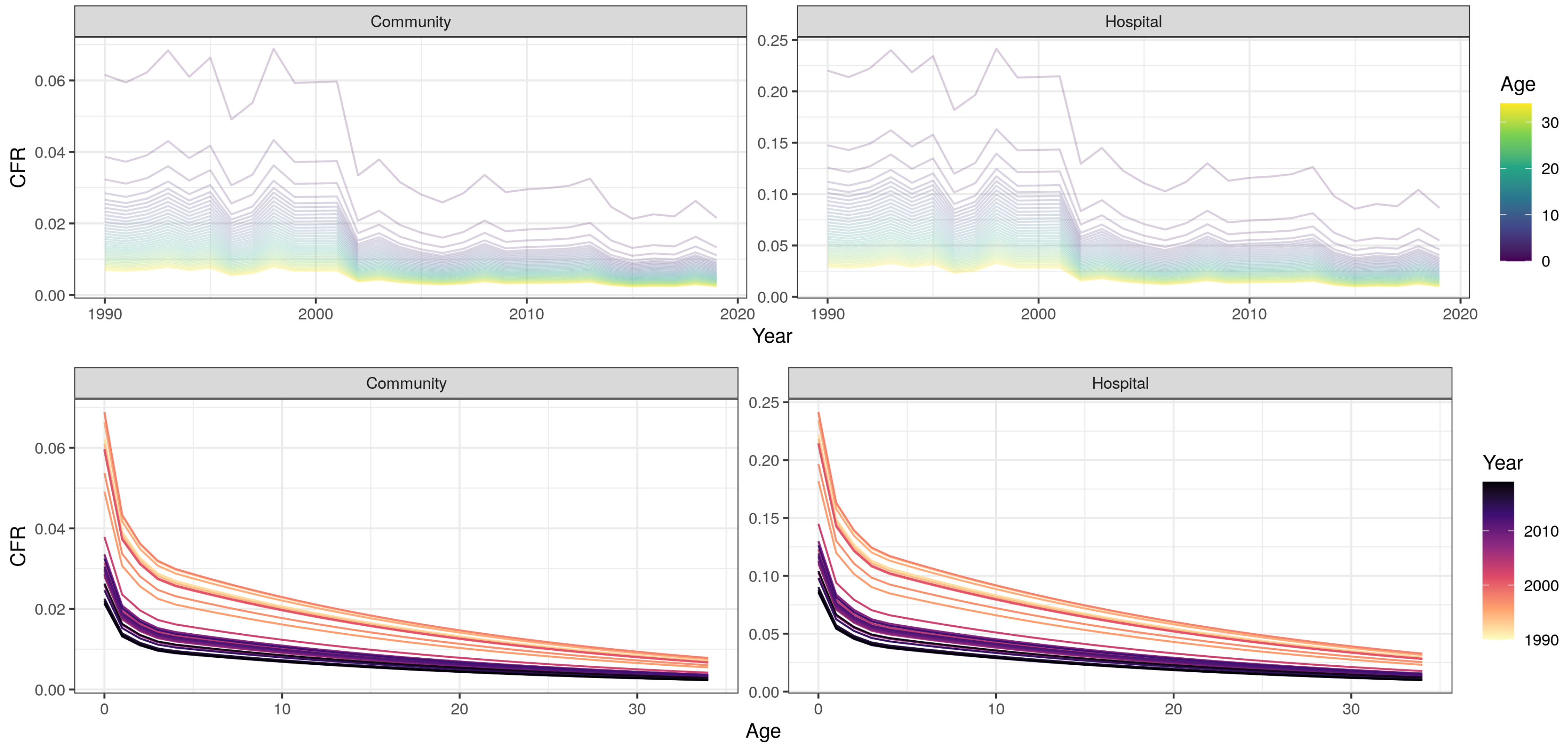
Community
Hospital
CFR
Year
Age
Community
Hospital
CFR
Age
Year

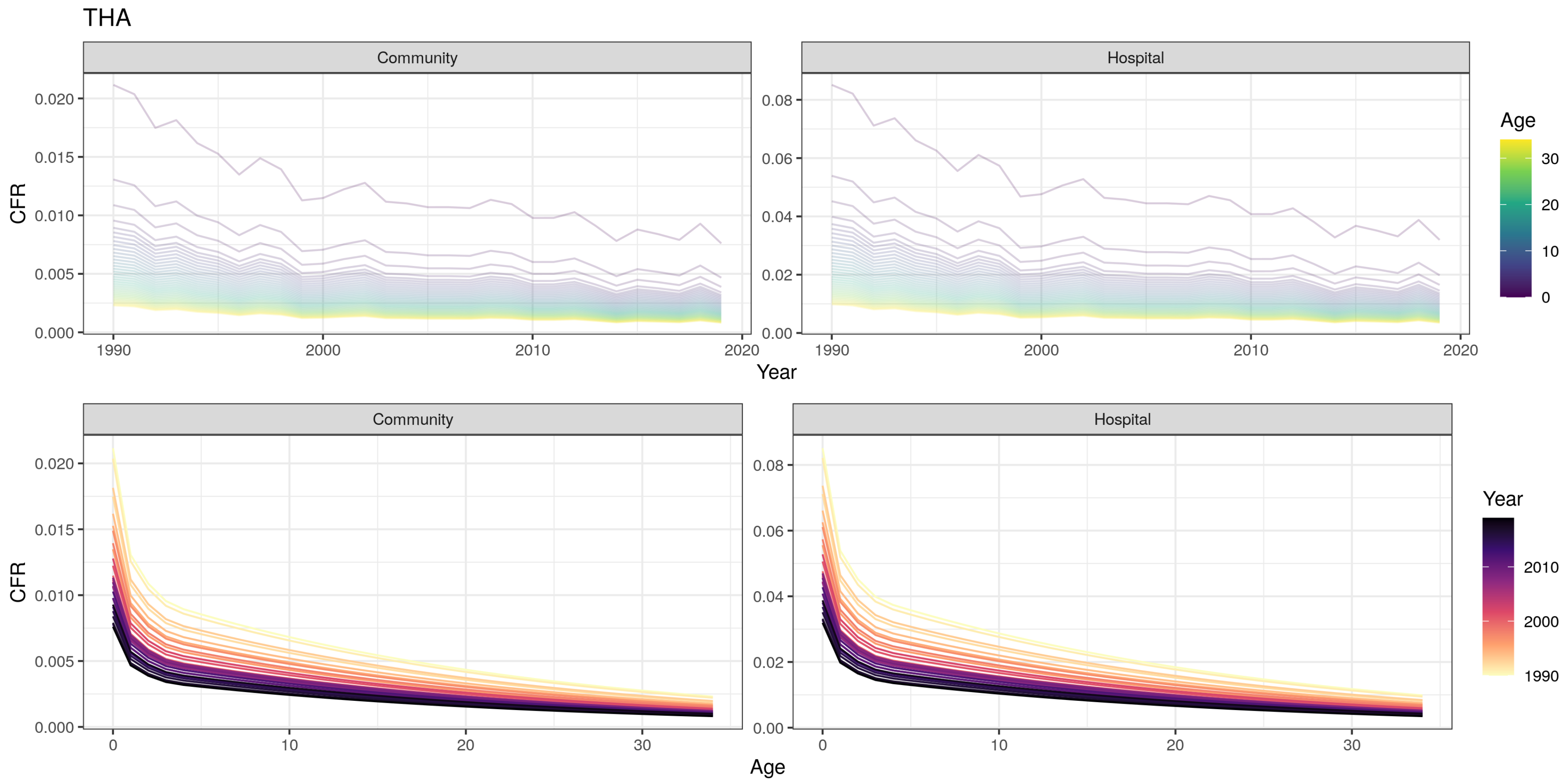
THA
Community
Hospital
CFR
Year
Age
0.000
0.005
0.010
0.015
0.020
0.00
0.02
0.04
0.06
0.08
1990
2000
2010
2020
0
10
20
30
Age
Year
2010
2000
1990

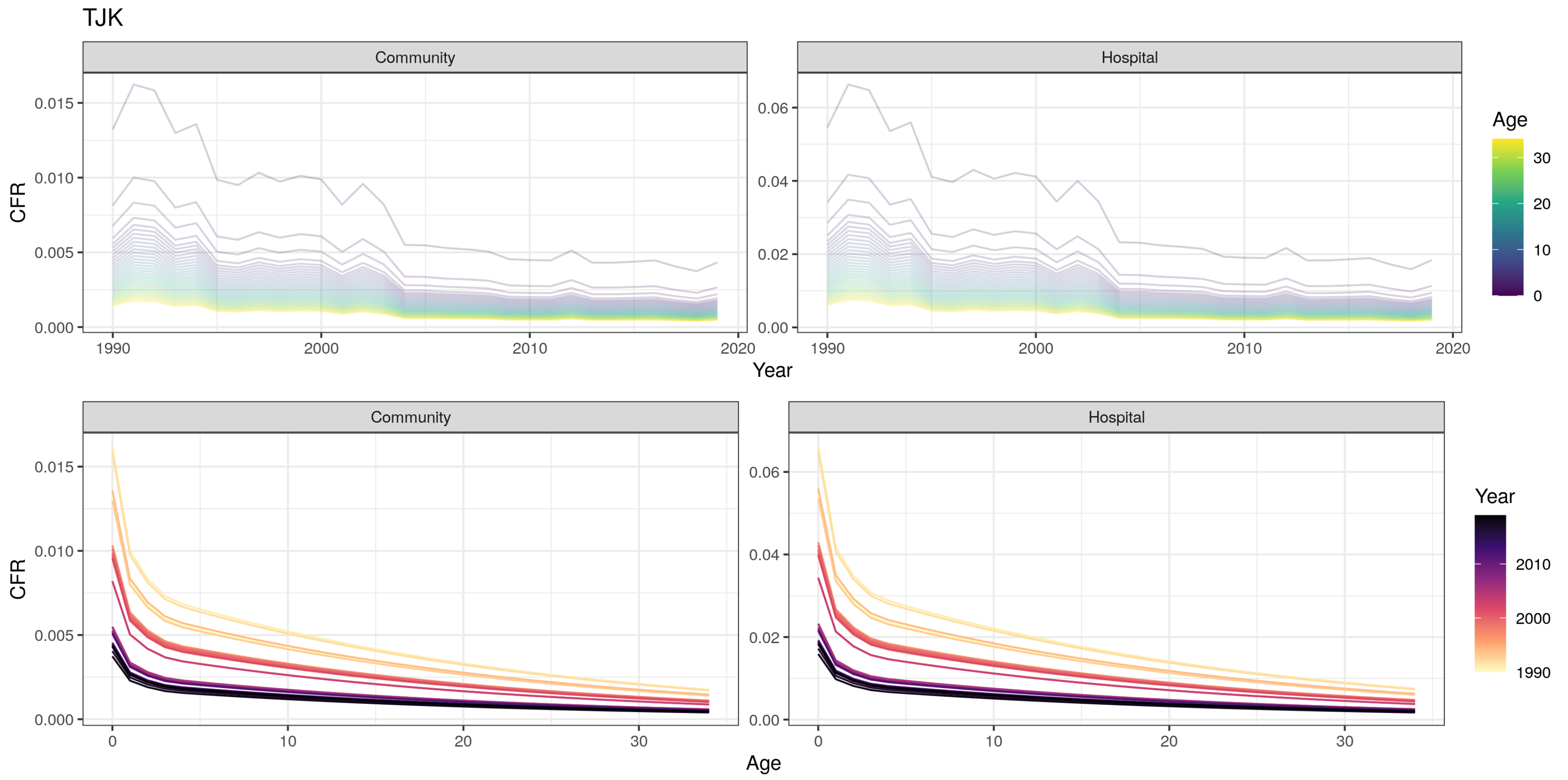

TJK
Community
Hospital
CFR
0.000
0.005
0.010
0.015
0.00
0.02
0.04
0.06
1990
2000
2010
2020
Year
Age
0
10
20
30
Community
Hospital
CFR
Age
Year
1990
2000
2010

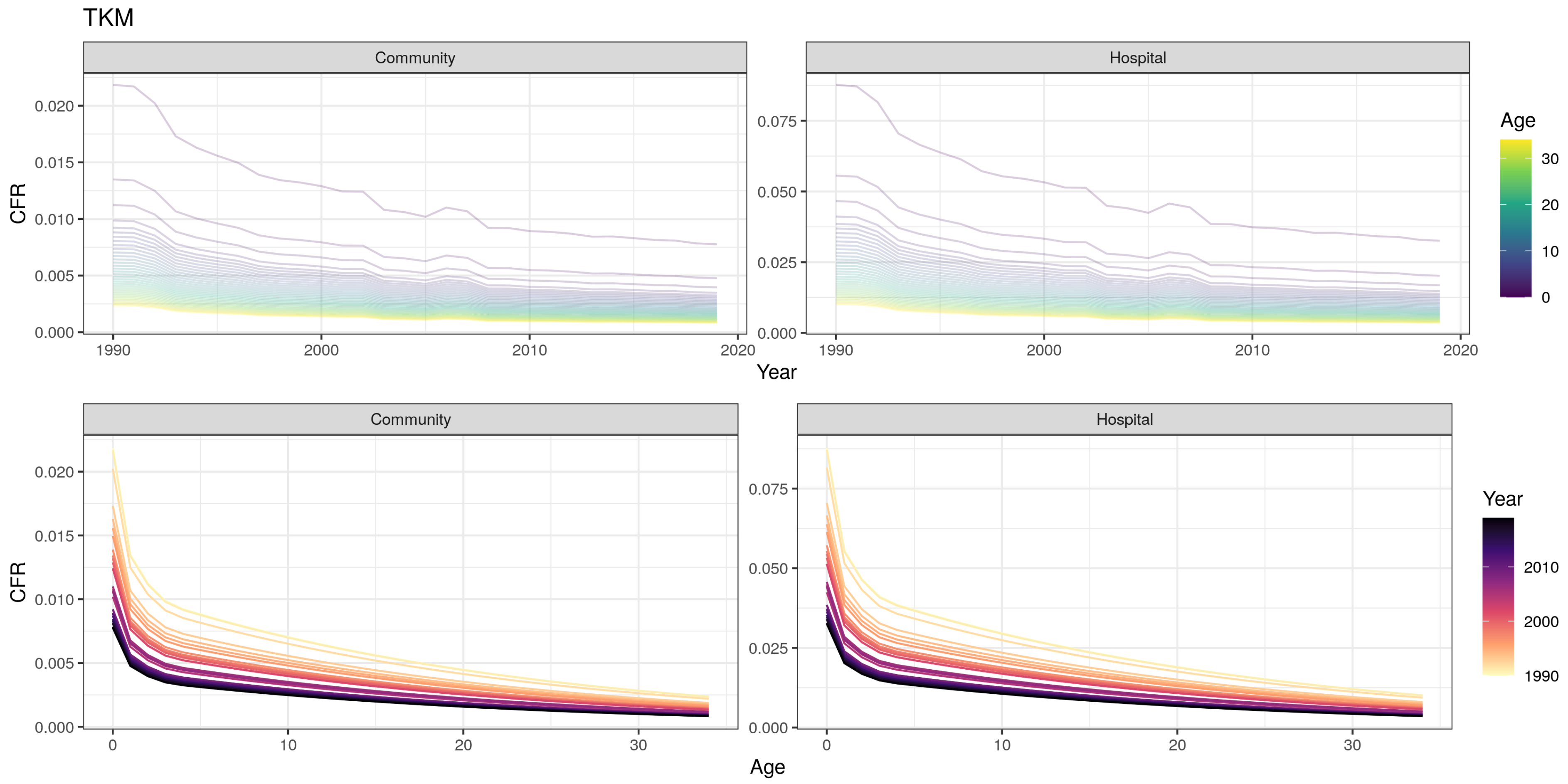

TKM
Community
Hospital
CFR
0.020
0.015
0.010
0.005
0.000
0.075
0.050
0.025
1990
2000
2010
2020
Year
Age
30
20
10
0
0
10
20
30
Year
2010
2000
1990

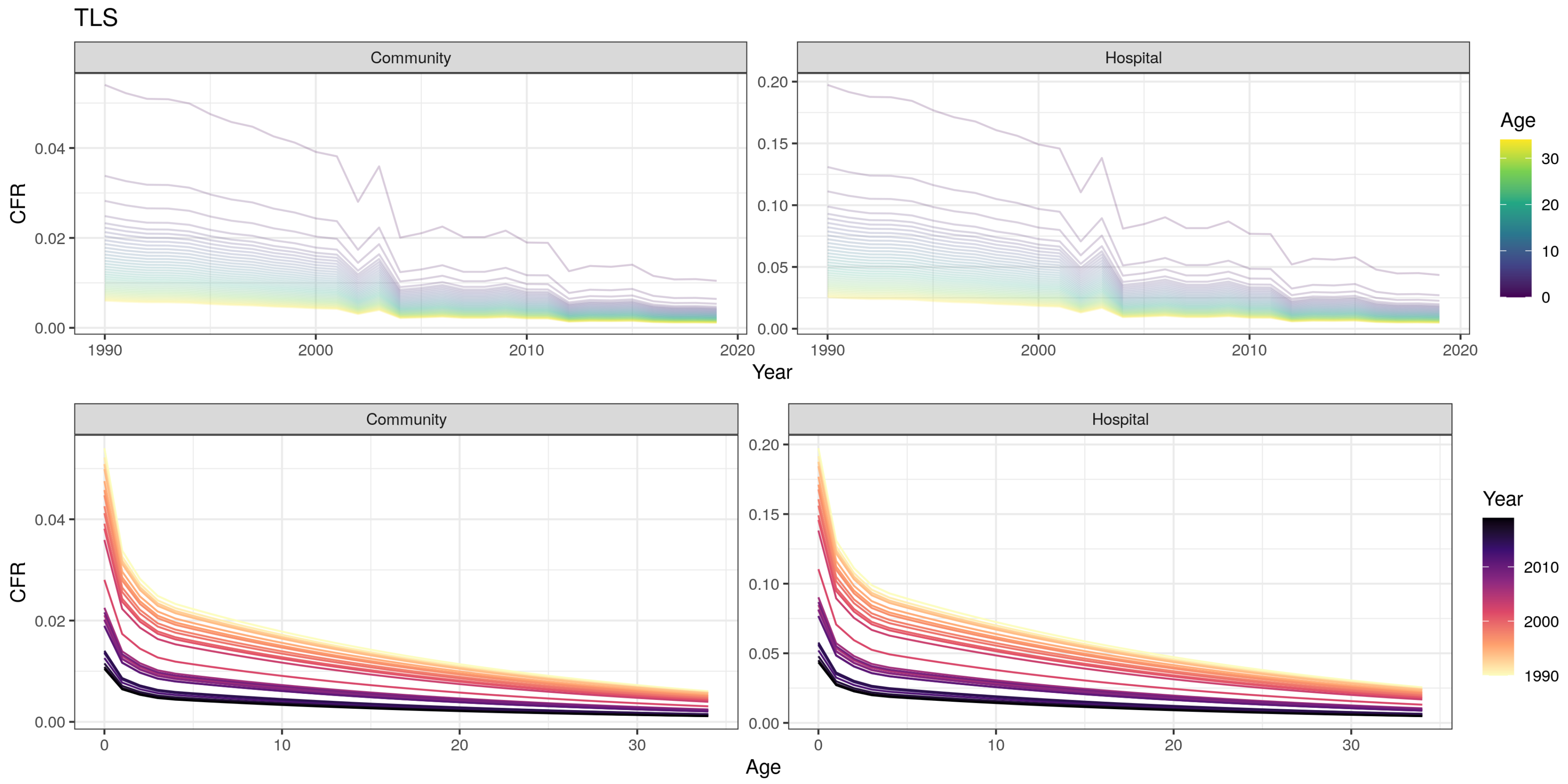
TLS
Community
Hospital
CFR
0.00
0.02
0.04
0.05
0.10
0.15
0.20
1990
2000
2010
2020
Year
Age
0
10
20
30
Community
Hospital
CFR
Age
Year
1990
2000
2010

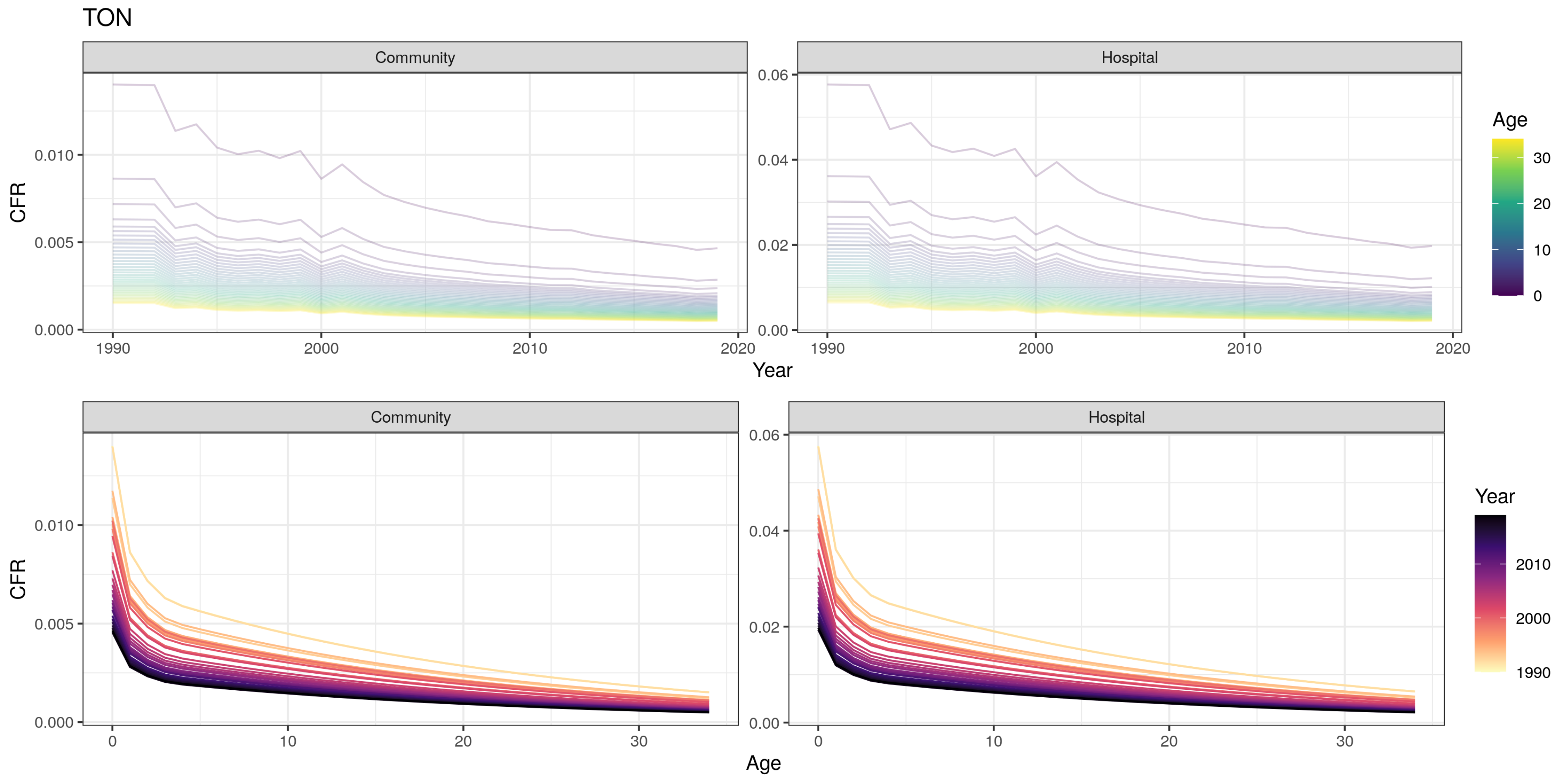

TON
Community
Hospital
CFR
0.000
0.005
0.010
0.00
0.02
0.04
0.06
1990
2000
2010
2020
Year
Age
0
10
20
30
Community
Hospital
CFR
0.000
0.005
0.010
0.00
0.02
0.04
0.06
0
10
20
30
Age
Year
1990
2000
2010

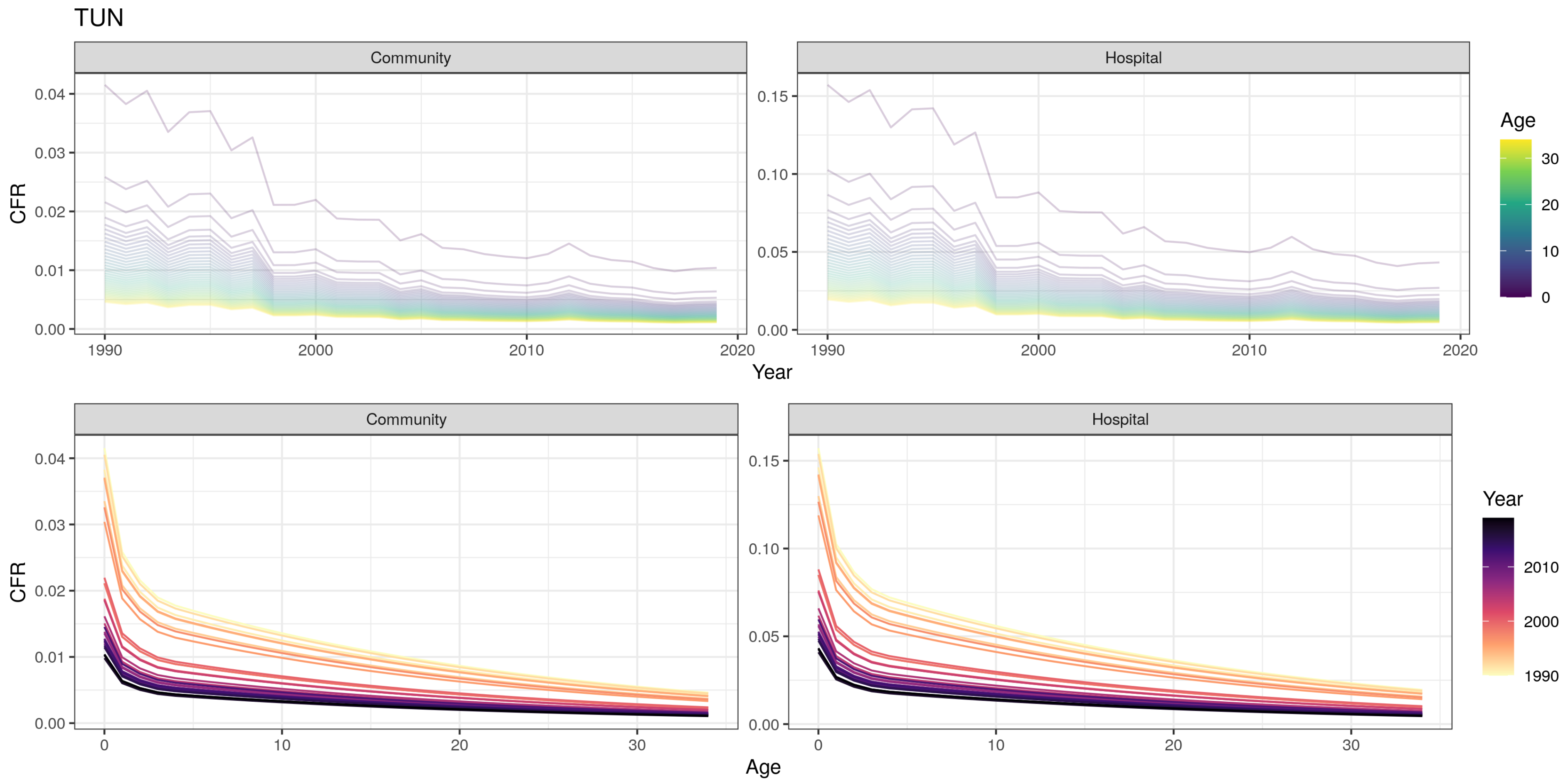

TUN
Community
Hospital
CFR
Year
Age
Community
Hospital
CFR
Age
Year

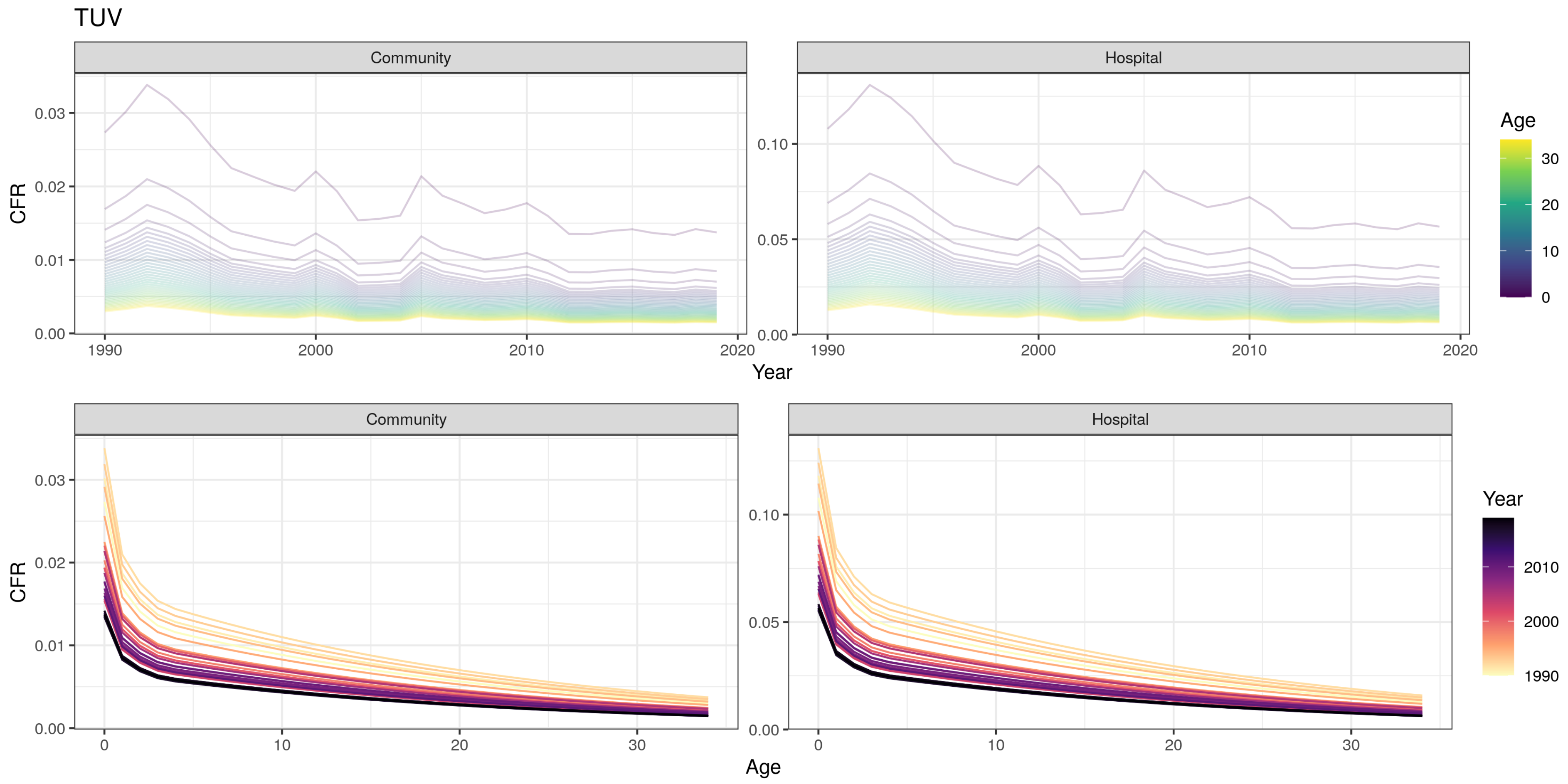
TUV
Community
Hospital
CFR
0.03
0.02
0.01
0.00
0.10
0.05
1990
2000
2010
2020
Year
Age
30
20
10
0
Community
Hospital
CFR
0
10
20
30
Age
Year
2010
2000
1990

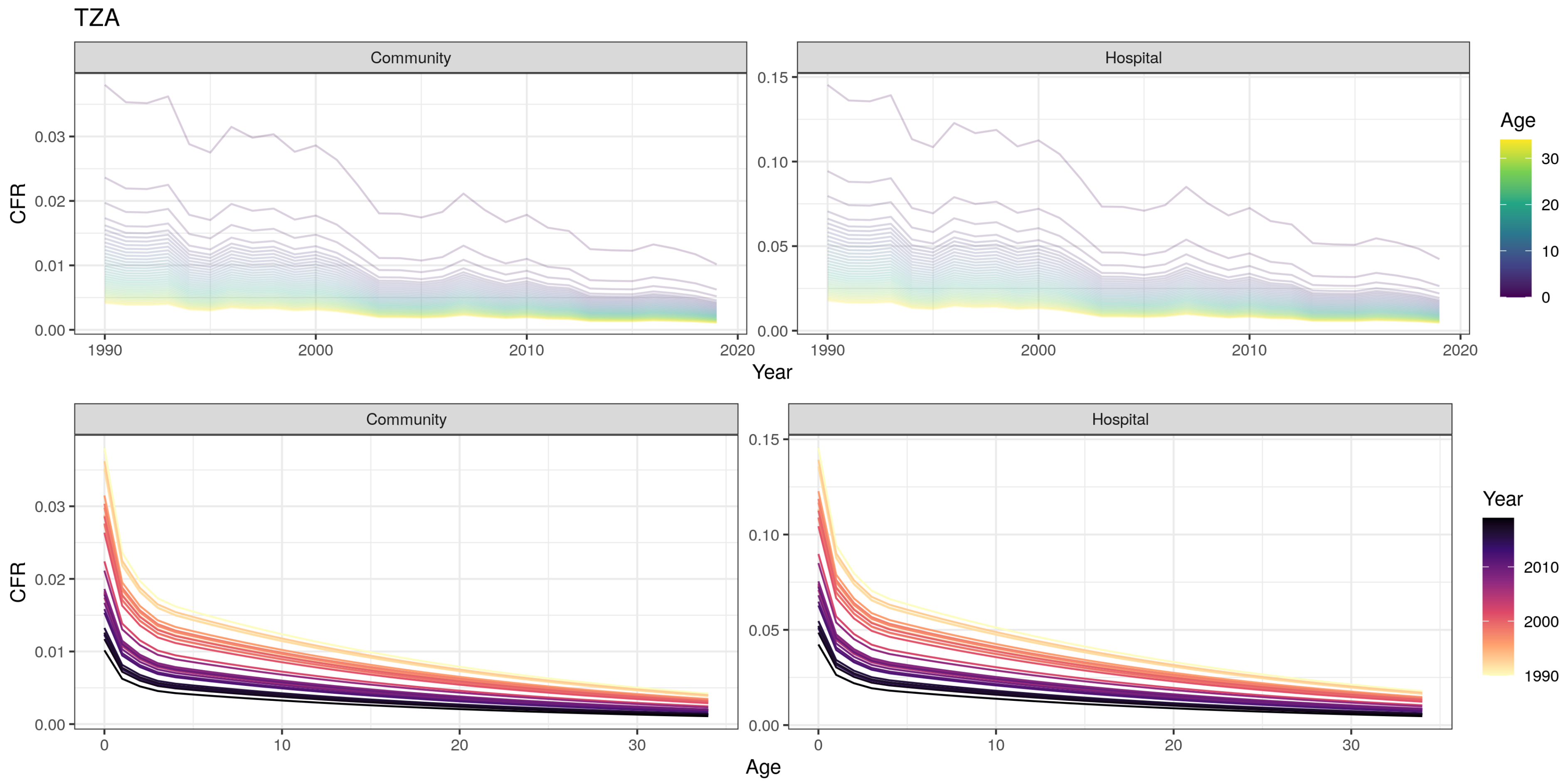
TZA
Community
Hospital
CFR
Year
Age
Community
Hospital
CFR
Age
Year
0.00
0.01
0.02
0.03
0.05
0.10
0.15
1990
2000
2010
2020
0
10
20
30

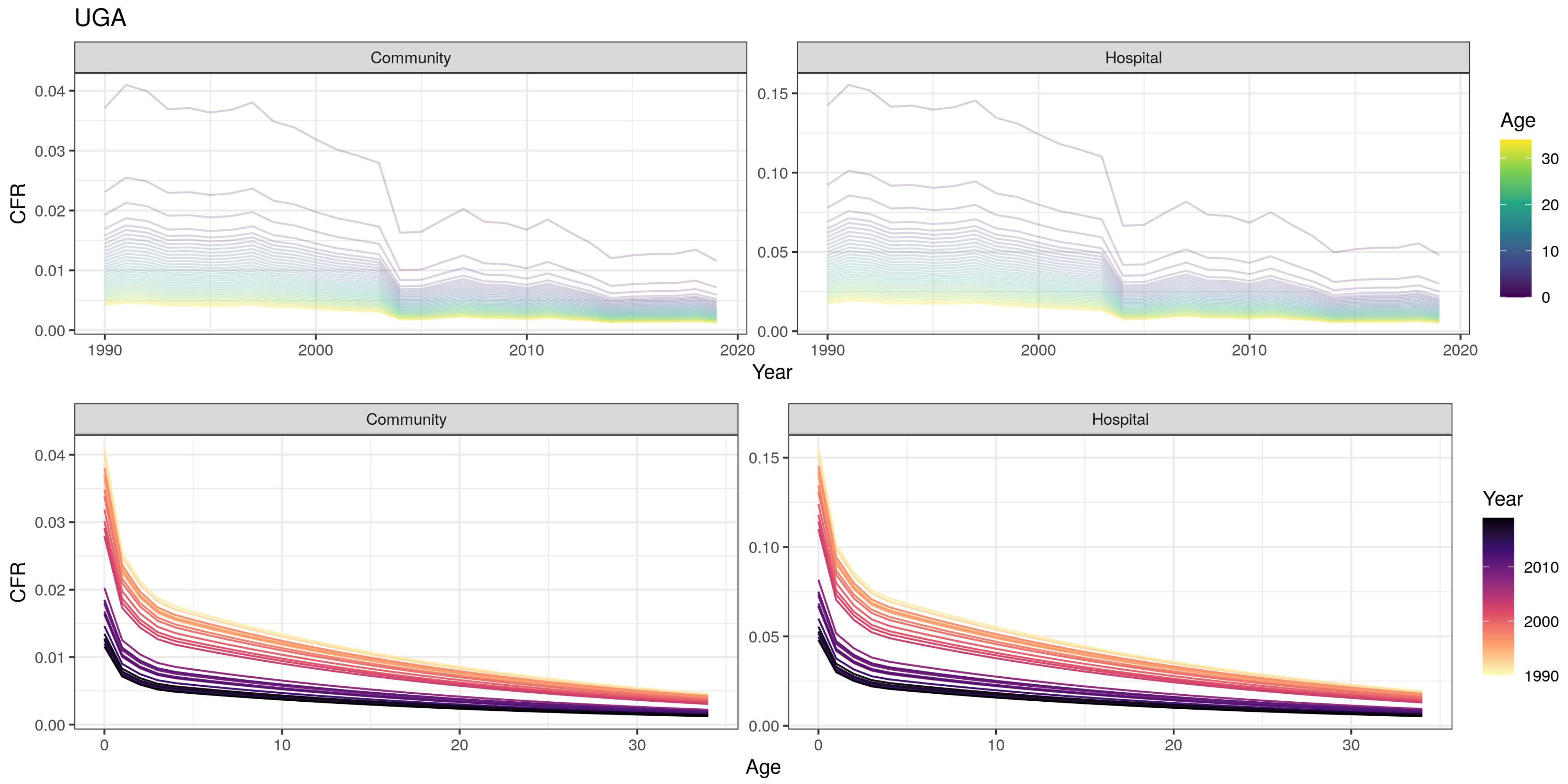

UGA
Community
Hospital
CFR
0.04
0.03
0.02
0.01
0.00
0.15
0.10
0.05
1990
2000
2010
2020
Year
Age
30
20
10
0
Community
Hospital
Age
Year
2010
2000
1990

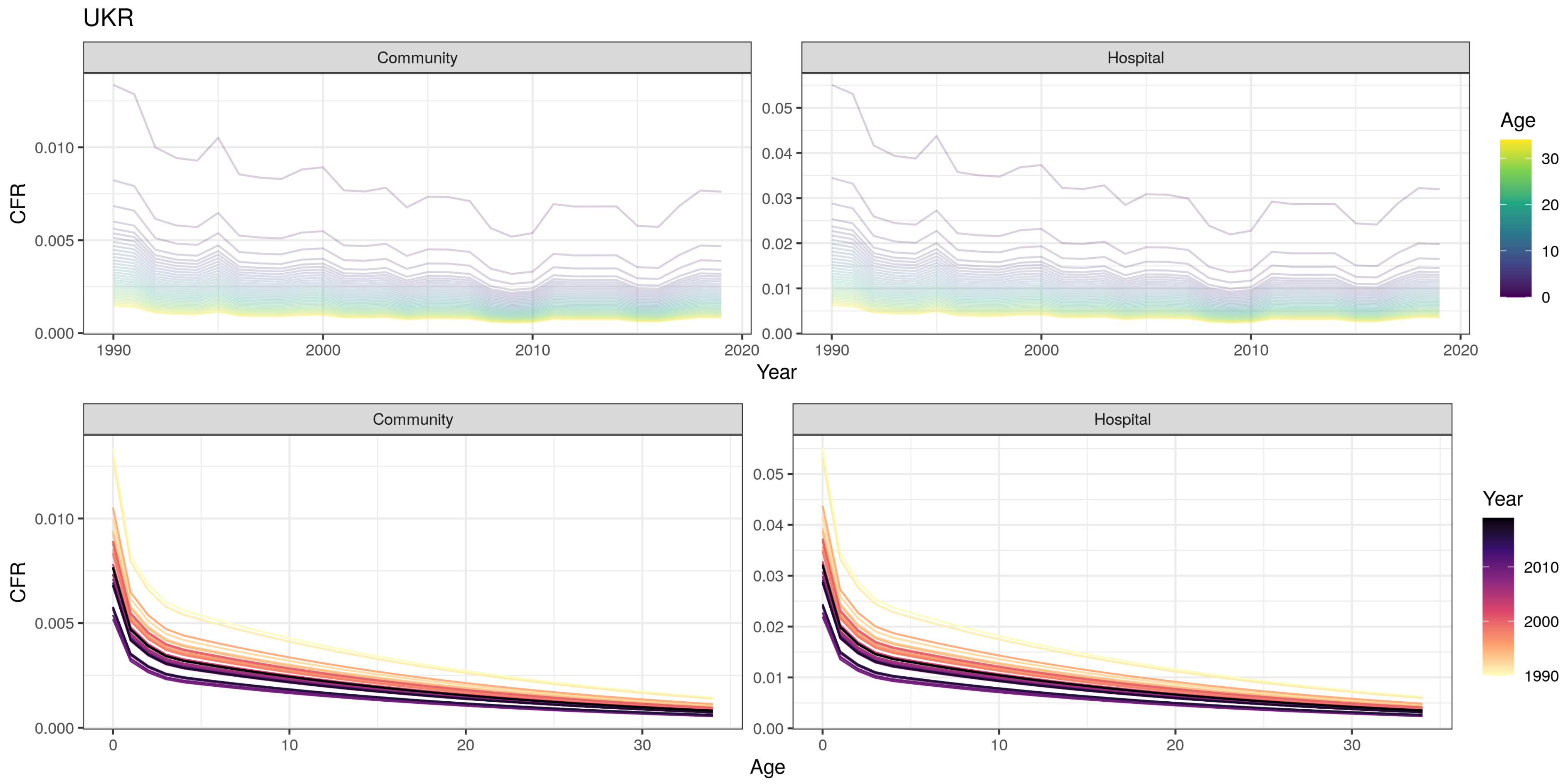

UKR
Community
Hospital
CFR
0.000
0.005
0.010
0.00
0.01
0.02
0.03
0.04
0.05
1990
2000
2010
2020
Year
Age
30
20
10
0
Community
Hospital
CFR
0
10
20
30
Age
Year
2010
2000
1990

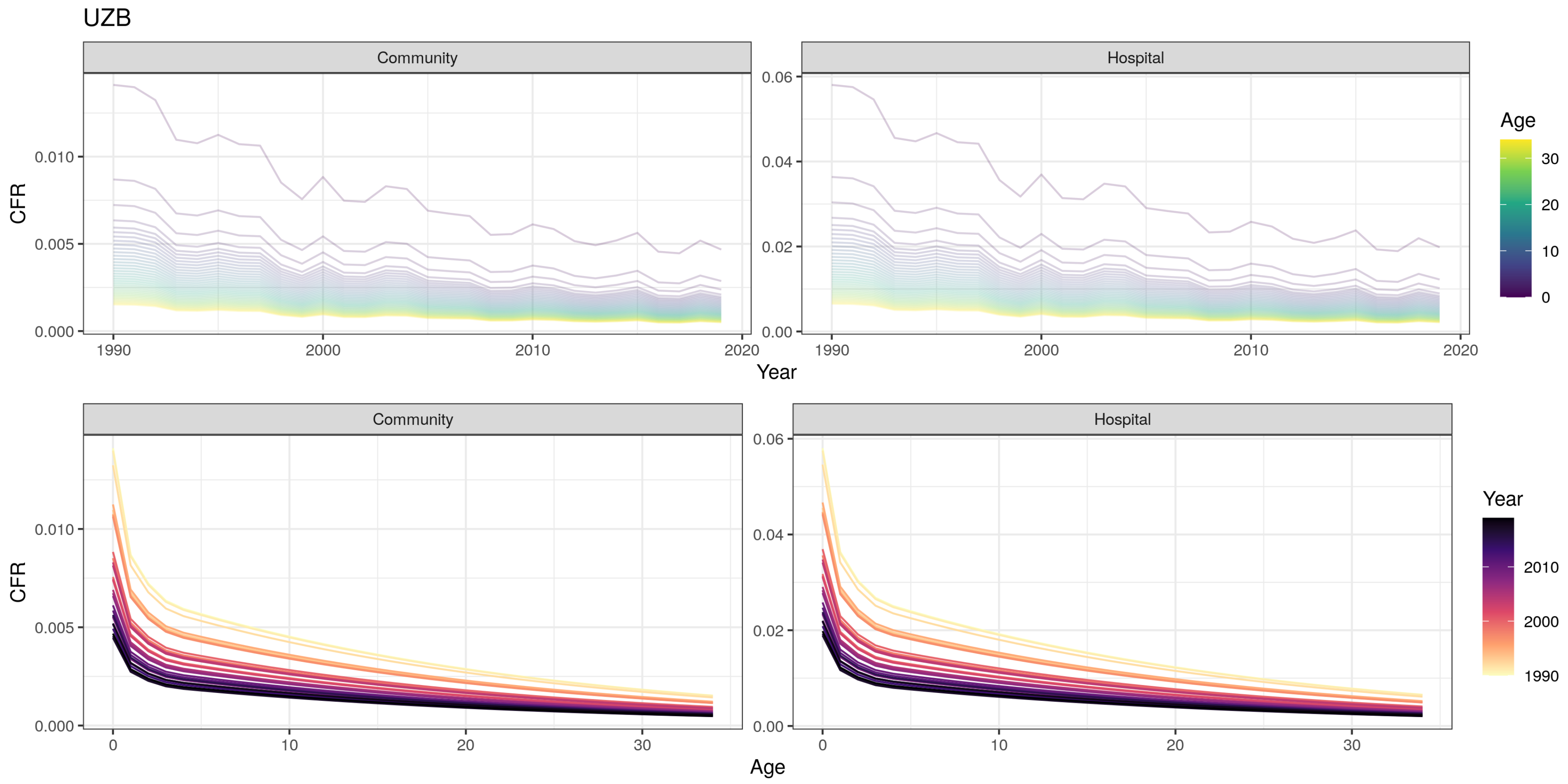

UZB
Community
Hospital
CFR
0.000
0.005
0.010
0.00
0.02
0.04
0.06
1990
2000
2010
2020
Year
Age
0
10
20
30
Year
2010
2000
1990

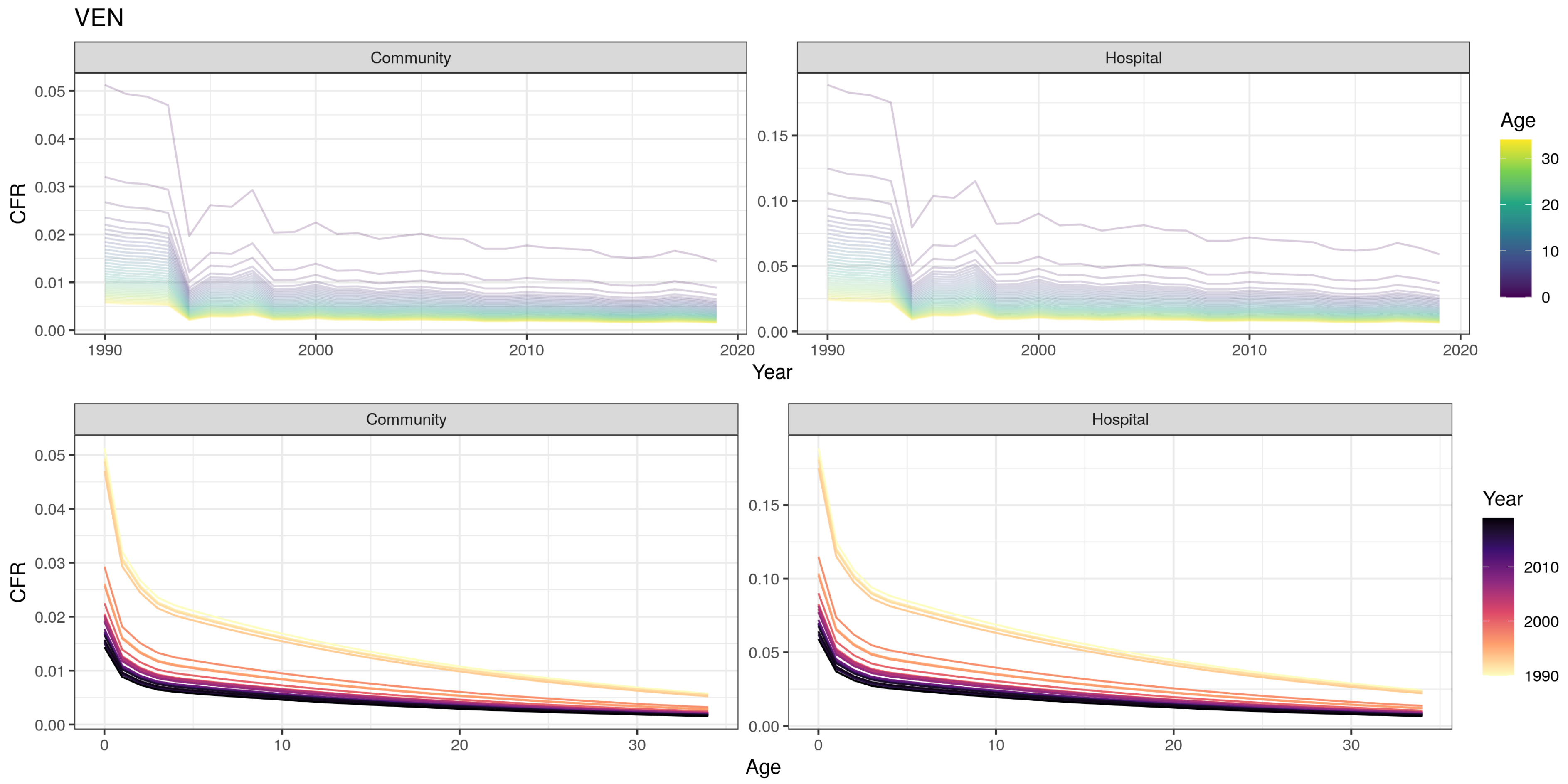

VEN
Community
Hospital
CFR
Year
Age
0.00
0.01
0.02
0.03
0.04
0.05
0.10
0.15
1990
2000
2010
2020
0
10
20
30

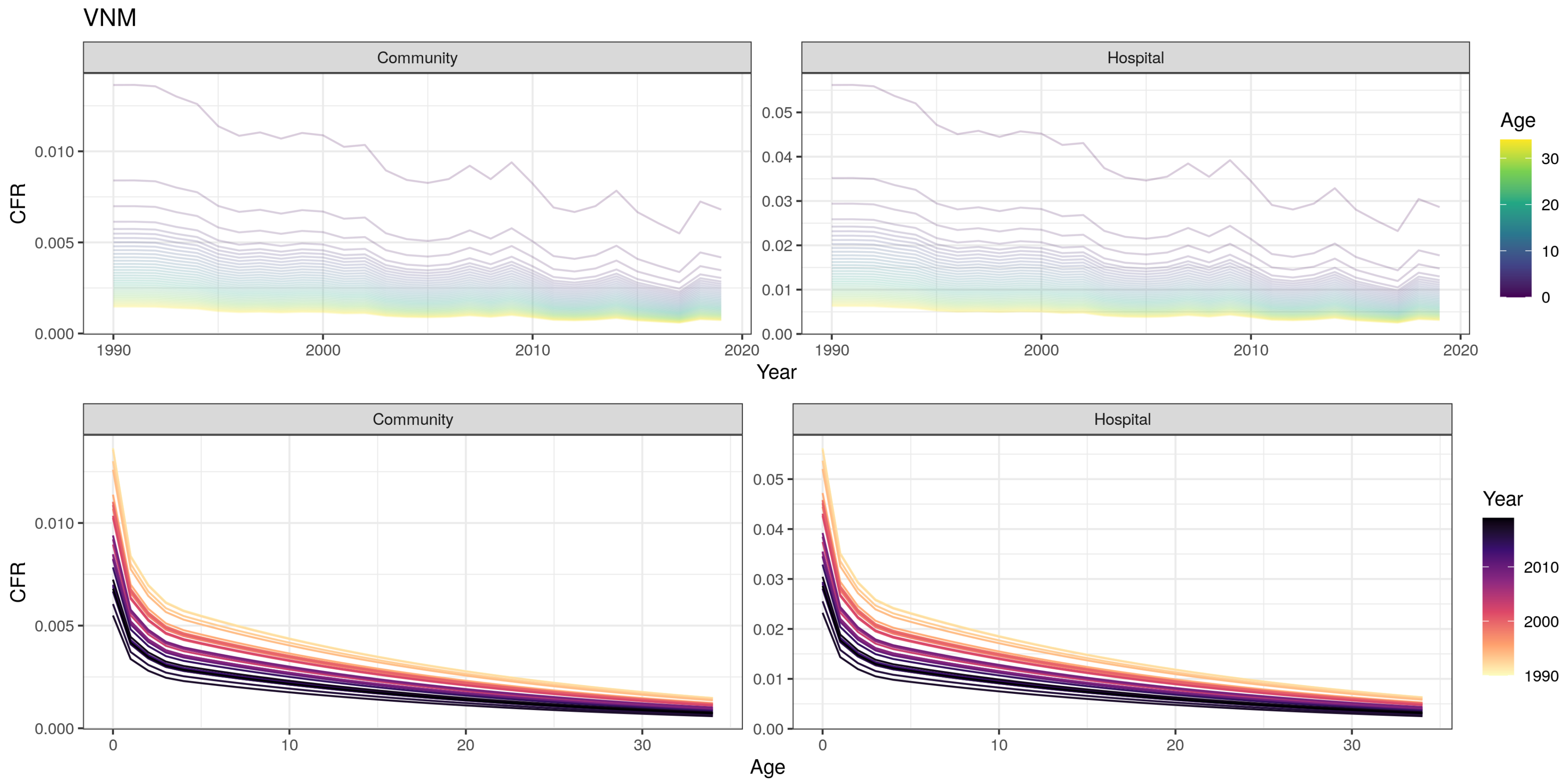
VNM
Community
Hospital
CFR
Year
Age
0.000
0.005
0.010
0.00
0.01
0.02
0.03
0.04
0.05
1990
2000
2010
2020
0
10
20
30
Age
Year

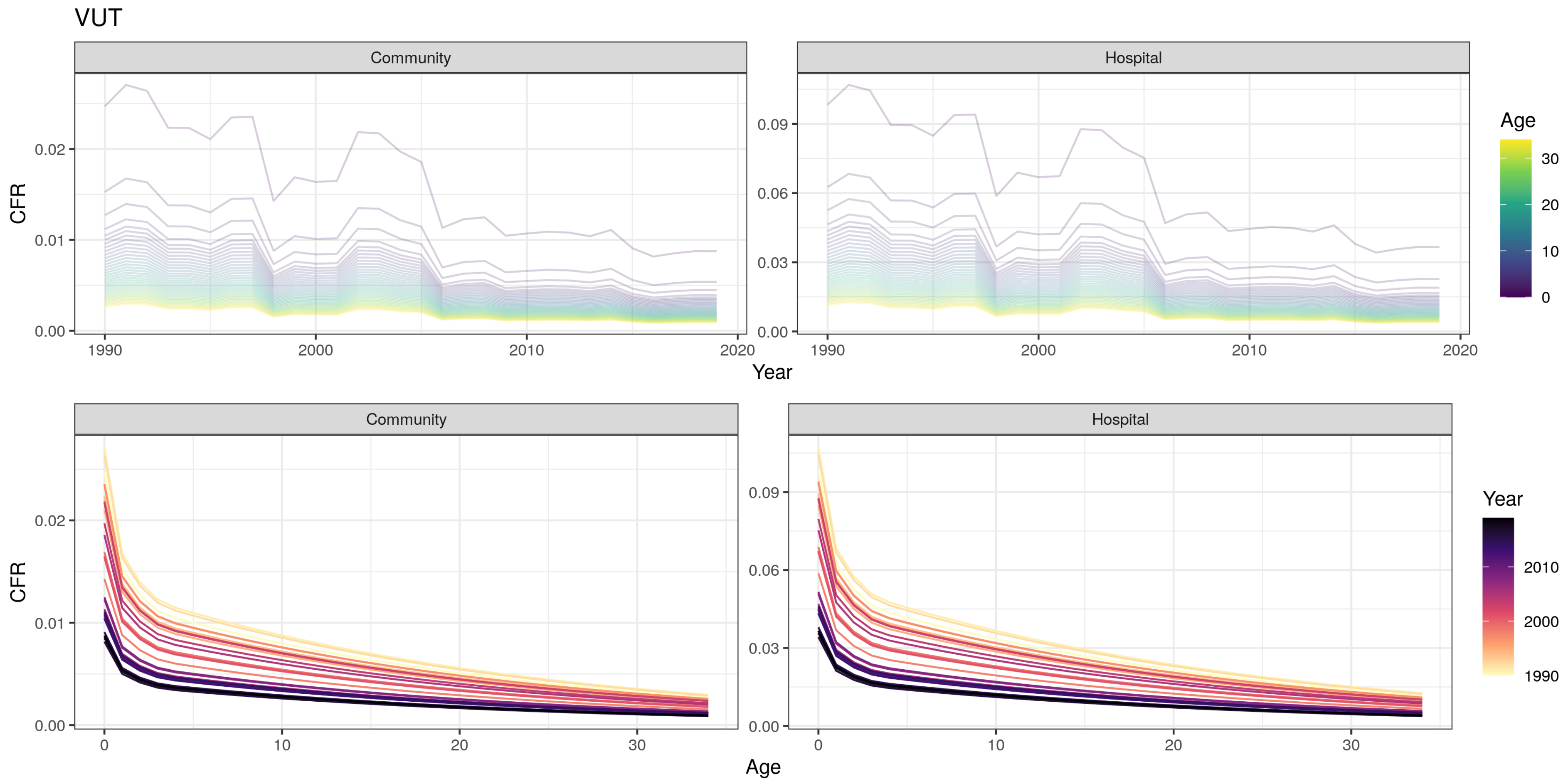
VUT
Community
Hospital
CFR
0.00
0.01
0.02
0.03
0.06
0.09
1990
2000
2010
2020
Year
Age
0
10
20
30
Community
Hospital
CFR
0.00
0.01
0.02
0.03
0.06
0.09
0
10
20
30
Age
Year
1990
2000
2010

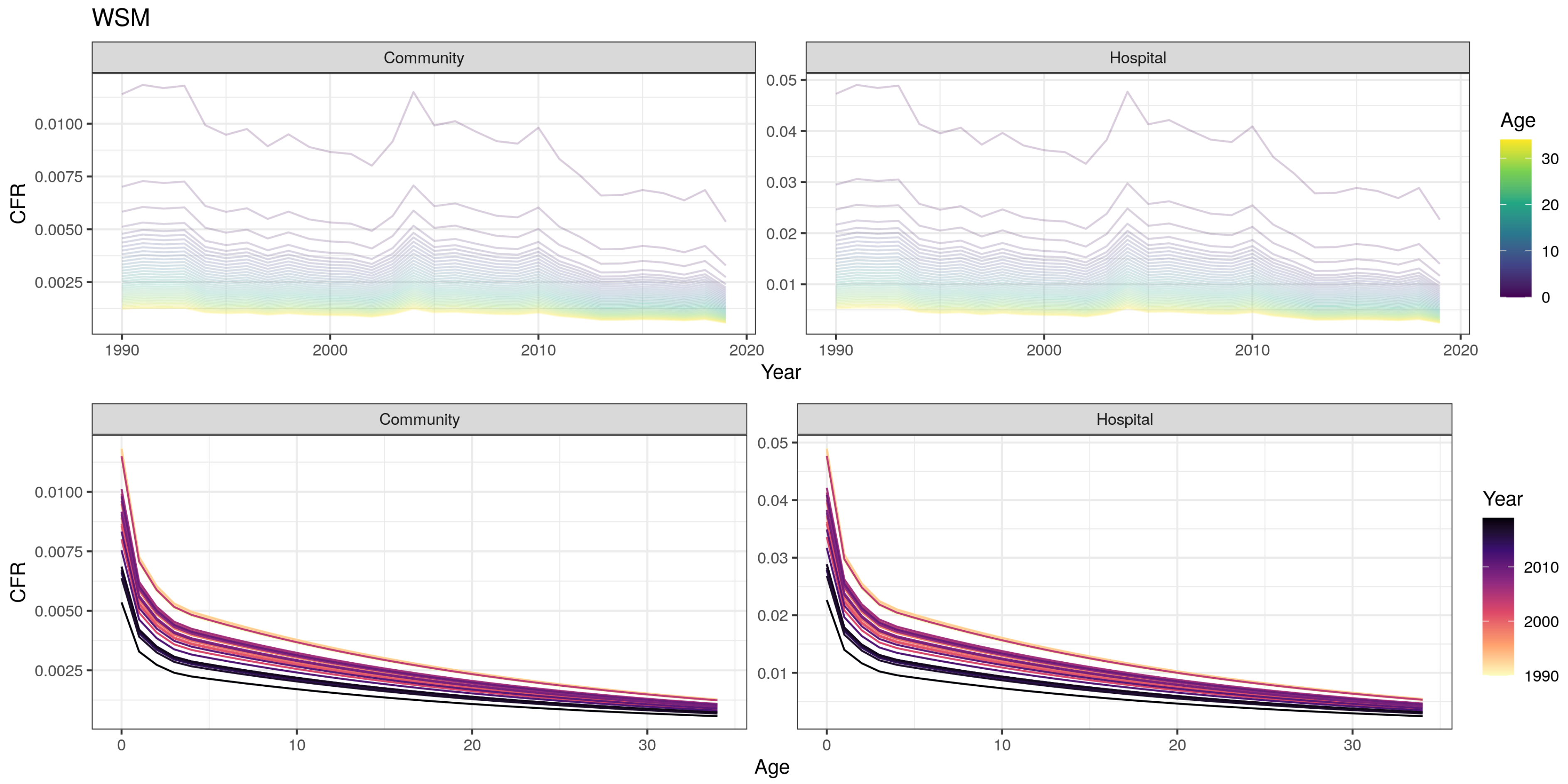

WSM
Community
Hospital
CFR
0.0100
0.0075
0.0050
0.0025
0.05
0.04
0.03
0.02
0.01
1990
2000
2010
2020
Year
Age
30
20
10
0
Community
Hospital
CFR
0
10
20
30
Age
Year
2010
2000
1990

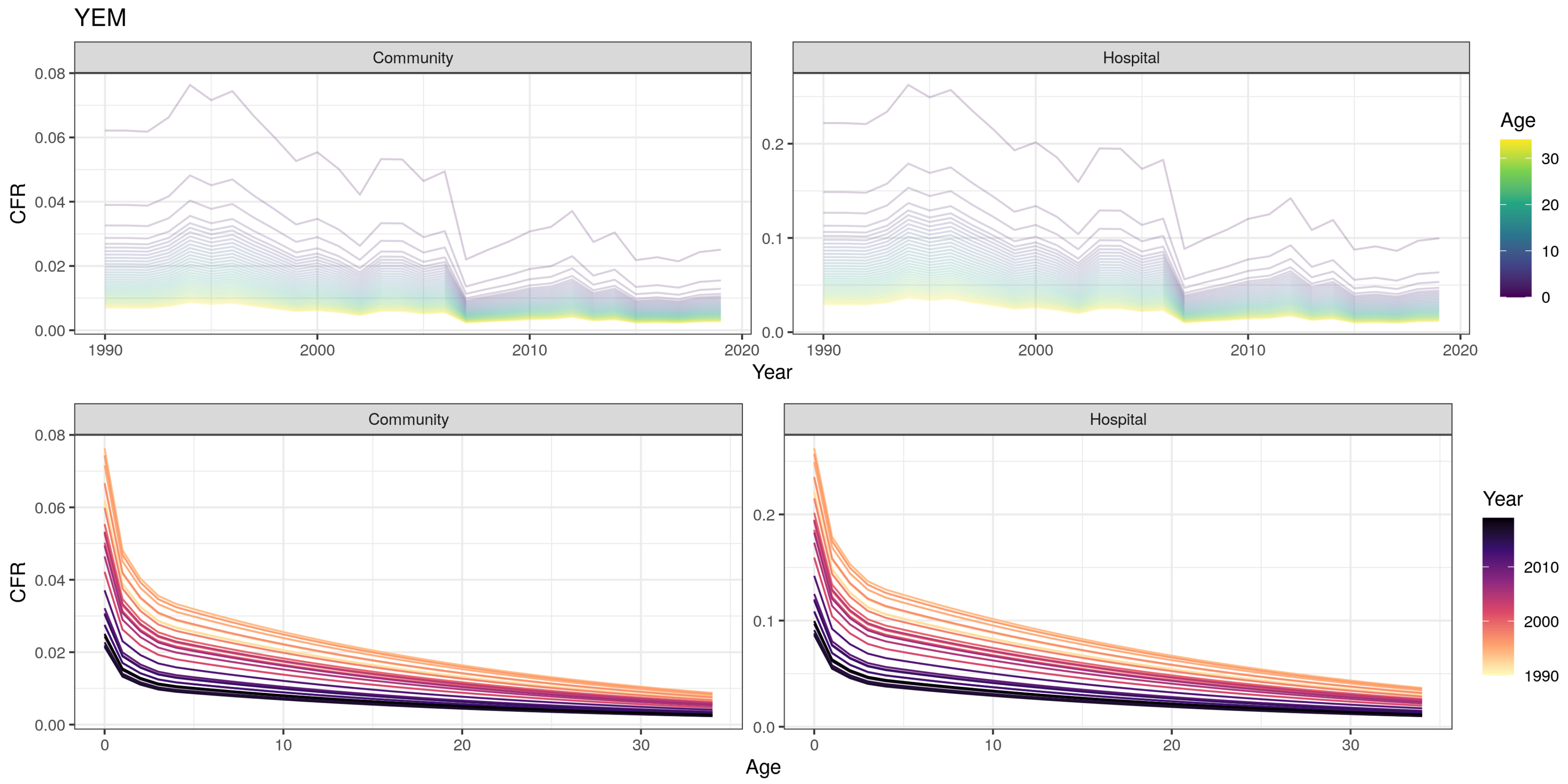

YEM
Community
Hospital
CFR
0.08
0.06
0.04
0.02
0.00
0.2
0.1
0.0
1990
2000
2010
2020
Year
Age
30
20
10
0
Community
Hospital
CFR
0
10
20
30
Age
Year
2010
2000
1990

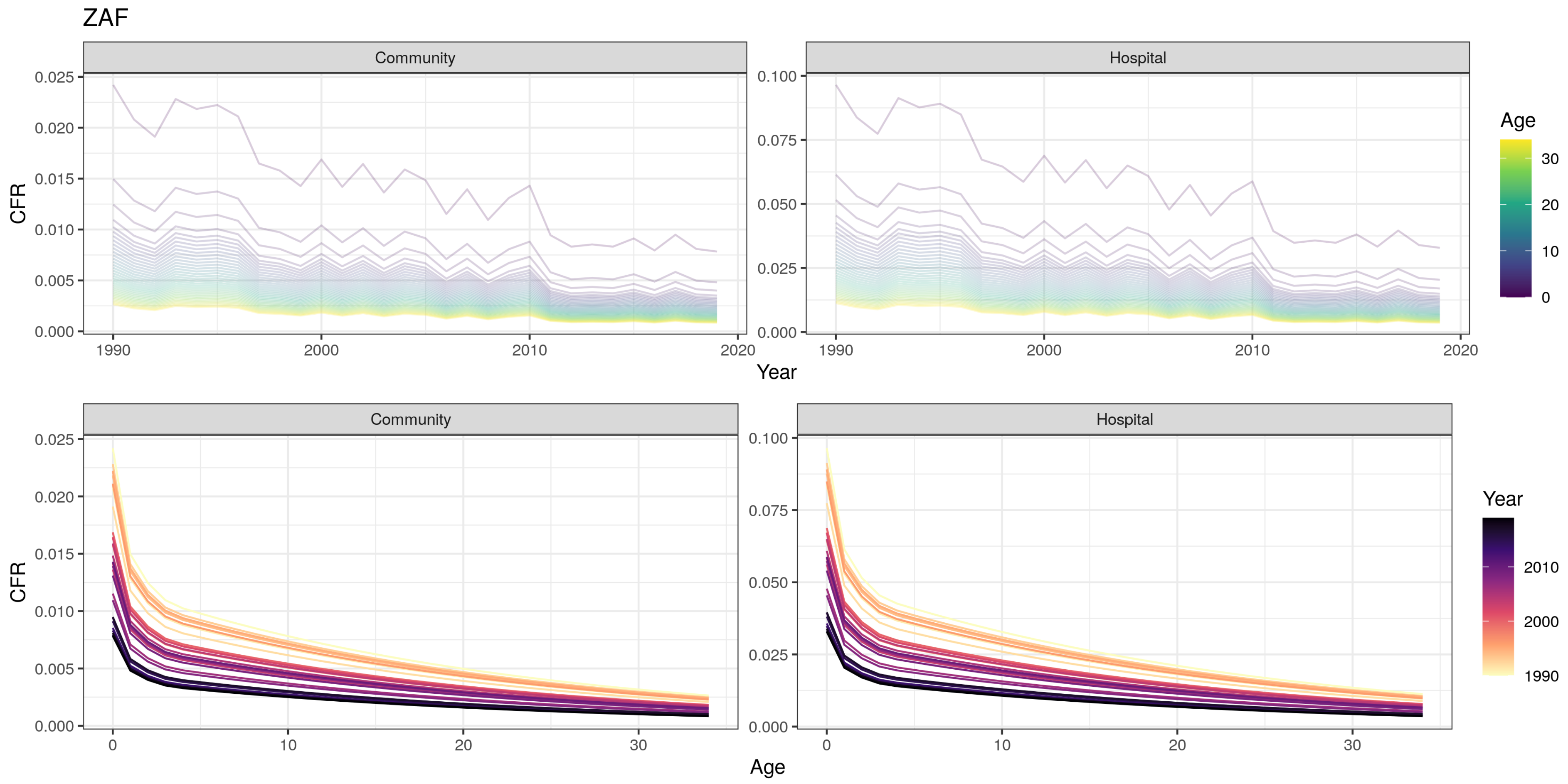
ZAF
Community
Hospital
CFR
Year
Age
Community
Hospital
CFR
Age
Year

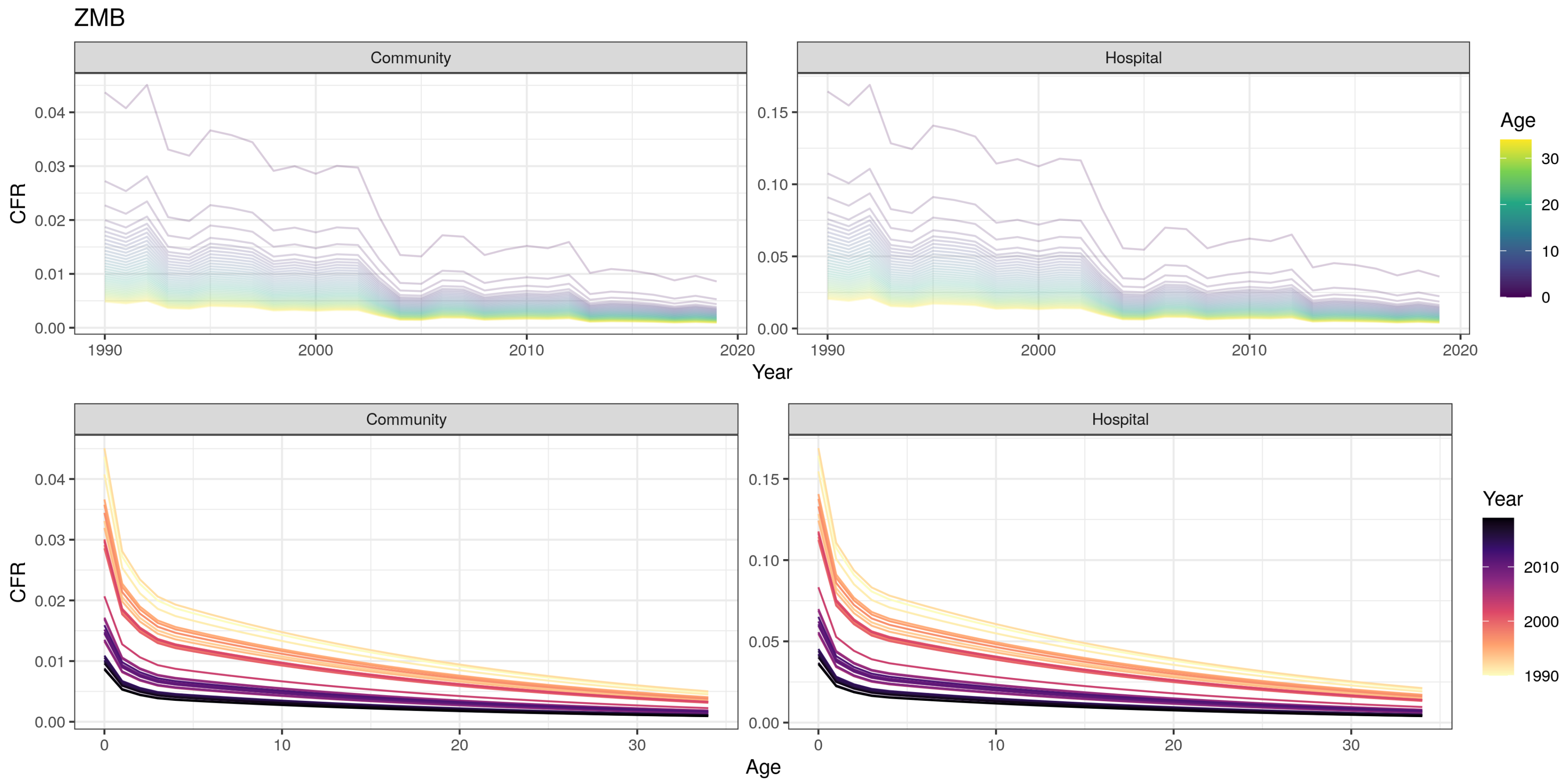

ZMB
Community
Hospital
Age
30
20
10
0
CFR
0.04
0.03
0.02
0.01
0.00
0.15
0.10
0.05
1990
2000
2010
2020
Year
Community
Hospital
Year
2010
2000
1990
CFR
0
10
20
30
Age

ZWE

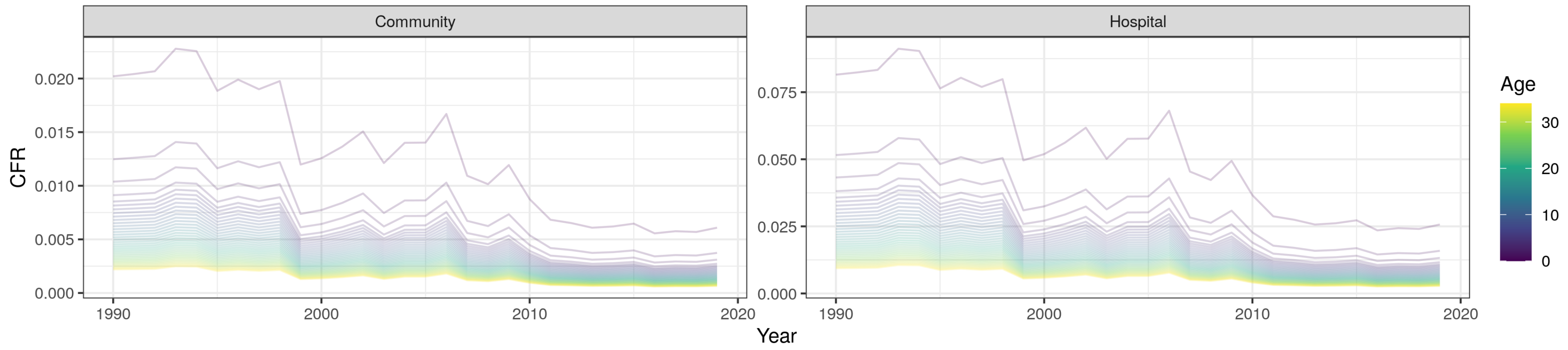
Community
Hospital
CFR
0.020
0.015
0.010
0.005
0.000
0.075
0.050
0.025
0.000
1990
2000
2010
2020
Year
Age
30
20
10
0

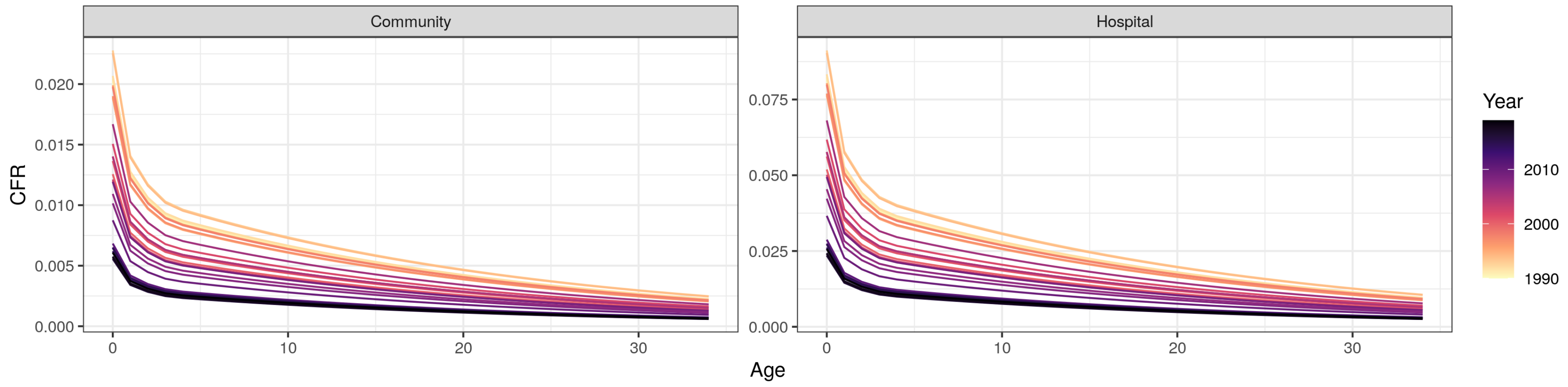
Community
Hospital
CFR
0.020
0.015
0.010
0.005
0.000
0.075
0.050
0.025
0.000
0
10
20
30
Age
Year
2010
2000
1990
